# Lifespan brain structural variation reveals shared organization across mental health conditions

**DOI:** 10.64898/2026.08.13.26360304

**Authors:** Meike D. Hettwer, Amin Saberi, Golia Shafiei, Aikaterina Manoli, Augustijn A. De Boer, Dag Alnæs, Pino Alonso, Celso Arango, Michal Assaf, Mihai Avram, Srinivas Balachander, Nerisa Banaj, Zeynep Başgöze, Marcelo C. Batistuzzo, Stephanie E.E.C. Bauduin, Francesco Benedetti, Sara Bertolin, Bianca Besteher, Laura Biagi, Robert J. Blair, Karina Blair, Sven Bölte, Stefan Borgwardt, Paolo Bosco, Paolo Brambilla, Beatrice Bravi, Brian P. Brennan, Willem B. Bruin, Geraldo F. Busatto, Murray J. Cairns, Sara Calderoni, Vince Calhoun, Rosa Calvo, Marta Cano, Vaughan J. Carr, Sean P. Carruthers, Georgia F. Caruana, Xavier Caseras, Stanley V. Catts, I-Jou Chi, Derin Cobia, Federica Colombo, Maria Beatriz Couto, Benedicto Crespo-Facorro, Kathryn R. Cullen, Udo Dannlowski, Mirella Dapretto, Adriana Di Martino, Gretchen J. Diefenbach, Annemiek Dols, Fabio Duran, Nadza Dzinalija, Christine Ecker, Stefan Ehrlich, Goi Khia Eng, Damien A. Fair, Afonso Fernandes, Jamie D. Feusner, Gregory A. Fonzo, Paola Fuentes-Claramonte, Nadine Gaab, Beata R. Godlewska, Benjamin I. Goldstein, Ali Saffet Gonul, Ian H. Gotlib, Hans J. Grabe, Melissa J. Green, Dominik Grotegerd, Oliver Gruber, Patricia Gruner, Abha R. Gupta, Ruben C. Gur, Raquel E. Gur, Shlomi Haar, Jarold P. Hamilton, Unn K. Haukvik, Frans A. Henskens, Asli C. Hinc, Yoshiyuki Hirano, Hao Hu, Matthew E. Hughes, Felice Iasevoli, Yanghee Im, Jonathan Ipser, Hammza Jabbar Abdl Sattar Hamoudi, Allison Jack, Delfina Janiri, Joost Janssen, Fern Jaspers-Fayer, Kyle M. Jensen, Jingwen Jin, Stefan Kaiser, Toshiharu Kamishikiryo, Melody J. Y. Kang, Andriana Karuk, Norbert Kathmann, Kody G. Kennedy, Minah Kim, Joseph A. King, Tilo Kircher, Anna Luisa Klahn, Daniel N. Klein, Kathrin Koch, Peter Kochunov, Azadeh Kushki, Jun Soo Kwon, Marilyn T. Lake, Mikael Landén, Luisa Lazaro, Irina Lebedeva, Nabulsi Leila, Meng Li, Christine Lochner, Carmel M. Loughland, Beatriz Luna, Karl Lundin Remnélius, Bradley J. MacIntosh, Matteo Mancini, Gisele G. Manfro, Rachel Marsh, Ignacio Martinez-Zalacain, David Mataix-Cols, Colm McDonald, Jane McGrath, Jose M. Menchon, Pedro Morgado, Bryan J. Mowry, Lilianne R. Mujica-Parodi, Emma Muñoz, Filippo Muratori, Declan Murphy, Benson Mwangi, Janardhanan C. Narayanaswamy, Jin Narumoto, Stener Nerland, Janina Neufeld, Benjamin T. Newman, Jared A. Nielsen, Erika L. Nurmi, Joseph O’Neill, Kirsten M. OHearn, Go Okada, Bob Oranje, Christos Pantelis, Nadine Parker, Kevin A. Pelphrey, Mary L. Phillips, John Piacentini, Maria Picó-Pérez, Rosanne Picotin, Alessandro Pigoni, Fabrizio Piras, Federica Piras, Edith Pomarol-Clotet, Giuseppe Pontillo, Daniel Porta-Casteràs, Maria J. Portella, Rebecca B. Price, Yann Quidé, Joaquim Radua, Elysha Ringin, Elena Rodriguez-Cano, Jaroslav Rokicki, Rafael Romero-Garcia, Susan Rossell, Hanyang Ruan, Katya Rubia, Matthew D. Sacchet, Yuki Sakai, Raymond Salvador, Gabriele Sani, Joao R. Sato, André Schmidt, Rodney J. Scott, Carl M. Sellgren, Lukas Sempach, Eiji Shimizu, Venkataram Shivakumar, Kang Sim, Jair C. Soares, Noam Soreni, Carles Soriano-Mas, Nuno Sousa, Frederike Stein, Jonas L. Steinhäuser-Meerz, Emily R. Stern, Thomas Straube, Jeffrey R. Strawn, Philip J. Sumner, Ibrahim Sungur, Philip R. Szeszko, Kristiina Tammimies, Alexander S. Tomyshev, Michela Tosetti, Laurens A. van de Mortel, John D. van Horn, Helena van Nieuwenhuizen, Tamsyn E. van Rheenen, Guido van Wingen, Daniela Vecchio, Ganesan Venkatasubramanian, Eduard Vieta, Enric Vilajosana, Yolanda Vives-Gilabert, Henry Völzke, Chris Vriend, Gregory L. Wallace, Zhen Wang, Martin Walter, Lei Wang, Sara Jane Webb, Lars T. Westlye, Sarah Whittle, Mark O. Wielpütz, Katharina Wittfeld, Will Woods, Mon-Ju Wu, Tony T. Yang, Lakshmi N. Yatham, Tokiko Yoshida, Abe Yoshinari, Je-Yeon Yun, Qing Zhao, Giovana B. Zunta-Soares, ENIGMA Autism Working Group, ENIGMA Anxiety Working Group, ENIGMA Bipolar Disorder Working Group, ENIGMA Major Depression Working Group, ENIGMA OCD Working Group, ENIGMA Schizophrenia Working Group, Odile A. van den Heuvel, Lianne Schmaal, Elena Pozzi, Ole A. Andreassen, Christopher R. K. Ching, Katherine E. Lawrence, Gaon S. Kim, Jan K. Buitelaar, Theo G.M. van Erp, Dan J. Stein, Daniel S. Pine, Anderson M. Winkler, Janna Marie Bas-Hoogendam, Andre Zugman, Nic J.A. van der Wee, Nynke A. Groenewold, Andre Marquand, Boris C. Bernhardt, Neda Jahanshad, Tyler M. Moore, Paul M. Thompson, Sophia I. Thomopoulos, Simon B. Eickhoff, Matthias Kirschner, Theodore D. Satterthwaite, Sofie L. Valk

## Abstract

Elucidating the neurobiological basis of neurodevelopmental and psychiatric conditions (NDPCs) remains challenging because brain alterations vary within diagnoses and overlap across them. Whether diverse alterations follow a systematic organization that may reflect shared vulnerabilities remains unknown. Here, we assembled 10,135 individuals with schizophrenia, autism, bipolar, obsessive-compulsive, generalized anxiety, and major depressive disorders, and 11,998 reference participants across six continents through the ENIGMA consortium. Using normative modeling, we quantified individual deviations in cortical thickness, surface area, and subcortical volumes relative to lifespan reference trajectories (5 to 80 years). We show that structural deviations converged along cortical axes reflecting connectome organization, maturation, and cytoarchitectonic diversity. These axes mirrored typical population variation, but their expression differed across diagnoses and partly scaled with symptom severity. Even rare and highly individualized extreme deviations followed this organization, concentrating in densely connected regions. Finally, brain structural deviations overlapped substantially across diagnoses, while differences between them increased toward the association cortex. Together, we provide large-scale evidence that structural deviations across NDPCs are systematically constrained by the brain’s intrinsic architecture. This shared organization provides a framework for reconciling individual variability with transdiagnostic similarities and motivates an integrative, systems-level understanding of mental health.

## Introduction

Nearly half of the global population will receive a mental health diagnosis during their lifetime^1^. A central paradox in understanding the neurobiological basis of neurodevelopmental and psychiatric conditions (NDPCs) is that individuals with the same diagnosis may show divergent brain alterations^2,3^, while different diagnoses share overlapping neurobiological patterns^4–7^. Moreover, psychiatric diagnoses frequently co-occur, with the presence of one psychiatric diagnosis substantially increasing the likelihood of receiving another across the lifespan^8,9^. Such co-occurrence is mirrored in shared genetic and environmental risk factors, overlapping alterations in brain structure and function, and symptoms that are present across diagnostic boundaries^7,10–15^. At the same time, substantial clinical and phenotypic heterogeneity exists within diagnostic groups^2,16^. Diagnostic categories thus provide only a coarse scaffold for elucidating the neurobiological mechanisms underlying mental illness. Dimensional frameworks such as the Hierarchical Taxonomy of Psychopathology^14^ and the Research Domain Criteria^15^ address this challenge by conceptualizing psychopathology along continuous, partially overlapping spectra. Yet, the extent to which these spectra are anchored in shared underlying patterns of brain structural variation remains unknown. In this large-scale study, we investigated whether brain structural variation across individuals and diagnoses follows systematic principles embedded in the brain’s intrinsic organization.

The human brain undergoes continuous change across the lifespan, shaping the emergence and manifestation of NDPCs^17–19^. Recent advances in computational psychiatry have established normative modeling as a powerful framework for characterizing lifespan trajectories and individual variation along them^20,21^. Analogous to growth charts, normative models estimate expected trajectories of brain features from demographic predictors such as age and sex. Individual’s brain features can then be expressed as statistical deviations from these reference trajectories. By intrinsically accounting for age-dependent interactions with brain maturation and ageing, deviations can yield insights into atypical development, adaptation to environmental stressors or pathological processes underlying NDPCs^22–24^. Prior normative modeling studies indicate that brain structural deviations in NDPCs are subtle and heterogeneous across individuals and brain regions^2,25–27^. This regional heterogeneity raises the question of whether diverse patterns of brain alteration follow a common organizational principle, impacting common functions and underlying shared symptom expression^2,28^.

Previous research has identified large-scale axes that systematically organize the brain^29–32^. Organizational axes, such as the sensorimotor-association axis, are refined throughout development and capture regional variation in neurobiology, function and connectivity. Crucially, this hierarchical organization creates spatial variation in vulnerability to diverse alterations. Brain structural alterations may thus unfold in a systematic manner across regions that are inter-connected, share neurobiological features, or exhibit synchronized developmental trajectories^5,28,29,33,34^. For instance, transdiagnostic studies have identified heightened vulnerability of highly connected network hubs within the transmodal association cortex. These hubs are metabolically demanding and thus susceptible to physiological stress, mature relatively late, and exhibit pronounced inter-individual variability^29,33,35^. Similarly, spatiotemporal variation in risk gene expression and plasticity are thought to impact vulnerability patterns^6,32^. Such large-scale organization may give rise to a shared, underlying pattern along which graded brain alterations emerge across diagnoses, potentially offering a neurobiological substrate for dimensional models of psychopathology such as HiTOP and RDoC. Studying such system-level markers is critical to gain a neurobiologically grounded understanding of transdiagnostic risk, shared clinical profiles, and epidemiological co-occurrence.

Here, we identified shared, systematic patterns of brain structural alterations across individuals with schizophrenia, autism, bipolar disorder, obsessive-compulsive disorder, generalized anxiety disorder, and major depression. To accomplish this, we unified the largest psychiatric neuroimaging sample to date. This sample includes cortical thickness, surface area, and subcortical volume data from 10,135 diagnosed individuals and 11,998 reference participants scanned across more than 170 sites and six continents. Using normative modeling, we quantified individual-level deviations from expected brain structural lifespan trajectories. These deviations converged along shared cortical axes that recapitulated the brain’s intrinsic organization. Across complementary analytic approaches, identified cortical axes captured systematic variation in transdiagnostic overlap, within-diagnosis variability, networks of co-occurring extreme deviations, and individual differences in symptom severity. Our global collaboration as part of the Enhancing NeuroImaging Genetics through Meta-Analysis (ENIGMA) consortium^36^ enabled well-powered analyses that were demographically diverse and methodologically harmonized, providing a unique synthesis not previously achievable. Thus, as described below, we demonstrate that macroscale brain structural variation across major psychiatric diagnoses is systematically organized along principles that extend beyond traditional diagnostic categories.

## Results

### The transdiagnostic ENIGMA sample

This study includes data from 10,135 individuals diagnosed primarily with one of six neurodevelopmental and psychiatric conditions (NDPCs; schizophrenia [SCZ; *n*=2,753], autism spectrum diagnosis [ASD; *n*=1,556], bipolar disorder [BP; *n*=1,370], obsessive-compulsive disorder [OCD; *n*=1,775], generalized anxiety disorder [ANXG; *n*=765], and major depressive disorder [MDD; *n*=1,916]) and a reference cohort (RC) of 11,998 comparison participants without any psychiatric or neurological diagnosis. We studied individuals aged 5-80 years, including data from 172 scan sites (102 study groups; **Fig. 1A-B**). We included NDPCs that often co-occur, and share genetic or phenotypic features^5,6,9,10^. Please see Supplementary Material (**Supplementary Tables S1-S13**) for details on sample demographics, exclusion criteria, and scan site information.

**Figure 1.**
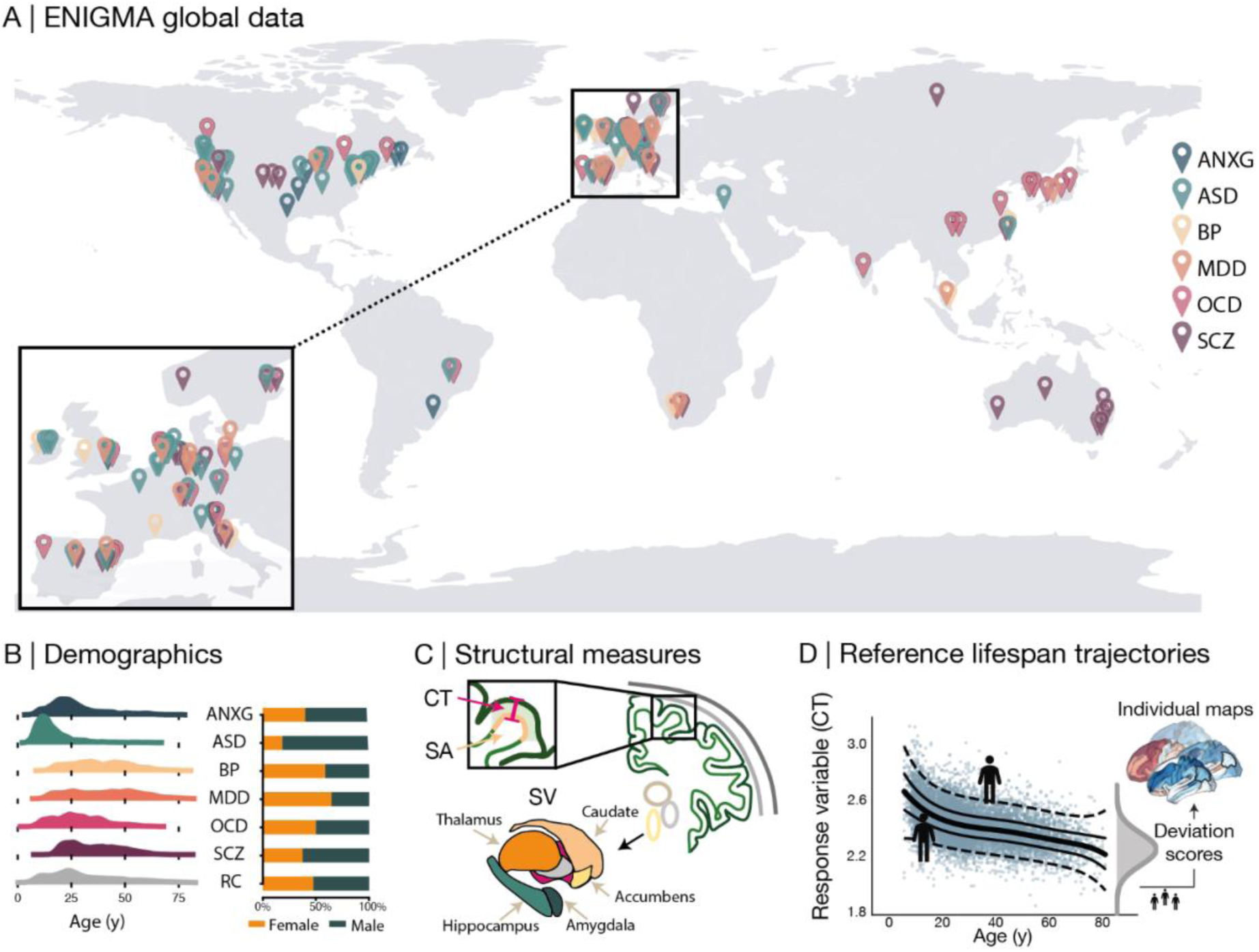
Aggregating a global, transdiagnostic dataset for a normative modeling framework. **A-C)** This study combines cortical thickness (CT), surface area (SA), and subcortical volume (SV) data from 10,132 individuals with a diagnosis of generalized anxiety (ANXG), autism spectrum diagnosis (ASD), bipolar (BP), major depressive (MDD), obsessive-compulsive (OCD), and schizophrenia spectrum (SCZ) disorders, and a reference cohort of *n*=11,998 between the ages 5 to 80y. **D)** We applied hierarchical Bayesian regression to derive an individual’s deviation from a lifespan reference trajectory for each brain region and measure.

### Normative modeling, group differences, and variability

We used FreeSurfer^37^-derived estimates of cortical thickness (CT), surface area (SA), and subcortical volumes (SV; **Fig. 1C**) for 68 cortical and 14 subcortical regions^38^. For each of these regions and metrics, we modeled lifespan trajectories in *n*=9,532 individuals without NDPCs (RC_train_; age range: 5 to 80 years; **Fig. 1D**; **Supplementary Table S14**). See **Supplementary Fig. S1** for model performance metrics and **Supplementary Table S15** and **Supplementary Fig. S2** for the assessment of site effects. Each individual’s brain structure was then expressed as a deviation from expected values, yielding regional deviation z-scores for individuals with NDPCs and a held-out RC_test_ cohort (*n*=2,466) set as a benchmark for unbiased group comparisons. All models are made publicly available under https://doi.org/10.5281/zenodo.21907827, allowing the community to derive deviation scores for their local cohorts.

Based on these individualized deviation scores, we first established group-level shifts between individuals with NDPCs and RCs (yielding Cohen’s *d* maps) as well as variability within diagnostic groups (**Fig. 2; Supplementary Fig. S3**). Group-shifts largely recapitulated spatial patterns reported in previous ENIGMA consortium publications^39^ (**Supplementary Tables S16-S20;** see **Supplementary Fig. S4-S8** for age-stratified analyses and interactions with age and sex). Interestingly, multiple NDPCs showed variability differences compared to RCs in CT and SV (*p*_FDR_<0.05) in more regions than they showed systematic shifts in group means (OCD, ASD, and ANXG for CT; ASD and ANXG for SA; ASD and OCD for SV; **Fig. 2B**; **Supplementary Tables S21-S23**). Overall, SCZ showed the largest effect sizes for group differences to RCs, whereas ASD showed the most pronounced inter-individual variability across metrics. See **Supplementary Fig. S9** and **Table S24** for alternative variability estimates. We further observed that spatial patterns of within-diagnosis variability were largely independent from their corresponding group-shift (i.e., Cohen’s *d*) maps, motivating the study of variability as a complementary dimension (all *p*_variogram_>0.05, see **Supplementary Table S25**).

**Figure 2.**
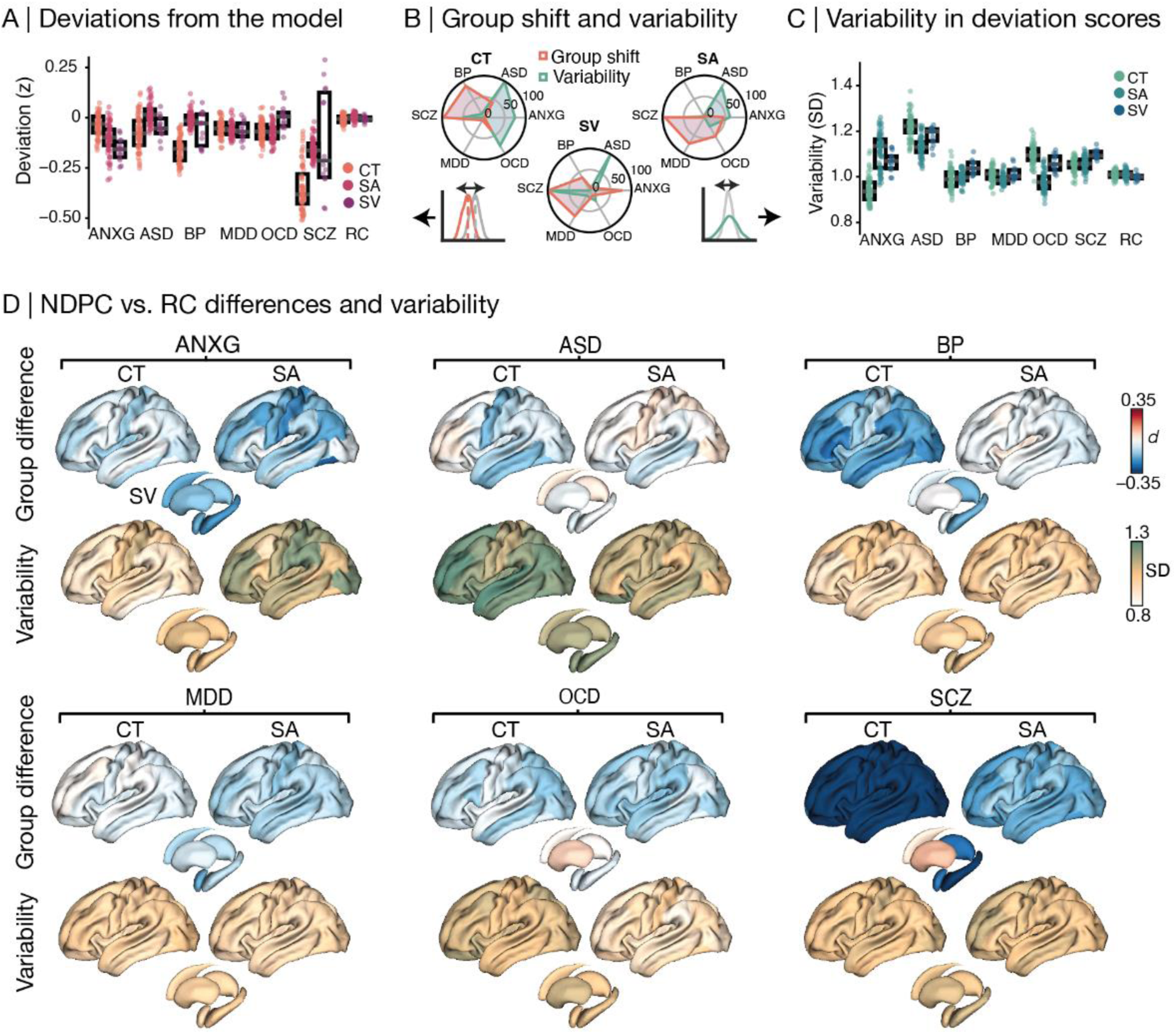
Group-shifts and variability in cortical thickness deviations. **A-C)** The six neurodevelopmental and psychiatric conditions (NDPCs) were differently characterized by group differences to the reference cohort and within-group variability (standard deviation). Data points in boxplots (**A** & **C**) depict regions, axes in radar plots (**B**) reflect percentages of regions with a significant group difference (NDPC>RC) in deviation strength (Cohen’s *d*; *p*_FDR_<0.05) or variability (Levene’s test, *p*_FDR_<0.05). **D)** Depicts spatial patterns of group differences and within-group variability per NDPC. Only left hemispheres are depicted to increase legibility, see **Supplementary Tables S16-18** and **S21-23,** and **Supplementary** Fig. 3 for both hemispheres. ANXG = generalized anxiety, ASD = autism spectrum diagnosis, BP = bipolar, MDD = major depressive, OCD = obsessive-compulsive, SCZ = schizophrenia spectrum disorders.

### Cortical axes capture co-organized deviations from the reference trajectory

Next, we investigated whether individual-level deviations were systematically co-organized, and whether these patterns reflect known principles of cortical organization. We first applied principal component analysis (PCA) to absolute CT deviation scores of the full NDPC and RC_test_ cohorts. Focusing on the magnitude of atypicality rather than the direction of deviations allowed us to capture the co-occurrence of deviations under the premise that both positive and negative deviations may be clinically relevant and can exist within the same individual. Deviations primarily covaried along a cortical axis that spanned from (para)limbic to dorsal heteromodal cortex (PC1; explaining 19.2% of the variance; **Fig. 3A**). A second axis contrasted unimodal (idiotypic) with ventral heteromodal/(para)limbic cortex (PC2; explaining 4.3% of the variance). Notably, these two axes differentially captured group-shifts and variability across NDPCs: while the magnitude of transdiagnostic group shifts predominantly varied along the paralimbic-to-heteromodal PC1 (*r*=0.65, *p*_variogram_<0.001; PC2: *r*=-0.31, *p*_variogram_=0.47), within-group variability was mostly structured along the unimodal-to-heteromodal PC2 (*r*=0.68, *p*_variogram_=0.035; PC1: *r*=0.04, *p*_variogram_=0.8; **Fig. 3B**). Axes of deviation were robust to leave-diagnoses-out and leave-sites-out robustness analyses (**Supplementary Table S26 & S27**), and observed across age groups as well as when leveraging signed deviation scores (**Supplementary Fig. S10 & S11A**).

**Figure 3.**
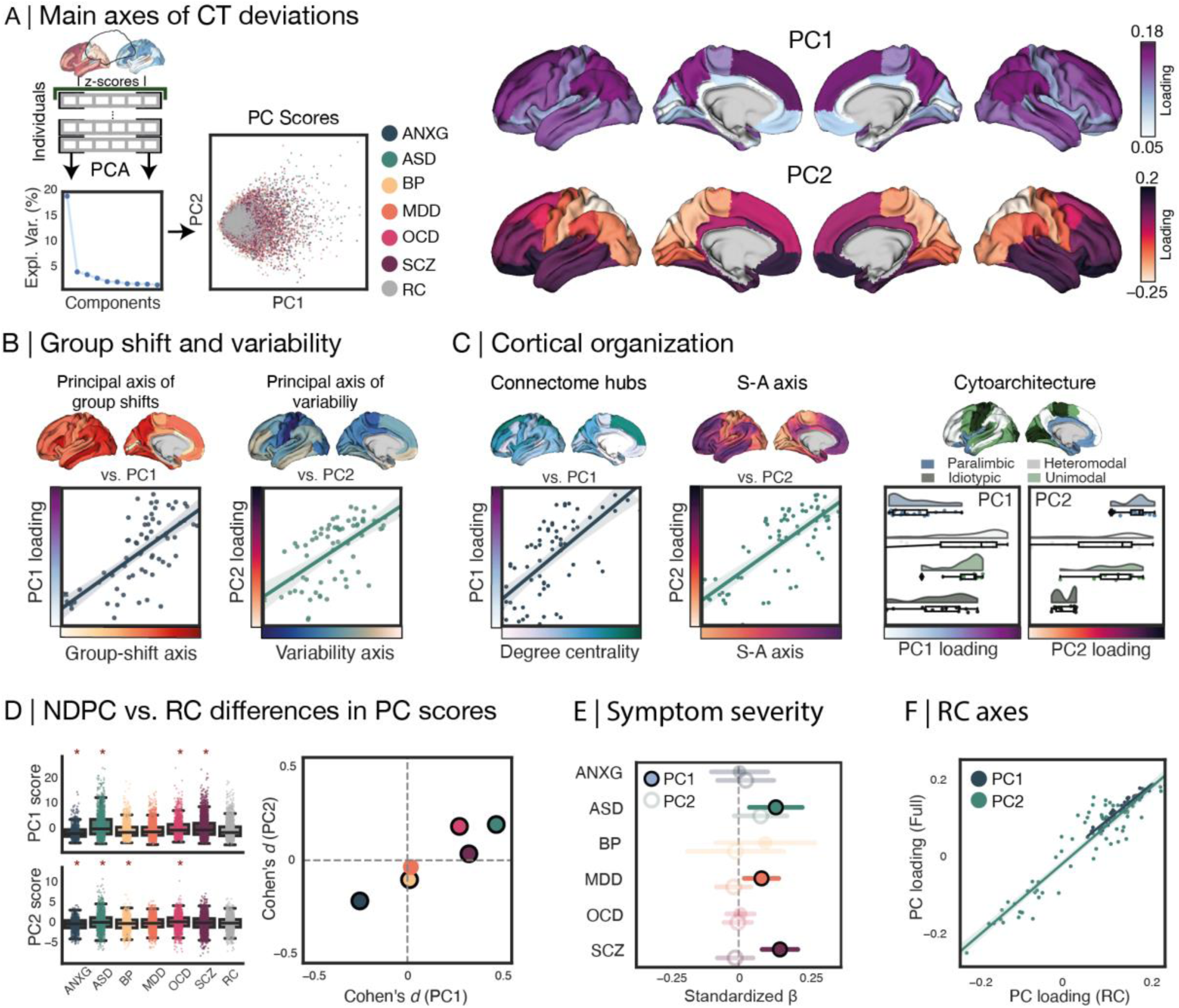
Principal axes of cortical thickness (CT) deviations. **A)** CT deviations are organized along two cortical axes as identified by a principal component analysis (PCA; left). **B)** CT axes differentially capture spatial patterns of transdiagnostic group shifts and within-diagnosis variability in deviation scores, which were identified by PCAs across Cohen’s *d* and variability maps, respectively. **C)** Left: The first axis (PC1) recapitulated the spatial distribution of normative connectome hubs (degree centrality, computed in 207 individuals from the Human Connectome Project – Young Adults cohort^40,41^). Middle: The second axis (PC2) further aligned with the sensorimotor-to-association axis^29^ (S-A axis) capturing the prominent hierarchical organization of the cortex. Right: PC loadings stratified across four cytoarchitectonic cortical types as defined by Mesulam^31^. **D)** Group differences in PC scores between individuals with and without a neurodevelopmental and psychiatric condition (NDPC). Left: Boxplot where each dot represents one individual, red stars indicate significant differences. Right: Cohen’s *d* values, framed in black if significant (*p*_FDR_<0.05). **E)** Associations between PC scores and symptom severity. Opacity indicates *p*_FDR_<0.05, horizontal lines indicate 95% confidence intervals. **F)** Association between PC loadings (i.e., spatial covariance patterns) derived solely in the reference cohort (RC) and the full sample including RCs and diagnosed individuals. * = *p*_FDR_<0.05; ANXG = generalized anxiety, ASD = autism spectrum diagnosis, BP = bipolar, MDD = major depressive, OCD = obsessive-compulsive, SCZ = schizophrenia spectrum disorders.

We then examined whether these large-scale deviation axes recapitulate the neurobiological organization of the cortex. To this end, we first identified densely connected network hubs in an independent cohort of healthy young adults (see Methods). Spatial correlations with the axes of deviations revealed that PC1, but not PC2, was significantly aligned with hubs of typical connectome organization. Specifically, PC1 highlighted differential CT deviations in densely connected hub regions compared to non-hub regions (PC1: *r* = 0.69, *p*_variogram_ <0.001; PC2: *r* = −0.33; *p*_variogram_ = 0.42; **Fig. 3C**). Furthermore, PC2 recapitulated a principal hierarchy of cortical organization and maturational timing, the sensorimotor-to-association axis^29^ (PC1: *r* = 0.11, *p*_variogram_ = 0.45; PC2: *r* = 0.70, *p*_variogram_ = 0.03).

### Individual variation along cortical axes

While CT deviations were largely structured along these shared cortical axes, we observed differences between diagnostic groups in terms of how strongly deviation patterns were expressed (**Fig. 3D**). For example, relative to individuals without NDPCs, ASD, SCZ, and OCD showed stronger deviations structured along PC1 towards the heteromodal pole (Cohen’s *d* range: 0.27 [OCD] to 0.47 [ASD]; *p*_FDR_<0.05), whereas ANXG showed an opposite trend (*d* = −0.25). At the same time, ASD and OCD further showed stronger variation along PC2 (*d* = 0.19 and 0.18, respectively; *p*_FDR_<0.05), whereas ANXG and BP diverged (*d* = −0.22 and −0.10, respectively; *p*_FDR_<0.05). We next investigated whether the expression of transdiagnostic axes at the individual level relates to symptom magnitude (**Supplementary Table S28**). In ASD, SCZ, and MDD cohorts, the magnitude of symptoms related to the respective diagnoses was positively associated with PC scores for the paralimbic-to-heteromodal PC1, indicating a stronger magnitude of deviations along this axis (standardized beta range: 0.08 [MDD] to 0.14 [SCZ]; *p*_FDR_<0.05; **Fig. 3E**). Together, these findings suggest that shared cortical axes capture subtle but meaningful variation across diagnoses and symptom severity, without implying that NDPC effects are confined to these axes.

Next, we asked to what extent axes of deviation observed across the full cohort reflect general principles of co-variation in the general population. PCA performed solely in the RC yielded highly similar cortical axes to those obtained in the full, transdiagnostic sample. Both the observed spatial patterns and differences between individuals with and without NDPCs were largely maintained when the axes were derived using only reference individuals without NDPCs (see **Fig. 3F** and **Supplementary Fig. S11B & C)**. Axes of CT deviations were highly correlated between the full and the RC samples (PC1: *r*=0.98, *p*_variogram_ <0.0001; PC2: *r*=0.86, *p*_variogram_<0.0001). Additional subspace analyses investigating whether NDPC-related alterations are embedded in typical structural covariance patterns are described in the **Supplementary Text**. Together, these analyses indicate that NDPC-related alterations largely unfold along pre-existing axes of typical population variation, while also retaining NDPC-specific structure beyond these dominant axes (particularly ASD).

To test whether these observations apply to other macrostructural brain measures, we expanded the analysis to SA and SV (see Supplementary Material). SA and SV findings support the notion that deviation patterns in NDPCs recapitulate coordinated co-variation observed in the general population (**Supplementary Fig. S12**). Individuals exhibiting stronger deviations along one axis (e.g., CT PC1) also tended to exhibit stronger deviations along axes of other structural features (e.g., SA PC1; see **Supplementary Fig. S13**). Finally, although age and sex influenced deviations from the reference trajectory in some NDPCs and regions (see **Supplementary Fig. S7 & S8**), these effects were largely independent from the PC-derived patterns overall, yet partly retained in ANXG (see **Supplementary Fig. S14**). See **Supplementary Tables S29 & S30** for investigations of time since diagnosis and medication effects.

### Extreme deviations from normative lifespan models are rare across the brain and heterogeneous across individuals

The cortical axes described above captured graded, subtle deviations shared across the full sample. We next asked whether this organization also extends to the rarer, particularly pronounced deviations within individuals. Specifically, we started by investigating how many individuals with NDPCs show deviations that exceed typical population variance outside the 2.5th-97.5th percentile (i.e., extreme deviations, *z* > 1.96 or *z* < −1.96, assessed separately). This approach also allowed us to determine how frequently extreme deviations occur in the same brain region across individuals sharing a diagnosis, thereby quantifying their spatial convergence or heterogeneity (**Fig. 4A** & **Supplementary Fig. S15-S17**). Notably, we found that at most 9.13% of individuals with the same diagnosis show converging extreme deviations. For example, extreme negative CT deviations in the same region were observed in maximally 4.22% of individuals with MDD (left isthmus of cingulate gyrus) and 9.13% of individuals with ASD (right inferior temporal gyrus). For extreme positive CT deviations, the maximum percentage of individuals sharing an extreme deviation in the same region ranged from 2.76% in the SCZ cohort (left pericalcarine cortex) to 9.13% in ASD (right rostral middle frontal gyrus).

**Figure 4.**
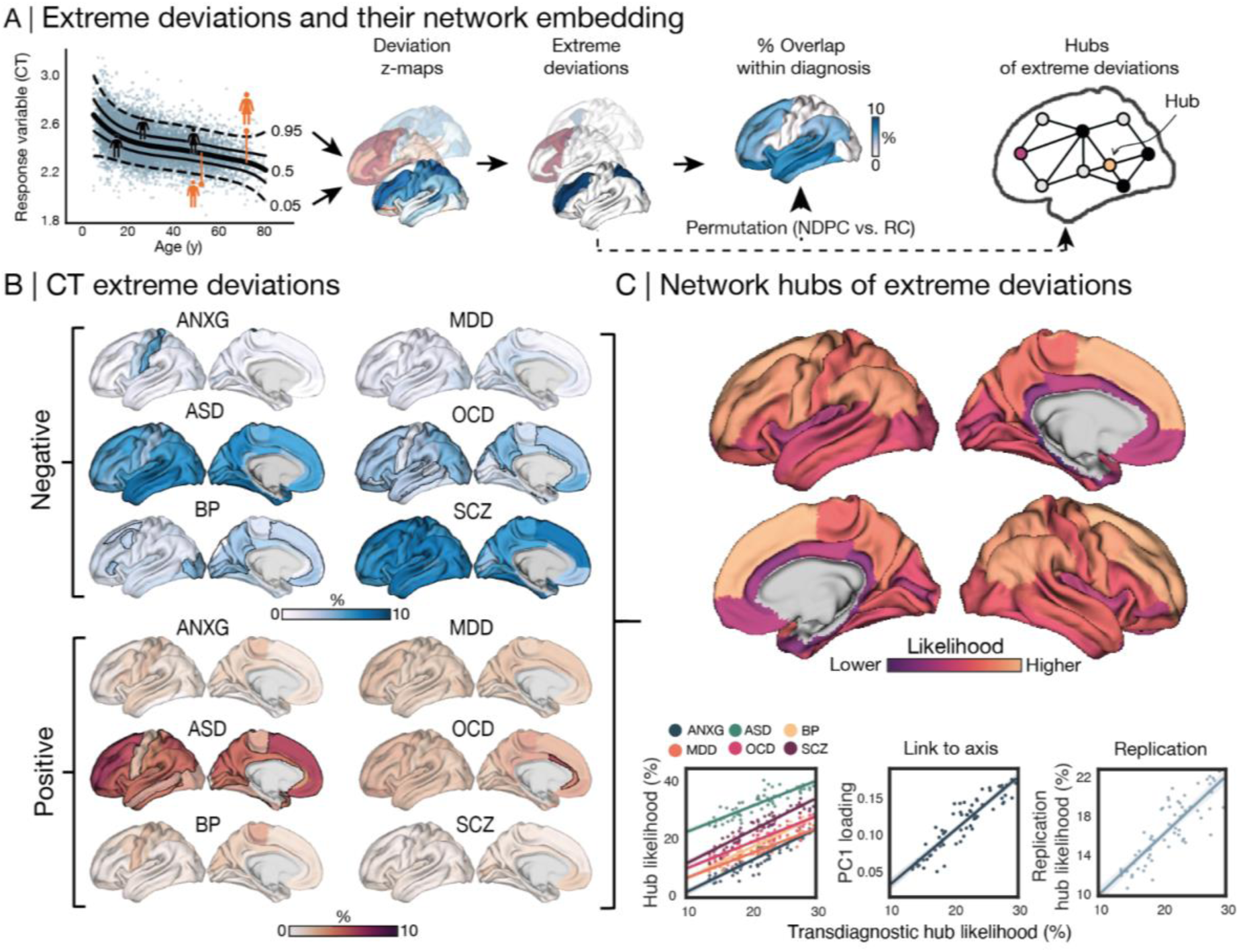
Extreme cortical thickness (CT) deviations and their shared network embedding. **A)** We defined values in the upper or lower 2.5th percentiles (|z| > 1.96) as extreme deviations and computed the percentage of individuals with the same diagnosis that show an extreme deviation in the same region. We then defined hubs of extreme deviations by asking ‘if one region shows an extreme deviation, how many of its most strongly connected regions also show an extreme deviation’ – using normative connectome data from the Human Connectome Project (HCP-YA^41^) as a reference connectome. **B)** Shows how often (%) individuals with the same diagnosis show significantly more extreme negative (blue) or positive (red) CT deviations in a given region than the reference cohort (10,000 permutations; *p*_FDR_<0.05; outlined in black). **C)** Depicts transdiagnostic hubs of extreme deviations. Bottom left: These hubs were consistent across diagnostic groups. Middle: The hub map was aligned with the observed principal axis of CT deviations (PC1; p<0.001). Right: The hub map could be replicated when leveraging the reference connectome of a second independent Cohort (Lausanne^42^). ANXG = generalized anxiety, ASD = autism spectrum diagnosis, BP = bipolar, MDD = major depressive, OCD = obsessive-compulsive, SCZ = schizophrenia spectrum disorders. RC = Reference cohort (here, RC_test_).

Comparing extreme deviation patterns of each NDPC cohort with RCs revealed greater regional accumulation of extreme negative deviations in all NDPCs except MDD (null models generated by shuffling group-labels 10,000 times; **Fig. 4B**). Greater regional accumulation of extreme positive deviations was only observed for ASD and OCD. NDPCs characterized by marked variability in our previous analyses again stood out (e.g., ASD, OCD, ANXG): individuals with the same diagnosis partly exhibited extreme deviations in opposite directions within the same regions, for example, in widespread heteromodal regions in ASD and the anterior cingulate cortex in OCD. This heterogeneity would be missed by traditional case-control contrasts. Of note, most individuals (both in RCs and NDPCs) showed at least one extreme deviation somewhere in the brain (see **Supplementary Fig. S18** & **Table S31**). Expanding this analysis to SA and SV further supported the notion that extreme deviations in macroscale brain structure are rare and heterogeneous at the regional level (see **Supplementary Fig. S15-S17**).

Despite being rare and heterogeneous, extreme deviations preferentially occurred in regions that also exhibited subtle group differences captured in Cohen’s *d* maps (see **Supplementary Tables S32).** Because these Cohen’s *d* patterns remained after excluding individuals with extreme deviations, extreme deviations are unlikely to be driving the observed group-level effects (see **Supplementary Table S33**). Rather, they appear to reflect the extreme end of a continuous distribution of structural variation present at the group-level.

### Extreme deviations are embedded in a transdiagnostic network reflecting typical connectome organization

Our findings thus far show that NDPC-related deviations converge along the normative organization of the cerebral cortex (**Fig. 3**), with extreme deviations representing particularly strong expressions of group-level tendencies (**Fig. 4**). We next investigated whether extreme deviations, despite being spatially heterogeneous, form a network of systematically co-occurring alterations. This notion builds on previous work suggesting that individuals with the same diagnosis share symptoms because their heterogeneous brain alterations impact the same networks^2,28^. We therefore investigated whether extreme deviations are more likely to co-occur in regions that are typically connected to each other. Specifically, we aimed to identify hubs where extreme deviations co-occur with extreme deviations in other parts of their network (‘co-alteration hub’)^15,17,23^. We leveraged structural and functional connectivity data from an unrelated subset of reference young adults of the Human Connectome Project (HCP-YA^40^) to derive a weighted reference connectome (see Methods). For each region and each participant, we asked: If region X shows an extreme deviation, how many of the regions that X is typically connected to also show extreme deviations? We found hubs of co-occurring extreme deviations, predominantly located in the dorsal prefrontal and parietal cortex (**Fig. 4C**). These hubs existed in each NDPC group, though their strength varied across groups (**Fig. 4C** bottom left). This convergence indicates a transdiagnostically shared network of extreme deviations. The likelihood of a region being a co-alteration hub increased gradually along the paralimbic-to-heteromodal axis of CT deviations (PC1: *r* = 0.91, *p*_variogram_ < 0.001) but not along PC2 (*r* = −0.46, *p*_variogram_ = 0.27). Alignment with PC1 is consistent with its correspondence to normative connectome organization described above (**Fig. 3C**). This hub pattern was replicated using a second independent reference connectome (**Fig. 4C** bottom right; *r* = 0.84, *p*_variogram_ < 0.001; See Supplementary Material for information on the Lausanne^42^ sample) and robust across age groups (see **Supplementary Figure S20**). Hubs of co-occurring extreme SA deviations were concentrated in the orbitofrontal cortex (see **Supplementary Fig. S19**). Collectively, these results indicate that extreme deviations are overall rare, yet systematically embedded within the brain’s typical connectome organization despite their heterogeneity.

### Brain structural deviations overlap substantially across diagnostic categories

So far, the observed systematic structure of brain structural deviations along shared axes and normative connectome organization suggests substantial brain phenotypic convergence across NDPCs. Previous meta-analytic studies have addressed such similarities by correlating group-level effect size maps^5,7,43,44^. Here, we leveraged our large-scale resource of individual-level data to quantify transdiagnostic convergence by measuring the distributional overlap of deviation scores across diagnoses (i.e., the shared area under two density functions; **Fig. 5A**). Overall, we observed pronounced similarities between NDPC cohorts, and between individuals with and without NDPCs (**Fig. 5B; see Supplementary Table S34** for an alternative similarity measure). CT distribution overlap was lowest between ANXG and SCZ (range: 79.2-96.3% across the cortex) and highest between MDD and OCD (range: 89.3-98.0%). All NDPC groups also showed strong overlaps with the RC_test_ sample (range: 79.8-96.6% for SCZ to 93.3-98.3% for MDD; see **Supplementary Table S35**). While overall overlap was high between NDPCs, it varied across the cortex (**Fig. 5A** – middle). Across all NDPC comparisons, we observed the highest transdiagnostic overlap in paralimbic, visual and motor areas, with more differentiation between NDPCs in the heteromodal cortex. Notably, this transdiagnostic overlap aligned with the first principal axis of CT deviations (*r*=-0.71, *p*_variogram_<0.001) but not with PC2 (*r*=-0.23, *p*_variogram_=0.162). The subtle differentiation of NDPCs in heteromodal cortex structure parallels the fact that these regions more frequently show NDPC vs. RC group shifts and extreme deviation hubs. However, the magnitude and direction of these effects vary across diagnoses.

**Figure 5.**
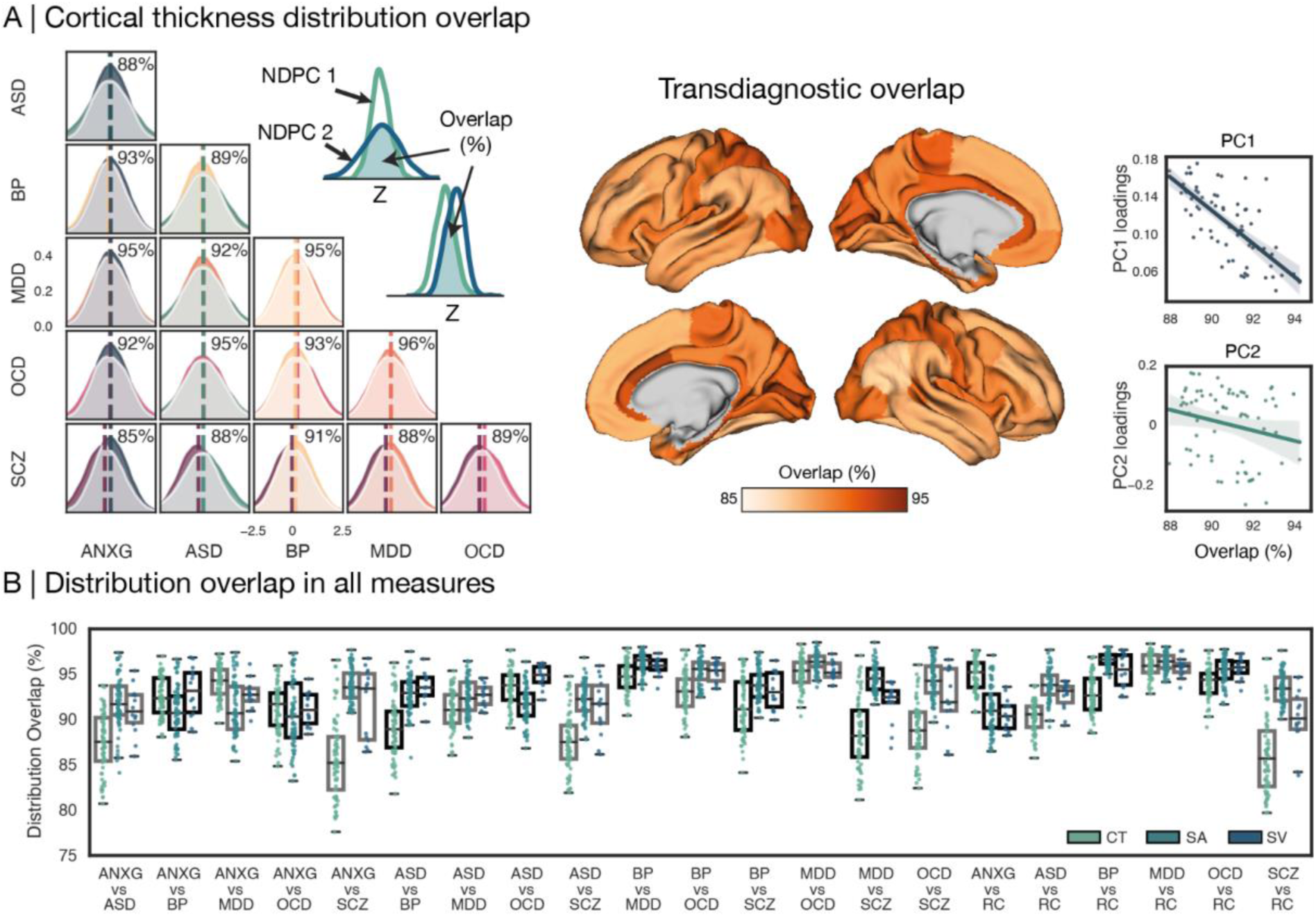
Overlapping distributions of brain structural deviations across neurodevelopmental and psychiatric conditions (NDPCs). **A)** The distribution overlap was computed as the shared area under two distributions. Left: Pair-wise distribution overlap in cortical thickness (CT) across individuals and cortical regions. Middle: Spatial variation in transdiagnostic CT overlap, computed as the average of all pair-wise comparisons per region. Right: Spatial association with the main axes of CT deviation, tested via Spearman’s rank order correlation. **B)** Boxplot depicting the pair-wise distribution overlap for CT, surface area (SA), and subcortical volume (SV). Each dot represents a brain region, boxes reflect the 25th - 75th percentile range with the median depicted by horizontal bars. ANXG = generalized anxiety disorder, ASD = autism spectrum diagnosis, BP = bipolar disorder, MDD = major depressive disorder, OCD = obsessive-compulsive disorder, SCZ = schizophrenia spectrum disorders. RC = Reference cohort (here, RC_test_).

Lastly, we observed that distribution overlap between NDPCs was even higher for SA and SV than for CT (see **Fig. 5B** and **Supplementary Fig. S21**). SA distribution overlap was lowest for ANXG and OCD (range: 83.8-97.3% across the cortex) and highest for MDD and OCD (range: 93.5-98.5%). SV distribution overlap was lowest between ANXG and ASD (range: 86.9-95.6% across subcortical structures) and highest for BP and MDD (range: 92.2-97.3%). Similarly, there was pronounced overlap with RCs (all NDPCs SA overlap > 85.7% and SV overlap > 88.6%). See **Supplementary Fig. S22** and **S23** for overlap results across age strata. Together, these findings demonstrate that macroscale brain structural alterations across NDPCs are highly overlapping and largely embedded within the broader landscape of typical brain variation, with diagnostic differentiation concentrated in the heteromodal cortex.

## Discussion

Brain structural variation in neurodevelopmental and psychiatric conditions (NDPCs) overlap across diagnoses yet remain heterogeneous within them. Here, we investigated whether diverse alterations follow a shared underlying structure that transcends diagnostic boundaries. To this end, we examined cortical thickness, surface area, and subcortical volumes in individuals with and without NDPCs relative to lifespan reference trajectories. Deviations in NDPCs reflected subtle and graded variation that was largely patterned along the brain’s intrinsic organization, rather than forming categorically distinct signatures. Accordingly, the cytoarchitectural, maturational, and connectomic architecture of the cortex appeared to constrain spatially co-occurring deviations in a systematic manner. Both transdiagnostic convergence and within-diagnosis variability shared a similar spatial structure. These patterns were more strongly expressed in several NDPCs at the group level, and scaled with symptom magnitude at the individual level. Together, current findings provide a systems-level framework for understanding shared patterns of vulnerability across diagnostic boundaries. These insights were gained in, to our knowledge, the largest and most geographically diverse psychiatric imaging cohort to date, increasing robustness to site- and population-specific biases.

Across a range of analytic approaches, we observed recurrent large-scale patterns of co-occurring brain structural deviations shared across six NDPCs. Previous studies have attributed such transdiagnostic similarities to shared genetic liability, overlapping environmental risk factors, lived experience, and also shared treatment effects^8,10,12,45^. Our findings suggest an additional perspective, namely that susceptibility and manifestation of psychopathological processes may be similarly expressed within the brain’s intrinsic organization. Consistent with this interpretation, recent work showed that statistical maps of CT differences across several NDPCs align with covariance patterns observed during typical maturation as a shared principle^46^. By deriving individual-level deviations from lifespan reference trajectories, we strengthen and extend these observations by showing that cortical organization constrains not only group-level alteration patterns, but also within-diagnosis heterogeneity, regional overlap, and the emergence of rare, network-embedded extreme deviations. Below, we propose two non-mutually exclusive mechanisms by which psychopathological variation may become anchored in the brain’s intrinsic architecture.

First, transdiagnostic alteration patterns may emerge because neurobiological properties associated with vulnerability are themselves systematically arranged within the cortical hierarchy^5,6,29,32,33,35,47^. We found that structural deviations recapitulate multiple dimensions of cortical organization which capture complementary aspects of brain structural vulnerability. For instance, axes of deviations segregated regions with cytoarchitectonically different laminar profiles, which in turn exhibit different plasticity levels across the lifespan^31,32^. Moreover, we found that strongly connected network hubs constrained the sub-networks in which both subtle and extreme deviations tended to co-occur, particularly at the heteromodal end of the paralimbic-to-heteromodal axis (CT PC1). Specifically, the dorsal prefrontal and parietal cortex, both members of the brain’s ‘rich club’ of densely interconnected network hubs^48^, showed coordinated extreme deviations. Previous work suggests that pathological alteration patterns recapitulate the metabolic demands and physiological stress of densely connected network hubs^33,35^. Their central position within the connectome embedding may both increase regional vulnerability related to metabolic stress and facilitate the synchronization of pathology across interconnected regions^33,35,49^. Moreover, previous meta-analytic approaches showed that co-occurring structural alterations across NDPCs not only preferentially arise in interconnected regions, but also scale with interregional similarity in gene expression, cytoarchitecture, and functional engagement^5,6,33,35,44,47^. Against this background, current findings from our global consortium support a framework in which the systematic spatial patterning of the cortical landscape constrains individual-level deviations across diagnostic boundaries.

Second, the progression of maturational programs across the cortex creates spatiotemporal variation in vulnerable developmental periods^29,34,50,51^. Cortical maturation unfolds asynchronously from early-maturing sensorimotor cortex to later-maturing heteromodal and paralimbic cortex, which retain plasticity well into adulthood. This protracted refinement is thought to support socioemotional maturation and learning, contributes to individual variability and provides an adaptive scaffold to refine network topology^52^. However, protracted malleability also allows atypical deviations in the timing and magnitude of neural system refinement to occur. Indeed, a broad range of NDPCs has been linked to disrupted developmental trajectories. Over 60% of individuals with an NDPC are diagnosed between ages 12 and 25^53^, demarcating adolescence as a transdiagnostic vulnerable period. Shared vulnerable periods and synchronized developmental aberrations may contribute to systematically structuring deviations across the cortex. Speculatively, the sensorimotor-association axis of maturational timing^29,54^ may underlie the unimodal-heteromodal (PC2) axis of deviations we observed, linking co-occurring deviations to synchronized plastic periods. Future longitudinal studies may address this question. Together, our findings demonstrate systematically interrelated brain structural alterations in regions that are not only highly connected and metabolically costly, but also developmentally protracted and plastic across the lifespan. These features may provide a common scaffold that channels transdiagnostic variation along pre-existing degrees of freedom.

Importantly, structural deviations largely fell within normative ranges, with substantial overlap among NDPCs and with RCs. Extending prior work that assessed transdiagnostic similarities via correlations of statistical maps^5,7,43,47^, our individual-level data revealed regional variation in transdiagnostic similarities. NDPCs exhibited a pronounced overlap of deviations, ranging from 79.2% to 98.5% across structural measures. However, we observed meaningful variation within these shared distributions, as greater symptom magnitude was associated transdiagnostically with stronger deviations along dominant axes of brain structural variation. This effect was independent of diagnosis and of the specific symptom domain assessed. Complementary to the mostly subtle deviations observed at the group-level, we observed extreme deviations that were rare across the cortex and heterogeneous across individuals. Yet, they exhibited a hub-like organization toward the heteromodal end of the paralimbic-heteromodal axis. These hubs indicate that even highly individualized extreme deviations are constrained by similar organizational principles. The rarity and individual heterogeneity of extreme deviations may limit their suitability as standalone biomarkers for psychiatric diagnoses. Our and previous findings^2,28^ instead suggest that their diagnostic and mechanistic relevance may emerge from their network context. Despite occurring in different regions across individuals, extreme deviations preferentially involved an overlapping set of inter-connected regions. This common network embedding may help reconcile pronounced individual heterogeneity with convergence within and across diagnoses. Previous work has further linked this notion to clinical phenomena: Spatially dispersed alterations may contribute to symptom diversity among individuals with the same diagnosis^2^. Their shared network embedding may in turn underlie common clinical features and transdiagnostic overlap. Overall, our findings indicate that NDPCs reflect graded variation within a shared neurobiological architecture. These findings thus offer a neurobiological account of lifespan co-occurrence and reinforce the conceptual shift from categorical boundaries toward continuous, system-level models of psychopathology.

Being the largest transdiagnostic lifespan modeling study to date, we also identified various avenues for improvement for future large-scale neuroimaging psychiatry research. First, this study is limited to the phenotypic data collected by the primary studies. For example, we lacked harmonized data on more detailed medication dosages, shared dimensional measures of symptom magnitude, genetic and environmental risk factors, and concurrent or lifetime comorbidities to study their interactions with present findings. Second, the cross-sectional nature of the study limits insights into the spatiotemporal spreading of coordinated structural deviations, limiting inference on individual-level progression. Third, the use of the Desikan-Killiany parcellation^38^ may have obscured regional heterogeneity and consequently biased transdiagnostic similarity by averaging across areas with distinct cytoarchitecture and vulnerability^29,55^. Fourth, hubs were defined using an independent connectome from young adults, whereas our normative models spanned the lifespan. Although network topology is broadly preserved across age^56^, age-related changes in connection strength may affect the likelihood of co-occurring structural alterations between two connected regions. Future studies should interrogate the relationship between macroscale anatomical and connectome architectural changes within the same participants. Finally, the definition of a reference cohort is a challenge in normative modeling. Here, we defined the reference based on individuals without known mental or neurological diagnoses. This operational definition does not exclude somatic conditions in the reference cohort, nor shall it imply that individuals with NDPCs are not part of the general population. Relatedly, clinical cohorts may underlie different study inclusion criteria. They may additionally exclude individuals whose symptom burden is too high to allow study participation, limiting the clinical spectrum captured in the sample. Last, sample sizes for symptom-severity analyses differed across diagnoses, resulting in variable statistical power across NDPCs. We therefore made our normative models publicly available to facilitate follow-up studies. These models enable comparable individual-level deviation scores to be derived in independent samples and additional diagnoses.

Taken together, our findings suggest that the neurobiology of diverse mental health diagnoses is better understood as systematic, graded variation within a shared organizational architecture than as a set of categorically distinct, diagnosis-specific signatures. Yet, diagnoses retain distinguishable degrees and directions of expression along this shared architecture. By showing that individual deviations converge along dominant patterns of cortical organization, we provide a principled framework for reconciling within-diagnosis variability with transdiagnostic similarities. More broadly, our results support a shift from a region- and diagnosis-centric perspective towards understanding mental health variation as being rooted in the brain’s intrinsic architecture. This systems-perspective motivates the study of shared mechanisms of risk and resilience and may inform integrative prevention strategies that transcend mental health diagnoses.

## Methods

### Participants

In a global collaborative effort, we aggregated data from 10,135 individuals with NDPCs and 11,998 comparison individuals (47.79% female; age: 30.61 +/- 15.70 y) without any psychiatric or neurodevelopmental diagnosis from 172 scan sites (102 study groups). The NDPC sample comprised individuals with schizophrenia spectrum disorder (SCZ; *n*=2753; 37.6% female; age: 35.29 +/-11.84 y), autism spectrum diagnosis (ASD; *n*=1556; 18.4% female; age: 16.03 +/-8.48 y), bipolar disorder (BP; *n*=1370; 58.7% female; age: 38.4 +/-13.2 y), obsessive-compulsive disorder (OCD; *n*=1775; 50.1% female; age: 28.2 +/- 11.5 y), generalized anxiety disorder (ANXG; *n*=765; 40.0 % female; age: 26.3 +/- 12.0 y), and major depressive disorders (MDD; *n*=1916; 64.8% female; age: 39.9 +/- 15.4 y). Our lifespan approach included individuals aged 5-80 years. This research complies with the ethical regulations as set by the Independent Research Ethics Committee at the Medical Faculty of the Heinrich Heine University Duesseldorf (study number 2018-317). The sites included in this study have received individual ethical approvals for collection and sharing of the data from their respective local ethics committees, and participants have given informed consent. See **Supplementary Tables S1-S13** for an overview of demographic characteristics and scan parameters for each site.

### Normative modeling

We used neuroimaging estimates of CT, SA, and SV, derived from the standard FreeSurfer pipeline^37^. These estimates were summarized in 68 cortical and 14 subcortical regions (Desikan atlas^38^). We trained a normative model for each of these regions and modalities using PCNtoolkit v1.0.0^57^. Normative models capture the mean and variance of a response variable (here: regional brain structural estimates) as a function of covariates (here: age, sex, and site) in a sufficiently large reference sample. We randomly sampled 80% of our comparison participants as our reference cohort (RC_train_; *n*=9532; 47.84% female; 30.77 +/- 15.76 years old; split within sites; see **Supplementary Table S14**) to establish normative lifespan trajectories. The held-out 20% of reference controls formed the test set (RC_test_; *n*=2466; 47.61% female; 30.01 +/- 15.45 years old). This test set allowed us to establish a reference benchmark for comparisons with NDPC groups without bias from overfitting to the reference cohort used to train the model.

In multi-site data like ours, study- and scanner-related variability introduces confounding effects on subsequent analyses. To account for such biases, we based our normative model on hierarchical Bayesian regression (HBR)^57,58^. HBR estimates signal and noise variance across sites by capturing mean and variance components separately for each site, while linking them through shared prior distributions. These shared priors assume that site and sex model parameters are drawn from a common distribution across sites, preventing overfitting on small batches for a specific site or sex. Moreover, we implemented HBR-SHASH, an extension of the HBR framework that flexibly models non-Gaussian data with heteroskedastic kurtosis and skewness and accommodates age-varying distribution shapes across the lifespan^57^. For all NDPCs and the RC_test_ set, we quantified individuals’ deviations from the expected reference curve, captured as z-scores^18,20,21^.

### Group differences and variability in deviation scores

As a first analytic step, we sought to recover, using our normative deviation scores, the group differences previously established by ENIGMA mega-analyses using raw structural values^59–64^. Mirroring ENIGMA mega-analyses, we computed Cohen’s *d* maps while sub-sampling a reference comparison group for each NDPC group via propensity score matching based on age and sex. For each NDPC separately, individuals from the RC_test_ cohort were selected based on their likelihood of being part of the NDPC group in terms of their age and sex (i.e., the propensity score derived from a logistic regression), thus optimizing the comparability of demographic distributions of NDPC and RC_test_ groups. We further studied intra-cohort variability among NDPCs by analyzing standard deviations (see **Supplementary Table S24** and **Figure S9** for other variability measures) and tested for group differences in variability using Levene’s test.

### Main axes of deviation

In order to identify systematic patterns of covarying structural deviations, we leveraged principal component analysis (PCA), identifying cortical axes of deviation from the normative model. We applied PCA to absolute deviation scores in the full cohort (NDPC cohorts and RC_test_) to capture which regions tend to show co-occurring deviations from the norm (in any direction). The rationale for using absolute scores here was to acknowledge that both positive and negative deviations from an expected norm can be interrelated phenomena with clinical consequences. NDPC vs. RC_test_ group differences in PC scores were computed via *t*-tests, using the same propensity score matching approach as described above to match RC groups. We used 50% of RCtest to fit the PCA and reserved the remaining 50% as an independent held-out comparison sample, allowing unbiased statistical comparisons between NDPC groups and typical variation. We further addressed the question of whether NDPC-related variation is embedded within the same low-dimensional space as typical inter-individual variation by again performing a PCA separately in the RC and the full sample. We then quantified the similarity between the resulting PC1-PC2 subspace using principal angles. Smaller angles indicate greater alignment of the underlying cortical variation, or little perturbation to typical patterns introduced by including individuals with NDPCs.

We then sought to determine whether regions that tend to show covarying group-shifts and variability are structured along similar axes as those capturing deviations from the normative trajectories in the full sample. To this end, we performed PCA on the Cohen’s *d* and variability maps separately, rescaling them to prevent NDPCs with larger effect sizes from dominating the patterns. The first principal component of each map was extracted to capture the transdiagnostic spatial organization of deviation magnitude and variability. We then examined the spatial relationship between these group-level deviation magnitude and variability axes and the axes of individual-level deviations using Spearman’s rank order correlations and variogram permutations^65^ (10,000 permutations).

### Contextualization of derived cortical axes

Focusing on CT axes in the main section of this work, we further contextualized derived patterns in relation to neurobiological principles of cortical organization. Specifically, we leveraged maps of connectome organization, the previously established sensorimotor-to-association (S-A) axis of cortical hierarchy^29^, and cytoarchitectonic differentiation described in the following section: first, to capture normative connectome organization, we accessed group-level structural (diffusion-weighted imaging; DWI) and functional (resting-state functional magnetic resonance imaging; rs-fMRI) connectivity data from an independent, healthy sample (Human Connectome Project; HCP^40^; *n* = 207, 83 males, mean age = 28.73 ± 3.73 years) via the ENIGMA Toolbox^39^. Please see Supplementary Material for further information on connectome processing. We generated a weighted connectome by multiplying structural and functional connectivity matrices, highlighting region pairs that are both structurally and functionally connected (as done previously, see^47^). The aim of combining multimodal connectomes was to acknowledge both anatomical networks and functional interactions. We then binarized the resulting matrix to keep each region’s 20% strongest connections. Based on this binary matrix, we computed normative degree centrality (hubs) as the sum of the strongest connections, assessing how often a region was a member of the top 20% connections of all other regions. Next, we accessed the S-A axis from previously published work^29^. Last, contextualization with cytoarchitecture was performed using Mesulam’s scheme of cortical laminar differentiation by stratifying CT axis loadings into idiotypic, unimodal, heteromodal, and paralimbic cortex^31,66^. Spatial associations between the CT axes, connectome hub, and S-A maps were assessed via Spearman’s correlations and tested for significance via variogram permutations, controlling for spatial auto-correlations (Brainsmash^65^; 10,000 permutations).

### Association of axes of deviations with symptom magnitude

The association between PC scores and individual symptom magnitude was computed separately for each diagnostic category, as different assessment tools were used for the different diagnoses. We selected the continuous total scores from the respective assessments, providing measures of overall symptom magnitude. We implemented general linear models to test the association between PC scores and symptom magnitude while controlling for age and sex. Anxiety symptoms were assessed in the ANXG cohort using the State-Trait Anxiety Inventory (STAI_T^67^; available for *n*=410). In the ASD cohort, we used the Calibrated Severity Score of the Autism Diagnostic Observation Schedule (ADOS-CSS^68^; available for *n*=491). OCD symptom magnitude was assessed based on the Yale-Brown Obsessive Compulsive Scale for adults (Y-BOCS^69^) and children (CY-BOCS), available for *n*=1,719. In both the SCZ and the BP cohorts, the Positive and Negative Symptom Scale (PANSS^70^; available for *n*=1,014 [SCZ] and 129 [BP]) was used to assess symptom magnitude. Last, MDD symptom magnitude was assessed based on the Hamilton Depression Rating Scale (HDRS; available for *n*=651).

### Heterogeneity of extreme deviations

Having characterized shared axes along which deviations vary across disorders, we investigated whether extreme deviations adhere to the same principles. To this end, we first examined what percentage of individuals show an extreme deviation (defined as |*z*|>1.96) in the same brain region. This yielded group-level maps estimated for negative and positive deviations separately^2,18,27,71^. NDPC vs. RC differences in extreme deviation percentages were then computed via non-parametric permutation tests, shuffling NDPC and RC labels 10,000 times (one-sided testing at *p*_FDR_<0.05). RC groups were matched to each NDPC group using propensity score matching in the same way as described for Cohen’s *d* map computation, taking age and sex into account. This analysis was performed for CT, SA, and SV.

### Network embedding of extreme deviations

After establishing extreme deviation percentage maps, we next asked whether extreme deviations, despite being spatially heterogeneous, co-occur in a shared network. To identify hubs of co-occurring extreme deviations, we again leveraged the normative, group-level weighted connectome derived from the HCP-YA cohort described above. For each brain region in each individual, we tested: if a region shows an extreme deviation (in any direction), how many of the regions that fall into its top 20% strongest connections also show an extreme deviation (in any direction). This yielded a ‘co-alteration hub likelihood map’, highlighting regions that are central to a network of co-occurring CT alterations within NDPC groups. We then averaged these maps first within diagnostic groups and then across them, to derive NDPC-specific and transdiagnostic hub maps. To address replicability, we repeated this analysis with a separate independent connectome dataset, using the same approach (Lausanne dataset^42^; *n* = 70 healthy young adults; 16 female, 25.3 ± 4.9 years; see Supplementary Material).

### Distribution overlap

Last, we sought to quantify how much individuals with different NDPCs overlap in terms of brain structural deviations. We specifically aimed to identify where in the brain this overlap is particularly high, as such regions are likely to be more relevant for future transdiagnostic investigations. We computed pairwise distribution overlap between each disorder pair, and with reference comparators, by quantifying distribution overlap^72,73^. The distribution overlap quantifies the shared area under two distributions using kernel density functions, thus providing an intuitive measure of similarities between two groups. As a sensitivity analysis, we also computed the Kolmogorov-Smirnov distance between pairwise distributions (see Supplementary Material). After computing the overlap for all NDPC pairs and for each brain region, we averaged overlap maps across all NDPC combinations to derive a transdiagnostic overlap map, highlighting spatial variation in transdiagnostic similarity.

## Author contributions

The authors confirm contribution to the paper as follows:

Study conception and design: M.D.H., S.L.V., T.S.

Data collection: D.A., P.A., C.A., Mi.A., Mih.A., Sr.B., N.B., Z.B., M.C.B., S.E.B., F.B., Sa.B., Bi.B., L.B., R.J.B., K.B., Sv.B., St.B., Pa.B., Pao.B., Be.B., B.P.B., W.B.B., G.F.B., M.J.C., S.C., V.C., R.C., Ma.C., V.J.C., S.P.C., G.C., X.C., S.V.C., I.C., D.C., F.C., Mar.C., B.C.-F., K.R.C., U.D., M.D., A.D.M., G.J.D., A.D., F.D., N.D., C.E., S.E., G.E., D.A.F., A.F., J.D.F., G.A.F., P.F.-C., G.N., B.R.G., B.I.G., A.G., I.H.G., H.J.G., M.J.G., D.G., O.G., P.G., A.R.G., R.C.G., R.E.G., S.H., J.P.H., U.K.H., F.A.H., A.C.H., Y.H., H.H., M.E.H., F.I., Y.I., J.I., H.J.A.S.H., A.J., D.J., Jo.J., F.J.-F., K.M.J., Ji.J., St.K., To.K., M.J.Y.K., An.K., N.K., K.G.K. Mi.K., J.A.K., Ti.K., Ann.K., D.N.K., K.K., P.K., Az.K., J.K., M.T.L., Mi.L., L.L., I.L., N.L., Me.L., C.L., C.M.L., B.L., K.L.R., Br.J.M., M.M., G.G.M., R.M., I.M.-Z., D.M.-C., C.M., J.M., J.M.M., P.M., Bry.J.M., L.R.M.-P., E.M., F.M., D.M., B.M., J.C.N., Ji.N., S.N., Ja.N., B.T.N., J.A.N., E.L.N., J.O., K.M.O., G.O., B.O., C.P., N.P., K.A.P., M.L.P., J.P., M.P.-P., R.P., A.P., Fa.P., Fe.P., E.P.-C., G.P., D.P.-C., M.J.P., R.B.P., Y.Q., Jo.R., E.R., E.R.-C., Ja.R., R.R.-G., S.R., H.R., K.R., M.D.S., Y.S., R.S., Ga.S., Jo.R.S., An.S., R.J.S., C.M.S., Lu.S., E.S., V.S., K.S., J.C.S., No.S., C.S.-M., Nu.S., Fr.S., J.L.S.-M., E.R.S., T.S., Je.R.S., P.J.S., I.S., P.R.S., K.T., A.S.T., M.T., L.A.V.D.M., J.D.V.H., H.V.N., T.E.V.R., G.V.W., D.V., G.V., Ed.V., En.V., Y.V.-G., H.V., C.V., G.L.W., Z.W., Ma.W., Le.W., Sa.W., La.W., Sar.W., M.O.W., K.W., W.W., Mo.W., T.T.Y., L.N.Y., T.Y., A.Y., J.Y., Q.Z., G.B.Z.-S.

Data Analysis and Software generation: M.D.H., S.L.V., A.S., A.A.A.d.B.

Draft paper preparation: M.D.H., S.L.V.

Draft paper revision: M.D.H., S.L.V., T.S.

All authors reviewed the results and approved the final version of the paper.

## Code availability

Custom code generated for this project was made publicly available under https://github.com/CNG-LAB/Enigmatics/. Normative models can be accessed via https://doi.org/10.5281/zenodo.21907827. This Github repository further contains instructions to transfer and apply the trained models to independent datasets. Our analysis code makes use of open software: Normative modeling was performed using the PCNToolkit (https://pcntoolkit.readthedocs.io). Brain visualizations were carried out using the ENIGMA Toolbox (https://enigma-toolbox.readthedocs.io) and Brainspace (https://brainspace.readthedocs.io/). Spatial permutations were performed using BrainSmash (https://brainsmash.readthedocs.io/).

## Data availability

Data generated for this study, as well as atlases used for contextualization, were made publicly available under Github https://github.com/CNG-LAB/Enigmatics/. Subject-level ENIGMA neuroimaging data is not publicly available but can be acquired by submitting a research proposal to the ENIGMA Working Groups (http://enigma.ini.usc.edu/). HCP group-level connectome data was accessed via the ENIGMA Toolbox (https://enigma-toolbox.readthedocs.io/en/latest/pages/05.HCP/index.html). Lausanne group-level connectome data is available at: https://zenodo.org/records/2872624#.XOJqE99fhmM. Processed developmental neuroimaging data from the Philadelphia Neurodevelopmental Cohort and the Healthy Brain Network included in this study can be accessed through the Reproducible Brain Charts initiative^74^.

## Acknowledgements

M.D.H. was supported by a Walter Benjamin Fellowship of the German Research Foundation (DFG; GZ: HE 10277/1-1), the Max Planck Society, and the German Federal Ministry of Education and Research (BMBF). Am.S. was supported by the Max Planck Society. Go.S. was supported by a postdoctoral fellowship from the Canadian Institutes of Health Research (CIHR). Ai.M. was supported by the Studienstiftung des deutschen Volkes and the Lise Meitner Excellence Program. P.A. was supported by Instituto de Salud Carlos III (ISCIII) grants PI22/00752 and PI25/01407 and Fundació La Marató TV3 grant 202201-30. C.A. was supported by the Spanish Ministry of Science and Innovation, Instituto de Salud Carlos III (ISCIII), co-financed by the European Union and European Regional Development Fund (ERDF), NextGenerationEU (PMP21/00051), grants PI19/01024 and PI22/01824, CIBERSAM, Madrid Regional Government (B2017/BMD-3740 AGES-CM-2), European Union Structural Funds, the European Union Seventh Framework Programme, the European Union H2020 Programme through the Innovative Medicines Initiative 2 Joint Undertaking (PRISM-2, grant 101034377; AIMS-2-TRIALS, grant 777394), Horizon Europe, the National Institute of Mental Health of the National Institutes of Health (1U01MH124639-01, ProNET; 5P50MH115846-03, FEP-CAUSAL), Fundación Familia Alonso, and Fundación Alicia Koplowitz. Mi.A. was supported by Hartford Hospital. Sr.B. was supported by a DHR-ICMR Young Medical Faculty PhD Grant 2024–25. M.C.B. was supported by the National Council for Scientific and Technological Development (CNPq; grant 310332/2025-7). Sa.B. was supported by grant CM21/00278, co-funded by the European Social Fund. L.B. and Pa.B. were supported by the Italian Ministry of Health (RC2025 L4). Sv.B. was supported by the Swedish Research Council, Vinnova, Formas, FORTE, Hjärnfonden, Stockholm Brain Institute, Autism and Asperger Association Stockholm, Queen Silvia Jubilee Fund, Solstickan Foundation, PRIMA Child and Adult Psychiatry, the Pediatric Research Foundation at Astrid Lindgren Children’s Hospital, the Swedish Foundation for Strategic Research, Jerring Foundation, the Swedish Order of Freemasons, Kempe-Carlgrenska Foundation, Sunnderdahls Handikappsfond, the Jeansson Foundation, and EU-AIMS, with support from the Innovative Medicines Initiative Joint Undertaking (grant 115300), comprising contributions from the European Union’s Seventh Framework Programme (FP7/2007–2013), European Federation of Pharmaceutical Industries and Associations companies, and Autism Speaks, as well as EU AIMS-2-TRIALS. M.J.C. was supported by the National Health and Medical Research Council (NHMRC). S.C. was supported by FIA-Fondazione Italiana Autismo and the Italian Ministry of Health (Ricerca Corrente 2026). V.C. was supported by NIH grant R01MH123610. R.C. was supported by Instituto de Salud Carlos III/Fondo de Investigaciones Sanitarias (PI09/1588), the European Regional Development Fund (FEDER), and Fundació La Marató-TV3 (091510). Ma.C. was supported by grant RYC2024-050082-I from the Spanish Ministry of Science, Innovation and Universities/State Research Agency (MICIU/AEI/10.13039/501100011033), co-funded by the European Social Fund Plus (FSE+). S.V.C. was supported by the NHMRC. D.C. was supported by T32 NS047987. B.C.-F. was supported by CIBERSAM G26. K.R.C. was supported by NIMH K23MH090421. U.D. was supported by the German Research Foundation (DFG; FOR2107 DA1151/5-1, DA1151/5-2, DA1151/9-1, DA1151/10-1, DA1151/11-1; SFB/TRR 393, project 521379614) and the Interdisciplinary Center for Clinical Research (IZKF) of the Medical Faculty of Münster (Dan3/016/26). M.D. was supported by NIMH R01MH117982 and NICHD P50HD055784. G.J.D. was supported by grant 129522 from the Hartford HealthCare Research Funding Initiative. N.D. acknowledges support from the International OCD Foundation Innovator Award 2021 awarded to Odile van den Heuvel and Chris Vriend. S.E. and J.A.K. were supported by the German Federal Ministry of Education and Research (BMBF) as part of the German Center for Child and Adolescent Health (DZKJ; 01GL2405B) and the ASD-Net research consortium (01EE1409A). Data associated with G.E. and E.R.S. were supported in part by NIMH/NIH grants R01MH126981, R01MH111794, and R33MH107589 awarded to E.R.S. J.D.F. was supported by R01MH085900. G.A.F. received salary support from the National Institute of Mental Health (R01MH132784, R01MH129694, R01MH125886) and philanthropic funding from the Effie and Wofford Cain Foundation. P.F.-C. was supported by a “la Caixa” Foundation Junior Leader Fellowship (LCF/BQ/PR22/11920017). G.N. was supported by R01HD065762. B.R.G. and K.R. were supported by the Medical Research Council. B.I.G. was supported by the Canadian Institutes of Health Research. I.H.G. was supported by NIMH R37MH101495. M.J.G., F.A.H., R.J.S., and S.R. acknowledge support from the NHMRC; F.A.H. additionally acknowledges support from the Pratt Foundation. P.G. was supported by K23MH115206. A.R.G. was supported by NIMH R01MH100028 through the Autism Center of Excellence Network. Y.H. was supported by the AMED Brain/MINDS Beyond Program (JP18dm0307002) and JSPS KAKENHI grants JP19K03309, JP22H01090, JP23K07004, JP23K22361, JP23K02956, JP24K21493, JP24K06547, JP25K06842, JP25H01085, and JP25K00879. Y.I., M.J.Y.K., and C.R.K.C. acknowledge support from R21MH139001, R01MH129742, R01MH131806, R01AG058854, and R01MH134962, as well as the Office of the Director, National Institutes of Health, under award S10OD032285. The content is solely the responsibility of the authors and does not necessarily represent the official views of the NIH. A.J. was supported by NIMH R01MH100028. St.K. was supported by the Swiss National Science Foundation (grants 169783 and 140351). To.K. acknowledges support for the Hiroshima cohort from the Japan Agency for Medical Research and Development (AMED; JP24wm0625204). N.K. was supported by the German Research Foundation (DFG). K.G.K. acknowledges support from the Ontario Mental Health Foundation and Canadian Institutes of Health Research grant MOP 136947 awarded to B.I.G. Mi.K. was supported by the Brain Science Convergence Research Program (RS-2023-00266120), Basic Science Research Program (RS-2026-25470841), and Basic Research Program (25-BR-05-05) of the Korea Brain Research Institute (KBRI), funded by the Ministry of Science and ICT. Ti.K. was supported in part by German Research Foundation consortium grants FOR 2107 and SFB/TRR 393 (“Trajectories of Affective Disorders”; 521379614), Germany’s Excellence Strategy (EXC 3066/1 “The Adaptive Mind”; 533717223), the DYNAMIC Center through the LOEWE program of the Hessian Ministry of Science and Arts (LOEWE1/16/519/03/09.001(0009)/98), and the Wellcome Trust-funded DIALOG consortium (314138/Z/24/Z). D.N.K. was supported by NIMH grant R01MH069942. Mi.L. acknowledges support for the St. Göran Bipolar Study from the Swedish Research Council (2022-01643), Hjärnfonden/Swedish Brain Foundation (FO2025-0004-HK-212), and Swedish Government under the LUA/ALF agreement (ALFGBG-1005343). L.L. and E.M. were supported by Fundació La Marató (project 091810); L.L. additionally acknowledges the Carlos III Health Institute (PI11/01419). C.L. was supported by the South African Medical Research Council. B.L. was supported by the Staunton Farm Foundation. G.G.M. was supported by CNPq. R.M. was supported by NIMH R21MH101441 and R01MH104648. D.M.-C. was supported by the Wellcome Trust and a pump-priming grant from the South London and Maudsley Trust (064846). C.M. acknowledges support for the NUIG sample from the Health Research Board (HRA_POR/2011/100). J.M.M. was supported by Instituto de Salud Carlos III (PI22/00956). P.M. was supported by national funds through the Foundation for Science and Technology (FCT), projects UID/06304/2025 and LA/P/0050/2020. L.R.M.-P. was supported by the Baszucki Foundation. D.M. acknowledges support from EU-AIMS/AIMS-2-TRIALS through the Innovative Medicines Initiative Joint Undertaking (grant 777394). Ja.N. was supported by Riksbankens Jubileumsfond. B.T.N. was supported by NIH R01MH100028. J.A.N. was supported by R01MH101486-04. J.O. was supported by NIMH R01MH085900 and R01MH081864. G.O. was supported by AMED grant JP24wm0625204. C.P. acknowledges that the Australian Schizophrenia Research Bank was supported by the NHMRC (Enabling Grant 386500), Pratt Foundation, Ramsay Health Care, Viertel Charitable Foundation, and Schizophrenia Research Institute. K.A.P. was supported by R01MH100028. J.P. was supported by 1R01MH081864. M.P.-P. was supported by grant RYC2021-031228-I from the Spanish Ministerio de Ciencia e Innovación (MCIN/AEI/10.13039/501100011033) and the European Union NextGenerationEU/PRTR. D.P.-C. was supported by the Spanish Ministry of Science, Innovation and Universities, State Research Agency, and European Union-NextGenerationEU/PRTR (JDC2023-0507775-I). R.R.-G. was supported by the Plan de Generación de Conocimiento (PID2021-122853OA-I00), Plan de Consolidación (CNS2023-143647) of the Agencia Estatal de Investigación, ERANET Neuron JTC 2023 (ERP-2023-23684211), and Consejería de Universidad, Investigación e Innovación, Junta de Andalucía (DGP_PIDI_2024_00089). Y.S. was supported by AMED grants JP23wm0625001 and JP24wm0625204. Jo.R.S. was supported by FAPESP. C.M.S. was supported by the Swedish Brain Foundation (2022-0357), Swedish Scientific Council (2023-02827), Swedish Society for Medical Research (CG-23-0298-B), and Swedish Federal Government under the LUA/ALF agreement. E.S. was supported by the AMED Brain/MINDS Beyond Program (JP18dm0307002). K.S. was supported by a Singapore Bioimaging Consortium research grant (RP C009/2006). J.C.S. was partially supported by NIMH 1R01MH085667-01A1, the John S. Dunn Foundation, and the Pat Rutherford Chair in Psychiatry at UTHealth Houston. C.S.-M. was supported by the Ministry of Science and Innovation, Spain (PID2022-139081OB-C22), and Marató de TV3 (202201-31). Fr.S. was supported by the German Research Foundation (DFG; STE3301/1-1, project 527712970), the Von Behring-Röntgen Society (72_0013), and the DFG Collaborative Research Centre/Transregio 393 (CRC/TRR 393; 521379614). M.T. was supported by Italian Ministry of Health grant RC 2026. J.D.V.H. was supported by 5R01MH100028 and 1S10OD038249. T.E.V.R. was supported by an Al and Val Rosenstrauss Fellowship from the Rebecca L. Cooper Medical Research Foundation. G.V. was supported by a Wellcome-DBT India Alliance grant (500236/Z/11/Z). Ed.V. acknowledges support from the Spanish Ministry of Science and Innovation (PI21/00787), Instituto de Salud Carlos III and FEDER, Secretaria d’Universitats i Recerca del Departament d’Economia i Coneixement (2021-SGR-01358), CERCA Programme, Generalitat de Catalunya, La Marató-TV3 Foundation (202234-30), European Union Horizon 2020 (grant 945151), Horizon Europe (grant 101057454), and EIT Health (EDIT-B). H.V. acknowledges that SHIP is part of the Community Medicine Research Network of University Medicine Greifswald, supported by the German Federal State of Mecklenburg-West Pomerania. Le.W. was supported by P50MH071616, R01MH056584, R01MH084803, and U01MH097435. Sa.W. was supported by R01MH10028. Sar.W. was supported by the Brain and Behavior Research Foundation. K.W. acknowledges support for SHIP from the German Federal Ministry of Education and Research (01ZZ9603, 01ZZ0103, 01ZZ0403), the Ministry of Cultural Affairs and Social Ministry of Mecklenburg-West Pomerania, and support for MRI scans in SHIP and SHIP-TREND from Siemens Healthineers and the Federal State of Mecklenburg-West Pomerania. W.W. was supported by the Australian National Imaging Facility. T.T.Y. was supported by the National Center for Complementary and Integrative Health (NCCIH; R61AT009864 and R33AT009864). L.N.Y. acknowledges that STOP-EM was funded by an unrestricted investigator-initiated grant from AstraZeneca. A.Y. was supported by the Japan Society for the Promotion of Science (KAKENHI 23K14825). O.A.V.D.H. was supported by R01MH138569. J.K.B. was supported by the EU-AIMS and AIMS-2-TRIALS programmes through Innovative Medicines Initiative Joint Undertaking grants 115300 and 777394, with contributions from the European Union’s FP7 and Horizon 2020 programmes, EFPIA companies, Autism Speaks, Autistica, and SFARI, and by the Horizon 2020-supported CANDY (847818) and R2D2 (101057385) programmes. The funders had no role in study design; data collection, analysis, or interpretation; manuscript preparation; or the decision to publish. T.G.V.E. was supported by R21MH097196. D.S.P. and A.Z. acknowledge support from Project ZIA-MH002781 in the National Institute of Mental Health Intramural Research Program. This research was supported in part by the Intramural Research Program of the National Institutes of Health; the contributions of NIH authors are considered works of the United States Government, and the findings and conclusions do not necessarily reflect the views of the NIH or the U.S. Department of Health and Human Services. A.M.W. was supported by NIH grants U54HG013247, R01MH138425, and R01MH139547. J.B.-H. was supported by a NWO Rubicon grant (Dutch Research Council 019.201SG.022), a Talent Acceleration grant from Medical Delta, and NeurolabNL (NWA.1418.22.025). An.M. was supported by European Research Council Consolidator Grant 101001118. N.J. was supported by R01MH134004. P.M.T. was supported by NIH grants R01MH134962, R01NS107513, U01MH136221, R01AG060610, and R01MH129742. The ENIGMA Working Groups acknowledge the NIH Big Data to Knowledge (BD2K) award for foundational support and consortium development (U54EB020403 to P.M.T.). S.I.T. was supported by NIH grants R01MH138569 and R01MH129742. S.B.E. was supported by the Max Planck School of Cognition. Ma.K. acknowledges funding from the Swiss National Science Foundation (P2SKP3_178175 and 32003B_219240). S.L.V. was supported by the Max Planck Society through the Lise Meitner Excellence Program, the Jacobs Foundation Research Fellowship, the Hector Research Career Development Award, and ERC Starting Grant “Social Connections” (101220063). SHIP is part of the Community Medicine Research net of the University of Greifswald, Germany, which is funded by the Federal Ministry of Education and Research (grants no. 01ZZ9603, 01ZZ0103, and 01ZZ0403), the Ministry of Cultural Affairs and the Social Ministry of the Federal State of Mecklenburg-West Pomerania. MRI scans in SHIP and SHIP-TREND have been supported by a joint grant from Siemens Healthineers, Erlangen, Germany and the Federal State of Mecklenburg-West Pomerania. We thank Anika Schlorhaufer for administrative support in preparing this manuscript, as well as additional members of the ENIGMA consortium, including Pedro Rosa and Sarah Durston. We would further like to acknowledge the late Professor Dan J. Stein for his longstanding leadership and collaboration within ENIGMA Working Groups and for his invaluable contributions to psychiatry, neuroscience, and mental health research. His intellectual generosity, mentorship, and unwavering support shaped this work and the broader field in lasting ways. His scientific legacy and his kindness to colleagues continue to inspire us.

## References

1. McGrath, J. J. et al. Age of onset and cumulative risk of mental disorders: a cross-national analysis of population surveys from 29 countries. Lancet Psychiatry 10, 668–681 (2023).

2. Segal, A. et al. Regional, circuit and network heterogeneity of brain abnormalities in psychiatric disorders. Nat. Neurosci. 26, 1–17 (2023).

3. Segal, A. et al. Embracing variability in the search for biological mechanisms of psychiatric illness. Trends Cogn. Sci. 0, (2024).

4. Goodkind, M. et al. Identification of a common neurobiological substrate for mental illness. JAMA Psychiatry 72, 305–315 (2015).

5. Hettwer, M. D. et al. Coordinated cortical thickness alterations across six neurodevelopmental and psychiatric disorders. Nat. Commun. 13, 6851 (2022).

6. Patel, Y. et al. Virtual Histology of Cortical Thickness and Shared Neurobiology in 6 Psychiatric Disorders. JAMA Psychiatry 78, 47 (2021).

7. Radonjic, N. V. et al. Structural brain imaging studies offer clues about the effects of the shared genetic etiology among neuropsychiatric disorders. Mol Psychiatry 26, 2101–2110 (2021).

8. Caspi, A. et al. Longitudinal Assessment of Mental Health Disorders and Comorbidities Across 4 Decades Among Participants in the Dunedin Birth Cohort Study. JAMA Netw. Open 3, e203221 (2020).

9. Plana-Ripoll, O. et al. Exploring comorbidity within mental disorders among a Danish national population. JAMA Psychiatry 76, 259–270 (2019).

10. Lee, P. H. et al. Genomic Relationships, Novel Loci, and Pleiotropic Mechanisms across Eight Psychiatric Disorders. Cell 179, 1469–1482.e11 (2019).

11. Newson, J. J., Pastukh, V. & Thiagarajan, T. C. Poor Separation of Clinical Symptom Profiles by DSM-5 Disorder Criteria. Front. Psychiatry 12, (2021).

12. McLaughlin, K. A., Colich, N. L., Rodman, A. M. & Weissman, D. G. Mechanisms linking childhood trauma exposure and psychopathology: a transdiagnostic model of risk and resilience. BMC Med. 18, 96 (2020).

13. Grotzinger, A. D. et al. Mapping the genetic landscape across 14 psychiatric disorders. Nature 649, 406–415 (2026).

14. Kotov, R. et al. The Hierarchical Taxonomy of Psychopathology (HiTOP): A dimensional alternative to traditional nosologies. J. Abnorm. Psychol. 126, 454–477 (2017).

15. Insel, T. et al. Research Domain Criteria (RDoC): Toward a New Classification Framework for Research on Mental Disorders. Am. J. Psychiatry 167, 748–751 (2010).

16. Young, G., Lareau, C. & Pierre, B. One quintillion ways to have PTSD comorbidity: Recommendations for the disordered DSM-5. Psychol. Inj. Law 7, 61–74 (2014).

17. Parkes, L. et al. Transdiagnostic dimensions of psychopathology explain individuals’ unique deviations from normative neurodevelopment in brain structure. Transl. Psychiatry 11, 1–13 (2021).

18. Rutherford, S. et al. Charting brain growth and aging at high spatial precision. eLife 11, e72904 (2022).

19. Ballester, P. L. et al. Brain age in mood and psychotic disorders: a systematic review and meta-analysis. Acta Psychiatr. Scand. 145, 42–55 (2022).

20. Marquand, A. F. et al. Conceptualizing mental disorders as deviations from normative functioning. Mol. Psychiatry 24, 1415–1424 (2019).

21. Rutherford, S. et al. The normative modeling framework for computational psychiatry. Nat. Protoc. 17, 1711–1734 (2022).

22. Paus, T., Keshavan, M. & Giedd, J. N. Why do many psychiatric disorders emerge during adolescence? Nat. Rev. Neurosci. 9, 947–957 (2008).

23. Dell’Osso, L., Lorenzi, P. & Carpita, B. The neurodevelopmental continuum towards a neurodevelopmental gradient hypothesis. J. Psychopathol. 25, 179–182 (2019).

24. Hallgrimsson, B. et al. The developmental-genetics of canalization. Semin. Cell Dev. Biol. 88, 67–79 (2019).

25. Sun, X. et al. Mapping Neurophysiological Subtypes of Major Depressive Disorder Using Normative Models of the Functional Connectome. Biol. Psychiatry 94, 936–947 (2023).

26. Wolfers, T. et al. Individual differences v. the average patient: mapping the heterogeneity in ADHD using normative models. Psychol. Med. 50, 314–323 (2020).

27. Wolfers, T. et al. Replicating extensive brain structural heterogeneity in individuals with schizophrenia and bipolar disorder. Hum. Brain Mapp. 42, 2546–2555 (2021).

28. Cash, R. F. H., Müller, V. I., Fitzgerald, P. B., Eickhoff, S. B. & Zalesky, A. Altered brain activity in unipolar depression unveiled using connectomics. Nat. Ment. Health 1, 174–185 (2023).

29. Sydnor, V. J. et al. Neurodevelopment of the association cortices: Patterns, mechanisms, and implications for psychopathology. Neuron 109, 2820–2846 (2021).

30. Huntenburg, J. M., Bazin, P.-L. & Margulies, D. S. Large-Scale Gradients in Human Cortical Organization. Trends Cogn. Sci. 22, 21–31 (2018).

31. Mesulam, M. M. From sensation to cognition. Brain J. Neurol. 121 (Pt 6), 1013–1052 (1998).

32. García-Cabezas, M. Á., Zikopoulos, B. & Barbas, H. The Structural Model: a theory linking connections, plasticity, pathology, development and evolution of the cerebral cortex. Brain Struct. Funct. 224, 985–1008 (2019).

33. Fornito, A., Zalesky, A. & Breakspear, M. The connectomics of brain disorders. Nat. Rev. Neurosci. 16, 159–172 (2015).

34. Sydnor, V. J. et al. Investigating hierarchical critical periods in human neurodevelopment. Neuropsychopharmacology 1–19 (2025) doi:10.1038/s41386-025-02246-5.

35. Vanasse, T. J. et al. Brain pathology recapitulates physiology: A network meta-analysis. Commun. Biol. 4, 301 (2021).

36. Thompson, P. M. et al. The ENIGMA Consortium: large-scale collaborative analyses of neuroimaging and genetic data. Brain Imaging Behav. 8, 153–182 (2014).

37. Fischl, B. FreeSurfer. NeuroImage 62, 774–781 (2012).

38. Desikan, R. S. et al. An automated labeling system for subdividing the human cerebral cortex on MRI scans into gyral based regions of interest. Neuroimage 31, 968–980 (2006).

39. Larivière, S. et al. The ENIGMA Toolbox: multiscale neural contextualization of multisite neuroimaging datasets. Nat. Methods 18, 698–700 (2021).

40. Glasser, M. F. et al. The minimal preprocessing pipelines for the Human Connectome Project. Neuroimage 80, 105–124 (2013).

41. Van Essen, D. C. et al. The Human Connectome Project: A data acquisition perspective. Neuroimage 62, 2222–2231 (2012).

42. Griffa, A., Aleman-Gomez, Y. & Hagmann, P. Structural and functional connectome from 70 young healthy adults. https://zenodo.org/records/2872624 (2019) doi: https://zenodo.org/record/2872624.

43. Opel, N. et al. Cross-Disorder Analysis of Brain Structural Abnormalities in Six Major Psychiatric Disorders: A Secondary Analysis of Mega- and Meta-analytical Findings From the ENIGMA Consortium. Biol. Psychiatry 88, 678–686 (2020).

44. Park, B. et al. Multiscale neural gradients reflect transdiagnostic effects of major psychiatric conditions on cortical morphology. Commun. Biol. 5, 1–14 (2022).

45. Kumar, K. et al. Cortical differences across psychiatric disorders and associated common and rare genetic variants. 2025.04.16.25325971 Preprint at 10.1101/2025.04.16.25325971 (2025).

46. Cao, Z. et al. Cortical profiles of numerous psychiatric disorders and normal development share a common pattern. Mol. Psychiatry 28, 698–709 (2023).

47. Hansen, J. Y. et al. Local molecular and global connectomic contributions to cross-disorder cortical abnormalities. Nat. Commun. 13, 4682 (2022).

48. van den Heuvel, M. P., Kahn, R. S., Goñi, J. & Sporns, O. High-cost, high-capacity backbone for global brain communication. Proc. Natl. Acad. Sci. U. S. A. 109, 11372–11377 (2012).

49. Raj, A. & Powell, F. Models of Network Spread and Network Degeneration in Brain Disorders. Biol. Psychiatry Cogn. Neurosci. Neuroimaging 3, 788–797 (2018).

50. Baum, G. L. et al. Development of structure–function coupling in human brain networks during youth. Proc. Natl. Acad. Sci. 117, 771–778 (2020).

51. Paquola, C. et al. Shifts in myeloarchitecture characterise adolescent development of cortical gradients. eLife 8, (2019).

52. Fakhar, K. & Astle, D. E. Embracing the suboptimal organization of the human brain. Trends Cogn. Sci. 10.1016/j.tics.2026.04.008 (2026) doi:10.1016/j.tics.2026.04.008.

53. Solmi, M. et al. Age at onset of mental disorders worldwide: large-scale meta-analysis of 192 epidemiological studies. Mol. Psychiatry 27, 281–295 (2022).

54. Sydnor, V. J. et al. Intrinsic activity development unfolds along a sensorimotor–association cortical axis in youth. Nat. Neurosci. 26, 638–649 (2023).

55. Paquola, C. et al. The architecture of the human default mode network explored through cytoarchitecture, wiring and signal flow. Nat. Neurosci. 28, 654–664 (2025).

56. Riedel, L., van den Heuvel, M. P. & Markett, S. Trajectory of rich club properties in structural brain networks. Hum. Brain Mapp. 43, 4239–4253 (2022).

57. de Boer, A. A. A. et al. Non-Gaussian normative modelling with hierarchical Bayesian regression. Imaging Neurosci. 2, imag–2–00132 (2024).

58. Kia, S. M. et al. Hierarchical Bayesian Regression for Multi-site Normative Modeling of Neuroimaging Data. in Medical Image Computing and Computer Assisted Intervention – MICCAI 2020 (eds Martel, A. L. et al.) 699–709 (Springer International Publishing, Cham, 2020). doi:10.1007/978-3-030-59728-3_68.

59. Boedhoe, P. S. W. et al. Cortical Abnormalities Associated With Pediatric and Adult Obsessive-Compulsive Disorder: Findings From the ENIGMA Obsessive-Compulsive Disorder Working Group. Am J Psychiatry 175, 453–462 (2018).

60. Hibar, D. P. et al. Cortical abnormalities in bipolar disorder: an MRI analysis of 6503 individuals from the ENIGMA Bipolar Disorder Working Group. Mol Psychiatry 23, 932–942 (2018).

61. Schmaal, L. et al. Cortical abnormalities in adults and adolescents with major depression based on brain scans from 20 cohorts worldwide in the ENIGMA Major Depressive Disorder Working Group. Mol Psychiatry 22, 900–909 (2017).

62. van Erp, T. G. M. et al. Cortical Brain Abnormalities in 4474 Individuals With Schizophrenia and 5098 Control Subjects via the Enhancing Neuro Imaging Genetics Through Meta Analysis (ENIGMA) Consortium. Biol Psychiatry 84, 644–654 (2018).

63. van Rooij, D. et al. Cortical and Subcortical Brain Morphometry Differences Between Patients With Autism Spectrum Disorder and Healthy Individuals Across the Lifespan: Results From the ENIGMA ASD Working Group. Am J Psychiatry 175, 359–369 (2018).

64. Harrewijn, A. et al. Cortical and subcortical brain structure in generalized anxiety disorder: findings from 28 research sites in the ENIGMA-Anxiety Working Group. Transl. Psychiatry 11, 502 (2021).

65. Burt, J. B., Helmer, M., Shinn, M., Anticevic, A. & Murray, J. D. Generative modeling of brain maps with spatial autocorrelation. NeuroImage 220, 117038 (2020).

66. Paquola, C. et al. A multi-scale cortical wiring space links cellular architecture and functional dynamics in the human brain. PLOS Biol. 18, e3000979 (2020).

67. Spielberger, C. D., Gonzalez-Reigosa, F., Martinez-Urrutia, A., Natalicio, L. F. S. & Natalicio, D. S. The State-Trait Anxiety Inventory. Rev. Interam. Psicol. J. Psychol. 5, (2017).

68. Lord, C. et al. Autism Diagnostic Observation Schedule: ADOS-2.

69. Goodman, W. K. et al. The Yale-Brown Obsessive Compulsive Scale: I. Development, Use, and Reliability. Arch. Gen. Psychiatry 46, 1006–1011 (1989).

70. Kay, S. R., Fiszbein, A. & Opler, L. A. The Positive and Negative Syndrome Scale (PANSS) for Schizophrenia. Schizophr. Bull. 13, 261–276 (1987).

71. Segal, A. et al. Multiscale Heterogeneity of White Matter Morphometry in Psychiatric Disorders. Biol. Psychiatry Cogn. Neurosci. Neuroimaging 10.1016/j.bpsc.2025.03.014 (2025) doi:10.1016/j.bpsc.2025.03.014.

72. ENIGMA Clinical High Risk for Psychosis Working Group. Normative Modeling of Brain Morphometry in Clinical High Risk for Psychosis. JAMA Psychiatry 81, 77–88 (2024).

73. Winter, N. R. et al. Quantifying Deviations of Brain Structure and Function in Major Depressive Disorder Across Neuroimaging Modalities. JAMA Psychiatry 79, 879–888 (2022).

74. Shafiei, G. et al. Reproducible Brain Charts: An open data resource for mapping brain development and its associations with mental health. Neuron 113, 3758–3779.e6 (2025).

