## Supplementary Material for "Lifespan brain structural variation reveals shared organization across mental health conditions"

Meike D. Hettwer[1,2,3,4], Amin Saberi[1,3,5], Golia Shafiei[2,4,6], Aikaterina Manoli[1,3,7], Augustijn A. De Boer[8,9], Dag Alnæs[10,11], Pino Alonso[12,13,14,15], Celso Arango[15,16,17], Michal Assaf[18,19], Mihai Avram[20], Srinivas Balachander[21], Nerisa Banaj[22], Zeynep Başgöze[23,24], Marcelo C. Batistuzzo[25,26], Stephanie E.E.C. Bauduin[27], Francesco Benedetti[28,29], Sara Bertolin[13,14,30], Bianca Besteher[31,32], Laura Biagi[33], Robert J. Blair[34,35], Karina Blair[36], Sven Bölte[37,38,39], Stefan Borgwardt[20], Paolo Bosco[33], Paolo Brambilla[40,41], Beatrice Bravi[42], Brian P. Brennan[43], Willem B. Bruin[44,45,46], Geraldo F. Busatto[47], Murray J. Cairns[48,49], Sara Calderoni[33,50], Vince Calhoun[51], Rosa Calvo[52,53,54,55], Marta Cano[15,56], Vaughan J. Carr[57,58], Sean P. Carruthers[59], Georgia F. Caruana[60], Xavier Caseras[61], Stanley V. Catts[62], I-Jou Chi[63,64], Derin Cobia[65], Federica Colombo[28], Maria Beatriz Couto[66,67], Benedicto Crespo-Facorro[15,68,69], Kathryn R. Cullen[70], Udo Dannlowski[32,71,72,73], Mirella Dapretto[74,75,76], Adriana Di Martino[77], Gretchen J. Diefenbach[19,78], Annemiek Dols[44,79], Fabio Duran[80], Nadza Dzinalija[81,82], Christine Ecker[83], Stefan Ehrlich[84], Goi Khia Eng[85,86], Damien A. Fair[24,87,88], Afonso Fernandes[66], Jamie D. Feusner[37,89,90], Gregory A. Fonzo[91], Paola Fuentes-Claramonte[30,92], Nadine Gaab[93,94], Beata R. Godlewska[95,96], Benjamin I. Goldstein[90], Ali Saffet Gonul[97], Ian H. Gotlib[98], Hans J. Grabe[99,100], Melissa J. Green[57], Dominik Grotegerd[71], Oliver Gruber[101], Patricia Gruner[19], Abha R. Gupta[102,103,104], Ruben C. Gur[4,105], Raquel E. Gur[4,105], Shlomi Haar[106], Jarold P. Hamilton[107], Unn K. Haukvik[108,109], Frans A. Henskens[110,111], Asli C. Hinc[97,112], Yoshiyuki Hirano[113,114], Hao Hu[115], Matthew E. Hughes[59,116], Felice Iasevoli[117], Yanghee Im[118], Jonathan Ipser[119], Hammza Jabbar Abdl Sattar Hamoudi[120], Allison Jack[121], Delfina Janiri[122,123], Joost Janssen[124], Fern Jaspers-Fayer[125], Kyle M. Jensen[126], Jingwen Jin[127], Stefan Kaiser[128], Toshiharu Kamishikiryo[129], Melody J. Y. Kang[118], Andriana Karuk[30,92], Norbert Kathmann[130], Kody G. Kennedy[131], Minah Kim[132,133,134], Joseph A. King[84], Tilo Kircher[135], Anna Luisa Klahn[136], Daniel N. Klein[137], Kathrin Koch[138], Peter Kochunov[139], Azadeh Kushki[89,140], Jun Soo Kwon[141,142], Marilyn T. Lake[119,143], Mikael Landén[136,144], Luisa Lazaro[15,52,53,54], Irina Lebedeva[145], Nabulsi Leila[118], Meng Li[31], Christine Lochner[146], Carmel M. Loughland[147], Beatriz Luna[148,149], Karl Lundin Remnélius[37,150], Bradley J. MacIntosh[151,152], Matteo Mancini[153,154,155], Gisele G. Manfro[156,157], Rachel Marsh[158,159], Ignacio Martinez-Zalacain[12,160], David Mataix-Cols[161], Colm McDonald[162], Jane McGrath[163], Jose M. Menchon[12,14,30], Pedro Morgado[66,67,164], Bryan J. Mowry[165,166], Lilianne R. Mujica-Parodi[167,168], Emma Muñoz[169], Filippo Muratori[33], Declan Murphy[170,171], Benson Mwangi[172], Janardhanan C. Narayanaswamy[173,174,175], Jin Narumoto[176], Stener Nerland[177,178], Janina Neufeld[37,179], Benjamin T. Newman[180,181], Jared A. Nielsen[65], Erika L. Nurmi[182], Joseph O`Neill[183,184], Kirsten M. OHearn[185], Go Okada[129], Bob Oranje[186], Christos Pantelis[60,187,188], Nadine Parker[10], Kevin A. Pelphrey[189], Mary L. Phillips[190], John Piacentini[76], Maria Picó-Pérez[66,191], Rosanne Picotin[101], Alessandro Pigoni[192], Fabrizio Piras[22], Federica Piras[22], Edith Pomarol-Clotet[30,92], Giuseppe Pontillo[117], Daniel Porta-Casteràs[56], Maria J. Portella[15,56,193], Rebecca B. Price[194], Yann Quidé[195,196], Joaquim Radua[197,198], Elysha Ringin[60], Elena Rodriguez-Cano[30,199], Jaroslav Rokicki[200,201], Rafael Romero-Garcia[202,203], Susan Rossell[204,205], Hanyang Ruan[138,206], Katya Rubia[207], Matthew D. Sacchet[208], Yuki Sakai[209,210,211], Raymond Salvador[30,92], Gabriele Sani[122,123], Joao R. Sato[212], André Schmidt[213], Rodney J. Scott[49,147,214], Carl M. Sellgren[161,215], Lukas Sempach[213,216], Eiji Shimizu[113,114], Venkataram Shivakumar[217], Kang Sim[218,219,220], Jair C. Soares[172], Noam Soreni[221,222], Carles Soriano-Mas[13,223,224], Nuno Sousa[66,225,226,227], Frederike Stein[135], Jonas L. Steinhäuser-Meerz[228,229,230], Emily R. Stern[85,86,231], Thomas Straube[232], Jeffrey R. Strawn[233], Philip J. Sumner[59], Ibrahim Sungur[97], Philip R. Szeszko[234,235], Kristiina Tammimies[37,236], Alexander S. Tomyshev[145], Michela Tosetti[33], Laurens A. van de Mortel[44,45], John D. van Horn[180,237], Helena van Nieuwenhuizen[137], Tamsyn E. van Rheenen[59,60], Guido van Wingen[44,45], Daniela Vecchio[238], Ganesan Venkatasubramanian[21], Eduard Vieta[197,239], Enric Vilajosana[52,54,240], Yolanda Vives-Gilabert[241], Henry Völzke[242], Chris Vriend[82,243,244], Gregory L. Wallace[245],

Zhen Wang[115], Martin Walter[31,32], Lei Wang[246], Sara Jane Webb[247], Lars T. Westlye[10,11,248], Sarah Whittle[249,250], Mark O. Wielpütz[251], Katharina Wittfeld[99], Will Woods[252], Mon-Ju Wu[172], Tony T. Yang[253,254,255,256], Lakshmi N. Yatham[125,257], Tokiko Yoshida[114,258], Abe Yoshinari[210,259], Je-Yeon Yun[141,260], Qing Zhao[115], Giovana B. Zunta-Soares[172], ENIGMA Autism Working Group, ENIGMA Anxiety Working Group, ENIGMA Bipolar Disorder Working Group, ENIGMA Major Depression Working Group, ENIGMA OCD Working Group, ENIGMA Schizophrenia Working Group, Odile A. van den Heuvel[81,82], Lianne Schmaal[261,262], Elena Pozzi[261,262], Ole A. Andreassen[10,248], Christopher R. K. Ching[118], Katherine E. Lawrence[118], Gaon S. Kim[118], Jan K. Buitelaar[263], Theo G.M. van Erp[264,265], Dan J. Stein[266], Daniel S. Pine[267], Anderson M. Winkler[268], Janna Marie Bas-Hoogendam[269,270,271], Andre Zugman[272], Nic J.A. van der Wee[270], Nynke A. Groenewold[119], Andre Marquand[8], Boris C. Bernhardt[273], Neda Jahanshad[274], Tyler M. Moore[4,6], Paul M. Thompson[118], Sophia I. Thomopoulos[118], Simon B. Eickhoff[3,5], Matthias Kirschner[128,275,276], Theodore D. Satterthwaite[2,4,6], Sofie L. Valk[1,3,5]

**ENIGMA Autism Working Group:** R. Bernier, D. van Rooij, A. Retico, L. Gallagher, S.Y. Bookheimer, E. Daly, C.M. Freitag; **ENIGMA Autism Working Group:** S. Khalsa, H.D. Critchley, M. Paulus; **ENIGMA Bipolar Disorder Working Group:** F. Scheffler, H.S. Temmingh; **ENIGMA Major Depression Working Group:** S.-M. Koopowitz, M.L. Oudega, A. Uyar; **ENIGMA OCD Working Group**: E. Stewart, Y.C.J. Reddy, D. Brandeis, T. Nakamae, A. Nakagawa, A. Watanabe, K. Yamada; **ENIGMA Schizophrenia Working Group:** U. Schall, A.C. Yang, M. Lee, P.T. Michie

#### Affiliations

[1] Max Planck Institute for Human Cognitive and Brain Sciences, Leipzig, Germany

[2] Penn Lifespan Informatics and Neuroimaging Center, University of Pennsylvania, Philadelphia, PA, USA

[3] Institute of Neuroscience and Medicine, Brain and Behaviour (INM-7), Research Center Jülich, Jülich, Germany

[4] Brain Behavior Laboratory, Department of Psychiatry, University of Pennsylvania Perelman School of Medicine, Philadelphia, PA, USA

[5] Institute of Systems Neuroscience, Medical Faculty and University Hospital Düsseldorf, Heinrich Heine University Düsseldorf, Düsseldorf, Germany

[6] Lifespan Brain Institute (LiBI) of Penn Medicine and CHOP, University of Pennsylvania, Philadelphia, PA, USA

[7] Faculty of Medicine, Leipzig University, Leipzig, Germany

[8] Donders Institute for Brain, Cognition and Behaviour, Radboud University Medical Centre, Nijmegen, the Netherlands

[9] Radboud University Medical Center, Nijmegen, the Netherlands

[10] Centre for Precision Psychiatry, Division of Mental Health and Addiction, Oslo University Hospital and University of Oslo, Oslo, Norway

[11] Department of Psychology, University of Oslo, Oslo, Norway

[12] Department of Psychiatry, Bellvitge Biomedical Research Institute, Bellvitge University Hospital, Barcelona, Spain

[13] Psychiatry and Mental Health Group, Institut d'Investigació Biomèrica de Bellvitge (IDIBELL), Bellvitge University Hospital, Barcelona, Spain

[14] Department of Clinical Sciences, Faculty of Medicine, University of Barcelona, Barcelona, Spain

[15] Centro de Investigación Biomédica en Red de Salud Mental (CIBERSAM), Instituto de Salud Carlos III, Madrid, Spain

[16] Instituto de Investigación del Hospital Universitario La Paz (IdiPAZ), Madrid, Spain

[17] School of Medicine, Universidad Autónoma de Madrid, Madrid, Spain

[18] Olin Neuropsychiatry Research Center, Institute of Living, Hartford Hospital, Hartford, CN, USA

[19] Department of Psychiatry, Yale School of Medicine, New Haven, CN, USA

[20] University of Luebeck, Department of Psychiatry and Psychotherapy, Luebeck, Germany

[21] Department of Psychiatry, National Institute of Mental Health and Neuro Sciences (NIMHANS), Bengaluru, India

[22] Laboratory of Neuropsychiatry, Department of Clinical Neuroscience and Neurorehabilitation, Santa Lucia Foundation Istituto di Ricovero e Cura a Carattere Scientifico (IRCCS), Rome, Italy
[23] University of Minnesota Medical School, Department of Psychiatry and Behavioral Sciences, Minneapolis, MN, USA
[24] University of Minnesota, Masonic Institute for the Developing Brain (MIDB), Minneapolis, MN, USA
[25] Department of Psychiatry, Faculdade de Medicina, Universidade de São Paulo, São Paulo, Brazil
[26] Department of Methods and Techniques in Psychology, Pontifical Catholic University, São Paulo, SP, Brazil
[27] Leiden University Medical Center (LUMC), Department of Psychiatry, Leiden, the Netherlands
[28] Psychiatry and Clinical Psychobiology Unit, Division of Neuroscience, Istituto di Ricovero e Cura a Carattere Scientifico (IRCCS) Ospedale San Raffaele, Milan, Italy
[29] University Vita-Salute San Raffaele, Milan, Italy
[30] Centro de Investigación Biomédica en Red de Salud Mental (CIBERSAM), Instituto de Salud Carlos III, Barcelona, Spain
[31] Department of Psychiatry and Psychotherapy, Jena University Hospital, Friedrich-Schiller-University Jena, Jena, Germany
[32] German Center for Mental Health (DZPG), partner site Halle-Jena-Magdeburg, Germany
[33] Istituto di Ricovero e Cura a Carattere Scientifico (IRCCS), Fondazione Stella Maris, Pisa, Italy
[34] Virginia Commonwealth University, Richmond, VA, USA
[35] Department of Clinical Medicine, Child and Adolescent Psychiatry, University of Copenhagen, Copenhagen, Denmark
[36] KabScientific
[37] Center of Neurodevelopmental Disorders (KIND), Department of Women's and Children's Health, Centre for Psychiatry Research, Karolinska Institutet and Region Stockholm, Stockholm, Sweden
[38] Child and Adolescent Psychiatry, Stockholm Health Care Services, Stockholm, Sweden
[39] Curtin Autism Research Group, Curtin School of Allied Health, Curtin University, Perth, Australia
[40] Department of Pathophysiology and Transplantation, University of Milan, Milan, Italy
[41] Department of Neurosciences and Mental Health, Istituto di Ricovero e Cura a Carattere Scientifico (IRCCS) Fondazione Ca' Granda Ospedale Maggiore Policlinico, Milan, Italy
[42] Psychiatry and Clinical Psychobiology Unit, Institute of Experimental Neuroscience, Istituto di Ricovero e Cura a Carattere Scientifico (IRCCS) San Raffaele Hospital, Milan, Italy
[43] McLean Hospital, Harvard Medical School, Belmont, MA, USA
[44] Amsterdam University Medical Center, University of Amsterdam, Department of Psychiatry, Amsterdam, The Netherlands
[45] Amsterdam Neuroscience, Amsterdam, The Netherlands
[46] Section Forensic Family and Youth Care, Institute of Education and Child Studies, Leiden University, Leiden, the Netherlands
[47] Hospital das Clinicas HCFMUSP, Faculdade de Medicina, Universidade de Sao Paulo, Sao Paulo, SP, Brazil
[48] School of Biomedical Sciences and Pharmacy, The University of Newcastle, Callaghan, Australia
[49] Hunter Medical Research Institute, New Lambton, Australia
[50] Department of Clinical and Experimental Medicine, University of Pisa, Pisa (Italy)
[51] Tri-institutional Center for Translational Research in Neuroimaging and Data Science (TReNDS), Georgia State University/Georgia Institute of Technology/Emory University, Atlanta, GA, USA
[52] Department of Medicine, Faculty of Medicine and Health Sciences, Institute of Neurosciences, University of Barcelona, Barcelona, Spain
[53] Department of Child and Adolescent Psychiatry and Psychology, Hospital Clinic of Barcelona, Barcelona, Spain
[54] Institut d'Investigacions Biomèdiques August Pi i Sunyer (IDIBAPS), Barcelona, Spain
[55] Centro de Investigación Biomédica en Red de Salud Mental (CIBERSAM), Madrid, Spain
[56] Sant Pau Mental Health Research Group, Institut de Recerca Sant Pau (IR SANT PAU), Barcelona, Spain
[57] School of Clinical Medicine, University of New South Wales, Sydney, Australia
[58] Monash University, Melbourne, Australia
[59] Centre for Mental Health and Brain Sciences, Swinburne University of Technology, Melbourne, Australia

[60] Department of Psychiatry, Faculty of Medicine, Dentistry, and Health Sciences, University of Melbourne, Parkville, Australia
[61] Centre for Neuropsychiatric Genetics and Genomics, Division of Psychological Medicine and Clinical Neurosciences, Cardiff University, Cardiff, UK
[62] Faculty of Health, Medicine and Behavioural Sciences, University of Queensland, Brisbane, Australia
[63] Department of Occupational Therapy, College of Health Sciences, Kaohsiung Medical University, Kaohsiung, Taiwan
[64] Biomedical Artificial Intelligence Academy, Kaohsiung Medical University, Kaohsiung, Taiwan
[65] Department of Psychology and Neuroscience, Brigham Young University, Provo, UT, USA
[66] Life and Health Sciences Research Institute (ICVS), School of Medicine, University of Minho, Braga, Portugal
[67] ICVS/3B's, PT Government Associate Laboratory, Braga/Guimarães, Portugal
[68] University hospital virgen del Rocío, ibis / CSIC, Sevilla, Spain
[69] University of Sevilla, Sevilla, Spain
[70] University of Minnesota, Child and Adolescent Mental Health (CAMH) Division, Minneapolis, MN, USA
[71] Institute for Translational Psychiatry, University of Münster, Münster, Germany
[72] Department of Psychiatry, Medical School and University Medical Center OWL, Protestant Hospital of the Bethel Foundation, Bielefeld University, Bielefeld, Germany
[73] Center for Intervention and Research on Adaptive and Maladaptive Brain Circuits Underlying Mental Health (C-I-R-C), partner site Halle-Jena-Magdeburg, Germany
[74] The Ahmanson Lovelace Brain Mapping Center, University of California, Los Angeles, CA, USA
[75] Department of Psychiatry and Biobehavioral Sciences, University of California, Los Angeles (UCLA) School of Medicine, Los Angeles, CA, USA
[76] Semel Institute for Neuroscience and Human Behavior, University of California, Los Angeles, CA, USA
[77] Child Mind Institute, Autism Center, New York, NY, USA
[78] Anxiety Disorders Center, Institute of Living, Hartford, CN, USA
[79] Department of Psychiatry, Utrecht University Medical Center (UMC), Utrecht, the Netherlands
[80] Laboratory of Psychiatric Neuroimaging (LIM-21), Departamento e Instituto de Psiquiatria, Hospital das Clinicas, Faculdade de Medicina Universidade de São Paulo (HCFMUSP), Faculdade de Medicina Universidade de São Paulo, Brazil
[81] Amsterdam University Medical Center, Department of Psychiatry, Department of Anatomy and Neuroscience, Vrije Universiteit Amsterdam, Amsterdam, The Netherlands
[82] Amsterdam Neuroscience, Compulsivity, Impulsivity and Attention program, Amsterdam, The Netherlands
[83] Department of Child and Adolescent Psychiatry, Psychosomatic Medicine and Psychotherapy, University Hospital Frankfurt, Goethe University, Frankfurt am Main, Germany
[84] Translational Developmental Neuroscience Section, Division of Psychological and Social Medicine and Developmental Neurosciences, Faculty of Medicine, Technische Universität Dresden, Dresden, Germany
[85] Clinical Research, Nathan Kline Institute for Psychiatric Research, Orangeburg, NY, USA
[86] Department of Psychiatry, New York University Grossman School of Medicine, New York, NY, USA
[87] Institute of Child Development, University of Minnesota, Minneapolis, MN, USA
[88] Department of Pediatrics, University of Minnesota, Minneapolis, MN, USA
[89] University of Toronto, Toronto, ON, Canada
[90] The Centre for Addiction and Mental Health, Hospital for Sick Children, Toronto, ON, Canada
[91] Charmaine and Gordon McGill Center for Psychedelic Research and Therapy, Department of Psychiatry and Behavioral Sciences, The University of Texas at Austin Dell Medical School, Austin, TX, USA
[92] FIDMAG Sisters Hospitallers Research Foundation, Barcelona, Spain
[93] Harvard Graduate School of Education, Cambridge, MA, USA
[94] Harvard University, Cambridge, MA, USA
[95] Department of Psychiatry, University of Oxford, Oxford, UK
[96] Oxford Health NHS Foundation Trust, Oxford, UK
[97] Standardization of Computational Anatomy Techniques for Cognitive and Behavioral Sciences (SoCAT) Lab, Department of Psychiatry, Faculty of the Medicine, Ege University, Izmir, Türkiye

[98] Department of Psychology, Stanford University, Stanford, CA, USA
[99] Department of Psychiatry and Psychotherapy, University Medicine Greifswald, Greifswald, Germany
[100] German Center of Neurodegenerative Diseases (DZNE) Site Rostock/Greifswald, Greifswald, Germany
[101] Section for Experimental Psychopathology and Neuroimaging, Department of General Psychiatry, Heidelberg University, Heidelberg, Germany
[102] Department of Pediatrics, Yale University School of Medicine, New Haven, CT, USA
[103] Yale Child Study Center, Yale University School of Medicine, New Haven, CT, USA
[104] Department of Neuroscience, Yale University School of Medicine, New Haven, CT, USA
[105] Lifespan Brain Institute, Children's Hospital of Philadelphia (CHOP), Philadelphia, PA, USA
[106] School of Psychology, University of Surrey, Guildford, UK
[107] Department of Clinical and Biological Psychology, University of Bergen, Bergen, Norway
[108] Centre for Research and Education in Forensic Psychiatry, Department of Mental Health and Addiction, Oslo University Hospital, Oslo, Norway
[109] Department of Adult Psychiatry, Institute of Clinical Medicine, University of Oslo, Norway
[110] ASRB University of Newcastle, NSW, Australia
[111] School of Medicine and Public Health, University of Newcastle, NSW, Australia
[112] Izmir City Hospital, Izmir, Turkey
[113] Research Center for Child Mental Development, Chiba University, Chiba, Japan
[114] United Graduate School of Child Development, The University of Osaka, Suita, Japan
[115] Shanghai Mental Health Center, Shanghai Jiao Tong University School of Medicine, Shanghai, China
[116] Australian National Imaging Facility, The University of Queensland, St Lucia, Australia
[117] School of Psychiatry, Department of Neuroscience, University of Naples "Federico II", Naples, Italy
[118] Imaging Genetics Center, Mark and Mary Stevens Neuroimaging and Informatics Institute, Keck School of Medicine of USC, University of Southern California, Marina del Rey, CA, USA
[119] Department of Psychiatry and Mental Health, Neuroscience Institute, University of Cape Town, Cape Town, South Africa
[120] Center of Excellence on Mood Disorders, Louis A. Faillace, MD, Department of Psychiatry and Behavioral Sciences at McGovern Medical School, UTHealth Houston, TX, USA
[121] Department of Psychology, George Mason University, Fairfax, VA, USA
[122] Department of Neuroscience, Section of Psychiatry, Università Cattolica del Sacro Cuore, Rome, Italy
[123] Department of Neuroscience, Head-Neck and Chest, Section of Psychiatry, Fondazione Policlinico Universitario Agostino Gemelli IRCCS, Rome, Italy
[124] Instituto de Investigación Sanitaria del Hospital Gregorio Marañón (IISGM), Madrid, Spain
[125] Department of Psychiatry, Faculty of Medicine, University of British Columbia, Vancouver, Canada
[126] Psychology Department, Georgia State University, Atlanta, GA, USA
[127] Department of Psychology, The University of Hong Kong, Hong Kong
[128] Division of Adult Psychiatry, Department of Psychiatry, Geneva University Hospitals, Geneva, Switzerland
[129] Department of Psychiatry and Neurosciences, Graduate School of Biomedical and Health Sciences, Hiroshima University, Hiroshima, Japan
[130] Department of Psychology, Humboldt University of Berlin, Berlin, Germany
[131] Centre for Youth Bipolar Disorder, Centre for Addiction and Mental Health, Toronto, ON, Canada
[132] Department of Psychiatry, Seoul National University College of Medicine, Seoul, South Korea
[133] Department of Neuropsychiatry, Seoul National University Hospital, Seoul, South Korea
[134] Institute of Human Behavioral Medicine, SNU-MRC, Seoul, South Korea
[135] University of Marburg, School of Medicine, Department of Psychiatry and Psychotherapy, Marburg, Germany
[136] Department of Psychiatry and Neurochemistry, Institute of Neuroscience and Physiology, University of Gothenburg, Gothenburg, Sweden
[137] Department of Psychology, Stony Brook University, Stony Brook, NY, USA
[138] Department of Neuroradiology, TUM University Hospital, School of Medicine and Health, Technical University of Munich (TUM), Munich, Germany
[139] The University of Texas Health Science Center Houston, TX, USA

[140] Holland Bloorview Kids Rehabilitation Hospital, Toronto, ON, Canada
[141] Seoul National University Hospital, Seoul, Republic of Korea
[142] Hanyang University Hospital, Seoul, South Korea
[143] Department of Paediatrics and Child Health, University of Cape Town, Cape Town, South Africa
[144] Department of Medical Epidemiology and Biostatistics, Karolinska Institutet, Stockholm, Sweden
[145] Russian Mental Health Research Center (RMHRC), Moscow, Russia
[146] SA MRC Unit on Risk and Resilience in Mental Disorders, Department of Psychiatry, Stellenbosch University, Stellenbosch, South Africa
[147] University of Newcastle, NSW, Australia
[148] Department of Psychology, University of Pittsburgh, Pittsburgh, PA, USA
[149] Department of Psychiatry, University of Pittsburgh Medical Center, University of Pittsburgh, Pittsburgh, PA, USA
[150] Child and Adolescent Psychiatry Unit, Department of Medical Sciences, Uppsala University, Uppsala, Sweden
[151] Sunnybrook Research Institute, Toronto, ON, Canada
[152] Centre for Addiction and Mental Health, Toronto, ON, Canada
[153] Enrico Fermi Research Center, Rome, Italy
[154] Neuroimaging Laboratory, Santa Lucia Foundation, Rome, Italy
[155] Cardiff University, Brain Research Imaging Centre (CUBRIC), Cardiff University, Cardiff, UK
[156] Department of Psychiatry, School of Medicine, Universidade Federal do Rio Grande do Sul, Porto Alegre, Brazil
[157] Anxiety outpatient Unit, Hospital de Clínicas de Porto Alegre, Porto Alegre, Brazil
[158] Columbia University Irving Medical Center, New York, NY, USA
[159] The New York State Psychiatric Institute, New York, NY, USA
[160] Department of Radiology, Bellvitge University Hospital, Barcelona, Spain
[161] Centre for Psychiatry Research, Department of Clinical Neuroscience, Karolinska Institutet, and Stockholm Health Care Services, Stockholm, Sweden
[162] Centre for Neuroimaging, Cognition and Genomics (NICOG), Clinical Neuroimaging Laboratory, College of Medicine Nursing and Health Sciences, University of Galway, Galway, Ireland
[163] Department of Psychiatry, Trinity College Dublin, Dublin, Ireland
[164] 2CA-Braga, Hospital de Braga, Braga, Portugal
[165] Queensland Brain Institute, The University of Queensland, Brisbane Australia
[166] Queensland Centre for Mental Health Research, The University of Queensland, Brisbane, Australia
[167] Department of Biomedical Engineering, State University of New York at Stony Brook, Stony Brook, NY, USA
[168] Martinos Center for Biomedical Imaging, Massachusetts General Hospital, Charlestown, MA, USA
[169] MRI Core Facility, IDIBAPS, Barcelona, Spain
[170] Institute of Psychiatry Psychology and Neuroscience, King's College London, London, UK
[171] NIHR Maudsley Biomedical Research Centre, King's College London, London, UK
[172] Center of Excellence on Mood Disorders, Louis A. Faillace, MD, Department of Psychiatry and Behavioral Sciences at McGovern Medical School, UTHealth Houston, Texas, USA
[173] Monash Health and Department of Psychiatry, School of Clinical Sciences, Monash University, Melbourne, VIC, Australia
[174] OCD Clinic, National Institute of Mental Health And Neurosciences (NIMHANS), India
[175] Deakin Institute for Mental and Physical Health and Clinical Translation, School of Medicine, Deakin University, Australia
[176] Kyoto Prefectural University of Medicine, Kyoto, Japan
[177] Division of Mental Health and Substance Abuse, Diakonhjemmet Hospital, Oslo, Norway
[178] Division of Mental Health and Addiction, Institute of Clinical Medicine, University of Oslo, Oslo, Norway
[179] Swedish Collegium for Advanced Study, Uppsala, Sweden
[180] Department of Psychology, University of Virginia, Charlottesville, VA, USA
[181] Department of Radiology and Medical Imaging, University of Virginia, Charlottesville, VA, USA

[182] University of California, Los Angeles, CA, USA
[183] Division of Child and Adolescent Psychiatry, UCLA Semel Institute for Neuroscience, Los Angeles, CA, USA
[184] UCLA Brain Research Institute, Los Angeles, CA, USA
[185] Department of Physiology and Pharmacology, Wake Forest University School of Medicine, Winston-Salem, NC, USA
[186] Center for Neuropsychiatric Schizophrenia Research (CNSR), Mental Health Center, Glostrup, Copenhagen University Hospital, Mental Health Services CPH, Copenhagen, Denmark
[187] Monash Institute of Pharmaceutical Sciences (MIPS), Monash University, Parkville, Australia
[188] Western Centre for Health Research and Education (WCHRE), University of Melbourne and Western Health, Sunshine Hospital, St Albans, Australia
[189] University of Virginia, Charlottesville, VA, USA
[190] University of Pittsburgh, Pittsburgh, PA, USA
[191] Department of Basic and Clinical Psychology and Psychobiology, Jaume I University, Castelló de la Plana, Spain
[192] Department of Neurosciences and Mental Health, Fondazione IRCCS Ca' Granda, Ospedale Maggiore Policlinico, Milan, Italy
[193] Department of Psychiatry and Legal Medicine, Universitat Autonoma de Barcelona, Barcelona, Spain
[194] University of Pittsburgh, PA, USA
[195] NeuroRecovery Research Hub, School of Psychology, University of New South Wales (UNSW) Sydney, Sydney, Australia
[196] Centre for Pain IMPACT, Neuroscience Research Australia, Randwick, Australia
[197] Institut d'Investigacions Biomèdiques August Pi i Sunyer (IDIBAPS), Centro de Investigación Biomédica en Red de Salud Mental (CIBERSAM), Barcelona, Spain
[198] Institute of Neurosciences, University of Barcelona, Barcelona, Spain
[199] Consorci Sanitari del Maresme, Barcelona, Spain
[200] Centre of Research and Education in Forensic Psychiatry (SIFER), Oslo University Hospital, Oslo, Norway
[201] Department of Electronic Systems, Vilnius Tech, Vilnius, Lithuania
[202] Department of Medical Physiology and Biophysics, Instituto de Biomedicina de Sevilla (IBiS), HUVR/CSIC/Universidad de Sevilla/CIBERSAM, ISCIII, Sevilla, Spain
[203] Department of Psychiatry, University of Cambridge, Cambridge, UK
[204] Centre for Mental Health and Brain Sciences, Faculty of Health, Arts & Design, Swinburne University of Technology, Melbourne, Australia
[205] University of Sydney, Sydney, Australia
[206] School of Medicine and Health, TUM-NIC Neuroimaging Center, Technical University of Munich, Munich, Germany.
[207] School of Academic Psychiatry, Institute of Psychiatry, Psychology and Neuroscience, Department of Child & Adolescent Psychiatry, King's College London, London, UK
[208] Meditation Research Program, Department of Psychiatry, Massachusetts General Hospital, Harvard Medical School, Boston, MA, USA
[209] ATR Brain Information Communication Research Laboratory Group, Kyoto, Japan
[210] Department of Psychiatry, Graduate School of Medical Science, Kyoto Prefectural University of Medicine, Kyoto, Japan
[211] XNef, Inc., Kyoto, Japan
[212] Center of Mathematics, Computing and Cognition, Universidade Federal do ABC, Santo Andre, Brazil
[213] University of Basel, Department of Clinical Research (DKF), Basel, Switzerland
[214] NSW Health Pathology, NSW, Australia
[215] Department of Physiology and Pharmacology, Karolinska Institutet, Stockholm, Sweden
[216] Center for Affective, Stress and Sleep Disorders, University Psychiatric Clinics (UPK) Basel, Basel, Switzerland
[217] Department of Integrative Medicine, National Institute of Mental Health and Neurosciences (NIMHANS), Bengaluru, India

[218] West Region, Institute of Mental Health, Singapore
[219] Yong Loo Lin School of Medicine, National University of Singapore, Singapore
[220] Lee Kong Chian School of Medicine, Nanyang Technological University, Singapore
[221] Department of Psychiatry and Behavioral Neurosciences, McMaster University, Hamilton, ON, Canada
[222] Pediatric OCD Consultation Clinic, Anxiety Treatment and Research Clinic, SJH Healthcare, Hamilton, ON, Canada
[223] Department of Social Psychology and Quantitative Psychology, Institute of Neurosciences, University of Barcelona, Barcelona, Spain
[224] Centro de Investigación Biomédica en Red de Salud Mental (CIBERSAM), Barcelona, Spain
[225] Centro Universitário Max Planck (UniMAX), São Paulo, Brazil
[226] Centro Universitários de Jaguaríuna (UniFAJ), São Paulo, Brazil
[227] Clinical Academic Center of Braga (2CA-Braga), Braga, Portugal
[228] Department of Medicine I, Faculty of Medicine and University Hospital Carl Gustav Carus, TUD Dresden University of Technology, Dresden, Germany
[229] Else Kröner-Fresenius Center for Digital Health, TUD Dresden University of Technology, Dresden, Germany
[230] Laureate Institute for Brain Research, Tulsa, OK, USA
[231] Neuroscience Institute, New York University School of Medicine, New York, NY, USA
[232] Institute of Medical Psychology and Systems Neuroscience, University Hospital Münster, Münster, Germany
[233] Department of Psychiatry and Behavioral Neuroscience, University of Cincinnati, College of Medicine, Cincinnati, OH, USA
[234] Icahn School of Medicine at Mount Sinai, New York, NY, USA
[235] James J. Peters VA Medical Center, Bronx, NY, USA
[236] Department of Highly Specialized Pediatric Orthopedics and Medicine, Astrid Lindgren Children's Hospital, Karolinska University Hospital, Stockholm, Sweden
[237] School of Data Science, University of Virginia, Charlottesville, VA, USA
[238] Laboratory of Neuropsychiatry, Department of Clinical and Behavioral Neurology, Santa Lucia Foundation Istituto di Ricovero e Cura a Carattere Scientifico (IRCCS), Rome, Italy
[239] Clinical Institute of Neuroscience, University of Barcelona, Hospital Clinic, Barcelona, Spain
[240] University of Vic – Central University of Catalonia, Barcelona, Spain
[241] Intelligent Data Analysis Laboratory, Department of Electronic Engineering, University of Valencia (UV), Valencia, Spain
[242] Institute for Community Medicine, University Medicine Greifswald, Greifswald, Germany
[243] Amsterdam University Medical Center, Department of Psychiatry, Vrije Universiteit Amsterdam, Amsterdam, The Netherlands
[244] Amsterdam University Medical Center, Department of Anatomy and Neurosciences, Vrije Universiteit Amsterdam, Amsterdam, The Netherlands
[245] Department of Speech, Language, and Hearing Sciences, The George Washington University, Washington, D.C., USA
[246] Rotman Research Institute, Baycrest Academy for Research and Education, Toronto, ON, Canada
[247] Center for Child Health, Behavior and Development, Seattle Children's Research Institute, Seattle, WA, USA
[248] K.G. Jebsen Centre for Neurodevelopmental Disorders, University of Oslo, Oslo, Norway
[249] School of Psychology, Deakin University, Melbourne, Australia
[250] Melbourne School of Psychological Sciences, The University of Melbourne, Melbourne, Australia
[251] Department of Diagnostic and Interventional Radiology, University Medicine Greifswald, Greifswald, Germany
[252] Swinburne University of Technology, Melbourne, Australia
[253] UCSF Department of Psychiatry and Behavioral Sciences, San Francisco, CA, USA
[254] Division of Child and Adolescent Psychiatry, UCSF, San Francisco, CA, USA
[255] Weill Institute for Neurosciences, UCSF, San Francisco, CA, USA
[256] UCSF School of Medicine, San Francisco, CA, USA
[257] Department of Psychiatry, Institute of Mental Health, Vancouver, BC, Canada

[258] Cognitive Behavioral Therapy Center, Chiba University Hospital, Chiba, Japan
[259] Sugimoto Psychiatric Clinic, Kyoto, Japan
[260] Yeongeon Student Support Center, Seoul National University College of Medicine, Seoul, Republic of Korea
[261] Centre for Youth Mental Health, The University of Melbourne, Parkville, Australia
[262] Orygen, The National Centre of Excellence in Youth Mental Health, Parkville, Australia
[263] Department of Medical Neuroscience, Donders Institute for Brain, Cognition and Behavior, Radboud University Medical Center, Nijmegen, The Netherlands
[264] Clinical Translational Neuroscience Laboratory, Department of Psychiatry and Human Behavior, University of California Irvine, Irvine, CA, USA
[265] Center for the Neurobiology of Learning and Memory, University of California Irvine, Irvine, CA, USA
[266] SAMRC Unit on Risk & Resilience in Mental Disorders, Department of Psychiatry & Neuroscience Institute, University of Cape Town, Cape Town, South Africa
[267] National Institute of Mental Health Intramural Research Program, Bethesda, MD, USA
[268] Department of Human Genetics, University of Texas Rio Grande Valley, Brownsville, Texas, USA
[269] Leiden University, Institute of Psychology, Leiden, The Netherlands
[270] Leiden University Medical Center (LUMC), Department of Psychiatry, Leiden, The Netherlands
[271] Leiden Institute for Brain and Cognition, Leiden, the Netherlands
[272] Section on Development and Affective Neuroscience (SDAN), National Institute of Mental Health (NIMH), Bethesda, Maryland, USA
[273] Montreal Neurological Institute and Hospital, McGill University, Montreal, QC, Canada
[274] Laboratory of Brain eScience, Mark and Mary Stevens Neuroimaging and Informatics Institute, Keck School of Medicine of USC, University of Southern California, Marina del Rey, CA, USA
[275] Psychiatric University Hospital Zurich, University of Zurich, Zurich, Switzerland
[276] Synapsy Center for Neuroscience and Mental Health Research, Faculty of Medicine, University of Geneva, Switzerland

### Supplementary Tables

**Table S1** Sample overview. 10
**Table S2** Demographic information of the autism spectrum diagnosis (ASD) cohort. 11
**Table S3** Demographic information of the bipolar disorder (BP) cohort. 13
**Table S4** Demographic information of the generalized anxiety disorder (ANXG) cohort. 14
**Table S5** Demographic information of the major depressive disorder (MDD) cohort. 15
**Table S6** Demographic information of the obsessive-compulsive disorder (OCD) cohort. 16
**Table S7** Demographic information of the schizophrenia spectrum disorder (SCZ) cohort. 17
**Table S8** Technical cohort information for the generalized anxiety disorder cohort. 18
**Table S9** Technical cohort information for the autism cohort. 22
**Table S10** Technical cohort information for the bipolar disorder cohort. 25
**Table S11** Technical cohort information for the MDD cohort. 28
**Table S12** Technical cohort information for the obsessive-compulsive disorder cohort. 32
**Table S13** Technical cohort information for the schizophrenia spectrum disorder cohort. 34
**Table S14** Train and test sub-sample demographics. 39
**Table S15** Batch effects in raw data and deviation (z) scores. 40
**Table S16** Group differences in cortical thickness between individuals with a neurodevelopmental and psychiatric condition (NDPC) and reference comparators. 41
**Table S17** Group differences in surface area between individuals with a neurodevelopmental and psychiatric condition (NDPC) and reference comparators. 43
**Table S18** Group differences in subcortical volumes between individuals with a neurodevelopmental and psychiatric condition (NDPC) and reference comparators. 45
**Table S19** Demographic information for reference cohort subgroups (RC) matched to individuals with a neurodevelopmental or psychiatric condition (NDPC) via propensity score matching for Cohen's d map computation. 46
**Table S20** Similarities between Cohen's d maps from current normative models and previously published ENIGMA maps. 47
**Table S21** Within-diagnosis variability in cortical thickness. 48
**Table S22** Within-diagnosis variability in surface area. 51
**Table S23** Within-diagnosis variability in subcortical volumes. 54
**Table S24** Robustness of variability findings. 55
**Table S25** Spearman's rho correlation between Cohen's d and variability (standard deviation) maps, for cortical thickness data. 56
**Table S26** Leave-one-NDPC-out axes of deviation. 57
**Table S27** Leave-sites-out axes of deviation. 58
**Table S28** Association between symptom magnitude and PC scores. 59
**Table S29** Linear regressions between time since diagnosis and PC scores for each neurodevelopmental and psychiatric condition (NDPC). 61
**Table S30** Differences in PC scores between medicated and unmedicated individuals. 62
**Table S31** Overall percentage of individuals showing at least one extreme deviation $|z|>1.96$ per disorder group and modality, in any region. 64
**Table S32** Spearman's correlations between original Cohen's d maps and extreme deviation overlap maps. 65
**Table S33** Spearman's correlations between original Cohen's d maps and those derived from samples without extreme deviations ($|z|> 1.96$). 66
**Table S34** Transdiagnostic overlap robustness analysis using Kolmogorov-Smirnov distance. 67
**Table S35** Pairwise distribution overlap between individuals with a neurodevelopmental or psychiatric condition (NDPC) and the reference cohort, computed for each region separately. 68

### Supplementary Figures

### Supplementary Text

#### Sample demographics

The initial sample included data from 11,071 individuals with primarily one of six mental disorders (autism [ASD], generalized anxiety disorder [ANXG], bipolar disorder [BP], major depressive disorder [MDD], obsessive-compulsive disorder [OCD], schizophrenia spectrum disorder [SCZ]) and a reference cohort (RC) of 13,317 individuals without any neuropsychiatric condition, across 186 scanning sites. Participants were excluded if they were outside of the age range of five to 80 years, or if they had missing demographic or diagnostic information, or if they were initially considered healthy controls but were diagnosed with a neuropsychiatric condition not studied in this work. We further excluded individuals who had missing neuroimaging data for 15% of brain regions or more per modality. Last, we excluded sites with fewer than ten RC individuals after applying exclusion criteria to ensure site effects can be estimated properly. See **Supplementary Table S1** to **Table S7** for an overview of demographic distributions and sample sizes across diagnostic groups and sites. Note that some sites contributed data to multiple ENIGMA working groups.

To increase the sample size in the younger age range of our lifespan models, we further included cortical thickness and surface area data from two developmental datasets: the Philadelphia Neurodevelopmental Cohort (PNC[1]) and the Healthy Brain Network cohort (HBN[2]). We only included individuals who did not meet diagnostic criteria (i.e., scored '2' or lower on the GOASSESS questionnaire) for normative model training (PNC: $n$ = 56, Age: 16.05y +/- 4.13, 25% female; HBN: $n$ = 196, Age: 10.21y +/- 3.55, 48.0% female).

#### Human Connectome Project (HCP)

To contextualize our macroscale brain structural findings in the transdiagnostic cohort, we further leveraged group-level structural and functional connectivity data from a healthy young adult sample of the Human Connectome Project (HCP-YA[3]; unrelated participants; $n$ = 207; 83 males, mean Age = 28.73 ± 3.73 years, range=22-36 years). This group-level data was accessed through the ENIGMA Toolbox[4], which provides details on data processing. Briefly, participants underwent anatomical, diffusion-weighted and resting-state functional MRI.

Structural connectivity matrices were derived from anatomically constrained tractography. As part of preprocessing, b0 intensity normalization and head motion corrections were applied. Moreover, susceptibility distortion and eddy currents were corrected. Streamlines were then reconstructed for 68 cortical regions, using the Desikan-Killiany parcellation[5]. A group-average structural connectome was generated using distance-dependent thresholding and subsequently log-transformed.
Resting-state functional MRI data underwent multiple preprocessing steps, including motion and distortion corrections, intensity normalization and intensity inhomogeneity corrections, brain extraction, normalization to MNI152 space, and ultimately the projection to a 2D cortical surface. Just as the structural connectivity data, functional timeseries data was parcellated according to the Desikan-Killiany atlas. At the subject-level, functional connectivity matrices were computed as pair-wise Pearson correlations between time series and Fisher's z-transformed. Z-transformed individual-level matrices were then averaged across individuals to derive a group-level connectome.

#### Lausanne dataset

We further leveraged the openly available Lausanne dataset as an independent cohort providing group-level structural and functional connectomes. Neuroimaging data were collected at the University Hospital Center and the University of Lausanne (Department of Radiology). The dataset comprises $n$ = 70 healthy young adults (25.3 ± 4.9 years; 16 females)[6] and is available at zenodo (https://zenodo.org/record/2872624#.XOJqE99fhmM). All participants provided informed consent and the Ethics Committee of Clinical Research of the Faculty of Biology and Medicine, University of Lausanne, approved the protocol. A 3-T MRI scanner (Siemens Medical; Trio) with a 32-channel head coil was used. Scanning sessions included an anatomical scan, diffusion-weighted imaging, and a resting-state functional MRI sequence sensitive to blood-oxygen-level-dependent (BOLD) signals. A magnetisation-prepared rapid acquisition gradient echo (MPRAGE) that is sensitive to white/gray matter contrast was implemented for the anatomical scan, with 1.2 mm slice thickness and 1 mm in-plane resolution. 128 diffusion-weighted volumes and one b0 volume were derived using a DSI sequence (voxel size: 2.2 × 2.2 × 3.0 mm; maximum $b$-value 8000 s/mm$^2$). Lastly, 280 functional images were generated for each participant using a gradient echo-planar imaging (EPI) sequence to derive functional images (slice thickness: 3.3 mm in-plane resolution and slice thickness with a 0.3 mm gap, TR 1920 ms). For the current study, we leveraged publicly shared[57]group-level connectomes of the Lausanne dataset, available at https://doi.org/10.5281/zenodo.6795748, which were previously used to perform epicenter mapping based on psychiatric case-control difference maps in cortical thickness[7].

Cortical structural grey matter data were parcellated according to the Desikan-Killiany atlas (68 regions)[5]. Each individual's structural connectome was reconstructed using deterministic streamline tractography as implemented in the Connectome Mapping Toolkit[8]. This procedure initiated 32 streamline propagations per diffusion direction per white matter voxel. Individual structural connectomes were combined using a group-level consensus method designed to retain the characteristic density and edge-length profiles observed across participants (see [9]). The resulting whole-brain binary network had a connection density of 24.6%. In the weighted connectome, connection strengths were defined as the mean of the log-transformed non-zero streamline densities and subsequently normalized to the range [0, 1].

Functional MRI data were pre-processed using a pipeline optimized for subsequent connectivity analyses[10]. Functional volumes were motion corrected (3 rotation and 3 translations, as captured by rigid body co-registration) and corrected for further physiological variables, including white matter and cerebrospinal fluid. A low-pass filter was then applied to the BOLD time-series (temporal Gaussian filter with full width at half maximum, i.e., 1.92 s). In order to allow stabilization of the time series, the first four time points were excluded. Moreover, motion scrubbing was applied following strategies described in ref[10]. Functional data was parcellated according to the Desikan-Killiany atlas, which was also used for diffusion data, yielding the same 68 cortical regions. Functional connectivity was then computed at the individual level by computing inter-regional zero-lag Pearson correlations of the BOLD time-series, yielding a connectivity matrix. The group-level matrix was derived by computing the mean connectivity of each connection (i.e., cell of the matrix) across individuals. The derived group-level functional connectome was based on n = 69 participants instead of 70, as one participant did not undergo a functional scan.

#### Data exclusion, quality control, and imputation

We excluded individuals below five and above 80 years old, as sample sizes in these age ranges were too low to ensure good model fit. Individuals with missing demographic information (age, sex, site, diagnosis) were also excluded. Moreover, individuals with 15% or more missing data per imaging modality (CT, SA, SV) were excluded from further analyses. Neuroimaging data were quality controlled according to the ENIGMA QC guidelines (https://enigma.ini.usc.edu/protocols/imaging-protocols/ ), performed by the individual sites. We excluded 2 scan sites for which no unaffected comparator participant data were available, and 15 scan sites with less than 10 unaffected comparators. If 15% of brain regions or more did not meet QC criteria in a given participant, the respective participant was excluded. If 15% of participants or more in a given site had missing data in the same brain region, the site was excluded. Additionally, we excluded regional values that were more than 4 SD away from the mean (per disorder group) or fell outside a biologically plausible range (i.e., 1-5mm for CT, 50-12000 for SA). Following these exclusions, we imputed neuroimaging-derived data if less than 15% of regions were excluded in a given modality and subject. Imputation was done via nearest neighbor imputation based on the top 10% most similar subjects in a given site. On average (across brain regions), 0.37% of ANXG, 0.85% of BP, 0.55% of SCZ, 1.26% of MDD, 1.32% of OCD, and 0.19% of ASD data were imputed.

#### Demographics for the train and test data

For the purpose of generating normative reference lifespan curves, the reference (i.e., control) cohort was split into training ($RC_{train}$; 80%) and test ($RC_{test}$; 20%). The purpose of holding out a $RC_{test}$ set was to allow for comparisons between NDPC and RC cohorts, where reference data yield the same generalization error in their predictions rather than overfitted forward predictions. The split was performed for each site separately, ensuring that site estimates can be learned properly. Age and sex distributions were comparable between both sub-samples (**Table S14**).

#### Normative model training

A hierarchical Bayesian regression (HBR) model with age, sex, and site was used to create models that map the normative variation in 68 cortical thickness (CT) regions, 68 surface area (SA) regions, and 14 subcortical volume (SV) regions, across age while accounting for sex and site as batch effects. We chose HBR as it has previously been shown to robustly accommodate site effects, which was of utmost relevance in our multi-site study. We specifically employed the SHASHb extension implemented in PCNToolkit[11], which draws on a flexible distribution from the sinh-arc family to model non-Gaussian data with heteroskedastic kurtosis and skewness. We used B-spline models with five knots to model non-linear trajectories as well as standard deviation (varying as a function of the covariates). Normative models were trained in the $RC_{train}$ sub-set and then applied to both the $RC_{test}$ and patient samples. The predictions from this model provided us with individualized deviation scores, i.e., $z$-scores, for each individual and region as follows:

$$z_{ij} = \frac{f(x_{ij}) - y_{ij}}{\sqrt{\sigma^2 + \sigma_*^2}}$$

Where i is the individual, j is the region (per modality), $f(x_{ij})$ is the predicted value, $y_{ij}$ is the observed value, $\sigma^2$ reflects random Gaussian noise, and $\sigma^{2*}$ is the model's predictive variance. We computed multiple metrics to assess model fit of our normative models: 1) Explained Variance (EXPV), Spearman's rank order correlation between observed and predicted values, and skewness and kurtosis of derived z-scores. See **Figure S1** for a summary of each metric and imaging modality.

#### Batch correction: Modelling of between-site heterogeneity by Hierarchical Bayesian Regression

While the large-scale collaborative effort of our transdiagnostic ENIGMA sample is a large step towards generalizable, representative psychiatric neuroimaging, it comes with the challenge of heterogeneities between sites in terms of, e.g., demographics, scanners, and study procedures. Here, we leveraged a type of normative modeling that is particularly suitable for accounting for multi-site data, namely HBR. Previous work[12] has demonstrated the suitability of this method for large, multi-site samples, retaining more clinically relevant variance than two-stage harmonization approaches or other popular tools such as ComBat[13], while successfully accommodating site variation. Moreover, many harmonization approaches assume that distributions resemble a Gaussian distribution. Our HBR model estimated heteroscedasticity (skewness and kurtosis) of distributions during the estimation of deviations from batch-specific reference curves.

To test whether our model appropriately estimated batch effects (**Fig. S2)**, we computed ANOVAs and Intra-class correlation coefficients (ICC) based on raw data and HBR-derived z-scores in the healthy control reference data. Both indicated that the HBR approach successfully learned and accounted for batch effects (all ANOVAs $p<0.05$, all ICC $< 0.05$; see **Table S15**)

To further assure that no single site drives variance patterns in the test cohort, we performed a leave-one-site-out sensitivity analysis. Specifically, we iterated through all 172 sites and recomputed the PCs for each modality. We then computed the spatial correlation between the derived PC maps and the original PCs based on the full sample. We repeated this analysis, leaving out 10% of sites (17) per iteration and again computed the spatial correlation of derived PC loadings with original PC loadings. We found that derived PC patterns of covarying structural deviations were highly robust to leaving out individual sites or subsets of sites (see **Table S27**), indicating that spatial patterns observed in this study were not driven by potential site biases.

#### Association between within-diagnosis variability maps and group-shift maps

Traditional t-tests / Cohen's d maps can pick up on various elements of the distribution properties of the groups being compared. For example, subtle group shifts may 'stretch out' a distribution if only a sub-set within a group shows a deviation in a certain direction (e.g., CT decrease). In this case, shifts in part of the group can lead to both increased variability and a significant group difference in mean. In a second scenario, variability within a group may increase in both directions away from the mean. When positive and negative deviations balance each other, the group mean remains unchanged. As a result, standard statistical tests contrasting two groups (in this case, individuals with and without NDPCs) may fail to detect a difference between groups, despite substantial alterations being present within subsets of individuals. To test 1) whether increases in variability may mask potential group differences or 2) whether group differences are accompanied by increases in variability, we tested the spatial associations between Cohen's *d* and variability (SD) maps via variogram permutations[14]. However, this analysis revealed that the two phenomena are widely independent, with unrelated spatial patterning across the cortex, highlighting the importance of studying both phenomena for complementary perspectives (**Table S25**).

#### Axes of deviation derived in the reference ($RC_{test}$) cohort

Our main findings describe cortical and subcortical axes of structural deviations from the reference lifespan trajectories computed in the full sample, i.e., including all patients and $RC_{test}$ individuals. The rationale of this approach was to capture systematic elements of co-occurring structural deviations independent of diagnosis. However, in order to find out if the derived axes predominantly capture normative patterns of variation, we re-computed the principal components in our $RC_{test}$ cohort. Specifically, we included 50% of the $RC_{test}$ sample to fit the PCA, allowing us to apply the PCA to both individuals with NDPCs and the held-out $RC_{test}$ subgroup for further analysis. We observed that axes of structural deviation derived in the full or $RC_{test}$ cohorts were highly similar (see **Fig. 3F and Supplementary Fig. S11**). This indicates that the observed dominant spatial pattern may largely reflect general principles of structural covariance in the brain. We further observed that group differences in PC scores between individuals with and without NDPCs, i.e., the degree to which individuals expressed the pattern captured by PCs, were largely replicable when deriving the PCs in the full or $RC_{test}$ sample, with the exception of BP for CT PC2 (**Supplementary Fig. S11**).

#### Quantifying the relationship between typical cortical organization and NDPC-related patterns

Deriving principal axes of structural deviations in the reference cohort (see above) suggested that much of the variation observed in NDPC cohorts is structured along similar spatial patterns as observed in the general population. We performed three additional analyses to further characterize the relationship between such typical covariance patterns and potential NDPC-related effects.

1. *Projections of Cohen's d maps onto the normative subspace:* Based on the cortical thickness deviation scores, we performed a principal component analysis including only the reference cohort to define the typical (sub-)cortical subspace. We retained the first two principal components, as in the main, which captured the dominant axes of typical variation. For each NDPC, we then orthogonally projected their Cohen's d maps onto the typical PC1-PC2 subspace. The proportion of each NDPC map explained by the typical subspace was quantified as:

$$R^2 = \frac{||\hat{d}\,||^2}{||d\,||^2}$$

   Where *d* denotes the original Cohen's d map and $\hat{d}$ its orthogonal projection onto the normative subspace. As such, this analysis yields the fraction of the total squared magnitude of the Cohen's d map captured by the main PCs of typical variation.

   This analysis revealed that much of NDPC-related alterations predominantly unfold along pre-existing axes of normative cortical variation, while a NDPC-specific component, most pronounced in autism and MDD, remains outside this shared structure. Specifically, we observed that Cohen's *d* patterns for Schizophrenia ($R^2$ = 0.97), bipolar disorder ($R^2$ = 0.89), and OCD ($R^2$ = 0.76) were largely captured by the normative subspace, anxiety is intermediate

($R^2 = 0.53$), whereas ASD ($R^2 = 0.26$) and MDD ($R^2 = 0.29$) contain much more structure outside the first two normative PCs (keeping in mind that effect sizes for MDD were generally small).

2. *Comparison of NDPC and typical eigenspaces:* In order to investigate whether NDPC effects exist in the same low-dimensional geometry as typical variation, Cohen's d maps for each NDPC were concatenated into a matrix, per structural measure. We then performed singular value decomposition (SVD) to estimate the dominant NDPC eigenspace. Next, we computed principal angles between the NDPC PC1-PC2 subspace and the typical PC1-PC2 subspace derived in the reference cohort. In this analysis, small principal angles indicate strong alignment between the two subspaces, whereas larger angles indicate NDPC-related variation outside the dominant typical eigenspace.

   This analysis revealed one dominant spatial axis along which the typical and NDPC-related sub-space were aligned (angle = 9.6°). However, they were not globally aligned across the full two-dimensional subspace, as indicated by the second angle (83.5°), indicating additional variance outside of the typical covariance structure.

3. *Stability of the typical eigenspace.* Last, we leveraged our individual-level data and investigated to which degree the inclusion of individuals with NDPCs altered the dominant covariance structure observed in the reference cohort. To this end, we compared PCA solutions derived from the RC alone with PCA solutions derived from the combined RC (*n*=2466) and NDPC samples (*n*=10,135). Similarity between the two-dimensional PC1-PC2 subspaces was quantified using principal angles.

   Across cortical thickness, surface area, and subcortical volume, the PC1–PC2 subspaces remained highly similar (maximum principal angles <11°), suggesting that including patients produces only minimal rotation of the dominant covariance structure.

Taken together, these analyses indicate that NDPC-related alterations unfold, at least in part, along pre-existing axes of normative cortical variation, while also retaining NDPC-specific structure outside of the dominant subspace of typical covariance.

#### Association between extreme deviations, sub-threshold shifts, and Cohen's *d* maps

To investigate whether group-level patterns found in our data and in the literature are driven by a subset of individuals showing extreme deviations, we performed two additional analyses: 1) We tested the spatial alignment of extreme deviation maps and the case-control Cohen's d maps, and 2) we re-computed the Cohen's d maps after removing individuals exhibiting extreme deviations (for each region separately, in both patients and control participants). While we found that extreme deviation maps were indeed correlated with Cohen's *d* maps, this was also the case for sub-threshold patterns (See **Table S32 & S33**). This observation indicates that group-level maps are reflective of both the influence of extreme deviations of a few, as well as sub-threshold but common shifts in NDPC cohorts.

#### Principles of structural deviations in surface area and subcortical volume

Having established transdiagnostic convergence and variability patterns in CT (see Main), we finally examined whether similar observations apply to SA and SV. We started by identifying axes of deviation for SA and SV, using the same PCA approach as described for CT (**Supplementary Fig. S12**). The primary axis of SA deviation (20.3% variance explained) closely aligned with normative connectome organization (PC1: r = 0.74, $p_{spin}$ <0.0001; PC2: r = 0, $p_{spin}$ = 0.997). Consistent with CT findings, this indicated that macroscale structural alterations preferentially follow the spatial arrangement of highly connected hubs in the cortex. Neither the first nor the weaker second SA component (explaining 4.0% of variance) captured the sensorimotor-to-association axis (both $p$>0.05). For SV, we observed a segregation of deviations in the bilateral putamen from the nucleus accumbens (27.6% of variance explained), alongside a segregation of bilateral caudate nucleus from the hippocampus (12.7% of variance explained; **Supplementary Fig. S12**). Across NDPCs, there was a general tendency that individuals exhibiting stronger deviations along one axis (e.g., CT PC1) also exhibited stronger deviations along axes of other structural features (e.g., SA PC1; see **Supplementary Fig. S13**). Similar to CT findings, we observed group differences in the extent to which individuals expressed deviations along shared axes (**Supplementary Fig. S12**). In particular, SV deviations showed stronger expression along one or both axes across all NPDCs.

Inspecting the prevalence and convergence of extreme deviations among patients with NDPCs again underscored the subtle and heterogeneous nature of structural alterations (**Supplementary Fig. S15** to **S17**). At maximum, 3.50% (BP) to 7.32% (ANXG) showed extreme negative deviations in SA converging in the same region; 2.98% (SCZ) to 6.68% (ASD) showed converging positive SA deviations. Leveraging the weighted reference connectome computed in the HCP-YA sample, we again investigated whether extreme SA deviations co-occurred in regions that are typically strongly connected to each other. We found that prefrontal, particularly orbitofrontal regions, formed hubs in which extreme deviations tended to follow a network-like structure (see **Supplementary Fig. S19**). For SV, 3.76% (MDD) to 8.68% (SCZ) showed converging negative SV deviations, while 3.13% (BP) to 5.48% (SCZ) showed positive SV deviations in the same region. Regional extreme deviation percentage was higher than in the reference cohort in at least one structural measure for each NDPC (see **Supplementary Fig. S15** to **S17**; as tested by 10.000 non-parametric permutations). Supporting CT findings, extreme deviations were overall sparse and spatially heterogeneous within NDPC groups.

### Supplementary Tables

**Table S1** *Sample overview.*

| NDPC | N | Age (SD) | % female |
|---|---|---|---|
| **ANXG** | 765 | 26.32 (+/- 12.05) | 40.00 |
| **ASD** | 1556 | 16.03 (+/- 8.48) | 18.44 |
| **BP** | 1370 | 38.37 (+/- 13.16) | 58.69 |
| **MDD** | 1916 | 39.89 (+/- 15.38) | 64.77 |
| **OCD** | 1775 | 28.24 (+/- 11.49) | 50.14 |
| **SCZ** | 2753 | 35.29 (+/- 11.84) | 37.60 |
| **RC** | 11,998 | 30.61 (+/- 15.70) | 47.79 |

ANXG = generalized anxiety, ASD = autism spectrum diagnosis, BP = bipolar, MDD = major depressive, OCD = obsessive-compulsive, SCZ = schizophrenia spectrum disorders, RC = Reference cohort.

**Table S2** *Demographic information of the autism spectrum diagnosis (ASD) cohort.*
Reference comparators (RC) refer to individuals without ASD from the respective study sites.

| | ASD | | | | RC | | | |
|---|---|---|---|---|---|---|---|---|
| **Site** | **N** | **Age (mean)** | **Age (std)** | **% female** | **N** | **Age (mean)** | **Age (std)** | **% female** |
| **ABIDE 1_Caltech** | 19 | 27.42 | 10.26 | 21.05 | 19 | 28.84 | 11.07 | 21.05 |
| **ABIDE 1_KKI** | 22 | 9.91 | 1.63 | 18.18 | 33 | 10.12 | 1.36 | 27.27 |
| **ABIDE 1_Leuven 1** | 14 | 21.86 | 4.11 | 0 | 13 | 23.38 | 3.07 | 0 |
| **ABIDE 1_Leuven 2** | 14 | 13.79 | 1.48 | 14.29 | 20 | 14.35 | 1.66 | 25 |
| **ABIDE 1_Max_Mun** | 23 | 26.39 | 15.15 | 13.04 | 28 | 26.32 | 10.39 | 10.71 |
| **ABIDE 1_NYU** | 78 | 14.59 | 7.02 | 12.82 | 103 | 15.77 | 6.3 | 23.3 |
| **ABIDE 1_OHSU** | 7 | 10.86 | 1.35 | 0 | 11 | 9.82 | 1.17 | 0 |
| **ABIDE 1_OLIN** | 20 | 16.7 | 3.42 | 15 | 16 | 16.94 | 3.68 | 12.5 |
| **ABIDE 1_Pitt** | 30 | 18.93 | 7.21 | 13.33 | 26 | 18.69 | 6.74 | 15.38 |
| **ABIDE 1_sSbl** | 15 | 35 | 10.43 | 0 | 15 | 33.73 | 6.61 | 0 |
| **ABIDE 1_SDSU** | 14 | 14.71 | 1.82 | 7.14 | 22 | 14.32 | 1.94 | 27.27 |
| **ABIDE 1_Stanford** | 20 | 10 | 1.62 | 20 | 20 | 9.85 | 1.57 | 20 |
| **ABIDE 1_Trinity** | 24 | 17.29 | 3.57 | 0 | 25 | 17.16 | 3.79 | 0 |
| **ABIDE 1_UM 1** | 52 | 12.79 | 2.44 | 15.38 | 49 | 14.04 | 3.18 | 32.65 |
| **ABIDE 1_UM 2** | 13 | 14.92 | 1.61 | 7.69 | 16 | 16.19 | 3.25 | 6.25 |
| **ABIDE 1_USM** | 54 | 22.37 | 7.8 | 0 | 41 | 21.32 | 7.71 | 0 |
| **ABIDE 1_Yale** | 28 | 12.75 | 3.01 | 28.57 | 27 | 12.74 | 2.81 | 29.63 |
| **ABIDE 2_BNI 1** | 28 | 37.32 | 16.37 | 0 | 29 | 39.59 | 15.09 | 0 |
| **ABIDE 2_EMC 1** | 25 | 8.16 | 1.31 | 20 | 26 | 8.08 | 1.06 | 15.38 |
| **ABIDE 2_ETH 1** | 11 | 20.45 | 3.72 | 0 | 24 | 23.88 | 4.55 | 0 |
| **ABIDE 2_GU 1** | 47 | 10.87 | 1.65 | 17.02 | 53 | 10.45 | 1.76 | 49.06 |
| **ABIDE 2_IP 1** | 22 | 15.18 | 4.9 | 36.36 | 32 | 24.12 | 11.66 | 68.75 |
| **ABIDE 2_IU 1** | 19 | 25.05 | 9.56 | 21.05 | 20 | 23.75 | 4.9 | 25 |
| **ABIDE 2_KKI 1** | 53 | 10.4 | 1.61 | 28.3 | 153 | 10.37 | 1.25 | 35.95 |
| **ABIDE 2_NYU 1** | 45 | 10.09 | 5.97 | 11.11 | 30 | 9.53 | 3.34 | 6.67 |
| **ABIDE 2-OHSU 1** | 36 | 11.92 | 2.21 | 19.44 | 56 | 10.38 | 1.64 | 51.79 |
| **ABIDE 2_OILH 2** | 18 | 21.72 | 4 | 16.67 | 32 | 23.81 | 3.67 | 40.62 |
| **ABIDE 2_SDSU 1** | 33 | 12.91 | 3.37 | 21.21 | 25 | 13.24 | 3.1 | 8 |
| **ABIDE 2_TCD 1** | 20 | 14.9 | 3.37 | 0 | 21 | 15.71 | 3.21 | 0 |
| **ABIDE 2_UCD 1** | 16 | 14.94 | 1.88 | 25 | 14 | 14.71 | 1.68 | 28.57 |
| **ABIDE 2_UCLA 1** | 15 | 11.93 | 2.28 | 6.67 | 15 | 9.87 | 2.2 | 33.33 |
| **ABIDE 2_USM 1** | 17 | 18.35 | 7.02 | 11.76 | 15 | 24.87 | 7.22 | 20 |
| **Barcelona** | 43 | 12 | 3.18 | 9.3 | 93 | 18.6 | 6 | 21.51 |
| **BRC** | 18 | 15 | 2 | 0 | 32 | 14.47 | 2.57 | 0 |
| **Cattes** | 44 | 26.52 | 6.36 | 40.91 | 23 | 26.09 | 6.21 | 34.78 |
| **CMU** | 14 | 26.36 | 5.84 | 21.43 | 13 | 26.85 | 5.74 | 23.08 |
| **Dresden** | 21 | 34.33 | 11.79 | 14.29 | 24 | 36.08 | 11.06 | 16.67 |
| **Fair** | 39 | 12.03 | 2.21 | 15.38 | 39 | 11.23 | 1.39 | 35.9 |
| **Frankfurt** | 12 | 17.5 | 3.68 | 16.67 | 12 | 18.67 | 1.3 | 16.67 |
| **FSM** | 14 | 5.43 | 0.51 | 50 | 16 | 5.31 | 0.48 | 56.25 |
| **GENDAAR Harvard 1** | 35 | 11.8 | 3.24 | 31.43 | 25 | 11.6 | 3.08 | 52 |
| **GENDAAR Seattle 1** | 21 | 12.29 | 3.02 | 61.9 | 10 | 10 | 1.94 | 50 |
| **GENDAAR Seattle 2** | 32 | 11.84 | 2.71 | 34.38 | 37 | 13 | 2.68 | 51.35 |

| | | | | | | | | |
|---|---|---|---|---|---|---|---|---|
| **GENDAAR Ucla 1** | 43 | 12.02 | 2.7 | 44.19 | 33 | 13.39 | 3.3 | 45.45 |
| **GENDAAR Ucla 2** | 11 | 12.73 | 3.44 | 63.64 | 16 | 12.19 | 2.97 | 50 |
| **GENDAAR Yale 1** | 40 | 12.65 | 2.86 | 60 | 36 | 13.31 | 2.75 | 47.22 |
| **HGGM** | 35 | 12.71 | 2.48 | 5.71 | 31 | 12.29 | 2.81 | 3.23 |
| **Liehoe** | 41 | 10.1 | 1.69 | 9.76 | 39 | 9.31 | 1.79 | 10.26 |
| **MRC 1** | 39 | 24.44 | 6.16 | 0 | 41 | 29 | 7.13 | 0 |
| **MRC 2** | 28 | 26.96 | 7.43 | 0 | 29 | 27.45 | 5.98 | 0 |
| **Pitt** | 30 | 15.97 | 5.1 | 16.67 | 64 | 16.56 | 6.19 | 18.75 |
| **Sao Paulo** | 15 | 12.8 | 4.36 | 0 | 18 | 12.11 | 3.53 | 0 |
| **RATSS** | 60 | 14.77 | 4.91 | 35 | 95 | 19.2 | 6.54 | 55.79 |
| **TCD 2** | 17 | 16.53 | 3.62 | 0 | 22 | 16.77 | 3.32 | 0 |
| **UMCU 1** | 35 | 13.57 | 4.21 | 8.57 | 33 | 15.42 | 5.5 | 9.09 |
| **Wougro 1** | 17 | 14.59 | 1.91 | 17.65 | 16 | 15.44 | 1.59 | 12.5 |

**Table S3** *Demographic information of the bipolar disorder (BP) cohort.*
Reference comparators (RC) refer to individuals without BP from the respective study sites.

| | BP | | | | RC | | | |
|---|---|---|---|---|---|---|---|---|
| ***Site*** | *N* | *Age (mean)* | *Age (std)* | *% female* | *N* | *Age (mean)* | *Age (std)* | *% female* |
| **Cardiff** | 86 | 40.23 | 8.33 | 65.12 | 60 | 38.25 | 9.21 | 65 |
| **CIAM** | 25 | 29.64 | 5.19 | 40 | 61 | 26.59 | 4.87 | 45.90 |
| **COGSBD** | 64 | 37.77 | 11.26 | 45.31 | 31 | 36.23 | 11.88 | 58.06 |
| **CYBD** | 78 | 17.32 | 1.51 | 64.1 | 75 | 16.97 | 1.83 | 50.67 |
| **FIDMAG** | 102 | 41.71 | 9.36 | 55.88 | 240 | 39.39 | 10.01 | 54.58 |
| **FOR2107 Marburg 1** | 49 | 43.08 | 11.81 | 63.27 | 397 | 34 | 12.56 | 60.96 |
| **FOR2107 Marburg 2** | 16 | 39.5 | 13.64 | 56.25 | 185 | 40.07 | 12.76 | 62.70 |
| **FOR2107 Münster** | 73 | 40.58 | 12.02 | 50.68 | 356 | 31.24 | 12.15 | 67.70 |
| **Idibaps** | 25 | 43.88 | 16.41 | 44 | 57 | 33.68 | 9.65 | 63.16 |
| **IGP** | 66 | 36.45 | 12.13 | 65.15 | 63 | 35.59 | 10.84 | 49.21 |
| **IMH** | 46 | 33.98 | 10.75 | 58.7 | 59 | 35.42 | 9.83 | 52.54 |
| **Montpellier** | 87 | 38.31 | 10.21 | 72.41 | 65 | 38.94 | 7.18 | 44.62 |
| **Nuig galway** | 45 | 39.87 | 10.43 | 57.78 | 80 | 36.17 | 11.13 | 41.25 |
| **Rome** | 189 | 44.04 | 12.49 | 50.79 | 221 | 44.69 | 15.87 | 57.47 |
| **SBA** | 36 | 38.19 | 9.23 | 50.00 | 56 | 36.8 | 10.6 | 60.71 |
| **SBP** | 139 | 39.79 | 12.23 | 63.31 | 85 | 38.82 | 14.65 | 51.76 |
| **Stop m UBC** | 55 | 22.85 | 4.75 | 52.73 | 38 | 23.71 | 4.57 | 50.00 |
| **TAMI**[15–17] | 136 | 47.01 | 13.35 | 67.65 | 436 | 43.32 | 16.38 | 60.55 |
| **UPenn (BD)** | 53 | 29.66 | 10.1 | 60.38 | 84 | 38.06 | 14.24 | 51.19 |

**Table S4** *Demographic information of the generalized anxiety disorder (ANXG) cohort.*
Reference comparators (RC) refer to individuals without ANXG from the respective study sites.

| | ANXG | | | | RC | | | |
|---|---|---|---|---|---|---|---|---|
| **Site** | **N** | **Age (mean)** | **Age (std)** | **% female** | **N** | **Age (mean)** | **Age (std)** | **% female** |
| **Barcelona** | 30 | 23.53 | 4.61 | 33.33 | 93 | 18.6 | 6.0 | 21.51 |
| **Boystown** | 49 | 15.8 | 1.37 | 40.82 | 44 | 15.39 | 1.62 | 47.73 |
| **Dresdensteinhauser** | 27 | 26.22 | 6.46 | 0 | 28 | 24.25 | 5.05 | 0 |
| **FOR2107 Marburg 1** | 64 | 34.7 | 12.82 | 70.31 | 397 | 34 | 12.56 | 60.96 |
| **FOR2107 Marburg 2** | 18 | 32.44 | 11.33 | 77.78 | 185 | 40.07 | 12.76 | 62.7 |
| **FOR2107 Münster** | 103 | 34.89 | 12.14 | 69.9 | 356 | 31.24 | 12.15 | 67.7 |
| **Harvard** | 180 | 24.98 | 4.54 | 32.78 | 42 | 25.43 | 3.88 | 38.1 |
| **Houston** | 6 | 20 | 12.47 | 33.33 | 201 | 32.35 | 13.96 | 37.31 |
| **IOL** | 32 | 38.78 | 13.41 | 34.38 | 16 | 39.25 | 15.04 | 18.75 |
| **Milan** | 33 | 45.06 | 15.13 | 39.39 | 63 | 34.29 | 12.56 | 41.27 |
| **Münster** | 25 | 29.16 | 10.12 | 28 | 29 | 27.83 | 9.2 | 41.38 |
| **Protaia** | 15 | 16.53 | 2.53 | 40 | 14 | 17.64 | 2.56 | 42.86 |
| **SDAN 2** | 66 | 13.21 | 2.86 | 28.79 | 65 | 13.58 | 2.72 | 40 |
| **Stonybrook** | 40 | 22.98 | 6.04 | 0 | 20 | 21.5 | 5.75 | 0 |
| **Stonybrook-klein** | 22 | 10.41 | 0.8 | 59.09 | 49 | 10.18 | 0.93 | 57.14 |
| **Sussex** | 19 | 29.79 | 7 | 10.53 | 21 | 28.67 | 9.45 | 14.29 |
| **UCSD** | 9 | 23.78 | 8.83 | 0 | 15 | 20.33 | 3.74 | 53.33 |
| **UPenn** | 27 | 15.78 | 3.4 | 48.15 | 428 | 14.22 | 4.06 | 51.4 |

**Table S5** *Demographic information of the major depressive disorder (MDD) cohort.*
Reference comparators (RC) refer to individuals without MDD from the respective study sites.

| | MDD | | | | RC | | | |
|---|---|---|---|---|---|---|---|---|
| *Site* | *N* | *Age (mean)* | *Age (std)* | *% female* | *N* | *Age (mean)* | *Age (std)* | *% female* |
| **BARC** | 39 | 46.79 | 7.69 | 79.49 | 23 | 46.39 | 8.35 | 73.91 |
| **Basel** | 84 | 39.69 | 11.71 | 55.95 | 54 | 25.93 | 3.94 | 53.7 |
| **CSAN** | 60 | 35.92 | 13.49 | 66.67 | 49 | 33.2 | 12.19 | 69.39 |
| **DCHS 1** | 6 | 30.17 | 7.57 | 100 | 23 | 29.65 | 7.04 | 100 |
| **DCHS 2** | 5 | 32.2 | 5.72 | 100 | 25 | 32.68 | 7.59 | 100 |
| **Episca** | 19 | 15.42 | 1.5 | 84.21 | 30 | 14.73 | 1.55 | 86.67 |
| **FIDMAG** | 34 | 49.26 | 12.37 | 61.76 | 240 | 39.39 | 10.01 | 54.58 |
| **FOR2107 Marburg 1** | 269 | 38 | 13.66 | 63.57 | 397 | 34 | 12.56 | 60.96 |
| **FOR2107 Marburg 2** | 64 | 40.88 | 13.88 | 62.5 | 185 | 40.07 | 12.76 | 62.7 |
| **FOR2107 Münster** | 336 | 35.28 | 12.6 | 66.07 | 356 | 31.24 | 12.15 | 67.7 |
| **Hiroshima 1** | 57 | 43.33 | 12.18 | 43.86 | 67 | 34.75 | 12.97 | 56.72 |
| **Hiroshima 2** | 16 | 40.5 | 11.48 | 62.5 | 48 | 42 | 11.58 | 75 |
| **Hiroshima 3** | 33 | 44.82 | 11.47 | 39.39 | 22 | 46.55 | 9.59 | 59.09 |
| **Hiroshima 4** | 44 | 46.36 | 12.23 | 52.27 | 16 | 43.31 | 8.62 | 50.00 |
| **IMH** | 22 | 40.09 | 7.61 | 45.45 | 59 | 35.42 | 9.83 | 52.54 |
| **Moraldilemma** | 24 | 19.42 | 2.19 | 100 | 45 | 18.51 | 1.79 | 100 |
| **Oxford** | 38 | 30.13 | 10.56 | 63.16 | 31 | 30.29 | 10.02 | 58.06 |
| **Rome** | 51 | 46.25 | 13.92 | 62.75 | 221 | 44.69 | 15.87 | 57.47 |
| **SF** | 73 | 15.63 | 1.36 | 64.38 | 78 | 15.37 | 1.26 | 48.72 |
| **SHIP-START-2 and SHIP-TREND-0** | 395 | 50.17 | 11.44 | 68.61 | 1170 | 51.6 | 13.52 | 46.92 |
| **Stanford T1w Aggregate** | 53 | 37.11 | 10.18 | 58.49 | 53 | 37.15 | 10.6 | 58.49 |
| **TAMI**[15–17] | 102 | 53.98 | 14.09 | 64.71 | 436 | 43.32 | 16.38 | 60.55 |
| **TIPS** | 24 | 46.17 | 11.92 | 58.33 | 61 | 48.07 | 14.61 | 47.54 |
| **UMN** | 68 | 15.88 | 1.78 | 76.47 | 38 | 16.24 | 2.2 | 65.79 |

**Table S6** *Demographic information of the obsessive-compulsive disorder (OCD) cohort.*
Reference comparators (RC) refer to individuals without OCD from the respective study sites.

| | OCD | | | | RC | | | |
|---|---|---|---|---|---|---|---|---|
| **Site** | **N** | **Age (mean)** | **Age (std)** | **% female** | **N** | **Age (mean)** | **Age (std)** | **% female** |
| **Benedetti** | 16 | 29.56 | 10.11 | 37.5 | 21 | 26.19 | 6.1 | 14.29 |
| **Beucke** | 80 | 32.52 | 9.97 | 48.75 | 80 | 31.94 | 10 | 50 |
| **Buitelaar** | 18 | 10.44 | 1.38 | 44.44 | 59 | 10.95 | 1.12 | 28.81 |
| **Cheng** | 19 | 29.84 | 8.99 | 68.42 | 34 | 31.29 | 7.7 | 73.53 |
| **Cheng 2** | 45 | 33.64 | 10.96 | 48.89 | 82 | 26.3 | 4.29 | 70.73 |
| **denys** | 20 | 33.5 | 10.42 | 85 | 19 | 40.21 | 11.65 | 73.68 |
| **Fitzgerald** | 44 | 14.32 | 2.56 | 56.82 | 43 | 13.14 | 2.47 | 48.84 |
| **Gruner** | 21 | 14.33 | 2.24 | 42.86 | 22 | 14.18 | 2.34 | 50 |
| **Heuvel 1** | 43 | 34.58 | 8.99 | 72.09 | 35 | 32.09 | 8.57 | 65.71 |
| **Heuvel 2** | 32 | 39 | 10.49 | 43.75 | 34 | 40.24 | 11.37 | 52.94 |
| **Hirano 1** | 67 | 27.4 | 11.03 | 55.22 | 49 | 28.8 | 9.25 | 53.06 |
| **Hoexter 1** | 39 | 32 | 10.41 | 53.85 | 15 | 29.93 | 9.43 | 46.67 |
| **Hoexter 2** | 26 | 12.5 | 2.5 | 42.31 | 23 | 12.04 | 2.62 | 52.17 |
| **Huyser** | 17 | 13.82 | 2.51 | 70.59 | 13 | 13.85 | 2.34 | 61.54 |
| **Koch** | 75 | 31.07 | 9.52 | 62.67 | 71 | 30.23 | 8.56 | 60.56 |
| **Kwonnmc** | 45 | 24.76 | 5.36 | 24.44 | 103 | 24.1 | 3.62 | 44.66 |
| **Kwonsnu** | 34 | 28.76 | 6.76 | 44.12 | 45 | 24.89 | 5.35 | 35.56 |
| **Lazaro** | 31 | 14.61 | 2.04 | 41.94 | 30 | 15 | 1.8 | 53.33 |
| **Lazaro** | 57 | 14.53 | 2.04 | 38.6 | 44 | 14.57 | 2.1 | 45.45 |
| **Marsh** | 21 | 12.14 | 3.69 | 52.38 | 13 | 9.15 | 2.58 | 46.15 |
| **Mataix_cols** | 37 | 39.35 | 11.13 | 59.46 | 29 | 37.66 | 11.14 | 68.97 |
| **Menchon** | 102 | 34.72 | 9.09 | 50.98 | 56 | 32.5 | 10.13 | 57.14 |
| **Morgado** | 58 | 27.76 | 7.45 | 53.45 | 52 | 27.75 | 6.18 | 63.46 |
| **Nakamae 1** | 81 | 31.77 | 9.09 | 53.09 | 48 | 30.44 | 7.9 | 54.17 |
| **Nakamae 2** | 34 | 32.82 | 9.74 | 64.71 | 39 | 29.77 | 7.36 | 53.85 |
| **Nakao** | 74 | 36.58 | 10.32 | 58.11 | 32 | 38.31 | 12.16 | 53.12 |
| **Nurmi** | 106 | 22.08 | 13.01 | 47.17 | 59 | 20.22 | 11.73 | 45.76 |
| **Reddy 1** | 31 | 28.26 | 6.5 | 41.94 | 20 | 25.9 | 5.66 | 30 |
| **Reddy 2** | 214 | 28.45 | 7.54 | 46.26 | 172 | 25.8 | 5.88 | 38.95 |
| **Simpson** | 33 | 29.67 | 8.12 | 48.48 | 33 | 28.24 | 8.06 | 48.48 |
| **Soreni** | 18 | 13.11 | 2.47 | 61.11 | 19 | 10.68 | 2.75 | 47.37 |
| **Spalletta** | 82 | 36.82 | 11.58 | 34.15 | 125 | 36.54 | 10.62 | 41.6 |
| **Stein** | 22 | 30.68 | 10.83 | 50 | 27 | 31.48 | 10.61 | 62.96 |
| **Stern** | 15 | 27.87 | 6.9 | 66.67 | 17 | 28.35 | 7.32 | 58.82 |
| **Stewart** | 27 | 15 | 2.65 | 59.26 | 30 | 14.03 | 3.5 | 60 |
| **Tolin** | 27 | 32.11 | 12.04 | 33.33 | 32 | 48 | 11.87 | 78.12 |
| **Walitza** | 14 | 31.71 | 7.93 | 50 | 15 | 32.6 | 9.36 | 73.33 |
| **Wang** | 50 | 29.72 | 9.41 | 46 | 36 | 26.14 | 7.63 | 47.22 |

**Table S7** *Demographic information of the schizophrenia spectrum disorder (SCZ) cohort.*
Reference comparators (RC) refer to individuals without SCZ from the respective study sites.

| | SCZ | | | | RC | | | |
|---|---|---|---|---|---|---|---|---|
| **Site** | **N** | **Age (mean)** | **Age (std)** | **% female** | **N** | **Age (mean)** | **Age (std)** | **% female** |
| **ASRB 0** | 27 | 41.11 | 8.4 | 44.44 | 22 | 39.32 | 15.05 | 63.64 |
| **ASRB 1** | 54 | 39.57 | 11.81 | 31.48 | 16 | 40.69 | 15.13 | 50 |
| **ASRB 2** | 19 | 37.68 | 8.71 | 42.11 | 25 | 42.76 | 12.38 | 60 |
| **ASRB 4** | 13 | 35.54 | 11.37 | 53.85 | 10 | 43.6 | 14.97 | 60 |
| **CIAM** | 16 | 31.12 | 6.48 | 31.25 | 61 | 26.59 | 4.87 | 45.9 |
| **CLING** | 49 | 32.43 | 9.36 | 26.53 | 323 | 25.18 | 5.28 | 59.13 |
| **Cobre** | 69 | 37.41 | 13.38 | 18.84 | 66 | 36.11 | 11.93 | 30.3 |
| **FIDMAG** | 160 | 39.64 | 11.86 | 22.5 | 240 | 39.39 | 10.01 | 54.58 |
| **FOR2107 Marburg 1** | 60 | 38.23 | 11.76 | 51.67 | 397 | 34 | 12.56 | 60.96 |
| **FOR2107 Marburg 2** | 38 | 38.45 | 11.92 | 44.74 | 185 | 40.07 | 12.76 | 62.7 |
| **FOR2107 Münster** | 34 | 37.32 | 10.72 | 55.88 | 356 | 31.24 | 12.15 | 67.7 |
| **Hubin** | 113 | 41.62 | 7.6 | 26.55 | 113 | 41.6 | 8.93 | 34.51 |
| **IGP** | 57 | 42.09 | 10.93 | 40.35 | 63 | 35.59 | 10.84 | 49.21 |
| **KASP** | 82 | 28.46 | 7.39 | 37.8 | 56 | 26.57 | 5.31 | 50 |
| **MCIC MGH** | 32 | 36.5 | 10.33 | 25 | 24 | 39.83 | 8.86 | 41.67 |
| **MCIC UMN** | 31 | 31.55 | 10.24 | 25.81 | 25 | 32.8 | 12.72 | 44 |
| **MCIC UMN** | 42 | 31.71 | 11.43 | 21.43 | 44 | 28.68 | 11.51 | 20.45 |
| **MPRC** | 206 | 35.42 | 12.84 | 37.86 | 231 | 37.1 | 15.21 | 58.44 |
| **NU** | 107 | 34.03 | 12.94 | 31.78 | 92 | 31.87 | 14.46 | 44.57 |
| **Pafip** | 352 | 29.86 | 8.79 | 39.2 | 195 | 29.12 | 7.7 | 38.46 |
| **Rome_sl** | 162 | 39.36 | 11.3 | 66.67 | 113 | 37.42 | 11.41 | 62.83 |
| **RSCZ** | 46 | 22.15 | 3.28 | 0 | 52 | 22.35 | 2.89 | 0 |
| **Score** | 140 | 26.17 | 6.32 | 29.29 | 54 | 25.93 | 3.94 | 55.56 |
| **TAMI**[15–17] | 258 | 42.95 | 12.36 | 56.2 | 436 | 43.32 | 16.38 | 60.55 |
| **Top_stop – Top3T** | 95 | 27.96 | 8.51 | 33.68 | 319 | 30.59 | 8.23 | 41.69 |
| **Top_stop Topge750** | 148 | 30.47 | 9.34 | 31.08 | 566 | 31.71 | 11.19 | 49.12 |
| **UCISZ** | 27 | 42.93 | 10.62 | 18.52 | 30 | 41.37 | 12.26 | 23.33 |
| **UNINA** | 49 | 37.45 | 9.67 | 30.61 | 55 | 42.4 | 15.66 | 47.27 |
| **UPenn (SCZ)** | 167 | 38.78 | 12.09 | 40.72 | 186 | 36.06 | 13.45 | 53.76 |
| **Voices** | 40 | 43.25 | 10.65 | 57.5 | 48 | 31.94 | 12.2 | 52.08 |
| **Zurich 2023** | 60 | 30.53 | 8.48 | 25 | 28 | 32.54 | 9.32 | 35.71 |

**Table S8** *Technical cohort information for the generalized anxiety disorder cohort.*

| Site | Country | Scanner | Acquisition | Slice Orientation | FreeSurfer Version | Exclusion criteria |
|---|---|---|---|---|---|---|
| Barcelona | Spain | Signa Excite system / GE | 1.5 T | 130-slice 3-dimensional SPGR sequence in the axial plane (time of repetition, 11.848 ms; time of echo, 4.2 ms; flip angle, 15°; field of view, 30 cm; matrix 256 × 256 pixels; in-plane resolution, 1.17 mm2; section thickness, 1.2 mm without inter-slice gap) | 6.0.0 | Exclusion criteria for individuals with GAD were current or past autism spectrum disorders, bipolar disorder, psychosis, or schizophrenia. |
| Boystown | U.S. | Siemens Skyra | 3.0 T | MP-RAGE, repetition time=2200ms, echo time=2.48ms; 230mm field of view; 8o flip angle; 256x208 matrix; thickness 1mm; voxel size 0.9x0.9x1mm3, distance factor 50% | 6.0.0 | Exclusion criteria for individuals with GAD were current or past autism spectrum disorders, bipolar disorder, psychosis, or schizophrenia. |
| Dresdensteinhauser | U.S. (Tulsa, OK, USA) | MR 750 (GE) | 3.0 T | FOV = 240×192 mm, matrix = 256×256, 186 axial slices, slice thickness = 0.9 mm, 0.938×0.938×0.9 mm3 voxel volume, TR = 5 ms, TE = 2.012 ms, SENSE acceleration factor R = 2, flip angle = 8°, delay time = 1400 ms, inversion time = 725 ms, sampling bandwidth = 31.25 kHz. | 6.0.0 | Exclusion criteria for individuals with GAD were current or past autism spectrum disorders, bipolar disorder, psychosis, or schizophrenia. |
| For2107 Marburg 1 | Germany | Tim Trio, Siemens | 3 T | 176 sagittal slices, slice gap 0.5mm, TR=1900ms, TE=2.26ms, inversion time=900ms, FA=9°, voxel size=1x1x1mm³ | 6.0.0 | Exclusion criteria for individuals with GAD were current or past autism spectrum disorders, bipolar disorder, psychosis, or schizophrenia. |
| For2107 Marburg 2 | Germany | Tim Trio, Siemens | 3 T | 176 sagittal slices, slice gap 0.5mm, TR=1900ms, TE=2.26ms, inversion time=900ms, FA=9°, voxel size=1x1x1mm³ | 6.0.0 | Exclusion criteria for individuals with GAD were current or past autism spectrum |

| | | | | | | |
|---|---|---|---|---|---|---|
| | | | | | | disorders, bipolar disorder, psychosis, or schizophrenia. |
| For2107 Münster | Germany | Prisma Siemens | 3 T | 192 sagittal slices, slice gap 0.5mm, TR=2130ms, TE=2.28ms, inversion time=900ms, FA=8°, voxel size=1x1x1mm³ | 6.0.0 | Exclusion criteria for individuals with GAD were current or past autism spectrum disorders, bipolar disorder, psychosis, or schizophrenia. |
| Harvard | U.S. | Magnetom Siemens | 3T | MPRAGE (MEMPRAGE) sequence on a Siemens MAGNETOM Connectom scanner (1.0 mm isotropic voxel size, FOV = 256 × 256 × 208 mm, TR/TI = 2530/1100 ms, flip angle = 7°, 4 echoes, GRAPPA acceleration factor 4) | 6.0.0 | Exclusion criteria for individuals with GAD were current or past autism spectrum disorders, bipolar disorder, psychosis, or schizophrenia. |
| Houston | U.S. | Philips Gyroscan Intera | 1.5T | Repetition time (TR) = 24 ms, echo time (TE) = 5 ms, flip angle = 40°, field of view (FOV) = 24 cm, Slice thickness = 1 mm, voxel dimension = 1 × 1 × 1 mm3 and matrix size = 256 × 256. | 6.0.0 | Exclusion criteria for individuals with GAD were current or past autism spectrum disorders, bipolar disorder, psychosis, or schizophrenia. |
| IOL | U.S. | Siemens Allegra | 3 T | 3D MPRAGE pulse sequence (repetition time (TR) = 2300 ms, echo time (TE) = 2.74 ms, inversion time (TI) = 900 ms, flip angle 8°, field of view (FOV) = 176 × 256 mm, matrix 176 × 256 × 176, voxel size 1 × 1 × 1 mm, pixel bandwidth 190 Hz; total scan time 7 min 37 s) | 6.0.0 | Exclusion criteria included current posttraumatic stress disorder, Past 6 months substance use disorder lifetime bipolar, psychotic, developmental or obsessive–compulsive disorder |
| Milan | Italy | Philips Achieva | 3T | 3D MPRAGE pulse sequence (repetition time (TR) = 2500 ms, echo time (TE) = 2.74 | 6.0.0 | Exclusion criteria for individuals with GAD were |

| | | | | | | |
|---|---|---|---|---|---|---|
| | | | | ms, inversion time (TI) = 900 ms, flip angle 8°, field of view (FOV) = 208 × 256 mm, matrix 208 × 256 × 176, voxel size 1 × 1 × 1 mm, pixel bandwidth 190 Hz; total scan time 2 min 38 s) | | current or past autism spectrum disorders, bipolar disorder, psychosis, or schizophrenia. |
| Münster | Germany | Magnetom PRISMA Siemens | 3T | MPRAGE sequence (TR = 2,130 ms, TE = 2.28 ms, voxel size = 1 mm isotropic, flip angle = 8°) with 192 slices. | 6.0.0 | Exclusion criteria for individuals with GAD were current or past autism spectrum disorders, bipolar disorder, psychosis, or schizophrenia. |
| Protaia | Brazil | GE Healthcare Signa HDxt | 3T | TE/TR=6.13/2.18 ms. FOV: 240 , voxel size 1x1x1 | 6.0.0 | Exclusion criteria for individuals with GAD were current or past autism spectrum disorders, bipolar disorder, psychosis, or schizophrenia. |
| SDAN 2 | U.S. | MR 750 (GE) - 2 scanners (1 included here) | 3T | (MPRAGE) with the following parameters: sagittal acquisition; 176 slices; 256x256 matrix; 1mm3 isotropic voxels; flip angle = 7°; repetition time [TR] = 7.7ms, echo time [TE] = 3.42ms | 6.0.0 | Exclusion criteria for individuals with GAD were current or past autism spectrum disorders, bipolar disorder, psychosis, or schizophrenia. |
| Stonybrook | U.S. | Siemens Trio scanner | 3 T | TR, 1900 ms; TE, 2.53 ms; flip angle, 9°; FOV, 176 × 250 × 250 mm; matrix, 176 × 256 × 256; voxel size, 1 × 1× 1 mm. | 6.0.0 | Exclusion criteria for individuals with GAD were current or past autism spectrum disorders, bipolar disorder, psychosis, or schizophrenia. |
| Stonybrook-klein | U.S. | Siemens | 3T | T1-weighted MPRAGE pulse sequence (TR = 1.9 s, TE = 2.53 ms, flip angle = 9°, slice | 6.0.0 | Exclusion criteria for individuals with GAD were |

| | | | | | | |
|---|---|---|---|---|---|---|
| | | | | thickness = 1 mm, in-plane resolution = 1×1 mm) | | current or past autism spectrum disorders, bipolar disorder, psychosis, or schizophrenia. |
| Sussex | UK | Magnetom Avanto | 1.5T | 176 axial slices, TR=1900ms, TE=3.37ms, TI=1100ms, FA=15°, voxel size=1×1×1mm$^3$, pixel bandwidth=130Hz/px, GRAPPA acceleration factor=2. | 6.0.0 | Exclusion criteria for individuals with GAD were current or past autism spectrum disorders, bipolar disorder, psychosis, or schizophrenia. |
| UCSD | U.S. | Signa EXCITE system / GE | 3 T | SPGR, TI = 450, TR = 8 ms, TE = 3 ms, FOV = 250 × 250 mm3, flip angle = 12°, 172 sagittally acquired slices with 1-mm thickness | 6.0.0 | Exclusion criteria for individuals with GAD were current or past autism spectrum disorders, bipolar disorder, psychosis, or schizophrenia. |
| Upenn (ANXG) | U.S. | Siemens TIM Trio | 3T | MPRAGE TR/TE/TI: 1810/3.5/1100 FOV: 180/240, Matrix 192/256/160, slice thick: 1 mm, Flip Angle 9, GRAPPA: 2 | 6.0.0 | Exclusion criteria for individuals with GAD were current or past autism spectrum disorders, bipolar disorder, psychosis, or schizophrenia. |

**Table S9** *Technical cohort information for the autism cohort.*

| Site | Location | Scanner | Acquisition sequence |
|---|---|---|---|
| **ABIDE 1_Caltech** | USA, Cal | 3T Siemens Trio | Coverage: 256x256, 176 slices, voxel size: 1x1x1mm, TE=2.73ms |
| **ABIDE 1 KKI** | Baltimore, USA | 3T Phillips Achieva | Coverage: 256x256, 200 slices, voxel size: 1x1x1mm, TE=3.7ms |
| **ABIDE 1 Leuven 1** | Leuven, Belgium | 3T Phillips Interna | Coverage: 256x256, 182 slices, voxel size: 1x1x1mm, TE=4.6ms |
| **ABIDE 1 Leuven 2** | Leuven, Belgium | 3T Phillips Interna | Coverage: 256x256, 182 slices, voxel size: 1x1x1mm, TE=4.6ms |
| **ABIDE 1 Max_Mun** | Munich, Germany | 3T Siemens Verio | Coverage: 256x256, 160 slices, voxel size: 1x1x1mm, TE=3.06ms |
| **ABIDE 1 NYU** | New york, NY, USA | 3T Siemens Allegra | Coverage: 256x256, 128 slices, voxel size: 1.3x1x1.3mm, TE=3.25ms |
| **ABIDE 1 Ohsu** | Portland, OR, USA | 3T Siemens Trio | Coverage: 256x256, 160 slices, voxel size: 1x1x1.1mm, TE=3.58ms |
| **ABIDE 1 Olin** | Hartford, CT, USA | 3T Siemens Allegra | Coverage: 256x256, 176 slices, voxel size: 1x1x1mm, TE=2.74ms |
| **ABIDE 1 Pitt** | Pittsburgh, PA, USA | 3T Siemens Allegra | Coverage: 269x269, 176 slices, voxel size: 1x1x1mm, TE=3.93ms |
| **ABIDE 1 sSBL** | Nijmegen, Netherlands | 3T Philips Intera | Coverage: 256x231, 170 slices, voxel size: 1x1x1 |
| **ABIDE 1 SDSU** | San diego, USA | 3T GE MR750 | Coverage: 256x256, 180 slices, voxel size: 1x1x1mm, TE=4.3 |
| **ABIDE 1 Stanford** | Stanford, CA, USA | 3T GR Signa | Coverage: 256x256, 132 slices, voxel size: 0.9x0.9x1mm, TE=1.8 |
| **ABIDE 1 Trinity** | Dublin, Ireland | 3T Philips Achieva | Coverage: 256x256, 160 slices, voxel size: 1x1x1, TE=3.9 |
| **ABIDE 1 UM 1** | Ann Arbor, Michigan, USA | 3T GE Signa | Coverage: 256x256, 128 slices, voxel size: 1x1x1mm, TE=1.8 |
| **ABIDE 1 UM 2** | Ann Arbor, Michigan, USA | 3T GE Signa | Coverage: 256x256, 128 slices, voxel size: 1x1x1mm, TE=1.8 |
| **ABIDE 1 USM** | Salt Lake City, Utah, USA | 3T Siemens Trio | Coverage: 256x240, 192 slices, voxel size: 1x1x1.2mm, TE=2.91 |
| **ABIDE 1 Yale** | New Haven, Connecticut, USA | 3T Siemens Magnetom | Coverage: 256x256, 160 slices, voxel size: 1x1x1mm, TE=1.73 |
| **ABIDE 2 BNI 1** | Phoenix, Arizona, USA | 3T Philips Ingenia | Coverage: 244x227, 170 slices, voxel size: 1.11x1.11x1.1mm, TE=3.1 |
| **ABIDE 2 EMC 1** | Rotterdam, Netherlands | 3T GE MR750 | Coverage: 256x256, 186 slices, voxel: 0.9x0.9x0.9 |
| **ABIDE 2 ETH 1** | Zürich, Switzerland | 3T Philips Achieva | Coverage: 256x256, 162 slices, voxel size: 0.89x0.89x0.89mm, TE=3.9 |
| **ABIDE 2 GU 1** | Washington, DC, USA | 3T Siemens TriTim | Coverage: 256x256, 176 slices, voxel size: 1x1x1mm, TE=3.5 |
| **ABIDE 2 IP 1** | Paris, France | 1.5T Siemens TriTim | Coverage: 256x256, 170 slices, voxel size: 1x1x1mm, TE=5.6 |
| **ABIDE 2 IU 1** | Bloomington, Indiana, USA | 3T Philips Achieva | Coverage: 256x256, 180 slices, voxel size: 0.7x0.7x0.7mm, TE=2.3 |
| **ABIDE 2 KKI 1** | Baltimore, Maryland, USA | 3T Philips Achieva | Coverage: 256x200, 200 slices, voxel size: 1x1x1mm, TE=3.7 |
| **ABIDE 2 NYU 1** | New York, NY, USA | 3T Siemens Allegra | Coverage: 256x256, 128 slices, voxel size: 1.3x1x1.3, TE=3.25 |
| **ABIDE 2 Ohsu 1** | Portland, Oregon, USA | 3T Siemens Skyra | Coverage: 256x256, 160 slices, voxel size: 1x1x1.1, TE=3.58 |
| **ABIDE 2 Oilh 2** | Hartford, Connecticut, USA | 3T Siemens TriTim | Coverage: 256x256, 208 slices, voxel size: 0.8x0.8x0.8, TE=2.88 |
| **ABIDE 2 SDSU 1** | San Diego, CA, USA | 3T GE MR750 | Coverage: 256x256, 176 slices, voxel size: 1x1x1, TE=3.172 |

| | | | |
|---|---|---|---|
| **ABIDE 2 TCD 1** | Dublin, Ireland | 3T Philips Achieva | Coverage: 256x256, 190 slices, voxel size: 0.9x0.9x0.9, TE=3.9 |
| **ABIDE 2 UCD 1** | Davis, California, USA | 3T Siemens TriTim | Coverage: 256x256, 192 slices, voxel size: 1x1x1, TE=3.16 |
| **ABIDE 2 UCLA 1** | Los Angeles, CA; USA | 3T Siemens TriTim | Coverage: 256x240, 160 slices, voxel size: 1x1x1.2, TE=2.86 |
| **ABIDE 2 USM 1** | Salt Lake City, Utah, USA | 3T Siemens TriTim | Coverage: 256x240, 220 slices, voxel size: 1x1x1 |
| **Barcelona** | Barcelona, Spain | | |
| **BRC** | London, UK | 3T GE Signa HDx | Coverage: 256x256, 166 slices, voxel size: 1x1x1 |
| **Cattes** | Nijmegen, The Netherlands | | |
| **CMU** | Pittsburgh, PA, USA | 3T Siemens Magnetom | Coverage: 256x256, voxel size: 1x1x1 |
| **Dresden** | Dresden, Germany | | |
| **Fair** | Portland, OR, USA | 3T Siemens Trio | Coverage: 256x256, 160 slices, voxel size: 1x1x1.1 |
| **Frankfurt** | Frankfurt, Germany | 1.5 T Siemens Sonata | Coverage: 256x240, 160 slices, voxel size: 1x1x1 |
| **FSM** | Pisa, Italy | 1.5T GE Signa | |
| **GENDAAR Harvard 1** | Cambridge, MA, USA | 3T Siemens Trio | Coverage: 256x256, 176 slices, voxel size: 1x1x1 |
| **GENDAAR Seattle 1** | Seattle, WA, USA | 3T Siemens Trio | Coverage: 256x256, 176 slices, voxel size: 1x1x1 |
| **GENDAAR Seattle 2** | Seattle, WA, USA | 3T Siemens Prisma | Coverage: 256x256, 176 slices, voxel size: 1x1x1 |
| **GENDAAR UCLA 1** | Los Angeles, CA, USA | 3T Siemens Trio | Coverage: 256x256, 176 slices, voxel size: 1x1x1 |
| **GENDAAR UCLA 2** | Los Angeles, CA, USA | 3T Siemens Prisma | Coverage: 256x256, 176 slices, voxel size: 1x1x1 |
| **GENDAAR Yale 1** | New Haven, CT, USA | 3T Siemens Trio | Coverage: 256x256, 176 slices, voxel size: 1x1x1 |
| **HGGM** | Madrid, Spain | 1.5T Philips Intera | Coverage: 256x256, 176 slices, voxel size: 1x1x1 |
| **Liehoe / Nijmegen3** | Nijmegen, The Netherlands | 1.5T Siemens Avanto | Coverage: 256x256, 176 slices, voxel size 1x1x1 |
| **MRC 1** | London, UK | 3T GE Signa HDx | Coverage: 256x256, 176 slices, voxel size= 1x1x1 |
| **MRC 2** | London, UK | 3T GE Signa HDx | Coverage: 256x256, 176 slices, voxel size= 1x1x1 |
| **Pitt** | Pittsburgh, PA, USA | 3T Siemens Allegra | Coverage: 256x256, 176 slices, voxel size: 1.05x1.05x1.05, TE=3.9ms |
| **Sao Paulo** | Sao Paulo, Brazil | 3T Philips Achieva | Coverage: 192x192, 160 slices, voxel size=1.36x1.36x1.2 |
| **RATSSSweden** | Stockholm, Sweden | 3T SE | Coverage: 240x240, 176 slices, TR=8.2 s |
| **TCD 2** | Dublin, Ireland | 3T Philips Achieva | Coverage: 256x256, 160 slices, voxel size= 1x1x1 |
| **UMCU 1** | Utrecht, The Netherlands | 1.5T Philips | Coverage: 256x256, 130 slices, voxel size= 1x1x1.5 |
| **Wougro 1/ Nijmegen1** | Nijmegen, The Netherlands | 1.5T Siemens Avanto | Coverage: 256x256, 160 slices, voxel size= 1x1x1 |

**Table S10** *Technical cohort information for the bipolar disorder cohort.*

| Site | Location | Scanner | Acquisition | Slice Orienta-tion | FreeSurfer Version | Diagnostic Interview |
|---|---|---|---|---|---|---|
| Barcelona / FIDMAG | Spain | 1.5T GE Signa | 3D T1-weighted enhanced fast gradient echo (EFGRE3D), matrix size = 512 × 512, 180 contiguous axial slices, voxel resolution = 0.47 × 0.47 × 1mm, no slice gap, TE = 5.19 ms, TR = 12.36 ms and inversion time (TI) = 450ms, flip angle = 20 degrees | Axial | 5.3.0 | Structured Clinical Interview for DSM-IV and Research Diagnostic Criteria (RDC). |
| Cardiff | UK | 3T GE HDx | 3D T1-weighted fast spoiled gradient recall (3D FSPGR), 172 slices, no gap, voxel size: 1x1x1, TE: 3ms, TR: 7.9ms, Flip angle: 20 | Axial | 5.1.0 | Consensus Consultant Diagnosis and Mini International Neuro-psychiatric Interview (MINI) |
| CIAM | South Africa (Cape Town) | 3T Siemens Allegra | 3D T1-weighted magnetization prepared rapid acquisition gradietn echo (MPRAGE); 128 slices, no gap, voxel size: 1.3x1.0x1.3, TE: 1.53/3.21/4.89/6.57ms, TR=2530ms, flip angle: 7 | Sagittal | 5.3.0 | Structured Clinical Interview for DSM-IV for Axis I Diagnoses |
| COGSBD | Australia | 3T SIEMENS Magnetom TrioTim | MP-RAGE, acquisition: saggital, axial slices: 176, 1mm slice thickness, no gap, matrix size 256 x 256, voxel size: 1.0 x 1.0 x 1.0mm3, T1: 900ms, TE: 2.52ms, TR: 1900ms, flip angle: 9 degrees | | 6.0.0 | MINI |
| CYBD | Canada | 3T Phillips | repetition time (TR) 9.5 ms, echo time (TE) 2.3 ms, inversion time (TI) 1400 ms, spatial resolution 0.94 mm × 1.17 mm × 1.2 m m (nearly 1 mm isotropic), 256 × 164 × 140 matrix, flip angle 8°, and scan duration 8 minutes 56 seconds. | | 6.0 | K-SADS-PL |
| For2107 Münster | Münster, Germany | 3T Siemens PRISMA | 3D T1-weighted magnetization prepared rapid acquisition gradient echo (MPRAGE). - Sagittal Acquisition Direction, # of Slices 192, 0mm Slice Gap, 1.0x1.0x1.0 Voxel Size (mm3), TI 900 ms, TE 2.28 ms, TR 1900 ms, Flip Angle 8 | Sagittal | 5.3 | |

| | | | | | | |
|---|---|---|---|---|---|---|
| FOR2107 Marburg | Marburg, Germany | 3T Siemens Magnetom TiroTim syngo MR B17 | 3D T1-weighted magnetization prepared rapid acquisition gradient echo (MPRAGE) - Sagittal Acquisition Direction, # of Slices 176, 0.5mm Slice Gap, 1.0x1.0x1.0 Voxel Size (mm3), TI 900 ms, TE 2.26 ms, TR 1900 ms, Flip Angle 9. | Sagittal | 5.3 | |
| Galway Bipolar Study / NUIG_Galway | Ireland | 1.5T Siemens Magnetom | T1-weighted magnetization prepared rapid gradient echo (MPRAGE) images: field of view (FOV) = 230 mm, repetition time (TR) = 1140 msec, echo time (TE) = 4.38 msec, matrix size 256X256, interpolated to 512X512, yielding an in-plane voxel size of 0.45 mmX0.45 mm2, and slice thickness 0.9 mm. | | 5.1.0 | Structured Clinical Interview for DSMIV-TR-Patient Edition for patients and SCID_NP for controls |
| IDIBAPS - Hospital Clinic | | 3T SIEMENS Prisma | 1mm slice thickness, no gap, matrix size 240*240; 0.9*0.9*1mm3 voxel resolution; TE 3.01ms; TR 2300ms; flip angle 9º | | | Structured Clinical Interview for DSM-IV |
| IGP | Australia | 3T Philips 3T Achieva TX | TR 8.9 ms, TE 4.1 ms, field of view 240 mm, matrix 268 × 268, 200 sagittal slices, slice thickness 0.9 mm (no gap). | Sagittal | 5.3 | Diagnosis was confirmed using the OPCRIT algorithm applied to interviewer ratings on the DIP, according to ICD-10 criteria |
| IMH study / SCDS_IMH | Singapor e | 3T Phillips Achieva | 180 axial slices of 0.9mm thickness with no gap, FOV = 230x230 mm2, matrix 256x204, voxel size =0.89x0.89x0.9 mm3, TR=7.2 s, TE=3.3 ms, FA=8° | Axial | 5.3 | SCID |
| Montpellier | | | | | | |
| SBP | Sweden | 1.5T GE Signa HDtx | 3D T1-weighted spoiled gradient recalled acquisition in steady state, 116 slices, no gap, voxel size: 0.7x0.7x1.8, TE:6ms, TR:21ms, flip angle=30 | | 5.3.0 | The clinical assessment instrument used in SBP was a Swedish version of the Affective Disorder Evaluation [1]. In addition, a structured psychiatric interview (M.I.N.I. International |

| | | | | | | |
|---|---|---|---|---|---|---|
| | | | | | | Neuropsychiatric Interview) covering other psychiatric diagnoses was completed. |
| SBA - Stanford Bipolar Aggregate STOP_M_UBC (STOP-EM First episdoe mania cohort) | USA | 1.5T GE Signa | 3D T1-weighted SPGR, 116 slices, voxel size: .86x.86 x 1.5 m3, TE: 1.7-3.0ms, TR: 8.3-10.1ms, flip angle = 15° | Sagittal | 5.3.0 | Structured Clinical Interview for DSM-IV |
| TAMI[15–17] | Taiwan | 3T Siemens Triomagnetom a tim system | T1-weighted MR scanning were obtained using a sagittal 3D magnetization-prepared rapid gradient echo (MPRAGE) sequence, with the following parameter settings: TE = 3.5 ms; matrix size = 256 × 256; slices = 192; slice thickness = 1 mm; and voxel size = 1.0 × 1.0 × 1.0 mm3. | | 7.4.1 | DSM-IV |
| Upenn (BD) | USA | 3T Siemens Tim Trio | 3D T1-weighted magnetization prepared rapid acquisition gradient echo (MPRAGE), 200 slices, no gap, voxel size: 0.9x0.9x1, TE:3.51ms, TR=1810ms, flip angle = 8 | | | Structured Clinical Interview for DSM-IV for Axis I Diagnoses |
| Rome | Italy | 3T Achieva MR scanner (Philips Medical Systems, Best, The Netherlands) | whole-brain T1-weighted images were obtained using a fast-field echo sequence (echo time/repetition: time = 5.3/11 ms, flip angleec = 9°, voxel sizeec = 1 × 1 × 1 mm3). | | 7.1.1 | SCID |

**Table S11** *Technical cohort information for the MDD cohort.*

| Site | Location | Scanner | Acquisition | Slice Orientation | FreeSurfer Version | Diagnostic Interview |
|---|---|---|---|---|---|---|
| **Barc / Barcelona** | Barcelona, Spain | 3T Philips Achieva | 3D MPRAGE images (Whole-brain T1-weighted); TR=6.7ms, TE=3.2ms; 170 slices, voxel size 0.89X0.89X1.2 mm. Image dimensions 288X288X170; field of view: 256X256X204; slice thickness: 1.2 mm; with a sagittal slice orientation, T1 contrast enhancement, flip angle: 8º, grey matter as a reference tissue, ACQ matrix MXP = 256X240 and turbo-field echo shots (TFE) = 218. | Sagittal | | DSM-IV-TR acc. to CIDI-interview and HAMD |
| **Basel** | Basel, Switzer-land | 3T Siemens Magnetom Prisma | 3D T1-weighted MPRAGE (TR = 2000 ms, TE = 3.37 ms, FOV 256 × 256 mm, voxel size = 1 × 1 × 1 mm, flip angle = 8°) | | | |
| **Csan** | | 3T Siemens MAGNETOM PRISMA | Whole-head t1-weighted MPRAGE (TR = 2300 ms, TE = 2.34 ms, FOV 250 × 250 mm, voxel size = 0.9 × 0.868 × 0.868 mm, flip angle = 8°) | | | M.I.N.I. Neuropsychiatric Interview |
| **Dchs** | Cape Town, South Africa | 3T Siemens Skyra | 3D multi-echo MPRAGE, voxel size 1 mm x 1mm x 1.5mm, TR = 2530 ms, TE = 1.69 x 3.55 x 5.41 x 7.27ms, FOV: 256x256mm, flip angle = 7° | | | MINI |
| **Episca** | Leiden, Netherlands Leiden | 3T Philips Achieva | a sagittal 3-dimensional gradient-echo T1-weighted image was acquired (repetition time = 9.8 ms; echo time = 4.6 ms; flip angle = 8°; 140 sagittal slices; no slice gap; field of view =256 × 256 mm; 1.17 × 1.17 × 1.2 mm voxels; duration = 4:56 min) | Sagittal | 5.3 | |
| **Fidmag** | Spain | 1.5T, GE Signa | matrix size = 512 × 512, 180 contiguous axial slices, voxel resolution = 0.47 × 0.47 × 1mm, no slice gap, TE = 5.19 ms, TR = 12.36 ms and inversion time (TI) = | Axial | 6.0 | |

| | | | | | | |
|---|---|---|---|---|---|---|
| | | | 450ms, flip angle = 20 degrees | | | |
| **For2107 Marburg** | Marburg, Germany | 3T Siemens Magnetom TiroTim syngo MR B17 | 3D T1-weighted magnetization prepared rapid acquisition gradient echo (MPRAGE) - Sagittal Acquisition Direction, # of Slices 176, 0.5mm Slice Gap, 1.0x1.0x1.0 Voxel Size (mm3), TI 900 ms, TE 2.26 ms, TR 1900 ms, Flip Angle 9. | Sagittal | 5.3 | |
| **For2107 Münster** | Münster, Germany | 3T Siemens PRISMA | 3D T1-weighted magnetization prepared rapid acquisition gradient echo (MPRAGE). - Sagittal Acquisition Direction, # of Slices 192, 0mm Slice Gap, 1.0x1.0x1.0 Voxel Size (mm3), TI 900 ms, TE 2.28 ms, TR 1900 ms, Flip Angle 8 | Sagittal | 5.3 | |
| **Hiroshima** | Hiroshima, Japan | 3T Siemens (Spectra, Verio.Dot), 3T GE (Signa HDxt) Site 1 = GE Signa HDxt 3.0T 2= GE Signa HDxt 3.0T 3 = SIEMENS MAGNETOM Spectra 3.0T 4 = SIEMENS MAGNETOM Verio.Dot 3.0T | T1 256x256x256 matrix of 1x1x1mm voxels (Siemens: ADNI MPRAGE (tfl), GRAPPA, 192 slices, GE: SPGR, 184 slices) | | | MINI |
| **IMH / Singapore** | Singapore, Singapore | 3T Phillips Achieva | 180 axial slices of 0.9mm thickness with no gap, FOV = 230x230 mm2, matrix 256x204, voxel size =0.89x0.89x0.9 mm3, TR=7.2 s, TE=3.3 ms, FA=8° | Axial | 5.3 | SCID |
| **Moral dilemma** | | 3T GE Signa Excite | 3D BRAVO sequence: 140 contiguous slices; repetition time, 7900 ms; echo time, 3000 ms; flip angle, 13°; in a 25.6-cm field of view, with a 256 × 256 pixel matrix and a slice | Axial | 5.3 | |

| | | | | | | |
|---|---|---|---|---|---|---|
| | | | thickness of 1 mm (1 mm gap). | | | |
| **Oxford** | Oxford, UK | 3T Siemens Tim Trio | Voxel resolution 0.78 x 0.8 x 0.78 mm on a 208 x 256 x 200 grid, TE/TI/TR= 4.8/1100/2040 ms | | | |
| **Rome** | Rome, Italy | 3T Achieva MR scanner (Philips Medical Systems, Best, The Netherlands) | whole-brain T1-weighted images were obtained using a fast-field echo sequence (echo time/repetition: time = 5.3/11 ms, flip angleec = 9°, voxel sizeec = 1 × 1 × 1 mm3). | | 7.1.1 | SCID |
| **SF / UCSF** | San Francisco, CA, USA | 3T GE MR750 | repetition time/echo time = 8.1 msec/3.17 msec, flip angle = 12°, 256 × 256 matrix, 1 × 1 × 1 mm voxels, 168 sagittal slices | Sagittal | | |
| **SHIP-START-2 and SHIP-TREND-0** | Greifswald, Germany Greifswald | 1.5T Siemens Avanto | 3D T1-weighted (MP-RAGE/ axial plane); TR=1900 msec; TE=3.4 msec; Flip angle=15°; voxel size 1 mm x 1 mm x 1 mm | Axial | FS 5.3 (cortical parameters), FS 5.1 (subcortical volumes) | M-CIDI interview |
| **Stanford T1w Aggregate** | Stanford, USA | 1.5T GE Signa Excite | Whole-brain T1-weighted images were collected using a spoiled gradient echo (SPGR) pulse sequence (116 sagittal slices; through-plane resolution = 1.5 mm; in-plane resolution = 0.86 x 0.86 mm; flip angle = 15 degrees; repetition time [TR] = 8.3-10.1 ms; echo time [TE] = 1.7-3.0; inversion time [TI] = 300 ms; matrix = 256 x 192). | Sagittal | 5.3 | |
| **TAMI**[15–17] | Taiwan | 3T Siemens Triomagnetom a tim system | T1-weighted MR scanning were obtained using a sagittal 3D magnetization-prepared rapid gradient echo (MPRAGE) sequence, with the following parameter settings: TE = 3.5 ms; matrix size = 256 × 256; slices = 192; slice thickness = 1 mm; and voxel size = 1.0 × 1.0 × 1.0 mm3. | Sagittal | 7.4.1 | diagnosed according to the Diagnostic and Statistical Manual of Mental Disorders, Fourth Edition, by two psychiatrists. |
| **Tips / Jena** | Germany | 3T Siemens Prisma fit | Scanner 1 (MP-RAGE): TR 2,300 ms, TE 3.03 ms, α 9°, 192 contiguous sagittal slices, FoV 256 | Sagittal | 5.3 | SCID |

| | | | | | |
|---|---|---|---|---|---|
| | | | mm, voxel resolution 1 × 1 × 1 mm; acquisition time 5:21 min | | |
| **UMN** | Minnesota, USA | 3.0 Tesla Tim Trio scanner; Siemens Corp | A 5-minute structural scan was acquired using a T1-weighted, high-resolution, magnetization-prepared gradient-echo sequence: repetition time, 2530 milliseconds; echo time, 3.65 milliseconds; inversion time, 1100 milliseconds; flip angle, 7°; field of view, 256 × 176 mm; voxel size, 1-mm isotropic; 224 slices; and generalized, autocalibrating, partially parallel acquisition acceleration factor, 2. | | Schedule for Affective Disorders and Schizophrenia for School-Age Children–Present and Lifetime Version and the Children's Depression Rating Scale–Revised (CDRS-R). |

**Table S12** *Technical cohort information for the obsessive-compulsive disorder cohort.*

| Site | Location | Scanner | Acquisition parameters |
|---|---|---|---|
| Benedetti | Milan, Italy | 3T Philips Gyroscan Intera | Matrix 256 x 256, 220 slices, Voxel size 1 x 1 x 1 mm |
| Beucke / Kathmann | Berlin, Germany | 1.5T Siemens Sonata | Matrix 256 x 224, 176 slices, Voxel size = 1mm ISO |
| Brennan | Massachusetts, USA | 3T Siemens TrioTim syngo MR B17 | Matrix 256 x 265, 128 slices, Voxel size = 1.3x1.0x1.3mm |
| Buitelaar / Rooij | Nijmegen, Netherlands | 3T Siemens PrismaFit | Matrix 256x256, 192 slices, 1x1x1 mm |
| Cheng | Kunming, China | 1.5T GE Signa Excite / 3T Philips Achieva | Matrix 256 x 256, 172 slices, Voxel size 0.93 x 0.93 x 0.9 mm/ Matrix 228 x 228, 230 slices, Voxel size 1.1 x 1.1 x 0.6 mm |
| Denys / van Wingen | Kunming, China | 3T Philips Intera | Matrix 256 x 256, 182/180 slices, Voxel size 1.2 x 0.833 x 0.833/ 1 x 0.5 x 0.5 |
| Fitzgerald | Michigan, USA | 3T GE Signa | Matrix 256 x 256, 124 slices, Voxel size 1.02 x 1.02 x 1.2 mm |
| Gruner | Connecticut, USA | 3T GE Signa | Matrix 256 x 256, 216 slices, Voxel size 0.976 x 0.976 x 1.0 mm |
| van den Heuvel | Amsterdam, Netherlands | 1.5T Siemens Sonata / 3T GE Healthcare Signa HDxt | Matrix 256 x 160, 160 slices, Voxel size 1 x 1 x 1.5 mm / Matrix 256 x 256, 172 slices, Voxel size 1 x 0.977 x 0.977 |
| Hirano | Chiba, Japan | 3T GE Discovery MR750 | Matrix 256 x 256, 178 slices, 1 x 1 x 1 mm |
| Hoexter | Sao Paulo, Brazil | 1.5T GE Signa / 3T Philips Achieva | Matrix 256 x 192, 248 slices, Voxel size 0.94 x 0.94 x 0.80 mm / Matrix 240 x 240, 208 slices, Voxel size 1 x 1 x1 mm |
| Huyser | Amsterdam, Netherlands | 3T Philips Intera MR | Matrix 256 x 256, 182 slices, Voxel size 1 x 1 x 1.2 mm |
| James | Oxford, UK | 1.5T Siemens Sonata | Matrix 256 x 256, 208 slices, Voxel size 1 x 1 x 1 mm |
| Koch | München, Germany | 3T Philips Ingenia | Matrix 240 x 240, 170 slices, Voxel size 1 x 1 x 1 mm |
| Kwon | Seoul, South Korea | 1.5T GE Signa / 1.5T Siemens Avanto / 3T Siemens Trio | Matrix 256 x 256, 124 slices, Voxel size 0.82 x 0.82 x 1.5 mm / Matrix 416 x 512, 160-208 slices, Voxel size 0.45 x 0.45 x 0.9 mm / Matrix 256 x 256, 208 slices, Voxel size 1 x 0.977 x 0.977 mm |
| Lázaro | Barcelona, Spain | 1.5T GE Signa LX / 3T Siemens Magnetom Tim | Matrix 256 x 256, 128 slices, Voxel size 1 x 1 x 1 mm / Matrix 256 x 256, 240 slices, Voxel size 1 x 1 x 1 mm |
| Marsh | New York, USA | 3T GE Signa | Matrix 256 x 256, 164 slices, Voxel size 0.976 x 0.976 x 1 mm |
| Mataix-Cols | Stockholm, Sweden | 1.5T GE Signa / 1.5T GE Signa HDx | Matrix 256 x 256, 124 slices, Voxel size 0.94 x 0.94 x 1.5 mm / Matrix 256 x 256, 146 slices, Voxel size 1.09 x 1.09 x 1.1 mm |
| Menchón / Soriano-Mas | Barcelona, Spain | 1.5T GE Signa Excite | Matrix 256 x 256, 130 slices, Voxel size 1.2 x 1.2 x 1.2 mm |
| Morgado | Braga, Portugal | 1.5T Siemens Avanto | Matrix 256 x 256, 176 slices, Voxel size 1 x 1 x 1 mm |
| Nakamae / Abe | Kyoto, Japan | 1.5T Philips Gyroscan Intera / 3T Philips Achieva | Matrix 256 x 256, 130 slices, Voxel size 0.98 x 0.98 x 1.5 mm / Matrix 256 x 256, 170 slices, Voxel size 1.0 x 1.0 x 1.0 mm |
| Nakao | Fukuoka, Japan | 3T Philips Achieva TX | Matrix 240 x 240, 190 slices, Voxel size 1.8 x 1.8 x 1.8 mm |
| Nurmi / O'Neill / Feusner / Piacentini | California, USA | 3T Siemens Trio | Matrix 256 x 256, 176 slices, Voxel size 1.0 x 1.0 x 1.0 mm |

| | | | |
|---|---|---|---|
| Reddy | Bangalore, India | 1.5T Siemens Vision / 3T Siemens Skyra / 3T Philips Achieva | Matrix 256 x 160, 160 slices, Voxel size 0.98 x 0.98 x 1 mm / Matrix 256 x 256, 192 slices, Voxel size 1.0 x 1.0 x 1.0 mm / Matrix 256 x 256, 165 slices, Voxel size 1.0 x 1.0 x 1.0 mm |
| Simpson / Marsh | New York, USA | 3T GE Signa | Matrix 256 x 256, 164 slices, Voxel size 0.976 x 0.976 x 1.0 mm |
| Soreni | Ontario, CAN | 3T GE Excite | Matrix 512 x 512, 148 slices, Voxel size 0.468 x 0.469 x 1 mm |
| Stein / Lochner | Cape Town, South Africa | 3T Siemens Allegra | Matrix 256 x 256, 160 slices, Voxel size 1.3 x 1.0 x 1.0 mm |
| Stern | New York, USA | 3T Siemens Allegra | Matrix 256 x 256, 208 slices, Voxel size 0.82 x 0.82 x 0.82 mm |
| Stewart | British Columbia, Canada | 3T GE Discovery 750 | Matrix 256 x 256, 164 slices, Voxel size 1 x 1 x 1 mm |
| Tolin | Connecticut, USA | Matrix 240 x 240, 160 slices, Voxel size 1 x 1 x 1 mm ISO / Matrix 240 x 240, 160 slices, Voxel size 1 x 1 x 1 mm ISO | |
| Walitza | Zürich, Switzerland | 3T Philips Achieva / 3T Philips Achieva | Matrix 256 x 256, 192 slices, Voxel size 1.0 x 1.0 x 1.0 mm / Matrix 256 x 256, 192 slices, Voxel size 1.0 x 1.0 x 1.0 mm |
| Zhao / Wang | Shanghai, China | 3T Siemens Verio | Matrix 256 x 256, 192 slices, Voxel size 1.0 x 1.0 x 1.0 mm |

**Table S13** *Technical cohort information for the schizophrenia spectrum disorder cohort.*

| Site | Location | Scanner | Acquisition | Slice Orientation | FreeSurfer Version | Diagnostic Interview |
|---|---|---|---|---|---|---|
| **ASRB** | Australia (Brisbane, Melbourne, Newcastle, Perth and Sydney) | Siemens Avanto 1.5T (5 identical scanners) | 176 slices of 1mm thickness, no gap with field-of view 250 x 250 mm2, repetition time 1980 ms, echo time 4.3 ms, data acquisition matrix 256 x 256, with a flip matrix of 15°, resulting in a voxel size of 0.98×0.98×1.0 mm3 | Sagital | v5.1.0 | ICD-10 |
| **CIAM** | South Africa | 3T Siemens Allegra | MPRAGE sequence :TR = 2530 ms, graded TE = 1.53, 3.21, 4.89, 6.57 ms, flip angle = 7°, FOV = 256 mm, slice thickness = 1.33 mm, 128 slices, voxel size 1.3x1.0x1.3, scan time 8:06. Single channel coil used | Sagital | v.5.3.0 | SCID using DSM-IV by clinically trained research team members. Only those participants which made a clear diagnosis of schizophrenia were included in our cohort. |
| **CLING** | Germany | 3T Magnetom TIM Trio | MRI scanning was performed on a 3.0-Tesla Magnetom TIM Trio (Siemens, Erlangen, Germany). A T1-weighted, 3D magnetization prepared rapid gradient echo sequence (MPRAGE) (TR/TE/TI/FA=2250 ms/3.26 ms/900 ms/9°; image matrix = 256 x 256; duration 8 min and 26 sec) was acquired generating 192 sagittal slices with a voxel size of 1 mm3." | Sagittal | v5.3.0 | ICD-10 and DSM-IV |
| **COBRE** | USA | 3T Siemens TIM Trio | T1-weighted images were acquired with a 5-echo multi-echo MPRAGE sequence [TE (echo times) = 1.64, 3.5, 5.36, 7.22, 9.08 ms, TR (repetition time) = 2.53 s, TI (inversion time) = 1.2 s, 7○ flip angle, number of excitations (NEX) = 1, slice thickness = 1 mm, FOV (field of view) = 256 mm, resolution = 256x256 | Sagittal | v5.3.0 | Structured Clinical Interview for DSM-IV Axis I Disorders (SCID) for diagnostic confirmation (consensus was reached by two research psychiatrists using the |

| | | | | | | |
|---|---|---|---|---|---|---|
| | | | | | | SCID-DSM-IV-TR, patient version) and evaluation for co-morbidities. |
| **FIDMAG** | Spain | 1.5T GE Signa | matrix size = 512 × 512, 180 contiguous axial slices, voxel resolution = 0.47 × 0.47 × 1mm, no slice gap, TE = 5.19 ms, TR = 12.36 ms and inversion time (TI) = 450ms, flip angle = 20 degrees | Axial | v5.3.0 | DSM-IV |
| **FOR2107 Marburg** | Germany | 3T Siemens Magnetom TrioTim Syngo | MPRAGE imaging sequence. 1 acquisition. Flip angle: 9 degrees. TE: 2.26 ms. TR: 1900 ms. TI: 900 ms. Acceleration factor: 2Field of view: 256. Image dimensions: 256x256x176 voxels. Voxel size: 1x1x1 mm. | Sagittal | v5.3.0 | DSM-IV-TR using SCID-I |
| **FOR2107 Münster** | Germany | 3T Siemens PRISMA | MPRAGE imaging sequence. 1 acquisition. Flip angle: 8 degrees. TE: 2.28 ms. TR: 2130 ms. TI: 900 ms. Acceleration factor: 2. Field of view: 256. Image dimensions: 256x256x192 voxels. Voxel size: 1.0x1.0x1.0mm. | Sagittal | v.5.3.0 | DSM-IV-TR using SCID-I |
| **HUBIN** | Sweden | 1.5T GE Signa | T1-weighted images, using a threedimensional spoiled gradient recalled (SPGR) pulse sequence, were acquired with the following parameters; 1.5 mm coronal slices, no gap, 35° flip angle, repetition time (TR) = 24 ms, echo time (TE) = 6.0 ms, number of excitations (NEX) = 2, field of view (FOV) = 24 cm, acquisition matrix = 256 × 192. T2-weighted images were acquired with the following parameters; 2.0 mm coronal slices, no gap, TR = 6,000 ms, TE = 84 ms, NEX = 2, FOV = 24 cm, acquisition matrix = 256 × 192. | Coronal | v5.3.0 | DSM-III-R/DSM-IV based on SCID-I and reviews of medical records |
| **IGP** | Australia | 3T Philips Achieva TX | MPRAGE imaging sequence. 200 acquisitions. Flip angle: 8 degrees. TE 4.1 ms. TR 8.9 ms. Field of view 240. Image | Sagittal | v5.3.0 | ICD-10 |

| | | | | | | |
|---|---|---|---|---|---|---|
| | | | dimensions 268x268. Voxel size 0.9x0.9x0.9. | | | |
| **KaSP** | Sweden | 3T GE | 3D IR prep fast SPGR, TR=7.904ms, TE=3.06ms, TI = 450ms, flip angle =12, 146 slices, voxel size = 0.934 x 0.934 x 1.2 mm3, matrix = 256 x 256 | Sagittal | v5.3.0 | SCID-I |
| **MCIC** | USA | 1.5 Siemens and 3T GE - 2 scanners | TR = 2530 ms for 3 T, TR = 12 ms for 1.5 T; TE =3.79 ms for 3 T, TE = 4.76 ms for 1.5 T; FA = 7 for 3 T, FA = 20 for 1.5 T; TI = 1100 for 3 T; Bandwidth = 181 for 3 T, Bandwidth = 110 for 1.5 T; 0.625×0.625 mm voxel size; slice thickness 1.5 mm; FOV 256×256×128 cm matrix; FOV = 16 cm (could be increased to 18 cm when needed for full brain coverage). | Coronal | v4.0.1 | A Structured Clinical Interview for DSM-IV (SCID/SCID-NP for controls) or the Comprehensive Assessment of Symptoms and History (CASH) were used to diagnose primary and co-morbid psychiatric disorders in controls and patients. |
| **MPRC1** | USA | 3T Siemens Allegro | T1-weighted, 3D MPRAGE, 1x1x1mm, TE/TR/TI=4.3/2500/1000ms, flip angle=8 degrees. | Sagittal | v5.3.0 | Patients at the MPRC clinic; Clinical interview with SCID DSM-IV. |
| **NU** | USA | 1.5T Vision | 1) 3D turbo-FLASH: TR=20 ms, TE=5.4 ms, flip=30˚, ACQ=1, 256x256 matrix, 1xl mm in-plane resolution, 180 slices, slice thickness 1 mm, 13:30 min scan time and 2) 3D MPRAGE (2-4repeats): TR=9.7 ms, TE=4 ms, flip=10˚, ACQ=1, 256x256 matrix, 1xl mm in-plane resolution, 128 slices, slice thickness 1.25 mm, 5:36 min scan time each | Axial | v5.3.0 | |
| **PAFIP3T** | Spain | 3T General Electric | Three-dimensional T1-weighted images, using a spoiled grass (SPGR) sequence acquired in the coronal plane with: echo time (TE)=5 ms, repetition time (TR)=24 ms, numbers of excitations (NEX)=2, rotation | Coronal | v5.0.0 | Diagnosis of schizophrenia was confirmed using the Structured Clinical Interview for DSM-IV (SCID-I). |

| | | | | | | |
|---|---|---|---|---|---|---|
| | | | angle=45°, field of view (FOV)=26×19.5 cm, slice thickness=1.5mm and a matrix of 256×192. | | | |
| **RomeSL** | Italy | 3T Siemens | T1-weighted, 3D MDEFT, 1x1x1xmm, TE/TR =2.4/7.92 ms, flip angle=15 | Sagittal | 6.0dev | MPRAGE imaging sequence. T1-weighted, 3D MDEFT, 1x1x1xmm, TE/TR =2.4/7.92 ms, flip angle=15. TE: 910 ms. Acceleration factor: 1. Field of view: 256. Image dimensions: 176x224x256 voxels. |
| **RSCZ** | Russia | 3T Philips Achieva | A turbo field echo sequence covering the whole brain. TR = 8,200 ms, TE = 3.7 ms, TI = 1,020 ms, flip angle = 8, SENSE factor = 1.5, FOV = 240 mm, voxel size of 0.83 × 0.83 mm with a slice thickness of 1 mm, no gap. | Sagittal | v5.3.0 | ICD-10 |
| **SCORE** | Switzerland | 3T Philips Achieva | MPRAGE: aquisition matrix: 256×256×176, isotropic spatial resolution: 1x1x1mm3, TI=1000ms, TR=2s, TE=3.4 ms, flip angle: 8° and bandwidth of 200 Hz/pixel | Sagittal | 6.0dev | DSM-IV |
| **TOP sTOP** | Norway | 3T GE Signa HDxT and 3T GE Discovery GE750 | TOPge750: repetition time (TR) 8.2 ms, echo time (TE) 3.2 ms, flip angle 12°, slice thickness 1.0 mm, 192 saggital slices; TOP3T: repetition time (TR) 7.8 ms, echo time (TE) 2.9 ms, flip angle 12°, slice thickness 1.2 mm, 166 saggital slices | Sagittal | v4.5.0 | SCID-IV |
| **TAMI**[15–17] | Taiwan (R. O. C.) | 3T Siemens Triomagnetom a tim system | T1-weighted MR scanning were obtained using a sagittal 3D magnetization-prepared rapid gradient echo (MPRAGE) sequence, with the following parameter settings: TE = 3.5 ms; matrix | Sagittal | v7.4.1 | diagnosed according to the Diagnostic and Statistical Manual of Mental Disorders, Fourth Edition, |

| | | | | | | |
|---|---|---|---|---|---|---|
| | | | size = 256 × 256; slices = 192; slice thickness = 1 mm; and voxel size = 1.0 × 1.0 × 1.0 mm3. | | | by two psychiatrists. |
| **UCISZ** | USA | 3T Philips Achieva | High-resolution structural imaging scans were acquired on on a 3T Philips Achieve using a T1 Turbo Fast Spin Echo (TFE) with 200 sagittal slices, 320x274 matrix size, 0.75mm3 isotropic voxels, TR = 11ms, TE =4.562ms, flip angle = 18°, Turbo = 180. | Sagittal | v6.0dev | SCID-I/P (DSM-IV-TR) |
| **UNINA** | Italy | 3T Siemens Trio | 3D T1-weighted Magnetization Prepared Rapid Acquisition Gradient Echo sequence (MPRAGE; TR=1900 ms; TE=3.4 ms; TI=900 ms; Flip Angle=9°; resolution=1x1x1 mm3; 160 axial slices) | Axial | v5.3.0 | PANSS |
| **UPenn (SCZ)** | USA | Siemens 3T | MPRAGE, TR=1810 ms, TE= 3.51 ms, TI=1100 ms, flip angle 9, FOV= 240 x 180 mm, matrix= 256 × 192, resolution = 0.9 x 0.9 mm, slices = 160, slice/skip thickness = 1 mm/0 mm | Axial | v.5.3.0 | SCID |
| **Voices** | Australia (Melbourne ) | 3T Siemens Tim Trio | 176 sagittal slices/brain of 1 mm thickness without gap; field of view = 256 × 256 mm2; repetition time/echo time = 1900/2.52 ms; data matrix size = 256 × 256; voxel dimensions = 1.0 × 1.0 × 1.0 mm3. All scans were conducted at a single site | Sagittal | v6.0.0 | MINI |
| **Zurich** | Switzerland | 3T Phillips | 3D T1-weighted images were acquired with an ultra fast gradient echo T1-weighted sequence(TR=8.4ms, TE=3.8ms, flip angle=8°) in 160 sagittal plan slices (1mm slice thickness, no slice gap) of 240×240mm2 resulting in 1x1x1mm3voxels. | Sagittal | v6.0.0 | structured Mini-International Neuropsychiatric Interview (MINI) for DSM-IV |

**Table S14** *Train and test sub-sample demographics.*

RC = Reference cohort, ANXG = generalized anxiety disorder, ASD = autism spectrum diagnosis, BP = bipolar disorder, MDD = major depressive disorder, OCD = obsessive-compulsive disorder, SCZ = Schizophrenia spectrum.

| | *N* | Age | % female | *N* sites |
|---|---|---|---|---|
| **Training** | | | | |
| **RC** | 9532 | 30.77 (+/- 15.76) | 47.84 | 172 |
| **RC Test** | | | | |
| **RC** | 2466 | 30.01 (+/- 15.45) | 47.61 | 172 |
| **NDPC Test** | | | | |
| **ANXG** | 765 | 26.32 (+/-12.05) | 40.00 | 18 |
| **ASD** | 1556 | 16.03 (+/-8.48) | 18.44 | 56 |
| **BP** | 1370 | 38.37 (+/-13.16) | 58.69 | 19 |
| **MDD** | 1916 | 39.89 (+/-15.38) | 64.77 | 24 |
| **OCD** | 1775 | 28.24 (+/-11.49) | 50.14 | 38 |
| **SCZ** | 2753 | 35.29 (+/- 11.84) | 37.60 | 31 |

**Table S15** *Batch effects in raw data and deviation (z) scores.*

| | **Raw CT** | **CT Z-scores** | **Raw SA** | **SA Z-scores** | **Raw SV** | **SV Z-scores** |
|---|---|---|---|---|---|---|
| Mean ICC | 0.20 | 0 | 0.06 | 0 | 0.15 | 0 |
| Median ICC | 0.21 | 0 | 0.05 | 0 | 0.12 | 0 |
| ICC < 0.05 (%) | 0.0 | 100 | 51.5 | 100 | 0 | 100 |
| ICC < 0.10 (%) | 1.50 | 100 | 88.2 | 100 | 35.7 | 100 |
| Mean F-statistic | 45.63 | 0.14 | 12.48 | 0.30 | 31.21 | 0.16 |
| Median F-statistic | 47.26 | 0.12 | 9.72 | 0.30 | 23.33 | 0.15 |
| Mean Eta² | 0.44 | 0.0025 | 0.18 | 0.006 | 0.34 | 0.0028 |
| p < 0.05 (%) | 100.0 | 0 | 100 | 0 | 100 | 0 |

**Table S16** *Group differences in cortical thickness between individuals with a neurodevelopmental and psychiatric condition (NDPC) and reference comparators.*

Cohen's *d* maps were computed between individuals with and without NDPCs. Reference comparators per NDPC were matched via propensity score matching, taking age and sex into account. As tests were based on deviation scores which were intrinsically corrected for age, sex, and site in the normative modelling step, these covariates were not included as covariates in t-tests / Cohen's *d* computation. FDR correction was performed at an alpha level <0.05 for each effect map. ANXG = generalized anxiety disorder, ASD = autism spectrum diagnosis, BP = bipolar disorder, MDD = major depressive disorder, OCD = obsessive-compulsive disorder, SCZ = Schizophrenia spectrum.

| **Structure** | **ANXG** | | **ASD** | | **BP** | | **MDD** | | **OCD** | | **SCZ** | |
|---|---|---|---|---|---|---|---|---|---|---|---|---|
| | ***d*** | **$P_{FDR}$** | ***d*** | **$P_{FDR}$** | ***d*** | **$P_{FDR}$** | ***d*** | **$P_{FDR}$** | ***d*** | **$P_{FDR}$** | ***d*** | **$P_{FDR}$** |
| **L bankssts** | 0 | 0.95 | -0.07 | 0.11 | -0.15 | 0 | -0.04 | 0.53 | -0.08 | 0.06 | -0.35 | 0 |
| **L caudalanteriorcingulate** | -0.14 | 0.07 | -0.02 | 0.66 | -0.05 | 0.18 | -0.01 | 0.93 | 0.04 | 0.29 | -0.14 | 0 |
| **L caudalmiddlefrontal** | -0.03 | 0.71 | -0.05 | 0.26 | -0.13 | 0 | 0.04 | 0.5 | -0.12 | 0 | -0.45 | 0 |
| **L cuneus** | 0.03 | 0.71 | -0.04 | 0.4 | -0.13 | 0 | -0.07 | 0.19 | -0.1 | 0.01 | -0.26 | 0 |
| **L entorhinal** | -0.02 | 0.81 | -0.22 | 0 | -0.04 | 0.25 | -0.03 | 0.59 | -0.1 | 0.02 | -0.13 | 0 |
| **L fusiform** | -0.05 | 0.68 | -0.15 | 0 | -0.2 | 0 | -0.09 | 0.06 | -0.07 | 0.09 | -0.44 | 0 |
| **L inferiorparietal** | -0.05 | 0.58 | -0.04 | 0.33 | -0.19 | 0 | -0.04 | 0.5 | -0.15 | 0 | -0.46 | 0 |
| **L inferiortemporal** | -0.12 | 0.11 | -0.15 | 0 | -0.17 | 0 | -0.03 | 0.59 | -0.09 | 0.03 | -0.43 | 0 |
| **L isthmuscingulate** | 0.06 | 0.57 | -0.01 | 0.9 | -0.15 | 0 | -0.05 | 0.43 | -0.05 | 0.2 | -0.26 | 0 |
| **L lateraloccipital** | -0.08 | 0.38 | -0.07 | 0.11 | -0.23 | 0 | -0.05 | 0.42 | -0.11 | 0 | -0.35 | 0 |
| **L lateralorbitofrontal** | -0.08 | 0.36 | 0 | 0.96 | -0.18 | 0 | -0.03 | 0.59 | -0.06 | 0.15 | -0.37 | 0 |
| **L lingual** | 0.01 | 0.82 | -0.05 | 0.28 | -0.22 | 0 | -0.05 | 0.43 | -0.09 | 0.03 | -0.32 | 0 |
| **L medialorbitofrontal** | -0.09 | 0.36 | 0.04 | 0.38 | -0.22 | 0 | -0.07 | 0.2 | -0.1 | 0.02 | -0.21 | 0 |
| **L middletemporal** | -0.03 | 0.71 | -0.17 | 0 | -0.25 | 0 | -0.06 | 0.26 | -0.14 | 0 | -0.46 | 0 |
| **L parahippocampal** | 0.07 | 0.45 | -0.1 | 0.02 | -0.1 | 0.02 | 0 | 0.94 | -0.03 | 0.46 | -0.21 | 0 |
| **L paracentral** | -0.05 | 0.61 | -0.06 | 0.13 | -0.07 | 0.07 | -0.05 | 0.43 | -0.04 | 0.34 | -0.35 | 0 |
| **L parsopercularis** | -0.04 | 0.71 | -0.06 | 0.14 | -0.25 | 0 | -0.09 | 0.08 | -0.14 | 0 | -0.45 | 0 |
| **L parsorbitalis** | -0.03 | 0.71 | -0.03 | 0.52 | -0.22 | 0 | -0.03 | 0.57 | -0.07 | 0.07 | -0.41 | 0 |
| **L parstriangularis** | -0.02 | 0.77 | 0.06 | 0.2 | -0.3 | 0 | -0.04 | 0.48 | -0.07 | 0.09 | -0.36 | 0 |
| **L pericalcarine** | -0.02 | 0.77 | -0.04 | 0.3 | -0.09 | 0.02 | 0.05 | 0.43 | -0.02 | 0.57 | -0.05 | 0.1 |
| **L postcentral** | -0.14 | 0.07 | -0.1 | 0.02 | -0.08 | 0.04 | 0 | 0.99 | -0.07 | 0.09 | -0.45 | 0 |
| **L posteriorcingulate** | -0.01 | 0.81 | 0.01 | 0.78 | -0.13 | 0 | -0.04 | 0.48 | -0.06 | 0.12 | -0.29 | 0 |
| **L precentral** | -0.04 | 0.71 | -0.17 | 0 | -0.15 | 0 | -0.06 | 0.35 | -0.04 | 0.33 | -0.46 | 0 |
| **L precuneus** | -0.08 | 0.37 | -0.05 | 0.28 | -0.18 | 0 | -0.07 | 0.18 | -0.14 | 0 | -0.41 | 0 |
| **L rostralanteriorcingulate** | -0.14 | 0.07 | -0.05 | 0.25 | -0.07 | 0.06 | -0.03 | 0.63 | -0.04 | 0.33 | -0.13 | 0 |
| **L rostralmiddlefrontal** | -0.11 | 0.14 | 0.04 | 0.33 | -0.23 | 0 | 0.02 | 0.74 | -0.1 | 0.02 | -0.42 | 0 |
| **L superiorfrontal** | -0.07 | 0.39 | 0 | 0.99 | -0.2 | 0 | 0.01 | 0.9 | -0.07 | 0.1 | -0.49 | 0 |
| **L superiorparietal** | -0.07 | 0.41 | -0.06 | 0.2 | -0.18 | 0 | -0.05 | 0.42 | -0.13 | 0 | -0.35 | 0 |
| **L superiortemporal** | -0.06 | 0.58 | -0.12 | 0 | -0.18 | 0 | -0.01 | 0.92 | -0.02 | 0.58 | -0.44 | 0 |
| **L supramarginal** | -0.04 | 0.71 | -0.06 | 0.17 | -0.23 | 0 | 0 | 0.99 | -0.12 | 0 | -0.47 | 0 |
| **L frontalpole** | -0.12 | 0.11 | 0.07 | 0.1 | -0.07 | 0.07 | 0.05 | 0.43 | -0.01 | 0.85 | -0.23 | 0 |

| | | | | | | | | | | | | |
|---|---|---|---|---|---|---|---|---|---|---|---|---|
| **L temporalpole** | -0.08 | 0.37 | -0.18 | 0 | -0.13 | 0 | -0.02 | 0.74 | 0 | 0.92 | -0.24 | 0 |
| **L transversetemporal** | 0.04 | 0.71 | -0.09 | 0.03 | -0.09 | 0.02 | -0.01 | 0.93 | 0 | 0.92 | -0.27 | 0 |
| **L insula** | -0.14 | 0.07 | -0.05 | 0.21 | -0.13 | 0 | -0.07 | 0.14 | -0.04 | 0.33 | -0.39 | 0 |
| **R bankssts** | 0 | 0.95 | -0.09 | 0.04 | -0.08 | 0.04 | -0.09 | 0.05 | -0.07 | 0.09 | -0.37 | 0 |
| **R caudalanteriorcingulate** | 0.02 | 0.77 | 0 | 0.96 | -0.01 | 0.82 | 0.02 | 0.74 | 0.02 | 0.53 | -0.15 | 0 |
| **R caudalmiddlefrontal** | -0.03 | 0.71 | 0.01 | 0.94 | -0.14 | 0 | -0.03 | 0.62 | -0.09 | 0.03 | -0.4 | 0 |
| **R cuneus** | 0.02 | 0.77 | 0 | 0.97 | -0.15 | 0 | -0.04 | 0.55 | -0.11 | 0.01 | -0.28 | 0 |
| **R entorhinal** | -0.04 | 0.71 | -0.22 | 0 | -0.09 | 0.02 | -0.02 | 0.65 | -0.04 | 0.33 | -0.09 | 0 |
| **R fusiform** | -0.13 | 0.07 | -0.21 | 0 | -0.22 | 0 | -0.11 | 0.05 | -0.07 | 0.09 | -0.48 | 0 |
| **R inferiorparietal** | 0.03 | 0.71 | 0.01 | 0.79 | -0.15 | 0 | 0.02 | 0.73 | -0.15 | 0 | -0.41 | 0 |
| **R inferiortemporal** | -0.03 | 0.71 | -0.18 | 0 | -0.13 | 0 | -0.03 | 0.6 | -0.04 | 0.32 | -0.39 | 0 |
| **R isthmuscingulate** | -0.04 | 0.68 | 0.01 | 0.94 | -0.23 | 0 | -0.08 | 0.14 | -0.03 | 0.4 | -0.29 | 0 |
| **R lateraloccipital** | -0.05 | 0.61 | -0.07 | 0.11 | -0.2 | 0 | -0.03 | 0.63 | -0.09 | 0.03 | -0.35 | 0 |
| **R lateralorbitofrontal** | -0.1 | 0.26 | 0.03 | 0.54 | -0.21 | 0 | 0.03 | 0.6 | -0.07 | 0.07 | -0.31 | 0 |
| **R lingual** | -0.07 | 0.45 | -0.08 | 0.06 | -0.25 | 0 | -0.08 | 0.08 | -0.08 | 0.05 | -0.34 | 0 |
| **R medialorbitofrontal** | -0.11 | 0.14 | 0.14 | 0 | -0.27 | 0 | 0.03 | 0.6 | -0.08 | 0.04 | -0.22 | 0 |
| **R middletemporal** | -0.08 | 0.36 | -0.14 | 0 | -0.25 | 0 | -0.1 | 0.05 | -0.06 | 0.11 | -0.39 | 0 |
| **R parahippocampal** | -0.02 | 0.81 | -0.14 | 0 | -0.17 | 0 | -0.1 | 0.05 | -0.06 | 0.12 | -0.26 | 0 |
| **R paracentral** | -0.02 | 0.77 | -0.09 | 0.03 | -0.1 | 0.01 | -0.02 | 0.71 | -0.02 | 0.53 | -0.32 | 0 |
| **R parsopercularis** | -0.02 | 0.77 | -0.04 | 0.41 | -0.2 | 0 | -0.01 | 0.77 | -0.08 | 0.05 | -0.39 | 0 |
| **R parsorbitalis** | 0.02 | 0.81 | 0.09 | 0.03 | -0.17 | 0 | 0.02 | 0.67 | -0.05 | 0.19 | -0.35 | 0 |
| **R parstriangularis** | -0.03 | 0.71 | 0.02 | 0.63 | -0.21 | 0 | -0.01 | 0.9 | -0.06 | 0.13 | -0.38 | 0 |
| **R pericalcarine** | -0.06 | 0.57 | -0.04 | 0.41 | -0.06 | 0.14 | 0.02 | 0.74 | 0.01 | 0.8 | -0.05 | 0.07 |
| **R postcentral** | -0.15 | 0.07 | -0.08 | 0.08 | -0.1 | 0.01 | -0.04 | 0.48 | -0.04 | 0.28 | -0.42 | 0 |
| **R posteriorcingulate** | -0.03 | 0.71 | 0.09 | 0.03 | -0.09 | 0.02 | -0.02 | 0.74 | -0.02 | 0.7 | -0.31 | 0 |
| **R precentral** | -0.07 | 0.39 | -0.15 | 0 | -0.12 | 0 | -0.05 | 0.42 | -0.04 | 0.28 | -0.41 | 0 |
| **R precuneus** | -0.08 | 0.36 | -0.07 | 0.11 | -0.2 | 0 | -0.02 | 0.74 | -0.14 | 0 | -0.38 | 0 |
| **R rostralanteriorcingulate** | -0.11 | 0.16 | 0.02 | 0.66 | -0.03 | 0.52 | 0.02 | 0.67 | 0.01 | 0.83 | -0.09 | 0 |
| **R rostralmiddlefrontal** | -0.08 | 0.36 | 0.12 | 0 | -0.17 | 0 | 0.07 | 0.21 | -0.09 | 0.02 | -0.37 | 0 |
| **R superiorfrontal** | -0.04 | 0.71 | 0.08 | 0.06 | -0.19 | 0 | 0.02 | 0.71 | -0.05 | 0.17 | -0.45 | 0 |
| **R superiorparietal** | -0.06 | 0.57 | -0.07 | 0.11 | -0.13 | 0 | 0 | 0.99 | -0.14 | 0 | -0.36 | 0 |
| **R superiortemporal** | -0.18 | 0.03 | -0.1 | 0.02 | -0.19 | 0 | -0.06 | 0.26 | 0 | 0.98 | -0.42 | 0 |
| **R supramarginal** | 0.02 | 0.78 | -0.08 | 0.05 | -0.12 | 0 | -0.03 | 0.6 | -0.05 | 0.21 | -0.47 | 0 |
| **R frontalpole** | -0.13 | 0.07 | 0.09 | 0.03 | -0.11 | 0.01 | 0.04 | 0.55 | 0.08 | 0.05 | -0.16 | 0 |
| **R temporalpole** | -0.08 | 0.36 | -0.19 | 0 | -0.11 | 0.01 | -0.02 | 0.74 | -0.05 | 0.19 | -0.21 | 0 |
| **R transversetemporal** | 0.03 | 0.71 | -0.1 | 0.02 | -0.11 | 0 | -0.04 | 0.49 | -0.03 | 0.46 | -0.32 | 0 |
| **R insula** | -0.14 | 0.07 | -0.15 | 0 | -0.13 | 0 | -0.1 | 0.05 | -0.08 | 0.05 | -0.38 | 0 |

**Table S17** *Group differences in surface area between individuals with a neurodevelopmental and psychiatric condition (NDPC) and reference comparators.*

Cohen's *d* maps were computed between individuals with and without NDPCs. Reference comparators per NDPC were matched via propensity score matching, taking age and sex into account. As tests were based on deviation scores which were intrinsically corrected for age, sex, and site in the normative modelling step, these covariates were not included as covariates in t-tests / Cohen's *d* computation. FDR correction was performed at an alpha level <0.05 for each effect map. ANXG = generalized anxiety disorder, ASD = autism spectrum diagnosis, BP = bipolar disorder, MDD = major depressive disorder, OCD = obsessive-compulsive disorder, SCZ = Schizophrenia spectrum.

| **Structure** | **ANXG** | | **ASD** | | **BP** | | **MDD** | | **OCD** | | **SCZ** | |
|---|---|---|---|---|---|---|---|---|---|---|---|---|
| | ***d*** | **$P_{FDR}$** | ***d*** | **$P_{FDR}$** | ***d*** | **$P_{FDR}$** | ***d*** | **$P_{FDR}$** | ***d*** | **$P_{FDR}$** | ***d*** | **$P_{FDR}$** |
| **L bankssts** | -0.2 | 0 | -0.11 | 0.05 | -0.06 | 0.43 | -0.15 | 0 | -0.06 | 0.13 | -0.2 | 0 |
| **L caudalanteriorcingulate** | -0.19 | 0 | 0.04 | 0.65 | 0.03 | 0.71 | -0.06 | 0.08 | -0.06 | 0.1 | -0.12 | 0 |
| **L caudalmiddlefrontal** | -0.2 | 0 | 0 | 1 | 0.04 | 0.64 | -0.08 | 0.02 | -0.07 | 0.08 | -0.14 | 0 |
| **L cuneus** | -0.08 | 0.19 | 0.03 | 0.77 | -0.01 | 0.93 | -0.07 | 0.03 | -0.08 | 0.04 | -0.15 | 0 |
| **L entorhinal** | 0.05 | 0.44 | 0.01 | 1 | -0.07 | 0.37 | -0.12 | 0 | -0.07 | 0.06 | -0.09 | 0 |
| **L fusiform** | -0.32 | 0 | -0.02 | 0.95 | -0.07 | 0.37 | -0.1 | 0 | -0.06 | 0.11 | -0.23 | 0 |
| **L inferiorparietal** | -0.19 | 0 | 0 | 1 | -0.07 | 0.37 | -0.12 | 0 | -0.05 | 0.18 | -0.2 | 0 |
| **L inferiortemporal** | -0.13 | 0.02 | -0.01 | 1 | -0.04 | 0.64 | -0.07 | 0.03 | -0.09 | 0.02 | -0.19 | 0 |
| **L isthmuscingulate** | -0.1 | 0.08 | 0.14 | 0.01 | 0.01 | 0.94 | -0.06 | 0.1 | -0.06 | 0.12 | -0.05 | 0.05 |
| **L lateraloccipital** | -0.03 | 0.61 | 0.06 | 0.36 | -0.07 | 0.37 | -0.1 | 0 | -0.12 | 0 | -0.18 | 0 |
| **L lateralorbitofrontal** | -0.1 | 0.09 | -0.06 | 0.38 | -0.01 | 0.98 | -0.11 | 0 | -0.12 | 0 | -0.2 | 0 |
| **L lingual** | -0.04 | 0.5 | 0.01 | 1 | -0.06 | 0.37 | -0.06 | 0.06 | -0.06 | 0.1 | -0.17 | 0 |
| **L medialorbitofrontal** | -0.06 | 0.35 | 0 | 1 | 0 | 1 | -0.09 | 0.01 | -0.07 | 0.08 | -0.12 | 0 |
| **L middletemporal** | -0.06 | 0.38 | -0.09 | 0.07 | -0.04 | 0.59 | -0.12 | 0 | -0.1 | 0.01 | -0.18 | 0 |
| **L parahippocampal** | -0.26 | 0 | 0.1 | 0.05 | -0.09 | 0.29 | -0.17 | 0 | 0.04 | 0.3 | -0.17 | 0 |
| **L paracentral** | -0.16 | 0.01 | 0.11 | 0.04 | -0.05 | 0.45 | -0.12 | 0 | -0.07 | 0.06 | -0.1 | 0 |
| **L parsopercularis** | -0.05 | 0.38 | 0.05 | 0.45 | 0 | 0.98 | -0.08 | 0.02 | -0.11 | 0 | -0.14 | 0 |
| **L parsorbitalis** | -0.07 | 0.26 | -0.04 | 0.64 | -0.05 | 0.49 | -0.06 | 0.07 | -0.1 | 0.01 | -0.19 | 0 |
| **L parstriangularis** | -0.04 | 0.49 | -0.03 | 0.8 | -0.04 | 0.64 | -0.07 | 0.04 | -0.15 | 0 | -0.12 | 0 |
| **L pericalcarine** | 0.01 | 0.88 | -0.02 | 1 | -0.07 | 0.37 | -0.1 | 0 | -0.08 | 0.03 | -0.13 | 0 |
| **L postcentral** | -0.25 | 0 | 0.03 | 0.84 | -0.02 | 0.87 | -0.16 | 0 | -0.14 | 0 | -0.2 | 0 |
| **L posteriorcingulate** | -0.13 | 0.02 | 0.02 | 0.99 | -0.01 | 0.92 | -0.05 | 0.13 | -0.09 | 0.02 | -0.11 | 0 |
| **L precentral** | -0.18 | 0 | 0.07 | 0.26 | -0.05 | 0.49 | -0.11 | 0 | -0.08 | 0.03 | -0.16 | 0 |
| **L precuneus** | -0.14 | 0.02 | 0.04 | 0.65 | -0.01 | 0.92 | -0.08 | 0.02 | -0.06 | 0.11 | -0.09 | 0 |
| **L rostralanteriorcingulate** | -0.09 | 0.12 | -0.01 | 1 | 0.09 | 0.29 | -0.04 | 0.19 | -0.05 | 0.18 | -0.13 | 0 |
| **L rostralmiddlefrontal** | -0.16 | 0.01 | 0 | 1 | -0.02 | 0.87 | -0.13 | 0 | -0.11 | 0 | -0.2 | 0 |
| **L superiorfrontal** | -0.1 | 0.09 | 0.01 | 1 | -0.02 | 0.87 | -0.09 | 0.01 | -0.14 | 0 | -0.21 | 0 |
| **L superiorparietal** | -0.15 | 0.01 | 0.05 | 0.45 | -0.04 | 0.54 | -0.08 | 0.03 | -0.13 | 0 | -0.14 | 0 |
| **L superiortemporal** | -0.14 | 0.02 | -0.08 | 0.17 | -0.02 | 0.87 | -0.12 | 0 | -0.14 | 0 | -0.19 | 0 |
| **L supramarginal** | -0.11 | 0.07 | -0.03 | 0.8 | -0.04 | 0.57 | -0.1 | 0 | -0.15 | 0 | -0.15 | 0 |
| **L frontalpole** | -0.04 | 0.49 | 0 | 1 | 0 | 0.99 | 0 | 0.98 | -0.05 | 0.16 | -0.09 | 0 |

| | | | | | | | | | | | | |
|---|---|---|---|---|---|---|---|---|---|---|---|---|
| **L temporalpole** | -0.15 | 0.01 | 0.02 | 0.98 | -0.02 | 0.87 | 0.01 | 0.88 | 0.01 | 0.82 | -0.02 | 0.43 |
| **L transversetemporal** | -0.15 | 0.01 | 0.01 | 1 | -0.06 | 0.37 | -0.12 | 0 | -0.19 | 0 | -0.2 | 0 |
| **L insula** | -0.01 | 0.8 | 0.02 | 0.99 | 0.05 | 0.52 | -0.07 | 0.04 | -0.16 | 0 | -0.15 | 0 |
| **R bankssts** | -0.13 | 0.02 | -0.01 | 1 | -0.07 | 0.37 | -0.15 | 0 | 0.01 | 0.82 | -0.18 | 0 |
| **R caudalanteriorcingulate** | -0.17 | 0.01 | -0.02 | 1 | -0.03 | 0.73 | -0.06 | 0.06 | -0.09 | 0.02 | -0.19 | 0 |
| **R caudalmiddlefrontal** | -0.11 | 0.06 | 0 | 1 | 0 | 0.98 | -0.1 | 0 | -0.07 | 0.06 | -0.15 | 0 |
| **R cuneus** | -0.06 | 0.38 | 0.01 | 1 | -0.09 | 0.29 | -0.05 | 0.12 | -0.12 | 0 | -0.11 | 0 |
| **R entorhinal** | -0.04 | 0.5 | 0.02 | 0.99 | 0.04 | 0.64 | -0.07 | 0.05 | -0.06 | 0.12 | -0.06 | 0.04 |
| **R fusiform** | -0.23 | 0 | 0 | 1 | -0.11 | 0.29 | -0.16 | 0 | -0.1 | 0.01 | -0.25 | 0 |
| **R inferiorparietal** | -0.17 | 0.01 | 0 | 1 | -0.1 | 0.29 | -0.16 | 0 | -0.04 | 0.25 | -0.18 | 0 |
| **R inferiortemporal** | -0.12 | 0.03 | -0.04 | 0.64 | -0.06 | 0.37 | -0.07 | 0.04 | -0.02 | 0.59 | -0.2 | 0 |
| **R isthmuscingulate** | -0.05 | 0.38 | 0.12 | 0.04 | 0.02 | 0.82 | -0.05 | 0.18 | -0.05 | 0.16 | -0.05 | 0.08 |
| **R lateraloccipital** | -0.05 | 0.38 | 0 | 1 | -0.06 | 0.37 | -0.09 | 0.01 | -0.16 | 0 | -0.19 | 0 |
| **R lateralorbitofrontal** | -0.05 | 0.38 | -0.05 | 0.45 | -0.01 | 0.98 | -0.14 | 0 | -0.09 | 0.02 | -0.17 | 0 |
| **R lingual** | -0.04 | 0.5 | -0.01 | 1 | -0.05 | 0.43 | -0.04 | 0.22 | -0.08 | 0.03 | -0.15 | 0 |
| **R medialorbitofrontal** | -0.04 | 0.5 | -0.05 | 0.47 | 0.06 | 0.37 | -0.09 | 0.01 | -0.06 | 0.09 | -0.19 | 0 |
| **R middletemporal** | -0.07 | 0.26 | -0.06 | 0.33 | -0.06 | 0.4 | -0.11 | 0 | -0.08 | 0.03 | -0.2 | 0 |
| **R parahippocampal** | -0.28 | 0 | 0.1 | 0.05 | -0.07 | 0.37 | -0.12 | 0 | -0.03 | 0.38 | -0.16 | 0 |
| **R paracentral** | -0.26 | 0 | 0.06 | 0.33 | -0.01 | 0.98 | -0.11 | 0 | -0.07 | 0.08 | -0.1 | 0 |
| **R parsopercularis** | -0.12 | 0.04 | 0.03 | 0.8 | -0.06 | 0.37 | -0.09 | 0.01 | -0.07 | 0.08 | -0.22 | 0 |
| **R parsorbitalis** | -0.04 | 0.5 | -0.06 | 0.33 | -0.08 | 0.29 | -0.06 | 0.08 | -0.14 | 0 | -0.15 | 0 |
| **R parstriangularis** | -0.06 | 0.35 | -0.05 | 0.47 | -0.02 | 0.87 | -0.08 | 0.03 | -0.12 | 0 | -0.13 | 0 |
| **R pericalcarine** | -0.04 | 0.46 | 0 | 1 | -0.06 | 0.4 | -0.05 | 0.15 | -0.06 | 0.12 | -0.1 | 0 |
| **R postcentral** | -0.28 | 0 | 0.02 | 0.95 | -0.02 | 0.87 | -0.1 | 0 | -0.04 | 0.29 | -0.19 | 0 |
| **R posteriorcingulate** | -0.11 | 0.06 | -0.05 | 0.45 | -0.01 | 0.92 | -0.11 | 0 | -0.16 | 0 | -0.2 | 0 |
| **R precentral** | -0.11 | 0.05 | 0.1 | 0.07 | -0.05 | 0.52 | -0.15 | 0 | -0.06 | 0.09 | -0.17 | 0 |
| **R precuneus** | -0.12 | 0.03 | 0.08 | 0.14 | 0.03 | 0.71 | -0.06 | 0.1 | -0.05 | 0.16 | -0.11 | 0 |
| **R rostralanteriorcingulate** | -0.13 | 0.02 | -0.01 | 1 | -0.03 | 0.73 | -0.11 | 0 | -0.09 | 0.02 | -0.21 | 0 |
| **R rostralmiddlefrontal** | -0.14 | 0.02 | 0 | 1 | 0 | 0.98 | -0.08 | 0.03 | -0.06 | 0.1 | -0.17 | 0 |
| **R superiorfrontal** | -0.17 | 0 | 0.01 | 1 | -0.05 | 0.45 | -0.15 | 0 | -0.16 | 0 | -0.23 | 0 |
| **R superiorparietal** | -0.14 | 0.01 | 0.1 | 0.07 | -0.05 | 0.45 | -0.11 | 0 | -0.11 | 0 | -0.16 | 0 |
| **R superiortemporal** | -0.12 | 0.05 | -0.04 | 0.65 | 0 | 0.98 | -0.08 | 0.03 | -0.2 | 0 | -0.18 | 0 |
| **R supramarginal** | -0.22 | 0 | 0.01 | 1 | -0.01 | 0.98 | -0.1 | 0 | -0.11 | 0 | -0.15 | 0 |
| **R frontalpole** | -0.04 | 0.5 | 0.02 | 0.99 | -0.03 | 0.71 | -0.02 | 0.57 | -0.05 | 0.18 | -0.08 | 0 |
| **R temporalpole** | -0.01 | 0.8 | 0 | 1 | -0.08 | 0.29 | 0.03 | 0.42 | -0.01 | 0.76 | -0.06 | 0.03 |
| **R transversetemporal** | -0.11 | 0.06 | 0.03 | 0.8 | 0.01 | 0.98 | -0.07 | 0.04 | -0.12 | 0 | -0.18 | 0 |
| **R insula** | -0.05 | 0.42 | 0 | 1 | -0.02 | 0.87 | -0.1 | 0.01 | -0.1 | 0.01 | -0.13 | 0 |

**Table S18** Group differences in subcortical volumes between individuals with a neurodevelopmental and psychiatric condition (NDPC) and reference comparators.

Cohen's *d* maps were computed between individuals with and without NDPCs. Reference comparators per NDPC were matched via propensity score matching, taking age and sex into account. As tests were based on deviation scores which were intrinsically corrected for age, sex, and site in the normative modelling step, these covariates were not included as covariates in t-tests / Cohen's *d* computation. FDR correction was performed at an alpha level <0.05 for each effect map. ANXG = generalized anxiety disorder, ASD = autism spectrum diagnosis, BP = bipolar disorder, MDD = major depressive disorder, OCD = obsessive-compulsive disorder, SCZ = Schizophrenia spectrum.

| | **ANXG** | | **ASD** | | **BP** | | **MDD** | | **OCD** | | **SCZ** | |
|---|---|---|---|---|---|---|---|---|---|---|---|---|
| **Structure** | *d* | $p_{FDR}$ | *d* | $p_{FDR}$ | *d* | $p_{FDR}$ | *d* | $p_{FDR}$ | *d* | $p_{FDR}$ | *d* | $p_{FDR}$ |
| **Laccumb** | -0.17 | 0 | -0.16 | 0 | -0.09 | 0.04 | -0.08 | 0.03 | -0.04 | 0.45 | -0.18 | 0 |
| **Lamyg** | -0.19 | 0 | -0.06 | 0.29 | -0.04 | 0.45 | -0.18 | 0 | -0.05 | 0.42 | -0.27 | 0 |
| **Lcaud** | -0.2 | 0 | 0.06 | 0.28 | -0.05 | 0.33 | -0.13 | 0 | 0.02 | 0.59 | 0.07 | 0.01 |
| **Lhippo** | -0.23 | 0 | -0.02 | 0.65 | -0.17 | 0 | -0.11 | 0 | -0.05 | 0.42 | -0.41 | 0 |
| **Lpal** | -0.06 | 0.26 | 0 | 0.91 | 0.08 | 0.1 | 0.01 | 0.87 | 0.04 | 0.45 | 0.29 | 0 |
| **Lput** | -0.16 | 0 | -0.02 | 0.65 | 0.01 | 0.77 | -0.08 | 0.01 | 0.09 | 0.05 | 0.13 | 0 |
| **Lthal** | -0.15 | 0 | 0.05 | 0.3 | -0.17 | 0 | -0.12 | 0 | 0.01 | 0.72 | -0.28 | 0 |
| **Raccumb** | -0.2 | 0 | -0.08 | 0.13 | -0.07 | 0.12 | -0.05 | 0.15 | 0.02 | 0.72 | -0.21 | 0 |
| **Ramyg** | -0.16 | 0 | -0.05 | 0.3 | -0.05 | 0.29 | -0.12 | 0 | -0.03 | 0.53 | -0.24 | 0 |
| **Rcaud** | -0.19 | 0 | 0.02 | 0.65 | -0.02 | 0.66 | -0.13 | 0 | 0.05 | 0.42 | 0.07 | 0.01 |
| **Rhippo** | -0.23 | 0 | -0.01 | 0.88 | -0.18 | 0 | -0.09 | 0.01 | 0 | 0.89 | -0.42 | 0 |
| **Rpal** | -0.09 | 0.1 | -0.05 | 0.3 | 0.02 | 0.66 | 0.02 | 0.51 | 0.03 | 0.59 | 0.21 | 0 |
| **Rput** | -0.16 | 0 | -0.06 | 0.28 | -0.01 | 0.89 | -0.04 | 0.26 | 0.11 | 0.02 | 0.13 | 0 |
| **Rthal** | -0.06 | 0.23 | 0.05 | 0.32 | -0.13 | 0 | -0.13 | 0 | -0.06 | 0.3 | -0.26 | 0 |

**Table S19** *Demographic information for reference cohort subgroups (RC) matched to individuals with a neurodevelopmental or psychiatric condition (NDPC) via propensity score matching for Cohen's d map computation.*

The NDPC column specifies the diagnostic group to which the respective RC subgroup was matched. ANXG = generalized anxiety disorder, ASD = autism spectrum diagnosis, BP = bipolar disorder, MDD = major depressive disorder, OCD = obsessive-compulsive disorder, SCZ = Schizophrenia spectrum.

| NDPC | $n_{RC}$ | Mean age | Age (STD) | Min age | Max age | # Male | # Female | # Sites |
|---|---|---|---|---|---|---|---|---|
| **ANXG** | 765 | 26.33 | 12.05 | 8 | 73 | 459 | 306 | 138 |
| **ASD** | 1556 | 21.84 | 8.69 | 5 | 64 | 995 | 561 | 167 |
| **BP** | 1370 | 38.49 | 13.98 | 13 | 78 | 776 | 594 | 135 |
| **MDD** | 1916 | 34.11 | 14.93 | 8 | 78 | 1141 | 775 | 159 |
| **OCD** | 1775 | 27.89 | 11.63 | 5 | 64 | 942 | 833 | 165 |
| **SCZ** | 2397 | 30.41 | 15.36 | 5 | 78 | 1266 | 1147 | 168 |

**Table S20** *Similarities between Cohen's d maps from current normative models and previously published ENIGMA maps.*

*Rho indicates Spearman correlations between current and previously reported effect size maps. Note that samples partially overlap; this is a consistency and sensitivity check for our normative model outputs rather than an independent replication. Significance of spatial associations was corrected for spatial autocorrelations using spatial variogram permutations (10,000 permutations).* ASD = autism spectrum diagnosis, BP = bipolar disorder, MDD = major depressive disorder, OCD = obsessive-compulsive disorder, SCZ = Schizophrenia spectrum, CT = Cortical thickness, SA = surface area.

| Disorder | Measure | Rho | $p_{variogram}$ |
|---|---|---|---|
| **ASD**[18] | CT | 0.77 | 0.0001* |
| **BP**[19] | CT | 0.74 | 0.0001* |
| **MDD**[20] | CT | 0.18 | 0.37 |
| **OCD**[21] | CT | 0.66 | 0.0001* |
| **SCZ**[22] | CT | 0.80 | 0.0001* |
| **BP** | SA | 0.28 | 0.013* |
| **MDD** | SA | 0.29 | 0.017* |
| **OCD** | SA | 0.72 | 0.0001* |
| **SCZ** | SA | 0.76 | 0.0001* |

* = significant at FDR p<0.05.

**Table S21** *Within-diagnosis variability in cortical thickness.*

*Variability was* measured by standard deviation (Sd)*. Differences in variability between individuals with and without a neurodevelopmental or psychiatric condition were estimated via Levene's test. The control groups were matched via propensity score matching, taking age and sex into account. FDR correction was performed at an alpha level <0.05.* ANXG = generalized anxiety disorder, ASD = autism spectrum diagnosis, BP = bipolar disorder, MDD = major depressive disorder, OCD = obsessive-compulsive disorder, SCZ = Schizophrenia spectrum.

| | ANXG | | ASD | | BP | | MDD | | OCD | | SCZ | |
|---|---|---|---|---|---|---|---|---|---|---|---|---|
| | **Sd** | **Levene (pFDR)** | **Sd** | **Levene (pFDR)** | **Sd** | **Levene (pFDR)** | **Sd** | **Levene (pFDR)** | **Sd** | **Levene (pFDR)** | **Sd** | **Levene (pFDR)** |
| L bankssts | 0.93 | 17.99 (<0.0001) | 1.24 | 23.04 (<0.0001) | 0.97 | 9.58 (0.02) | 0.99 | 7.17 (0.08) | 1.05 | 0.01 (0.91) | 1.05 | 0.5 (0.54) |
| L caudal anterior cingulate | 0.92 | 3.13 (0.1) | 1.15 | 26.31 (<0.0001) | 1.02 | 0.02 (0.91) | 1.03 | 1.99 (0.67) | 1.2 | 49.28 (<0.0001) | 1.05 | 8.55 (0.01) |
| L caudal middle frontal | 0.93 | 6.33 (0.02) | 1.22 | 45.37 (<0.0001) | 1.04 | 1.31 (0.44) | 1 | 0.18 (0.97) | 1.13 | 29.25 (<0.0001) | 1.05 | 8.67 (0.01) |
| L cuneus | 0.96 | 5.48 (0.03) | 1.23 | 40.38 (<0.0001) | 1.03 | 0 (0.95) | 1.02 | 0.31 (0.95) | 1.02 | 0.06 (0.82) | 1.02 | 0.01 (0.95) |
| L entorhinal | 0.86 | 14.07 (<0.0001) | 1.21 | 55.42 (<0.0001) | 0.99 | 0.02 (0.91) | 0.99 | 0.01 (0.97) | 1.13 | 34.08 (<0.0001) | 1.03 | 5.97 (0.04) |
| L fusiform | 0.89 | 26.1 (<0.0001) | 1.26 | 44.21 (<0.0001) | 0.99 | 2.1 (0.3) | 1.03 | 0.11 (0.97) | 1.1 | 4.93 (0.04) | 1.06 | 1.74 (0.24) |
| L inferior parietal | 0.93 | 11.36 (<0.0001) | 1.26 | 53.06 (<0.0001) | 0.97 | 0.76 (0.55) | 1.02 | 0.59 (0.89) | 1.09 | 6.71 (0.02) | 1.04 | 1.72 (0.24) |
| L inferior temporal | 0.86 | 35.17 (<0.0001) | 1.24 | 42.15 (<0.0001) | 0.98 | 2.39 (0.27) | 0.97 | 4.03 (0.3) | 1.07 | 2.63 (0.12) | 1.06 | 3.41 (0.11) |
| L isthmus cingulate | 0.94 | 7.4 (0.01) | 1.18 | 38.49 (<0.0001) | 1.03 | 0.14 (0.77) | 1.08 | 9.78 (0.08) | 1.16 | 19.69 (<0.0001) | 1.06 | 8.5 (0.01) |
| L lateral occipital | 0.95 | 9.91 (<0.0001) | 1.22 | 35.25 (<0.0001) | 1.03 | 0.04 (0.88) | 1.05 | 0.69 (0.89) | 1.11 | 4.7 (0.04) | 1.1 | 9.17 (0.01) |
| L lateralorbitofrontal | 0.91 | 12.43 (<0.0001) | 1.24 | 36.97 (<0.0001) | 0.98 | 3.06 (0.2) | 0.99 | 0.29 (0.95) | 1.15 | 20.73 (<0.0001) | 1.06 | 1.82 (0.24) |
| L lingual | 0.92 | 11.61 (<0.0001) | 1.23 | 43.16 (<0.0001) | 1.01 | 0.22 (0.74) | 1 | 0.48 (0.89) | 1.08 | 6.54 (0.02) | 1.06 | 3.59 (0.1) |
| L medial orbitofrontal | 0.9 | 14.92 (<0.0001) | 1.25 | 59.54 (<0.0001) | 1 | 0.98 (0.51) | 0.95 | 2.74 (0.52) | 1.17 | 36.12 (<0.0001) | 1.04 | 6.08 (0.03) |
| L middle temporal | 0.93 | 7.05 (0.01) | 1.25 | 53.02 (<0.0001) | 0.95 | 2.12 (0.3) | 0.98 | 0.48 (0.89) | 1.06 | 5.03 (0.03) | 1.05 | 9.07 (0.01) |
| L para-hippocampal | 0.98 | 0.35 (0.57) | 1.11 | 19.68 (<0.0001) | 1.01 | 0.36 (0.68) | 1.03 | 0.84 (0.87) | 1.09 | 22.11 (<0.0001) | 1.02 | 2.35 (0.18) |
| L paracentral | 0.98 | 2 (0.18) | 1.16 | 25.63 (<0.0001) | 1.11 | 9.62 (0.02) | 1.01 | 0.45 (0.89) | 1.11 | 20.09 (<0.0001) | 1.06 | 7.67 (0.02) |
| L pars opercularis | 0.99 | 7.53 (0.01) | 1.21 | 31.21 (<0.0001) | 0.97 | 6.39 (0.08) | 1.02 | 0.21 (0.97) | 1.09 | 5.38 (0.03) | 1.1 | 18.08 (<0.0001) |
| L pars orbitalis | 0.9 | 19.64 (<0.0001) | 1.2 | 22.71 (<0.0001) | 0.94 | 7.41 (0.06) | 1 | 0.9 (0.87) | 1.09 | 2.91 (0.1) | 1.06 | 0.36 (0.6) |
| L pars triangularis | 0.94 | 10.91 (<0.0001) | 1.23 | 41.99 (<0.0001) | 0.98 | 4.67 (0.11) | 1.03 | 0.05 (0.97) | 1.1 | 4.6 (0.04) | 1.08 | 3.43 (0.11) |
| L pericalcarine | 0.99 | 0.47 (0.52) | 1.23 | 37.5 (<0.0001) | 1.02 | 0.4 (0.68) | 0.96 | 7.24 (0.08) | 1.02 | 1.23 (0.29) | 1.08 | 4.85 (0.05) |
| L postcentral | 1.11 | 1.8 (0.2) | 1.12 | 13.49 (<0.0001) | 1.03 | 0.19 (0.74) | 1.01 | 0.07 (0.97) | 1.06 | 3.5 (0.07) | 1.1 | 21.59 (<0.0001) |
| L posterior cingulate | 0.98 | 5.32 (0.03) | 1.21 | 28.98 (<0.0001) | 1.04 | 0.07 (0.84) | 1.06 | 0.99 (0.87) | 1.14 | 13.65 (<0.0001) | 1.07 | 4.43 (0.06) |
| L precentral | 1.03 | 0.5 (0.51) | 1.15 | 23.42 (<0.0001) | 1.06 | 0.29 (0.71) | 1.01 | 0.02 (0.97) | 1.06 | 4.75 (0.04) | 1.05 | 5.28 (0.05) |
| L precuneus | 0.95 | 9.17 (0.01) | 1.25 | 36.45 (<0.0001) | 1.04 | 0.48 (0.66) | 1.06 | 0.02 (0.97) | 1.14 | 9.66 (<0.0001) | 1.06 | 0.24 (0.66) |
| L rostral anterior cingulate | 0.94 | 6.05 (0.02) | 1.15 | 34.13 (<0.0001) | 0.94 | 3.08 (0.2) | 1.01 | 0.95 (0.87) | 1.14 | 41.41 (<0.0001) | 1.06 | 17.71 (<0.0001) |
| L rostral middle frontal | 0.92 | 12.08 (<0.0001) | 1.29 | 60.29 (<0.0001) | 0.93 | 14.18 (<0.0001) | 1.02 | 0.93 (0.87) | 1.1 | 9.56 (<0.0001) | 1.03 | 2.66 (0.16) |
| L superior frontal | 0.95 | 4.83 (0.04) | 1.26 | 53.72 (<0.0001) | 1.01 | 0.14 (0.77) | 1 | 0.04 (0.97) | 1.11 | 12.89 (<0.0001) | 1.06 | 9.51 (0.01) |

| | | | | | | | | | | | | |
|---|---|---|---|---|---|---|---|---|---|---|---|---|
| | | | 1.2 | 27.3 | 1.0 | | 1.0 | | | | | |
| L superior parietal | 1 | 3.89 (0.06) | 1 | (<0.0001) | 2 | 1.12 (0.48) | 2 | 0.28 (0.95) | 1.13 | 8.64 (0.01) | 1.07 | 2.01 (0.22) |
| | 0.9 | | | 65.06 | 0.9 | | 1.0 | | | | | 19.92 |
| L superior temporal | 8 | 2.08 (0.17) | 1.3 | (<0.0001) | 6 | 5.2 (0.1) | 2 | 1.29 (0.87) | 1.07 | 7 (0.01) | 1.09 | (<0.0001) |
| | 0.9 | 12.6 | 1.2 | 31.98 | 0.9 | | 1.0 | | | | | |
| L supramarginal | 2 | (<0.0001) | 1 | (<0.0001) | 6 | 2.96 (0.2) | 1 | 0.58 (0.89) | 1.07 | 6.99 (0.01) | 1.06 | 5.26 (0.05) |
| | 0.8 | 24.6 | 1.1 | 24.67 | 0.9 | 17.89 | | | | 12.48 | | |
| L frontal pole | 7 | (<0.0001) | 6 | (<0.0001) | 3 | (<0.0001) | 1 | 0.02 (0.97) | 1.09 | (<0.0001) | 0.97 | 5.02 (0.05) |
| | 0.8 | 15.58 | | 46.16 | 0.9 | | 0.9 | | | 14.03 | | |
| L temporal pole | 9 | (<0.0001) | 1.2 | (<0.0001) | 5 | 5.44 (0.09) | 9 | 0 (0.97) | 1.09 | (<0.0001) | 0.98 | 0.58 (0.51) |
| L transverse | | | | 35.59 | 1.0 | | 1.0 | | | 9.16 | | |
| temporal | 1 | 0.81 (0.4) | 1.2 | (<0.0001) | 3 | 3.42 (0.18) | 3 | 6.31 (0.12) | 1.1 | (<0.0001) | 1.05 | 8.28 (0.01) |
| | 0.9 | | 1.2 | 21.95 | 0.9 | | 0.9 | | | | | |
| L insula | 9 | 2.44 (0.14) | 2 | (<0.0001) | 6 | 5 (0.1) | 7 | 3.81 (0.31) | 1.11 | 6.98 (0.01) | 1.08 | 5.14 (0.05) |
| | 0.9 | | 1.2 | 35.1 | 0.9 | | | | | | | |
| R bankssts | 8 | 8.85 (0.01) | 4 | (<0.0001) | 7 | 0.36 (0.68) | 1 | 0.05 (0.97) | 1.07 | 3.9 (0.06) | 1.05 | 5.76 (0.04) |
| R caudal anterior | 0.9 | | 1.1 | 30.05 | 1.0 | | | | | 21.14 | | 10.76 |
| cingulate | 4 | 4.7 (0.04) | 7 | (<0.0001) | 2 | 0.48 (0.66) | 1 | 0.5 (0.89) | 1.14 | (<0.0001) | 1.04 | (0.01) |
| R caudal middle | 0.9 | 11.06 | 1.2 | 36.54 | 1.0 | | | | | 16.11 | | |
| frontal | 2 | (<0.0001) | 3 | (<0.0001) | 1 | 0.29 (0.71) | 1 | 0.09 (0.97) | 1.11 | (<0.0001) | 1.01 | 0.01 (0.94) |
| | | 9.74 | 1.2 | 46.94 | 1.0 | | 1.0 | | | | | |
| R cuneus | 0.9 | (<0.0001) | 2 | (<0.0001) | 2 | 0.94 (0.51) | 6 | 8.6 (0.08) | 1.04 | 3.56 (0.07) | 1.06 | 9.83 (0.01) |
| | | 11.72 | 1.2 | 74.9 | 0.9 | | | | | 10.52 | | 12.12 |
| R entorhinal | 0.9 | (<0.0001) | 5 | (<0.0001) | 8 | 0.57 (0.64) | 1 | 0.09 (0.97) | 1.09 | (<0.0001) | 1.06 | (<0.0001) |
| | 0.8 | 22.76 | 1.2 | 56.89 | | | 1.0 | | | | | 10.95 |
| R fusiform | 7 | (<0.0001) | 8 | (<0.0001) | 1 | 0.77 (0.55) | 1 | 0.06 (0.97) | 1.05 | 0.55 (0.49) | 1.08 | (0.01) |
| | 0.8 | 14.9 | 1.2 | 54.91 | 0.9 | | 1.0 | | | | | |
| R inferior parietal | 8 | (<0.0001) | 6 | (<0.0001) | 5 | 4.46 (0.12) | 1 | 0 (0.97) | 1.07 | 5.8 (0.02) | 1.01 | 1.12 (0.35) |
| | 0.8 | 16.55 | 1.2 | 64.4 | 0.9 | | 0.9 | | | 10.42 | | |
| R inferior temporal | 7 | (<0.0001) | 9 | (<0.0001) | 6 | 0.42 (0.68) | 5 | 2.7 (0.52) | 1.08 | (<0.0001) | 1.03 | 3.11 (0.13) |
| | 0.9 | | 1.1 | 15.99 | 1.0 | | 1.0 | | | 11.79 | | |
| R isthmus cingulate | 7 | 2.12 (0.17) | 4 | (<0.0001) | 4 | 1.3 (0.44) | 6 | 7.22 (0.08) | 1.12 | (<0.0001) | 1.06 | 9.97 (0.01) |
| | 0.9 | | 1.2 | 56.08 | 1.0 | | 1.0 | | | | | |
| R lateral occipital | 3 | 6.08 (0.02) | 3 | (<0.0001) | 5 | 1.48 (0.43) | 3 | 1.54 (0.86) | 1.1 | 6.46 (0.02) | 1.06 | 9.23 (0.01) |
| R | 0.8 | 24.23 | 1.2 | 60.08 | 0.9 | | 0.9 | | | 25 | | |
| lateralorbitofrontal | 8 | (<0.0001) | 7 | (<0.0001) | 4 | 3.85 (0.14) | 8 | 0 (0.97) | 1.13 | (<0.0001) | 1.05 | 5.51 (0.04) |
| | 0.9 | 10.3 | 1.2 | 41.75 | 0.9 | | 0.9 | | | | | |
| R lingual | 2 | (<0.0001) | 3 | (<0.0001) | 9 | 2.53 (0.25) | 9 | 2.06 (0.67) | 1.03 | 0.09 (0.78) | 1.05 | 2.95 (0.14) |
| R medial | 0.8 | 16.69 | 1.2 | 53.08 | 0.9 | | 0.9 | | | 48.43 | | |
| orbitofrontal | 8 | (<0.0001) | 4 | (<0.0001) | 9 | 1.16 (0.48) | 9 | 0.06 (0.97) | 1.19 | (<0.0001) | 1.06 | 4.68 (0.06) |
| | 0.8 | 24.25 | 1.3 | 67.43 | 0.9 | | 0.9 | | | | | |
| R middle temporal | 8 | (<0.0001) | 2 | (<0.0001) | 5 | 4.03 (0.14) | 8 | 0.93 (0.87) | 1.07 | 3.4 (0.08) | 1.05 | 1.67 (0.25) |
| | 0.9 | | 1.1 | 26.59 | 0.9 | | 1.0 | | | 12.87 | | |
| R para-hippocampal | 4 | 3.05 (0.1) | 6 | (<0.0001) | 7 | 5.82 (0.09) | 3 | 0.59 (0.89) | 1.1 | (<0.0001) | 1.05 | 5.03 (0.05) |
| | 0.9 | | 1.1 | 24.64 | 1.0 | | 1.0 | | | | | |
| R paracentral | 7 | 8.92 (0.01) | 9 | (<0.0001) | 9 | 1.01 (0.51) | 3 | 1.01 (0.87) | 1.12 | 7.35 (0.01) | 1.06 | 2.56 (0.16) |
| | 0.9 | | | 26.23 | 0.9 | | 1.0 | | | 13.41 | | |
| R pars opercularis | 6 | 5.52 (0.03) | 1.2 | (<0.0001) | 9 | 1.4 (0.44) | 2 | 0.12 (0.97) | 1.09 | (<0.0001) | 1.07 | 6.06 (0.03) |
| | 0.9 | | 1.2 | 40.12 | 0.9 | | 1.0 | | | 11.57 | | |
| R pars orbitalis | 3 | 5.01 (0.04) | 2 | (<0.0001) | 1 | 12.31 (0.01) | 3 | 1.11 (0.87) | 1.1 | (<0.0001) | 1.01 | 1.03 (0.37) |
| | 0.9 | | 1.2 | 50.28 | 0.9 | | 1.0 | | | 10.06 | | |
| R pars triangularis | 5 | 7.63 (0.01) | 7 | (<0.0001) | 6 | 5.43 (0.09) | 3 | 0.67 (0.89) | 1.11 | (<0.0001) | 1.06 | 2.82 (0.14) |
| | 1.0 | | 1.1 | 24.48 | 1.0 | | 0.9 | | | | | 12.51 |
| R pericalcarine | 2 | 0.08 (0.78) | 7 | (<0.0001) | 1 | 0.46 (0.66) | 6 | 5.01 (0.21) | 0.98 | 3.6 (0.07) | 1.09 | (<0.0001) |
| | | | 1.1 | 26.87 | 1.0 | | 1.0 | | | | | 19.28 |
| R postcentral | 1.1 | 2.16 (0.17) | 6 | (<0.0001) | 1 | 0.19 (0.74) | 1 | 0.05 (0.97) | 1.03 | 0.82 (0.39) | 1.08 | (<0.0001) |
| | 0.9 | | | 30.58 | 1.0 | | 1.0 | | | 17.86 | | |
| R posteriorcingulate | 4 | 7.19 (0.01) | 1.2 | (<0.0001) | 3 | 0.81 (0.55) | 2 | 0.93 (0.87) | 1.12 | (<0.0001) | 1.07 | 9.24 (0.01) |
| | 1.0 | | 1.1 | 14.73 | 1.0 | | 1.0 | | | | | |
| R precentral | 5 | 2.82 (0.12) | 4 | (<0.0001) | 2 | 1.64 (0.4) | 2 | 0.44 (0.89) | 1.07 | 1.34 (0.27) | 1.02 | 0.01 (0.95) |
| | 0.9 | 9.41 | 1.2 | 41.37 | 1.0 | | 1.0 | | | 14.95 | | |
| R precuneus | 5 | (<0.0001) | 4 | (<0.0001) | 1 | 1.39 (0.44) | 3 | 0.48 (0.89) | 1.13 | (<0.0001) | 1.05 | 1.38 (0.3) |
| R rostral anterior | 0.8 | | 1.1 | 26.12 | 0.9 | | 0.9 | | | 26.81 | | 11.99 |
| cingulate | 9 | 6.38 (0.02) | 3 | (<0.0001) | 4 | 3.83 (0.14) | 9 | 0.01 (0.97) | 1.13 | (<0.0001) | 1.05 | (<0.0001) |
| R rostral middle | 0.8 | 29.67 | 1.3 | 86.21 | 0.9 | | 1.0 | | | 11.18 | | |
| frontal | 6 | (<0.0001) | 8 | (<0.0001) | 6 | 5.37 (0.09) | 1 | 0.11 (0.97) | 1.09 | (<0.0001) | 1.03 | 0.68 (0.47) |

| | | | | | | | | | | | | |
|---|---|---|---|---|---|---|---|---|---|---|---|---|
| R superior frontal | 0.94 | 5.79 (0.03) | 1.32 | 64.92 (<0.0001) | 1.01 | 0.19 (0.74) | 1.02 | 0.09 (0.97) | 1.11 | 14.15 (<0.0001) | 1.07 | 6.11 (0.03) |
| R superior parietal | 0.98 | 4.6 (0.04) | 1.2 | 30.36 (<0.0001) | 1.01 | 3.33 (0.18) | 1.04 | 0.06 (0.97) | 1.12 | 5.16 (0.03) | 1.07 | 2.44 (0.17) |
| R superior temporal | 0.94 | 9.72 (<0.0001) | 1.31 | 67.89 (<0.0001) | 0.97 | 6.02 (0.09) | 1.02 | 0 (0.99) | 1.08 | 6.32 (0.02) | 1.09 | 7.48 (0.02) |
| R supramarginal | 0.92 | 9.72 (<0.0001) | 1.23 | 27.91 (<0.0001) | 0.95 | 8.75 (0.03) | 0.99 | 1.31 (0.87) | 1.09 | 6.38 (0.02) | 1.04 | 0 (0.98) |
| R frontal pole | 0.87 | 25.21 (<0.0001) | 1.12 | 9.34 (<0.0001) | 0.93 | 7.15 (0.06) | 0.99 | 0.28 (0.95) | 1.09 | 11.62 (<0.0001) | 0.97 | 6.65 (0.03) |
| R temporal pole | 0.91 | 10.8 (<0.0001) | 1.21 | 32.81 (<0.0001) | 0.9 | 16.22 (<0.0001) | 0.92 | 7.7 (0.08) | 1.15 | 18.43 (<0.0001) | 1 | 0.29 (0.64) |
| R transverse temporal | 1.04 | 0.21 (0.65) | 1.21 | 27.78 (<0.0001) | 1.06 | 4.12 (0.14) | 1.05 | 2.07 (0.67) | 1.04 | 0.16 (0.72) | 1.05 | 1.74 (0.24) |
| R insula | 0.98 | 1.13 (0.32) | 1.2 | 34.59 (<0.0001) | 0.95 | 4.78 (0.11) | 0.95 | 4.18 (0.3) | 1.09 | 9.66 (<0.0001) | 1.02 | 0.67 (0.47) |

**Table *S*22** Within-diagnosis variability in surface area.

Variability was measured by standard deviation (Sd). Differences in variability between individuals with and without a neurodevelopmental or psychiatric condition were estimated via Levene's test. The control groups were matched via propensity score matching, taking age and sex into account. FDR correction was performed at an alpha level <0.05. ANXG = generalized anxiety disorder, ASD = autism spectrum diagnosis, BP = bipolar disorder, MDD = major depressive disorder, OCD = obsessive-compulsive disorder, SCZ = Schizophrenia spectrum.

| | ANXG | | ASD | | BP | | MDD | | OCD | | SCZ | |
|---|---|---|---|---|---|---|---|---|---|---|---|---|
| **Structure** | **Sd** | **Levene (pFDR)** | **Sd** | **Levene (pFDR)** | **Sd** | **Levene (pFDR)** | **Sd** | **Levene (pFDR)** | **Sd** | **Levene (pFDR)** | **Sd** | **Levene (pFDR)** |
| L bankssts | 1.03 | 1.03 (0.39) | 1.12 | 11.32 (<0.0001) | 0.93 | 2.75 (0.45) | 0.96 | 0.1 (0.89) | 0.96 | 1.53 (0.35) | 1.05 | 10.33 (0.01) |
| L caudal anterior cingulate | 1.1 | 7.51 (0.01) | 1.11 | 10.74 (<0.0001) | 1.03 | 0.91 (0.66) | 1.04 | 1.66 (0.56) | 1 | 0.15 (0.8) | 1.03 | 1.06 (0.4) |
| L caudal middle frontal | 0.96 | 1.02 (0.39) | 1.12 | 13.88 (<0.0001) | 1 | 0.37 (0.82) | 0.97 | 0.52 (0.65) | 0.98 | 0 (0.98) | 1.04 | 7.13 (0.02) |
| L cuneus | 1.19 | 9.98 (<0.0001) | 1.12 | 4.2 (0.05) | 1.04 | 0.21 (0.86) | 0.99 | 1.27 (0.57) | 0.97 | 7.31 (0.03) | 1.05 | 0.32 (0.64) |
| L entorhinal | 1.2 | 11.11 (<0.0001) | 1.08 | 1.49 (0.24) | 0.99 | 1.29 (0.61) | 0.97 | 1.14 (0.57) | 0.93 | 8.14 (0.02) | 1.07 | 10.54 (0.01) |
| L fusiform | 1.06 | 0.01 (0.94) | 1.16 | 10.75 (<0.0001) | 0.98 | 13.11 (0.02) | 1.01 | 1.59 (0.56) | 1.04 | 0.86 (0.48) | 1.07 | 0.72 (0.49) |
| L inferior parietal | 1.09 | 4.33 (0.06) | 1.15 | 18.43 (<0.0001) | 1.04 | 0.01 (0.99) | 0.99 | 3.01 (0.43) | 0.98 | 1.7 (0.34) | 1.06 | 1.33 (0.34) |
| L inferior temporal | 1.11 | 3.5 (0.09) | 1.14 | 7.54 (0.01) | 0.99 | 1.77 (0.61) | 0.98 | 0.67 (0.63) | 0.98 | 2.15 (0.27) | 1.09 | 9.77 (0.01) |
| L isthmus cingulate | 1.14 | 12.12 (<0.0001) | 1.14 | 12.97 (<0.0001) | 1 | 1.21 (0.61) | 1.02 | 0.02 (0.97) | 1.06 | 3.53 (0.14) | 1.02 | 0 (0.98) |
| L lateral occipital | 1.24 | 18.63 (<0.0001) | 1.05 | 0.17 (0.7) | 0.99 | 1.48 (0.61) | 1 | 1 (0.57) | 0.93 | 14.26 (<0.0001) | 1.07 | 4.75 (0.06) |
| L lateralorbitofrontal | 1.15 | 12.41 (<0.0001) | 1.22 | 50.31 (<0.0001) | 1.01 | 0.69 (0.69) | 0.96 | 5.11 (0.27) | 0.98 | 0.55 (0.59) | 1.09 | 9.99 (0.01) |
| L lingual | 1.17 | 16.89 (<0.0001) | 1.13 | 5.7 (0.02) | 1.01 | 0.25 (0.85) | 1 | 0.2 (0.85) | 1 | 2.62 (0.22) | 1.04 | 0.57 (0.54) |
| L medial orbitofrontal | 1.14 | 4.62 (0.05) | 1.14 | 10.49 (<0.0001) | 0.99 | 2.08 (0.58) | 0.98 | 1.04 (0.57) | 1.01 | 0.02 (0.93) | 1.09 | 8.44 (0.01) |
| L middle temporal | 1.14 | 9.39 (<0.0001) | 1.17 | 24.76 (<0.0001) | 1 | 0.09 (0.92) | 0.97 | 1.4 (0.57) | 0.93 | 8.6 (0.02) | 1.1 | 16.66 (<0.0001) |
| L para-hippocampal | 0.99 | 0.57 (0.54) | 1.16 | 21.92 (<0.0001) | 0.97 | 1.45 (0.61) | 1.01 | 1.66 (0.56) | 1.07 | 6.29 (0.05) | 1.02 | 1.68 (0.28) |
| L paracentral | 1.01 | 0.19 (0.7) | 1.17 | 15.05 (<0.0001) | 1.03 | 0.3 (0.85) | 1 | 1.45 (0.57) | 1.02 | 0.15 (0.8) | 1.03 | 0.34 (0.64) |
| L pars opercularis | 1.03 | 1.34 (0.32) | 1.14 | 15.44 (<0.0001) | 0.93 | 6.19 (0.18) | 0.97 | 0.66 (0.63) | 0.99 | 0.2 (0.79) | 1.01 | 0.07 (0.83) |
| L pars orbitalis | 1.19 | 25.03 (<0.0001) | 1.08 | 2.3 (0.15) | 1.04 | 3.7 (0.34) | 0.97 | 1.85 (0.56) | 0.94 | 7.03 (0.04) | 1.06 | 8.04 (0.01) |
| L pars triangularis | 1.15 | 16.17 (<0.0001) | 1.06 | 1.27 (0.27) | 0.97 | 1.26 (0.61) | 0.96 | 3.04 (0.43) | 0.98 | 0.95 (0.47) | 1.02 | 0.69 (0.49) |
| L pericalcarine | 1.15 | 7.9 (0.01) | 1.13 | 5.21 (0.03) | 1.01 | 0.42 (0.82) | 0.99 | 0.14 (0.88) | 0.95 | 5.04 (0.08) | 1.05 | 3.9 (0.09) |
| L postcentral | 1.23 | 14.09 (<0.0001) | 1.15 | 8.02 (0.01) | 1.04 | 0.12 (0.91) | 1 | 1.01 (0.57) | 1.04 | 0.01 (0.94) | 1.08 | 3.19 (0.13) |
| L posterior cingulate | 1.11 | 2.6 (0.16) | 1.1 | 6.7 (0.01) | 0.98 | 11.5 (0.02) | 1.03 | 0.17 (0.88) | 1.01 | 1.29 (0.39) | 1.03 | 0.3 (0.64) |
| L precentral | 1.15 | 15.88 (<0.0001) | 1.13 | 18.26 (<0.0001) | 1.04 | 0.03 (0.98) | 0.99 | 1.1 (0.57) | 0.98 | 1.53 (0.35) | 1.09 | 11.81 (<0.0001) |
| L precuneus | 1.23 | 25.58 (<0.0001) | 1.13 | 10.96 (<0.0001) | 1.01 | 0.74 (0.68) | 0.98 | 2 (0.56) | 0.97 | 6.04 (0.05) | 1.06 | 5.24 (0.05) |
| L rostral anterior cingulate | 1.11 | 6.16 (0.02) | 1.12 | 15.25 (<0.0001) | 1.02 | 0 (1) | 0.97 | 2.73 (0.43) | 0.96 | 2.27 (0.26) | 1.08 | 8.81 (0.01) |
| L rostral middle frontal | 1.2 | 22.72 (<0.0001) | 1.13 | 12.26 (<0.0001) | 1.03 | 0.24 (0.85) | 0.99 | 0.73 (0.63) | 0.93 | 14.4 (<0.0001) | 1.12 | 20.13 (<0.0001) |

| | | | | | | | | | | | | |
|---|---|---|---|---|---|---|---|---|---|---|---|---|
| L superior frontal | 1.13 | 16.88 (<0.0001) | 1.14 | 24.15 (<0.0001) | 1.05 | 2.96 (0.45) | 1 | 0.01 (0.97) | 0.94 | 2.41 (0.25) | 1.11 | 37.4 (<0.0001) |
| L superior parietal | 1.18 | 16.88 (<0.0001) | 1.11 | 9.46 (<0.0001) | 1.01 | 0.61 (0.72) | 0.97 | 7.46 (0.22) | 0.93 | 9.89 (0.01) | 1.06 | 2.92 (0.14) |
| L superior temporal | 1.15 | 10.35 (<0.0001) | 1.11 | 18.45 (<0.0001) | 0.98 | 4.1 (0.32) | 0.97 | 3.97 (0.35) | 0.93 | 8.98 (0.02) | 1.06 | 5.84 (0.04) |
| L supramarginal | 1.24 | 37.98 (<0.0001) | 1.05 | 1.29 (0.27) | 0.99 | 1.75 (0.61) | 0.96 | 5.78 (0.27) | 0.88 | 27.36 (<0.0001) | 1.08 | 7.38 (0.02) |
| L frontal pole | 1.01 | 0.08 (0.79) | 1.04 | 0 (0.99) | 0.99 | 0.83 (0.68) | 0.98 | 0.72 (0.63) | 0.97 | 2.8 (0.21) | 0.98 | 1.9 (0.25) |
| L temporal pole | 0.99 | 0.42 (0.59) | 1.09 | 8.74 (<0.0001) | 1 | 1.21 (0.61) | 1 | 0 (0.98) | 1 | 0.85 (0.48) | 1.01 | 0.94 (0.44) |
| L transverse temporal | 1.04 | 0.39 (0.59) | 1.1 | 3.36 (0.08) | 0.99 | 0.4 (0.82) | 1 | 0.04 (0.93) | 1.01 | 0.01 (0.94) | 1.03 | 1.43 (0.33) |
| L insula | 1.13 | 13.33 (<0.0001) | 1.15 | 17.92 (<0.0001) | 0.99 | 2.05 (0.58) | 0.97 | 4.43 (0.34) | 0.93 | 9.2 (0.02) | 1.06 | 4.49 (0.07) |
| R bankssts | 1.02 | 0.39 (0.59) | 1.18 | 21.42 (<0.0001) | 0.94 | 6.4 (0.18) | 0.97 | 2.59 (0.43) | 0.98 | 1.35 (0.39) | 1.07 | 6.06 (0.04) |
| R caudal anterior cingulate | 1.02 | 0.35 (0.61) | 1.08 | 11.49 (<0.0001) | 1 | 0.14 (0.91) | 1.03 | 0.77 (0.63) | 0.97 | 0.19 (0.79) | 1.02 | 1.7 (0.28) |
| R caudal middle frontal | 1.01 | 0.3 (0.62) | 1.14 | 35.58 (<0.0001) | 1 | 0.06 (0.95) | 0.99 | 0.12 (0.88) | 0.98 | 0.14 (0.8) | 1.06 | 13.35 (<0.0001) |
| R cuneus | 1.25 | 20.8 (<0.0001) | 1.08 | 3.75 (0.06) | 1.04 | 0.29 (0.85) | 0.94 | 10.82 (0.07) | 0.96 | 12.88 (<0.0001) | 1.05 | 2.64 (0.17) |
| R entorhinal | 1.05 | 2.21 (0.19) | 1.12 | 13.31 (<0.0001) | 1 | 0.02 (0.98) | 0.99 | 0.52 (0.65) | 0.99 | 1.22 (0.4) | 1.01 | 0.09 (0.83) |
| R fusiform | 1.09 | 3.98 (0.07) | 1.15 | 15.07 (<0.0001) | 0.99 | 1.13 (0.61) | 1.02 | 0 (0.97) | 1 | 0.06 (0.88) | 1.07 | 5.18 (0.05) |
| R inferior parietal | 1.16 | 9.4 (<0.0001) | 1.14 | 10.75 (<0.0001) | 0.97 | 0.75 (0.68) | 0.99 | 0 (0.98) | 0.95 | 5.71 (0.06) | 1.06 | 3.3 (0.13) |
| R inferior temporal | 1.13 | 8.03 (0.01) | 1.17 | 28.8 (<0.0001) | 0.98 | 2.41 (0.51) | 0.97 | 3.69 (0.37) | 0.99 | 0.07 (0.86) | 1.07 | 5.97 (0.04) |
| R isthmus cingulate | 1.09 | 1.69 (0.27) | 1.12 | 11.88 (<0.0001) | 0.97 | 1.39 (0.61) | 1 | 0.75 (0.63) | 1.05 | 4.34 (0.11) | 1.02 | 0.33 (0.64) |
| R lateral occipital | 1.26 | 33.71 (<0.0001) | 1.08 | 6.93 (0.01) | 0.99 | 0 (1) | 0.99 | 0.13 (0.88) | 0.92 | 9.09 (0.02) | 1.08 | 18.39 (<0.0001) |
| R lateralorbitofrontal | 1.12 | 7.54 (0.01) | 1.21 | 46.71 (<0.0001) | 0.99 | 0.26 (0.85) | 1 | 0.68 (0.63) | 0.95 | 4.16 (0.11) | 1.1 | 21.28 (<0.0001) |
| R lingual | 1.21 | 16.59 (<0.0001) | 1.06 | 0.12 (0.74) | 1.04 | 1.07 (0.62) | 0.97 | 1.91 (0.56) | 0.95 | 5.59 (0.06) | 1.06 | 3.21 (0.13) |
| R medial orbitofrontal | 1.18 | 18.32 (<0.0001) | 1.15 | 20.26 (<0.0001) | 1.01 | 0.03 (0.98) | 0.98 | 1.71 (0.56) | 0.96 | 3.25 (0.16) | 1.08 | 9.39 (0.01) |
| R middle temporal | 1.16 | 11.98 (<0.0001) | 1.22 | 29.72 (<0.0001) | 1.02 | 0.02 (0.98) | 0.98 | 4.07 (0.35) | 0.98 | 1.14 (0.41) | 1.11 | 8.67 (0.01) |
| R para-hippocampal | 1.01 | 0.54 (0.54) | 1.21 | 33.49 (<0.0001) | 0.97 | 9.22 (0.05) | 1.05 | 0.45 (0.68) | 1.07 | 0.85 (0.48) | 1.03 | 0.06 (0.83) |
| R paracentral | 1.01 | 0.12 (0.75) | 1.15 | 7.62 (0.01) | 0.98 | 1.35 (0.61) | 0.97 | 1.24 (0.57) | 1.04 | 2.16 (0.27) | 1.04 | 1.17 (0.38) |
| R pars opercularis | 0.98 | 0.33 (0.61) | 1.15 | 31.02 (<0.0001) | 0.98 | 1.26 (0.61) | 1.02 | 2.63 (0.43) | 1.03 | 1.56 (0.35) | 1.1 | 15.74 (<0.0001) |
| R pars orbitalis | 1.15 | 14.64 (<0.0001) | 1.13 | 11.17 (<0.0001) | 1 | 1.18 (0.61) | 0.97 | 3.48 (0.39) | 0.96 | 4.25 (0.11) | 1.06 | 4.5 (0.07) |
| R pars triangularis | 1.09 | 4.94 (0.04) | 1.11 | 22.89 (<0.0001) | 1 | 0.58 (0.72) | 0.96 | 0.44 (0.68) | 0.97 | 0.63 (0.56) | 1.05 | 11.04 (0.01) |
| R pericalcarine | 1.18 | 8.33 (0.01) | 1.06 | 2.03 (0.17) | 1.04 | 0.73 (0.68) | 0.99 | 0.63 (0.63) | 0.94 | 13.3 (<0.0001) | 1.07 | 5.72 (0.04) |
| R postcentral | 1.13 | 5.97 (0.02) | 1.18 | 18.88 (<0.0001) | 1.01 | 3.93 (0.32) | 0.99 | 5.94 (0.27) | 0.98 | 6.35 (0.05) | 1.04 | 0.79 (0.47) |
| R posteriorcingulate | 1.1 | 1.15 (0.36) | 1.1 | 11.5 (<0.0001) | 0.99 | 1.03 (0.62) | 1.01 | 1.03 (0.57) | 1.02 | 0 (0.97) | 1.06 | 2.99 (0.14) |
| R precentral | 1.15 | 18.32 (<0.0001) | 1.18 | 29.72 (<0.0001) | 1 | 0.12 (0.91) | 0.99 | 0.04 (0.93) | 0.97 | 1.61 (0.35) | 1.06 | 5.17 (0.05) |

| | | | | | | | | | | | | |
|---|---|---|---|---|---|---|---|---|---|---|---|---|
| R precuneus | 1.22 | 27.51 (<0.0001) | 1.14 | 20.62 (<0.0001) | 1 | 0.01 (0.99) | 0.99 | 0.07 (0.93) | 0.95 | 3.98 (0.11) | 1.05 | 5.81 (0.04) |
| R rostral anterior cingulate | 1.04 | 0.88 (0.42) | 1.21 | 45.95 (<0.0001) | 0.97 | 4.63 (0.27) | 0.97 | 2.64 (0.43) | 1 | 0.25 (0.76) | 1.02 | 0.89 (0.44) |
| R rostral middle frontal | 1.15 | 15.25 (<0.0001) | 1.13 | 11.41 (<0.0001) | 1.01 | 0.11 (0.91) | 0.97 | 1.37 (0.57) | 0.9 | 13.06 (<0.0001) | 1.07 | 8.82 (0.01) |
| R superior frontal | 1.13 | 25.42 (<0.0001) | 1.16 | 40.95 (<0.0001) | 1 | 0 (1) | 0.98 | 0.01 (0.97) | 0.94 | 1.28 (0.39) | 1.09 | 20.99 (<0.0001) |
| R superior parietal | 1.16 | 19.36 (<0.0001) | 1.09 | 4.43 (0.04) | 1 | 2.73 (0.45) | 0.98 | 5.35 (0.27) | 0.95 | 10.6 (0.01) | 1.07 | 0.45 (0.59) |
| R superior temporal | 1.11 | 12.34 (<0.0001) | 1.22 | 49.39 (<0.0001) | 1.02 | 0 (1) | 0.98 | 1.08 (0.57) | 0.98 | 0.3 (0.74) | 1.1 | 18.06 (<0.0001) |
| R supramarginal | 1.14 | 20.21 (<0.0001) | 1.11 | 15.52 (<0.0001) | 0.98 | 5.04 (0.24) | 0.99 | 0.52 (0.65) | 0.95 | 2.13 (0.27) | 1.05 | 2.41 (0.19) |
| R frontal pole | 1 | 1.56 (0.28) | 1.08 | 1.41 (0.25) | 0.97 | 1.2 (0.61) | 1 | 0.06 (0.93) | 1.01 | 0.08 (0.86) | 1.02 | 0 (0.98) |
| R temporal pole | 0.97 | 4.82 (0.05) | 1.11 | 4.98 (0.03) | 1.01 | 0.1 (0.91) | 1 | 0.05 (0.93) | 0.98 | 5.18 (0.07) | 0.99 | 2.33 (0.2) |
| R transverse temporal | 0.99 | 1.56 (0.28) | 1.2 | 37.72 (<0.0001) | 0.99 | 5.81 (0.18) | 1.05 | 1.43 (0.57) | 1.08 | 4.83 (0.08) | 1.03 | 0.06 (0.83) |
| R insula | 1.18 | 22.16 (<0.0001) | 1.14 | 16.57 (<0.0001) | 0.98 | 3.01 (0.45) | 0.97 | 2.25 (0.5) | 0.97 | 4.15 (0.11) | 1.06 | 5.81 (0.04) |

**Table S23** *Within-diagnosis variability in subcortical volumes.*

Variability was measured by standard deviation (Sd). Differences in variability between individuals with and without a neurodevelopmental or psychiatric condition were estimated via Levene's test. The control groups were matched via propensity score matching, taking age and sex into account. FDR correction was performed at an alpha level <0.05. ANXG = generalized anxiety disorder, ASD = autism spectrum diagnosis, BP = bipolar disorder, MDD = major depressive disorder, OCD = obsessive-compulsive disorder, SCZ = Schizophrenia spectrum.

| | **ANXG** | | **ASD** | | **BP** | | **MDD** | | **OCD** | | **SCZ** | |
|---|---|---|---|---|---|---|---|---|---|---|---|---|
| **Structure** | Sd | Levene (pFDR) | Sd | Levene (pFDR) | Sd | Levene (pFDR) | Sd | Levene (pFDR) | Sd | Levene (pFDR) | Sd | Levene (pFDR) |
| Laccumb | 0.99 | 0.21 (0.91) | 1.2 | 51.82 (<0.0001) | 1.04 | 5.47 (0.07) | 0.99 | 0.03 (0.86) | 1.11 | 16.06 (<0.0001) | 1.12 | 33.7 (<0.0001) |
| Lamyg | 1.08 | 4.38 (0.13) | 1.21 | 27.13 (<0.0001) | 1.11 | 7.12 (0.05) | 0.99 | 0.21 (0.82) | 1.04 | 1.95 (0.18) | 1.09 | 16.35 (<0.0001) |
| Lcaud | 1.04 | 0.01 (0.94) | 1.16 | 22.32 (<0.0001) | 1 | 0.01 (0.94) | 1 | 0.98 (0.67) | 1.03 | 3.67 (0.09) | 1.09 | 14.84 (<0.0001) |
| Lhippo | 1.04 | 0.02 (0.94) | 1.22 | 28.47 (<0.0001) | 1.07 | 0.59 (0.69) | 1.03 | 0.24 (0.82) | 1.09 | 3.65 (0.09) | 1.11 | 13.24 (<0.0001) |
| Lpal | 1.13 | 1.97 (0.34) | 1.12 | 12.03 (<0.0001) | 1.01 | 0.01 (0.94) | 1.08 | 7.43 (0.05) | 1.09 | 8.66 (0.01) | 1.07 | 8.65 (<0.0001) |
| Lput | 1.09 | 1.62 (0.36) | 1.17 | 23.78 (<0.0001) | 1.01 | 0.02 (0.94) | 1.01 | 1.24 (0.67) | 1.12 | 19.33 (<0.0001) | 1.1 | 25.63 (<0.0001) |
| Lthal | 1.06 | 1.28 (0.4) | 1.16 | 21.7 (<0.0001) | 1.06 | 6.07 (0.06) | 1 | 0.93 (0.67) | 1.04 | 5.14 (0.05) | 1.07 | 10.26 (<0.0001) |
| Raccumb | 1 | 0.06 (0.94) | 1.23 | 46.57 (<0.0001) | 1.06 | 2.22 (0.32) | 1.02 | 0.09 (0.86) | 1.08 | 3.08 (0.11) | 1.09 | 15.68 (<0.0001) |
| Ramyg | 1.08 | 3.14 (0.21) | 1.21 | 31.73 (<0.0001) | 1.03 | 1.85 (0.35) | 0.99 | 1.11 (0.67) | 1.06 | 5.14 (0.05) | 1.08 | 20.62 (<0.0001) |
| Rcaud | 1.03 | 0.01 (0.94) | 1.21 | 46.49 (<0.0001) | 1.01 | 0.18 (0.93) | 1 | 0.28 (0.82) | 1.02 | 2.42 (0.14) | 1.09 | 13.7 (<0.0001) |
| Rhippo | 1.05 | 1.87 (0.34) | 1.19 | 25.58 (<0.0001) | 1.06 | 3.22 (0.2) | 1.04 | 2.22 (0.64) | 1.05 | 4.47 (0.07) | 1.1 | 23.3 (<0.0001) |
| Rpal | 1.12 | 9.37 (0.02) | 1.14 | 24.05 (<0.0001) | 1.01 | 0.85 (0.62) | 1.09 | 13.38 (<0.0001) | 1 | 0 (0.95) | 1.07 | 10.36 (<0.0001) |
| Rput | 1.13 | 4.88 (0.13) | 1.21 | 26.24 (<0.0001) | 1.01 | 0.12 (0.93) | 1 | 0.03 (0.86) | 1.1 | 10.53 (0.01) | 1.14 | 28.48 (<0.0001) |
| Rthal | 1.09 | 17.74 (<0.0001) | 1.09 | 13.4 (<0.0001) | 1.05 | 8 (0.05) | 0.98 | 0.39 (0.82) | 1.01 | 2.49 (0.14) | 1.11 | 26.82 (<0.0001) |

**Table S24** *Robustness of variability findings*.

Spearman's correlations between variability maps derived from standard deviations and maps derived from interquartile range (IQR), median absolute deviation MAD), or weighted standard deviation (wSD). ANXG = generalized anxiety disorder, ASD = autism spectrum diagnosis, BP = bipolar disorder, MDD = major depressive disorder, OCD = obsessive-compulsive disorder, SCZ = Schizophrenia spectrum, CT = cortical thickness, SA = surface area, SV = subcortical volumes.

| | **ANXG** | **ASD** | **BP** | **MDD** | **OCD** | **SCZ** |
|---|---|---|---|---|---|---|
| CT IQR | 0.74*** | 0.74*** | 0.86*** | 0.72*** | 0.65*** | 0.64*** |
| SA IQR | 0.84*** | 0.79*** | 0.77*** | 0.64*** | 0.85*** | 0.84*** |
| SV IQR | 0.77*** | 0.32 | 0.65* | 0.84** | 0.87*** | 0.81** |
| CT MAD | 0.75*** | 0.74*** | 0.84*** | 0.74*** | 0.67*** | 0.65*** |
| SA MAD | 0.83*** | 0.81*** | 0.77*** | 0.67*** | 0.85*** | 0.83*** |
| SV MAD | 0.73** | 0.28 | 0.67** | 0.94*** | 0.82** | 0.78** |
| CT wSD | 0.98*** | 0.74*** | 0.96*** | 0.94*** | 0.96*** | 0.93*** |
| SA wSD | 1*** | 0.92*** | 0.96*** | 0.93*** | 0.99*** | 0.99*** |
| SV wSD | 0.98*** | 0.76*** | 0.91*** | 0.97*** | 0.98*** | 0.97*** |

Variogram permutation. *** = p<0.001, ** = p<0.01, *=p<0.05

**Table S25** *Spearman's rho correlation between Cohen's d and variability (standard deviation) maps, for cortical thickness data.*

Significance was assessed via variogram permutations (10,000 permutations). ANXG = generalized anxiety disorder, ASD = autism spectrum diagnosis, BP = bipolar disorder, MDD = major depressive disorder, OCD = obsessive-compulsive disorder, SCZ = Schizophrenia spectrum.

| **Disorder** | **Rho** | $p_{variogram}$ | $p_{variogram}$ **FDR** |
|---|---|---|---|
| ANXG | 0.16 | 0.41 | 0.83 |
| ASD | 0.02 | 0.93 | 0.98 |
| BP | 0.15 | 0.24 | 0.72 |
| MDD | -0.11 | 0.60 | 0.90 |
| OCD | 0.005 | 0.98 | 0.98 |
| SCZ | -0.18 | 0.24 | 0.72 |

**Table S26** *Leave-one-NDPC-out axes of deviation.*

Spearman's rho correlation between original principal components (PCs) computed in the full test sample and PCs derived after leaving one neurodevelopmental or psychiatric condition (NDPC) or reference cohort (RC) out. Note that PCs are direction-less and may flip, leading to negative correlations. Variogram test. All $p<0.0001$. ANXG = generalized anxiety disorder, ASD = autism spectrum diagnosis, BP = bipolar disorder, MDD = major depressive disorder, OCD = obsessive-compulsive disorder, SCZ = Schizophrenia spectrum, CT = cortical thickness, SA = surface area, SV = subcortical volumes.

| Modality | PC | ANXG | ASD | BP | MDD | OCD | SCZ | RC |
|---|---|---|---|---|---|---|---|---|
| **CT** | PC1 | 0.999 | 0.993 | 0.999 | 0.998 | 0.998 | 0.997 | 0.998 |
| **CT** | PC2 | 0.999 | 0.983 | 0.997 | 0.997 | 0.988 | 0.995 | 0.997 |
| **SA** | PC1 | 0.999 | 0.998 | 0.999 | 0.999 | 0.999 | 0.997 | 0.998 |
| **SA** | PC2 | 0.996 | 0.991 | 0.996 | 0.996 | 0.997 | 0.991 | 0.997 |
| **SV** | PC1 | 0.996 | 0.982 | 0.996 | 1 | 1 | 0.991 | 0.996 |
| **SV** | PC2 | 1.0 | -0.996 | 0.987 | 0.991 | 0.996 | 0.991 | 0.978 |

**Table S27** *Leave-sites-out axes of deviation.*

Spearman's rho correlations for the spatial associations between principal components (PC1 and PC2) derived from the full sample compared to a subsample where either one site or 10% of sites were left out. This procedure was repeated iteratively until each site, or set of sites, was left out once. The table reports minimum, maximum, and mean correlations across these iterations. CT = Cortical thickness, SA = surface area, SV = subcortical volume.

| **Measure** | **Test** | **PC** | **Minimum Rho** | **Maximum Rho** | **Mean Rho** | **Standard deviation** | **Number of iterations** |
|---|---|---|---|---|---|---|---|
| **CT** | Leave-1-site-out | PC1 | 1.0 | 1.0 | 1.0 | 0 | 172 |
| **CT** | Leave-1-site-out | PC2 | 0.99 | 1.0 | 1.0 | 0 | 172 |
| **CT** | Leave-10%-sites-out | PC1 | 1.0 | 1.0 | 1.0 | 0 | 10 |
| **CT** | Leave-10%-sites-out | PC2 | 0.98 | 1.0 | 0.99 | 0.01 | 10 |
| **SA** | Leave-1-site-out | PC1 | 1.0 | 1.0 | 1.0 | 0 | 172 |
| **SA** | Leave-1-site-out | PC2 | 1.0 | 1.0 | 1.0 | 0 | 172 |
| **SA** | Leave-10%-sites-out | PC1 | 1.0 | 1.0 | 1.0 | 0 | 10 |
| **SA** | Leave-10%-sites-out | PC2 | 0.99 | 1.0 | 1.0 | 0 | 10 |
| **SV** | Leave-1-site-out | PC1 | 0.99 | 1.0 | 1.0 | 0 | 172 |
| **SV** | Leave-1-site-out | PC2 | 0.98 | 1.0 | 1.0 | 0 | 172 |
| **SV** | Leave-10%-sites-out | PC1 | 0.98 | 1.0 | 0.99 | 0.01 | 10 |
| **SV** | Leave-10%-sites-out | PC2 | 0.96 | 1.0 | 0.99 | 0.01 | 10 |

**Table S28** *Association between symptom magnitude and PC scores.*

Linear regressions between symptom severity and PC scores were controlled for age and sex. Statistical values (except for $R^2$) refer to the PC term in the model. STAI_T = State-Trait Anxiety Inventory (Trait subscale), ADOS_CSS = Autism Diagnostic Observation Schedule calibrated severity score, HDRS = Hamilton Depression Scale, Y-BOCS = Yale-Brown Obsessive Compulsive Scale, PANSS = Positive and negative symptom score, NDPC = neurodevelopmental or psychiatric condition; ANXG = generalized anxiety disorder, ASD = autism spectrum diagnosis, BP = bipolar disorder, MDD = major depressive disorder, OCD = obsessive-compulsive disorder, SCZ = Schizophrenia spectrum. FDR $p<0.05$

| Measure | NDPC | Symptom measure | *n* | $\beta_{standardized}$ | t | SE | $R^2$ | $P_{FDR}$ |
|---|---|---|---|---|---|---|---|---|
| CT – PC1 | ANXG | STAI_T | 411 | 0.003 | 0.064 | 0.049 | 0.043 | 0.949 |
| CT – PC2 | ANXG | STAI_T | 411 | 0.024 | 0.492 | 0.049 | 0.043 | 0.949 |
| CT – PC1 | ASD | ADOS_CSS | 463 | 0.127 | 2.766 | 0.046 | 0.039 | 0.032* |
| CT – PC2 | ASD | ADOS_CSS | 463 | 0.075 | 1.629 | 0.046 | 0.028 | 0.312 |
| CT – PC1 | OCD | Y-BOCS | 129 | 0.090 | 1.055 | 0.086 | 0.134 | 0.704 |
| CT – PC2 | OCD | Y-BOCS | 129 | -0.011 | -0.130 | 0.084 | 0.127 | 0.949 |
| CT – PC1 | MDD | HDRS | 1108 | 0.079 | 2.654 | 0.030 | 0.025 | 0.032* |
| CT – PC2 | MDD | HDRS | 1108 | -0.023 | -0.770 | 0.030 | 0.020 | 0.882 |
| CT – PC1 | BP | PANSS_Total_combined | 1719 | 0.006 | 0.257 | 0.024 | 0.016 | 0.949 |
| CT – PC2 | BP | PANSS_Total_combined | 1719 | -0.003 | -0.120 | 0.024 | 0.016 | 0.949 |
| CT – PC1 | SCZ | PANSS_Total_combined | 1014 | 0.143 | 4.630 | 0.031 | 0.039 | 0.00005* |
| CT – PC2 | SCZ | PANSS_Total_combined | 1014 | -0.014 | -0.444 | 0.031 | 0.019 | 0.949 |
| SA – PC1 | ANXG | STAI_T | 411 | -0.032 | -0.630 | 0.051 | 0.044 | 0.818 |
| SA – PC2 | ANXG | STAI_T | 411 | -0.104 | -2.135 | 0.049 | 0.053 | 0.170 |
| SA – PC1 | ASD | ADOS_CSS | 463 | -0.001 | -0.032 | 0.046 | 0.023 | 0.994 |
| SA – PC2 | ASD | ADOS_CSS | 463 | -0.0003 | -0.008 | 0.046 | 0.023 | 0.994 |
| SA – PC1 | OCD | Y-BOCS | 129 | -0.085 | -0.979 | 0.087 | 0.133 | 0.659 |
| SA – PC2 | OCD | Y-BOCS | 129 | 0.002 | 0.023 | 0.084 | 0.126 | 0.994 |
| SA – PC1 | MDD | HDRS | 1108 | 0.037 | 1.225 | 0.030 | 0.020 | 0.659 |
| SA – PC2 | MDD | HDRS | 1108 | 0.017 | 0.567 | 0.030 | 0.019 | 0.818 |
| SA – PC1 | BP | PANSS_Total_combined | 1719 | 0.012 | 0.505 | 0.024 | 0.016 | 0.818 |
| SA – PC2 | BP | PANSS_Total_combined | 1719 | -0.049 | -2.031 | 0.024 | 0.018 | 0.170 |
| SA – PC1 | SCZ | PANSS_Total_combined | 1014 | 0.149 | 4.797 | 0.031 | 0.041 | 0.00005* |
| SA – PC2 | SCZ | PANSS_Total_combined | 1014 | -0.033 | -1.071 | 0.031 | 0.020 | 0.659 |

| | | | | | | | | |
|---|---|---|---|---|---|---|---|---|
| SV – PC1 | ANXG | STAI_T | 411 | -0.026 | -0.527 | 0.049 | 0.043 | 0.994 |
| SV – PC2 | ANXG | STAI_T | 411 | 0.016 | 0.322 | 0.049 | 0.043 | 0.994 |
| SV – PC2 | ASD | ADOS_CSS | 463 | -0.037 | -0.795 | 0.046 | 0.024 | 0.8536 |
| SV – PC1 | ASD | ADOS_CSS | 463 | -0.078 | -1.680 | 0.046 | 0.029 | 0.375 |
| SV – PC1 | OCD | Y-BOCS | 129 | -0.018 | -0.208 | 0.088 | 0.127 | 0.994 |
| SV – PC2 | OCD | Y-BOCS | 129 | 0.007 | 0.088 | 0.084 | 0.126 | 0.994 |
| SV – PC1 | MDD | HDRS | 1108 | 0.0002 | 0.008 | 0.030 | 0.019 | 0.994 |
| SV – PC2 | MDD | HDRS | 1108 | 0.060 | 2.020 | 0.030 | 0.023 | 0.262 |
| SV – PC1 | BP | PANSS_Total_combined | 1719 | 0.028 | 1.179 | 0.024 | 0.016 | 0.716 |
| SV – PC2 | BP | PANSS_Total_combined | 1719 | 0.019 | 0.810 | 0.024 | 0.016 | 0.854 |
| SV – PC1 | SCZ | PANSS_Total_combined | 1014 | 0.100 | 3.162 | 0.032 | 0.029 | 0.019* |
| SV – PC2 | SCZ | PANSS_Total_combined | 1014 | -0.005 | -0.162 | 0.031 | 0.019 | 0.994 |

**Table S29** *Linear regressions between time since diagnosis and PC scores for each neurodevelopmental and psychiatric condition (NDPC).*

Models were controlled for age and sex. Statistical values (except for $R^2$) refer to the PC term in the model. ANXG = generalized anxiety disorder, ASD = autism spectrum diagnosis, BP = bipolar disorder, MDD = major depressive disorder, OCD = obsessive-compulsive disorder, SCZ = Schizophrenia spectrum, CT = cortical thickness, SA = surface area, SV = subcortical volumes. FDR $p<0.05$

| Measure | NDPC | PC | n | $\beta_{standardized}$ | SE | R2 | $p_{PC\text{-}FDR}$ |
|---|---|---|---|---|---|---|---|
| CT | ANXG | PC1 | 179 | 0.016 | 0.063 | 0.312 | 0.803 |
| CT | ANXG | PC2 | 179 | 0.054 | 0.063 | 0.315 | 0.569 |
| CT | MDD | PC1 | 1515 | -0.015 | 0.023 | 0.175 | 0.615 |
| CT | MDD | PC2 | 1515 | 0.014 | 0.023 | 0.175 | 0.615 |
| CT | BP | PC1 | 1111 | 0.024 | 0.023 | 0.42 | 0.569 |
| CT | BP | PC2 | 1111 | 0.019 | 0.023 | 0.42 | 0.569 |
| CT | SCZ | PC1 | 1323 | 0.038 | 0.017 | 0.623 | 0.151 |
| CT | SCZ | PC2 | 1323 | -0.016 | 0.017 | 0.621 | 0.569 |
| CT | OCD | PC1 | 1531 | 0.043 | 0.02 | 0.395 | 0.151 |
| CT | OCD | PC2 | 1531 | -0.027 | 0.02 | 0.394 | 0.568 |
| SA | ANXG | PC1 | 179 | 0.04 | 0.063 | 0.314 | 0.677 |
| SA | ANXG | PC2 | 179 | -0.009 | 0.063 | 0.312 | 0.883 |
| SA | MDD | PC1 | 1515 | 0.016 | 0.024 | 0.175 | 0.677 |
| SA | MDD | PC2 | 1515 | 0.014 | 0.023 | 0.175 | 0.677 |
| SA | BP | PC1 | 1111 | -0.022 | 0.023 | 0.42 | 0.677 |
| SA | BP | PC2 | 1111 | 0.031 | 0.023 | 0.421 | 0.606 |
| SA | SCZ | PC1 | 1323 | -0.026 | 0.017 | 0.622 | 0.606 |
| SA | SCZ | PC2 | 1323 | -0.013 | 0.017 | 0.621 | 0.677 |
| SA | OCD | PC1 | 1531 | 0.028 | 0.02 | 0.394 | 0.606 |
| SA | OCD | PC2 | 1531 | -0.004 | 0.02 | 0.393 | 0.883 |
| SV | ANXG | PC1 | 179 | 0.016 | 0.063 | 0.312 | 0.884 |
| SV | ANXG | PC2 | 179 | 0.066 | 0.063 | 0.316 | 0.6 |
| SV | MDD | PC1 | 1515 | 0.013 | 0.023 | 0.175 | 0.739 |
| SV | MDD | PC2 | 1515 | -0.015 | 0.023 | 0.175 | 0.728 |
| SV | BP | PC1 | 1111 | -0.034 | 0.023 | 0.421 | 0.369 |
| SV | BP | PC2 | 1111 | 0.003 | 0.023 | 0.42 | 0.899 |
| SV | SCZ | PC1 | 1323 | 0.025 | 0.017 | 0.622 | 0.369 |
| SV | SCZ | PC2 | 1323 | -0.014 | 0.017 | 0.621 | 0.688 |
| SV | OCD | PC1 | 1531 | -0.033 | 0.02 | 0.394 | 0.369 |
| SV | OCD | PC2 | 1531 | -0.032 | 0.02 | 0.394 | 0.369 |

**Table S30** *Differences in PC scores between medicated and unmedicated individuals.*
General linear models testing for group differences in medicated compared to unmedicated individuals in principal component (PC) scores, controlling for age and sex. For SCZ and BP, medication categories were tested in separate, mutually exclusive contrasts. The control group consisted of patients not receiving any of the medication classes included in the respective comparison, and were matched in n via propensity score matching (taking age and sex into account). Statistical values (except R2 refer to the medication term of the linear model). ANXG = generalized anxiety disorder, ASD = autism spectrum diagnosis, BP = bipolar disorder, MDD = major depressive disorder, OCD = obsessive-compulsive disorder, SCZ = Schizophrenia spectrum. *pFDR <0.05

| NDPC | Measure | N (n per group) | $\beta_{standardized}$ | SE | $R^2$ | p FDR |
|---|---|---|---|---|---|---|
| MDD – Antidepressants | CT – PC1 | 334 (167) | 0.144 | 0.116 | 0.011 | 0.621 |
| MDD – Antidepressants | CT – PC2 | 334 (167) | -0.259 | 0.115 | 0.036 | 0.293 |
| OCD – Any | CT – PC1 | 1672 (836) | 0.171 | 0.049 | 0.008 | 0.03* |
| OCD – Any | CT – PC2 | 1672 (836) | 0.039 | 0.049 | 0.001 | 0.682 |
| ASD – Any | CT – PC1 | 680(340) | 0.013 | 0.076 | 0.031 | 0.945 |
| ASD – Any | CT – PC2 | 680 (340) | 0.068 | 0.076 | 0.013 | 0.654 |
| SCZ – Atypical antipsychotics | CT – PC1 | 462 (231) | -0.057 | 0.093 | 0.003 | 0.768 |
| SCZ – Atypical antipsychotics | CT – PC2 | 462 (231) | -0.047 | 0.093 | 0.009 | 0.842 |
| SCZ – Typical antipsychotics | CT – PC1 | 84 (42) | 0.618 | 0.208 | 0.134 | 0.118 |
| SCZ – Typical antipsychotics | CT – PC2 | 84 (42) | -0.199 | 0.222 | 0.018 | 0.654 |
| SCZ – Typical & atypical antipsychotics | CT – PC1 | 412 (206) | 0.252 | 0.099 | 0.019 | 0.223 |
| SCZ – Typical & atypical antipsychotics | CT – PC2 | 412 (206) | 0.143 | 0.099 | 0.008 | 0.621 |
| BP – Atypical antipsychotics | CT – PC1 | 62 (31) | 0.22 | 0.26 | 0.018 | 0.654 |
| BP – Atypical antipsychotics | CT – PC2 | 62 (31) | 0.395 | 0.253 | 0.072 | 0.621 |
| BP – Atypical antipsychotics | CT – PC1 | 442 (221) | 0.003 | 0.095 | 0.009 | 0.977 |
| BP – Atypical antipsychotics | CT – PC2 | 442 (221) | 0.115 | 0.095 | 0.007 | 0.621 |
| BP – Lithium | CT – PC1 | 144 (72) | 0.067 | 0.167 | 0.025 | 0.901 |
| BP – Lithium | CT – PC2 | 144 (72) | -0.178 | 0.164 | 0.054 | 0.623 |
| BP – Antidepressants | CT – PC1 | 306 (153) | 0.217 | 0.114 | 0.018 | 0.494 |
| BP – Antidepressants | CT – PC2 | 306 (153) | 0.117 | 0.114 | 0.014 | 0.623 |
| MDD - Antidepressants | SA – PC1 | 334 (167) | 0.118 | 0.112 | 0.075 | 0.623 |
| MDD - Antidepressants | SA – PC1 | 334 (167) | 0.146 | 0.116 | 0.013 | 0.621 |
| OCD – Any | SA – PC2 | 1672 (836) | 0.036 | 0.049 | 0.018 | 0.708 |
| OCD – Any | SA – PC2 | 1672 (836) | 0.029 | 0.049 | 0 | 0.768 |
| ASD – Any | SA – PC1 | 680 (340) | -0.015 | 0.076 | 0.015 | 0.945 |
| ASD - Any | SA – PC2 | 680 (340) | 0.088 | 0.077 | 0.011 | 0.623 |
| SCZ – Atypical antipsychotics | SA – PC1 | 462 (231) | -0.097 | 0.093 | 0.012 | 0.623 |
| SCZ – Atypical antipsychotics | SA – PC2 | 462 (231) | -0.01 | 0.093 | 0.001 | 0.972 |
| SCZ – Typical antipsychotics | SA – PC1 | 84 (42) | -0.267 | 0.216 | 0.072 | 0.621 |
| SCZ – Typical antipsychotics | SA – PC2 | 84 (42) | -0.045 | 0.223 | 0.006 | 0.945 |

| | | | | | | |
|---|---|---|---|---|---|---|
| SCZ – Typical & atypical antipsychotics | SA – PC1 | 412 (206) | -0.03 | 0.099 | 0.005 | 0.94 |
| SCZ – Typical & atypical antipsychotics | SA – PC2 | 412 (206) | -0.094 | 0.099 | 0.008 | 0.654 |
| BP – Typical antipsychotics | SA – PC1 | 62 (31) | 0.32 | 0.249 | 0.103 | 0.621 |
| BP – Typical antipsychotics | SA – PC2 | 62 (31) | -0.466 | 0.253 | 0.072 | 0.53 |
| BP – Atypical antipsychotics | SA – PC1 | 442 (221) | 0.033 | 0.094 | 0.041 | 0.929 |
| BP – Atypical antipsychotics | SA – PC2 | 442 (221) | -0.121 | 0.095 | 0.008 | 0.621 |
| BP – Lithium | SA – PC1 | 144 (72) | 0.071 | 0.164 | 0.059 | 0.888 |
| BP – Lithium | SA – PC2 | 144 (72) | 0.145 | 0.167 | 0.027 | 0.654 |
| BP – Antidepressants | SA – PC1 | 306 (153) | -0.007 | 0.112 | 0.056 | 0.976 |
| BP – Antidepressants | SA – PC2 | 306 (153) | -0.116 | 0.114 | 0.013 | 0.623 |
| MDD - Antidepressants | SV – PC1 | 334 (167) | 0.194 | 0.116 | 0.021 | 0.621 |
| MDD - Antidepressants | SV – PC2 | 334 (167) | -0.147 | 0.116 | 0.018 | 0.621 |
| OCD - Any | SV – PC1 | 1672 (836) | 0.054 | 0.049 | 0.012 | 0.623 |
| OCD – Any | SV – PC2 | 1672 (836) | 0.113 | 0.049 | 0.007 | 0.293 |
| ASD – Any | SV – PC1 | 680 (340) | -0.115 | 0.076 | 0.019 | 0.621 |
| ASD – Any | SV – PC2 | 680 (340) | -0.084 | 0.076 | 0.011 | 0.623 |
| SCZ – Atypical antipsychotics | SV – PC1 | 462 (231) | -0.078 | 0.093 | 0.013 | 0.654 |
| SCZ – Atypical antipsychotics | SV – PC2 | 462 (231) | 0.024 | 0.093 | 0.016 | 0.94 |
| SCZ – Typical antipsychotics | SV – PC1 | 84 (42) | 0.308 | 0.214 | 0.083 | 0.621 |
| SCZ – Typical antipsychotics | SV – PC2 | 84 (42) | -0.426 | 0.216 | 0.067 | 0.494 |
| SCZ – Typical & atypical antipsychotics | SV – PC1 | 412 (206) | 0.025 | 0.098 | 0.03 | 0.94 |
| SCZ – Typical & atypical antipsychotics | SV – PC2 | 412 (206) | 0.071 | 0.1 | 0.003 | 0.713 |
| BP – Typical antipsychotics | SV – PC1 | 62 (31) | 0.068 | 0.251 | 0.088 | 0.94 |
| BP – Typical antipsychotics | SV – PC2 | 62 (31) | -0.012 | 0.248 | 0.108 | 0.976 |
| BP – Atypical | SV – PC1 | 442 (221) | -0.084 | 0.093 | 0.05 | 0.654 |
| BP – Atypical antipsychotics | SV – PC2 | 442 (221) | 0.126 | 0.095 | 0.017 | 0.621 |
| BP – Lithium | SV – PC1 | 144 (72) | -0.236 | 0.162 | 0.077 | 0.621 |
| BP – Lithium | SV – PC2 | 144 (72) | -0.016 | 0.166 | 0.034 | 0.972 |
| BP – Antidepressants | SV – PC1 | 306 (153) | -0.078 | 0.114 | 0.025 | 0.722 |
| BP – Antidepressants | SV – PC2 | 306 (153) | -0.021 | 0.114 | 0.019 | 0.945 |

*= $p_{FDR}<0.05$. Note that for OCD, medication effects further interacted with symptom severity for PC1(beta: 0.17, SE = 0.05, pFDR = 0.0013) but not PC2 (beta: 0.02, SE = 0.05, pFDR = 0.64)

**Table S31** *Overall percentage of individuals showing at least one extreme deviation |z|>1.96 per disorder group and modality, in any region.*

ANXG = generalized anxiety disorder, ASD = autism spectrum diagnosis, BP = bipolar disorder, MDD = major depressive disorder, OCD = obsessive-compulsive disorder, SCZ = Schizophrenia spectrum, CT = cortical thickness, SA = surface area, SV = subcortical volumes.

| Measure | Diagnosis | % any extreme deviation |
|---|---|---|
| CT | ANXG | 73.20 |
| CT | ASD | 84.64 |
| CT | BP | 78.32 |
| CT | MDD | 80.11 |
| CT | OCD | 88.06 |
| CT | SCZ | 83.40 |
| CT | RC | 76.93 |
| SA | ANXG | 81.05 |
| SA | ASD | 84.19 |
| SA | BP | 76.86 |
| SA | MDD | 77.92 |
| SA | OCD | 75.94 |
| SA | SCZ | 81.08 |
| SA | RC | 77.98 |
| SV | ANXG | 38.30 |
| SV | ASD | 50.32 |
| SV | BP | 39.34 |
| SV | MDD | 36.69 |
| SV | OCD | 42.65 |
| SV | SCZ | 45.66 |
| SV | RC | 33.94 |

**Table S32** *Spearman's correlations between original Cohen's d maps and extreme deviation overlap maps*. Positive and negative extreme deviation maps were assessed separately, and p-values were adjusted for spatial auto-correlation via variogram permutations for cortical thickness (CT) and surface area (SA), and subcortical volume (SV; 10,000 permutations). ANXG = generalized anxiety disorder, ASD = autism spectrum diagnosis, BP = bipolar disorder, MDD = major depressive disorder, OCD = obsessive-compulsive disorder, SCZ = Schizophrenia spectrum.

| Diagnosis | Measure | Rho | P_spin |
|---|---|---|---|
| ANXG | CT Negative | -0.125 | 0.537 |
| ANXG | CT Positive | 0.295 | 0.025 |
| ASD | CT Negative | -0.646 | 0.002 |
| ASD | CT Positive | 0.634 | 0.0001 |
| BP | CT Negative | -0.381 | 0.014 |
| BP | CT Positive | 0.500 | 0.001 |
| MDD | CT Negative | -0.379 | 0.022 |
| MDD | CT Positive | 0.301 | 0.024 |
| OCD | CT Negative | -0.378 | 0.009 |
| OCD | CT Positive | 0.413 | 0.037 |
| SCZ | CT Negative | -0.850 | 0.0001 |
| SCZ | CT Positive | 0.767 | 0.0001 |
| ANXG | SA Negative | -0.210 | 0.376 |
| ANXG | SA Positive | 0.487 | 0.0004 |
| ASD | SA Negative | -0.567 | 0.0002 |
| ASD | SA Positive | 0.4312 | 0.0002 |
| BP | SA Negative | -0.398 | 0.002 |
| BP | SA Positive | 0.399 | 0.002 |
| MDD | SA Negative | -0.573 | 0.0001 |
| MDD | SA Positive | 0.252 | 0.016 |
| OCD | SA Negative | -0.141 | 0.184 |
| OCD | SA Positive | 0.532 | 0.0001 |
| SCZ | SA Negative | -0.698 | 0.0001 |
| SCZ | SA Positive | 0.379 | 0.0006 |
| ANXG | SV Negative | -0.068 | 0.844 |
| ANXG | SV Positive | 0.763 | 0.002 |
| ASD | SV Negative | -0.904 | 0.0001 |
| ASD | SV Positive | 0.381 | 0.181 |
| BP | SV Negative | -0.784 | 0.002 |
| BP | SV Positive | 0.196 | 0.420 |
| MDD | SV Negative | -0.574 | 0.023 |
| MDD | SV Positive | 0.826 | 0.0002 |
| OCD | SV Negative | -0.519 | 0.048 |
| OCD | SV Positive | 0.359 | 0.216 |
| SCZ | SV Negative | -0.952 | 0.0001 |
| SCZ | SV Positive | 0.950 | 0.0001 |

**Table S33** *Spearman's correlations between original Cohen's d maps and those derived from samples without extreme deviations (|z|> 1.96).*

P-values were adjusted for spatial auto-correlation via variogram permutations (10,000 permutations). ANXG = generalized anxiety disorder, ASD = autism spectrum diagnosis, BP = bipolar disorder, MDD = major depressive disorder, OCD = obsessive-compulsive disorder, SCZ = Schizophrenia spectrum. CT = cortical thickness, SA = surface area, SV = subcortical volume.

| Diagnosis | Measure | Rho | P_spin |
|---|---|---|---|
| ANXG | CT | 0.636 | 0.0001 |
| ASD | CT | 0.520 | 0.0003 |
| BP | CT | 0.618 | 0.0001 |
| MDD | CT | 0.524 | 0.0002 |
| OCD | CT | 0.350 | 0.005 |
| SCZ | CT | 0.714 | 0.0001 |
| ANXG | SA | 0.478 | 0.0001 |
| ASD | SA | 0.597 | 0.0001 |
| BP | SA | 0.533 | 0.0001 |
| MDD | SA | 0.535 | 0.0001 |
| OCD | SA | 0.576 | 0.0001 |
| SCZ | SA | 0.483 | 0.0001 |
| ANXG | SV | 0.792 | 0.0007 |
| ASD | SV | 0.839 | 0.0001 |
| BP | SV | 0.954 | 0.0001 |
| MDD | SV | 0.854 | 0.0002 |
| OCD | SV | 0.866 | 0.0002 |
| SCZ | SV | 0.980 | 0.0001 |

**Table S34** *Transdiagnostic overlap robustness analysis using Kolmogorov-Smirnov distance.*

We tested the robustness of observed transdiagnostic overlap patterns by comparing similarity maps computed as shared area under any two distributions to the Kolmogorov-Smirnov distance between distributions. Note that negative Spearman correlations indicate similar results, as Kolmogorov-Smirnov distance is a distance / dissimilarity measure (where 0 = complete overlap, 1 = maximal distance). P-values were derived from spin tests (for cortical measures) and variogram tests (for subcortical measures) using 10,000 permutations. CT = cortical thickness, SA = surface area, SV = subcortical volumes.

| **Measure** | **Spearman's rho** | $p_{variogram}$ |
|---|---|---|
| CT | -0.96 | <0.0001 |
| SA | -0.85 | <0.0001 |
| SV | -0.92 | <0.0001 |

**Table S35** *Pairwise distribution overlap between individuals with a neurodevelopmental or psychiatric condition (NDPC) and the reference cohort, computed for each region separately.*

ANXG = generalized anxiety disorder, ASD = autism spectrum diagnosis, BP = bipolar disorder, MDD = major depressive disorder, OCD = obsessive-compulsive disorder, SCZ = Schizophrenia spectrum, CT = cortical thickness, SA = surface area, SV = subcortical volumes.

| Diagnosis | Mean overlap | Minimum overlap | Maximum overlap | Measure |
|---|---|---|---|---|
| ANXG | 95.15 | 91.44 | 98.53 | CT |
| ASD | 90.68 | 86.24 | 94.31 | CT |
| BP | 93.18 | 89.23 | 97.08 | CT |
| MDD | 96.43 | 93.31 | 98.40 | CT |
| OCD | 94.06 | 89.28 | 97.41 | CT |
| SCZ | 86.4 | 79.82 | 96.62 | CT |
| ANXG | 91.14 | 85.99 | 96.92 | SA |
| ASD | 94.06 | 90.36 | 98.03 | SA |
| BP | 96.72 | 94.72 | 98.26 | SA |
| MDD | 96.31 | 93.68 | 98.17 | SA |
| OCD | 95.60 | 91.75 | 97.97 | SA |
| SCZ | 93.68 | 90.61 | 97.80 | SA |
| ANXG | 91.05 | 88.61 | 94.64 | SV |
| ASD | 92.28 | 88.91 | 93.77 | SV |
| BP | 95.31 | 92.31 | 97.82 | SV |
| MDD | 95.93 | 93.77 | 97.89 | SV |
| OCD | 95.60 | 93.20 | 97.28 | SV |
| SCZ | 89.81 | 83.24 | 94.78 | SV |

### Supplementary Figures

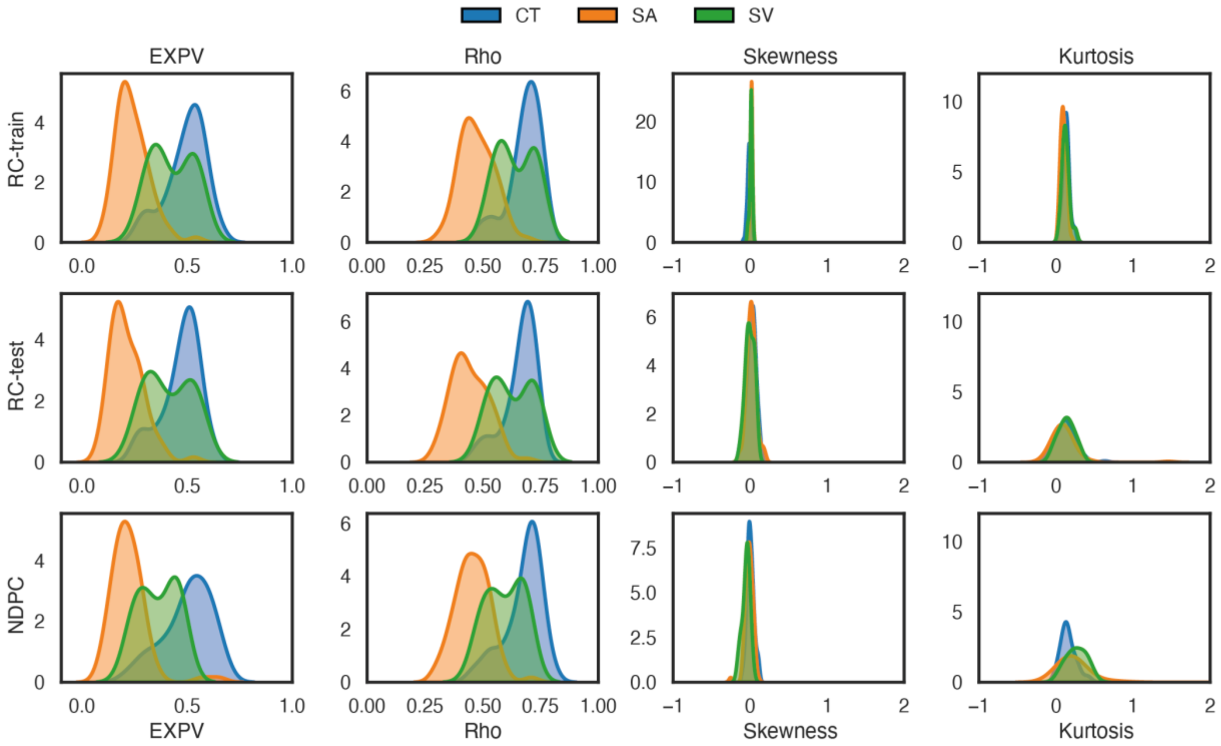

**Figure S1** *Model fit evaluation.*
EXPV = Explained Variance, Rho = Spearman's correlation between true and predicted values, CT = cortical thickness, SA = surface area, SV = subcortical volume. NDPC = Individuals with a neurodevelopmental or psychiatric condition.

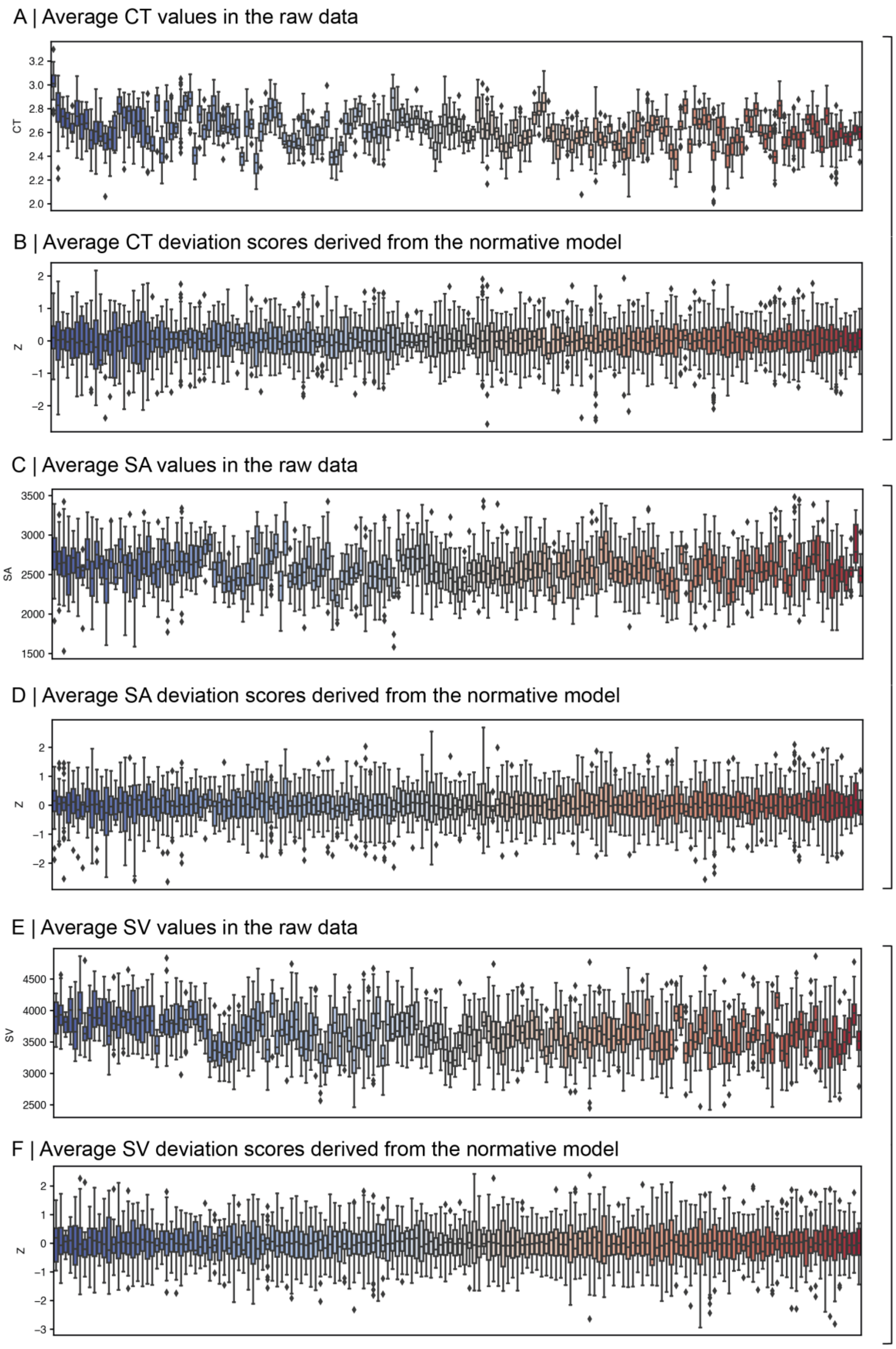

**Figure S2** *Site distributions in raw data and HBR-derived deviation scores (Z) for cortical thickness (CT), surface area (SA) and subcortical volume (SV).*

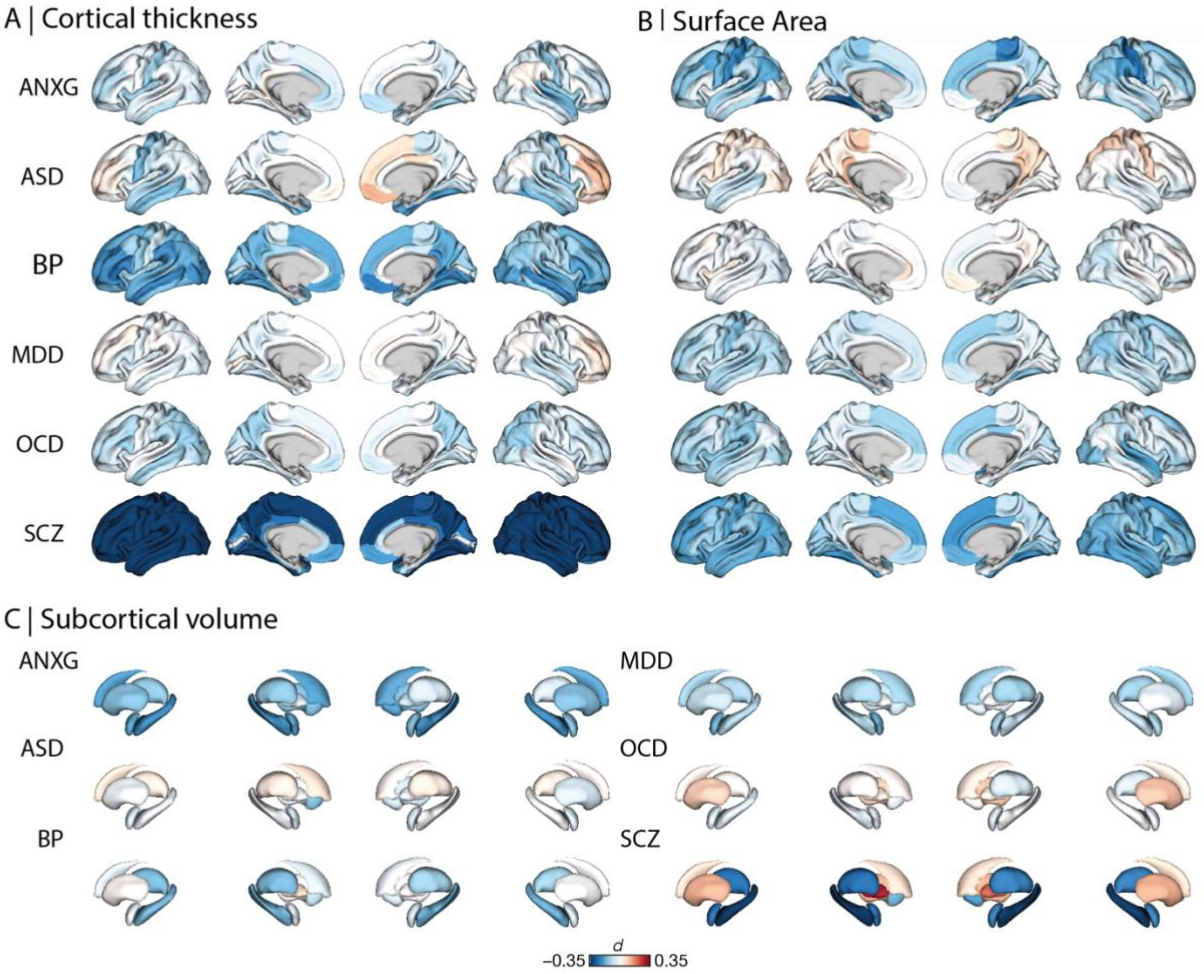

**Figure S3** *Group differences between individuals with and without neurodevelopmental and psychiatric conditions in cortical thickness, surface area, and subcortical volume.*

Cohen's *d* maps were computed between groups that were matched via propensity score matching, taking age and sex into account. As tests were based on deviation scores, which were intrinsically corrected for age, sex, and site in the normative modelling step, these covariates were not included as covariates in t-tests / Cohen's *d* computation. ANXG = Generalized anxiety disorder; ASD = Autism spectrum diagnosis; BP = Bipolar disorder; MDD = Major depressive disorder; OCD = Obsessive-compulsive disorder; SCZ = Schizophrenia.

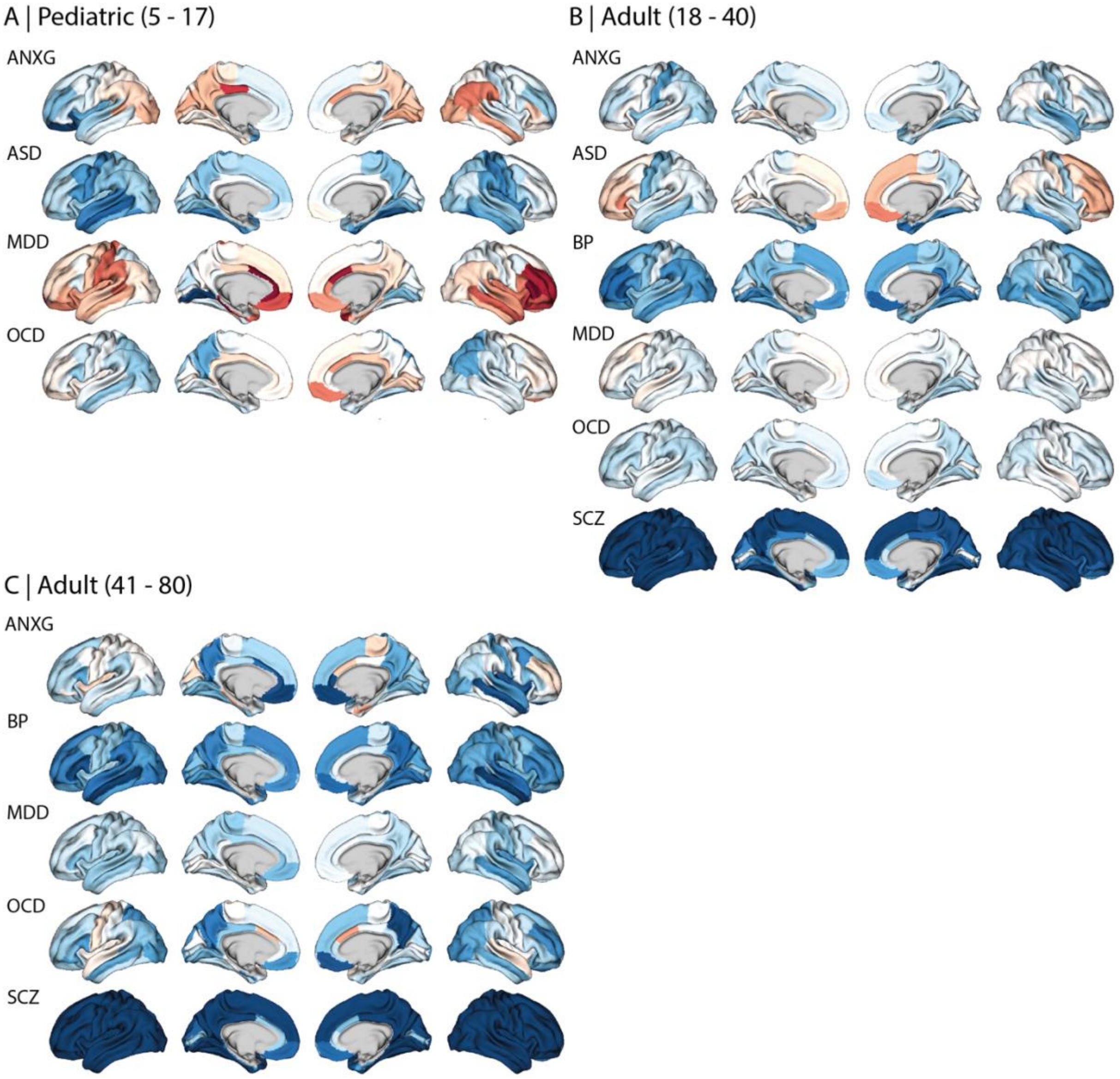

**Figure S4** *Group differences in cortical thickness across age strata.*
Cohen's *d* maps were computed using the same formula as described for the full sample Cohen's *d* maps. They were only computed for groups for which at least 100 diagnosed individuals were available for the respective age group. Reference comparators (RCs) were matched via propensity score matching, taking age and sex into account and matching sample sizes between groups. Sample sizes for the pediatric age strata were: 154 individuals with generalized anxiety disorder (ANXG), 531 individuals with an autism spectrum diagnosis (ASD) for cortical, 490 for subcortical data, 153 individuals with major depressive disorder (MDD), and 360 individuals with obsessive-compulsive disorder (OCD). Sample sizes for the adult (18-40 years) age strata were: 511 individuals with ANXG, 414 individuals with ASD, 720 individuals with bipolar disorder (BP), 805 individuals with MDD, 1173 individuals with OCD, 1356 individuals with schizophrenia spectrum disorder (SCZ). Sample sizes for the adult (41-80 years) age strata were: 100 individuals with ANXG, 579 individuals with BP, 579 individuals with MDD, 242 individuals with OCD, 579 individuals with SCZ.

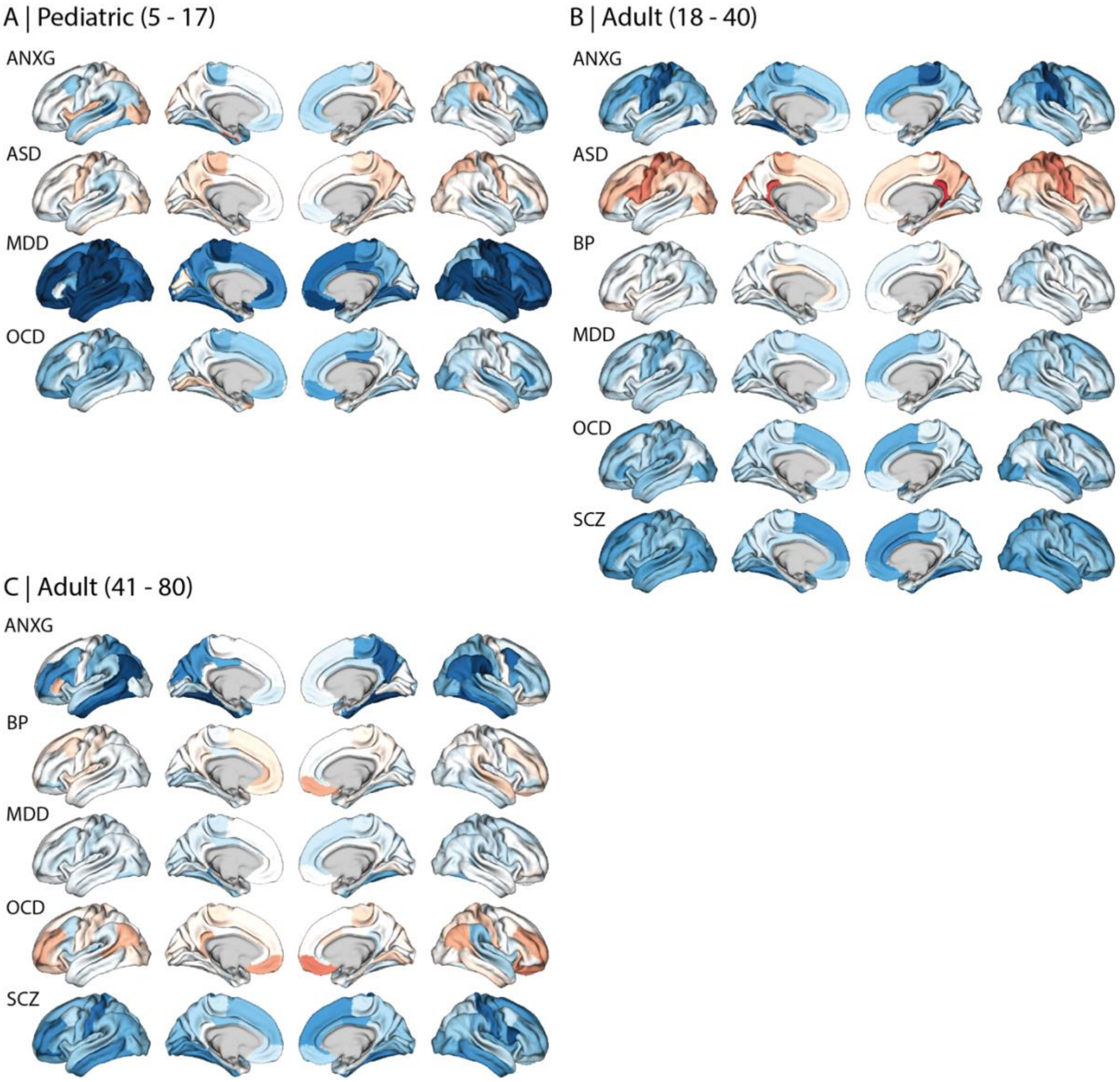

**Figure S5** *Group differences in surface area across age strata.*
Cohen's d maps were computed using the same formula as described for the full sample Cohen's d maps. They were only computed for groups for which at least 100 diagnosed individuals were available for the respective age group. Reference comparators (RCs) were matched via propensity score matching, taking age and sex into account and matching sample sizes between groups. Sample sizes for the pediatric age strata were: 154 individuals with generalized anxiety disorder (ANXG), 531 individuals with an autism spectrum diagnosis (ASD) for cortical, 490 for subcortical data, 153 individuals with major depressive disorder (MDD), and 360 individuals with obsessive-compulsive disorder (OCD). Sample sizes for the adult (18-40 years) age strata were: 511 individuals with ANXG, 414 individuals with ASD, 720 individuals with bipolar disorder (BP), 805 individuals with MDD, 1173 individuals with OCD, 1356 individuals with schizophrenia spectrum disorder (SCZ). Sample sizes for the adult (41-80 years) age strata were: 100 individuals with ANXG, 579 individuals with BP, 579 individuals with MDD, 242 individuals with OCD, 579 individuals with SCZ.

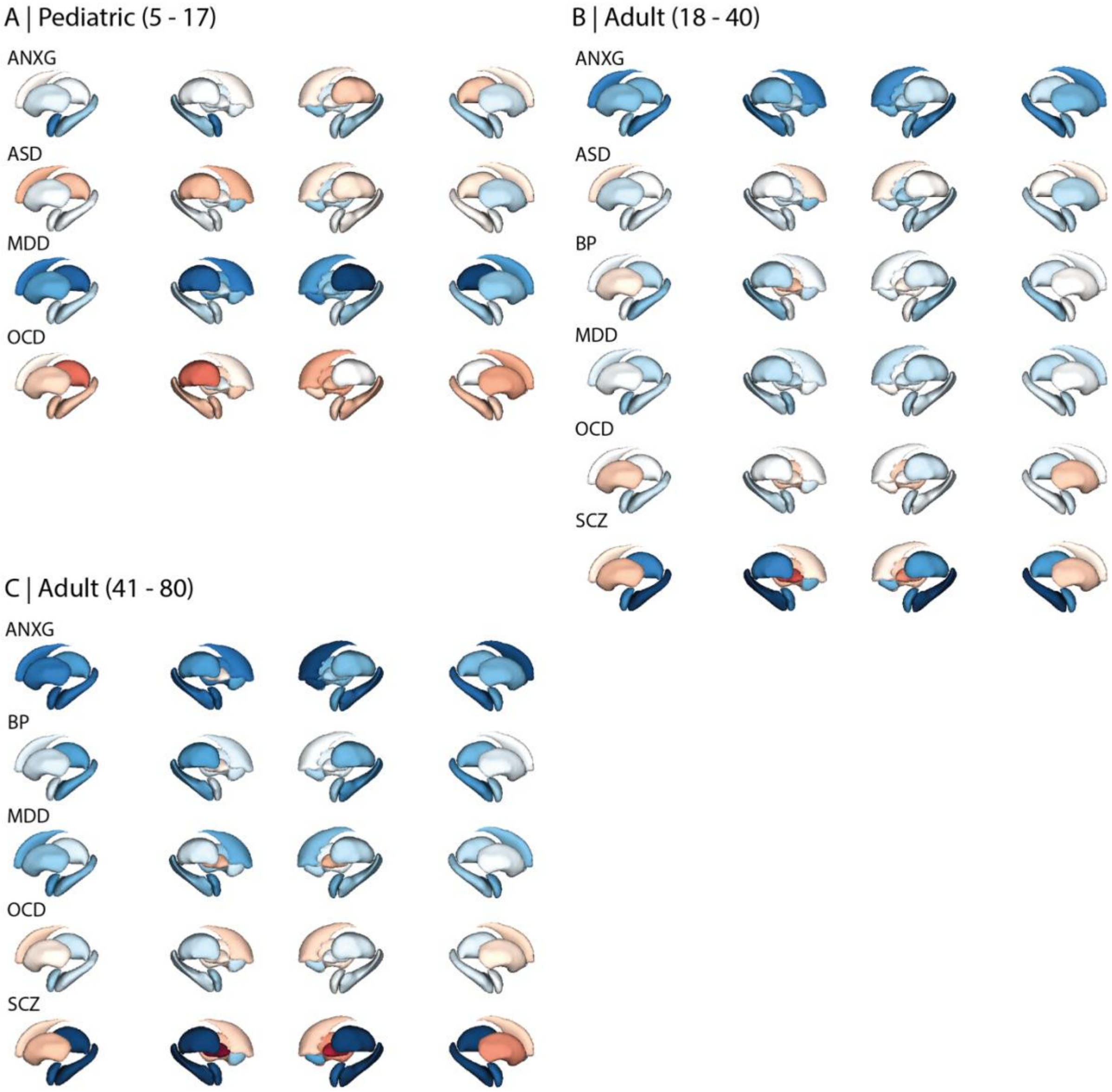

**Figure S6** *Group differences in subcortical volume across age strata.*
Cohen's d maps were computed using the same formula as described for the full sample Cohen's d maps. They were only computed for groups for which at least 100 diagnosed individuals were available for the respective age group. Reference comparators (RCs) were matched via propensity score matching, taking age and sex into account and matching sample sizes between groups. Sample sizes for the pediatric age strata were: 154 individuals with generalized anxiety disorder (ANXG), 531 individuals with an autism spectrum diagnosis (ASD) for cortical, 490 for subcortical data, 153 individuals with major depressive disorder (MDD), and 360 individuals with obsessive-compulsive disorder (OCD). Sample sizes for the adult (18-40 years) age strata were: 511 individuals with ANXG, 414 individuals with ASD, 720 individuals with bipolar disorder (BP), 805 individuals with MDD, 1173 individuals with OCD, 1356 individuals with schizophrenia spectrum disorder (SCZ). Sample sizes for the adult (41-80 years) age strata were: 100 individuals with ANXG, 579 individuals with BP, 579 individuals with MDD, 242 individuals with OCD, 579 individuals with SCZ.

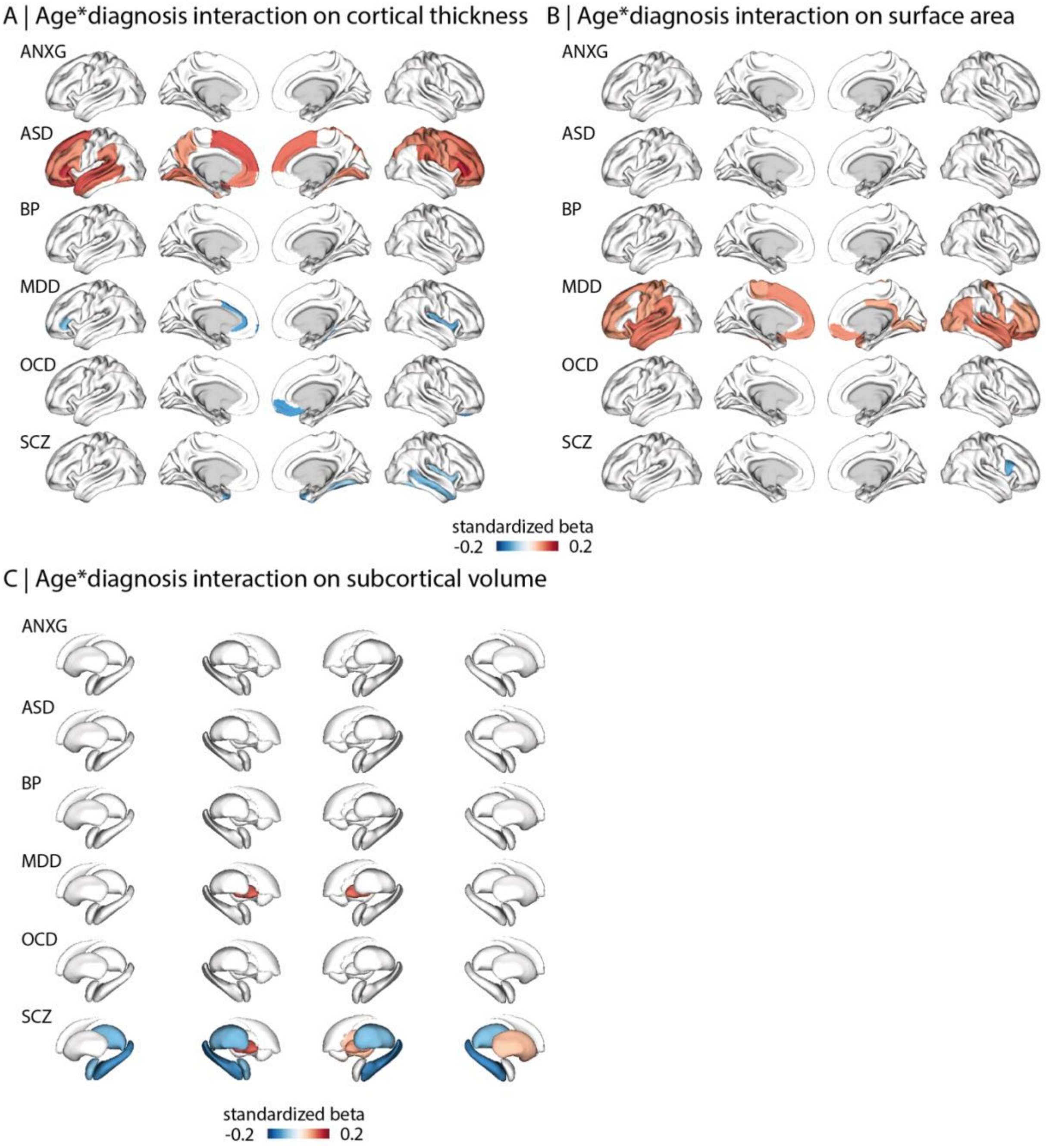

**Figure S7** *Interaction between age and diagnosis for cortical thickness, surface area, and subcortical volume.* Standardized general linear models were used to examine diagnosis × age interactions while controlling for main effects of age, sex, and diagnosis. All p<0.05 FDR.

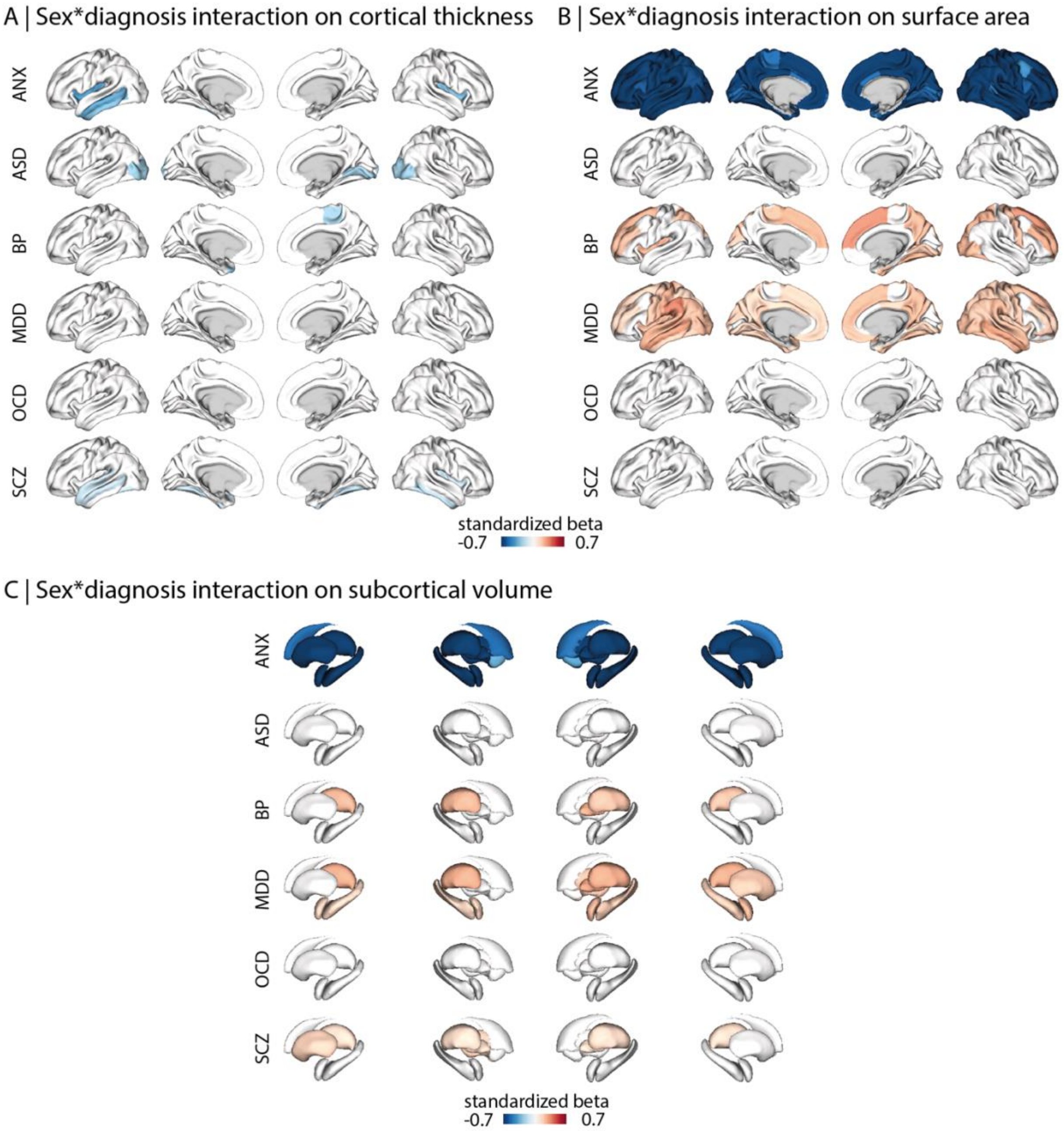

**Figure S8** *Interaction between sex and diagnosis for cortical thickness, surface area, and subcortical volume.* Standardized general linear models were used to examine diagnosis × sex interactions while controlling for main effects of age, sex and diagnosis. All p<0.05 FDR.

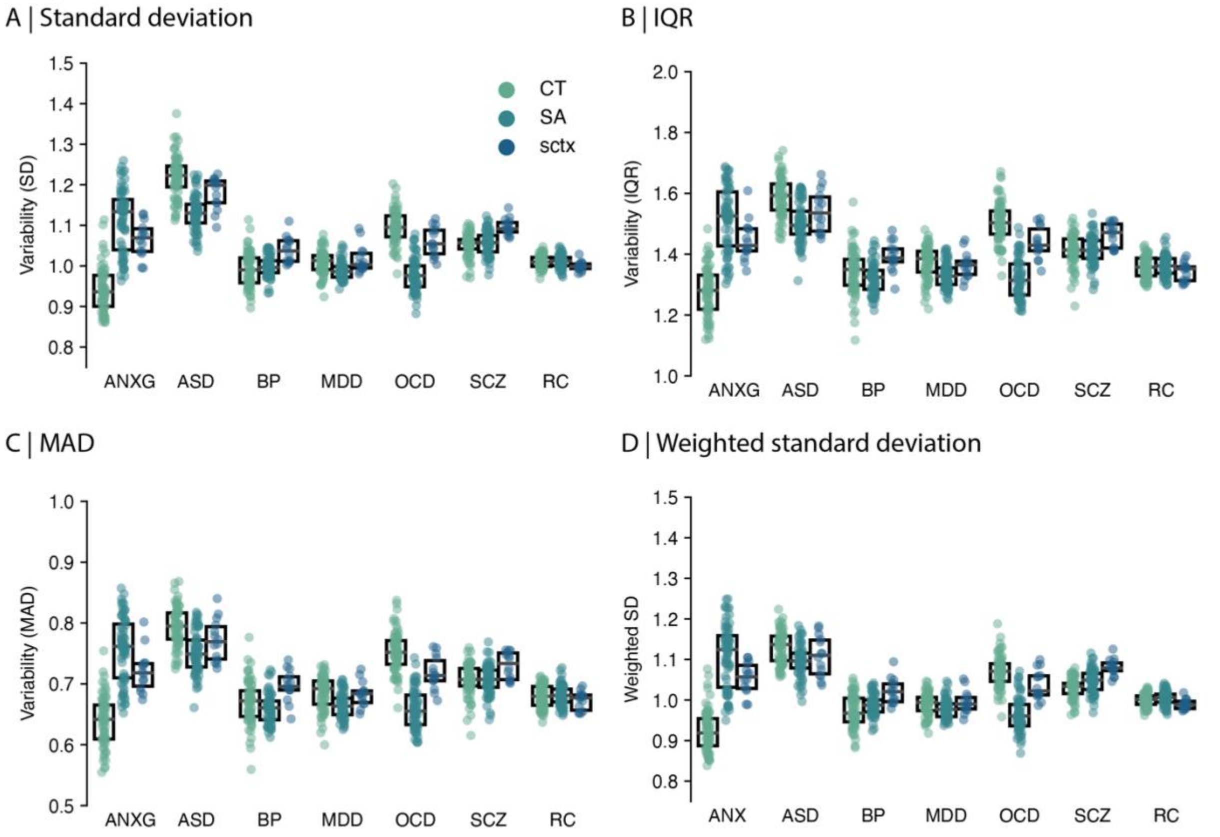

**Figure S9** *Different measures of within-diagnosis variability.*

A) standard deviation (SD), B) interquartile range (IQR), C) mean absolute deviation (MAD), and D) weighted standard deviation (SD computed within sites and aggregated by weighting by sample size per site). Each dot represents a region in which variability is estimated across individuals.

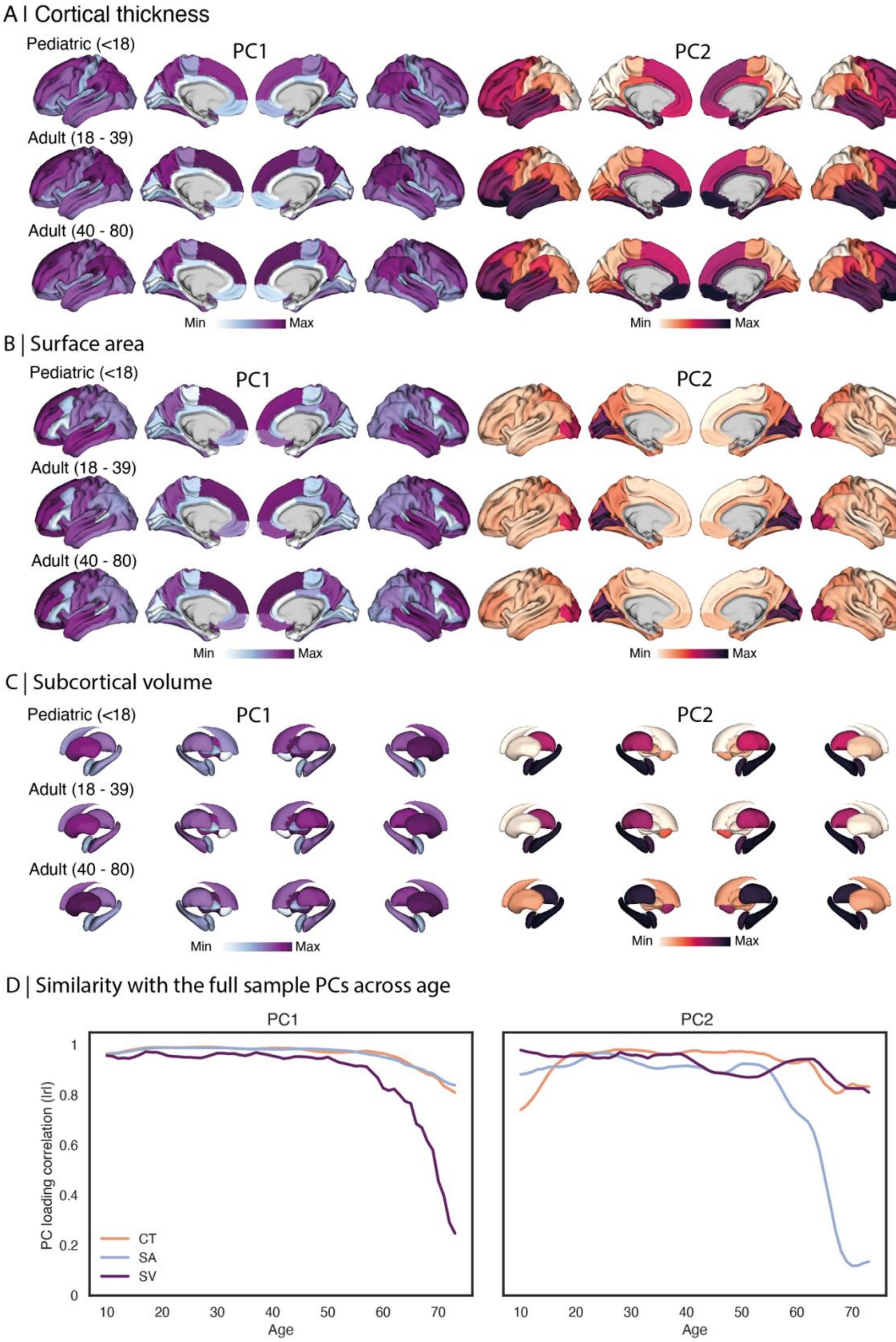

**Figure S10** *Main axes of structural deviations across three age strata.*
Principal components were derived as described in the main, but separately for pediatric participants (5-17 years), younger adults (18 – 39 years) and relatively older adults (40 – 80 years). They were computed in the full sample for cortical thickness (CT; A), surface area (SA; B) and subcortical volume (SV; C). D) depicts the similarity (Spearman's rho) of spatial patterns (i.e., principal component loadings) derived within 10-year age windows and

the full age range. Sliding windows were moved by 1 year in every iteration. PC1 = First principal component, PC2 = Second component.

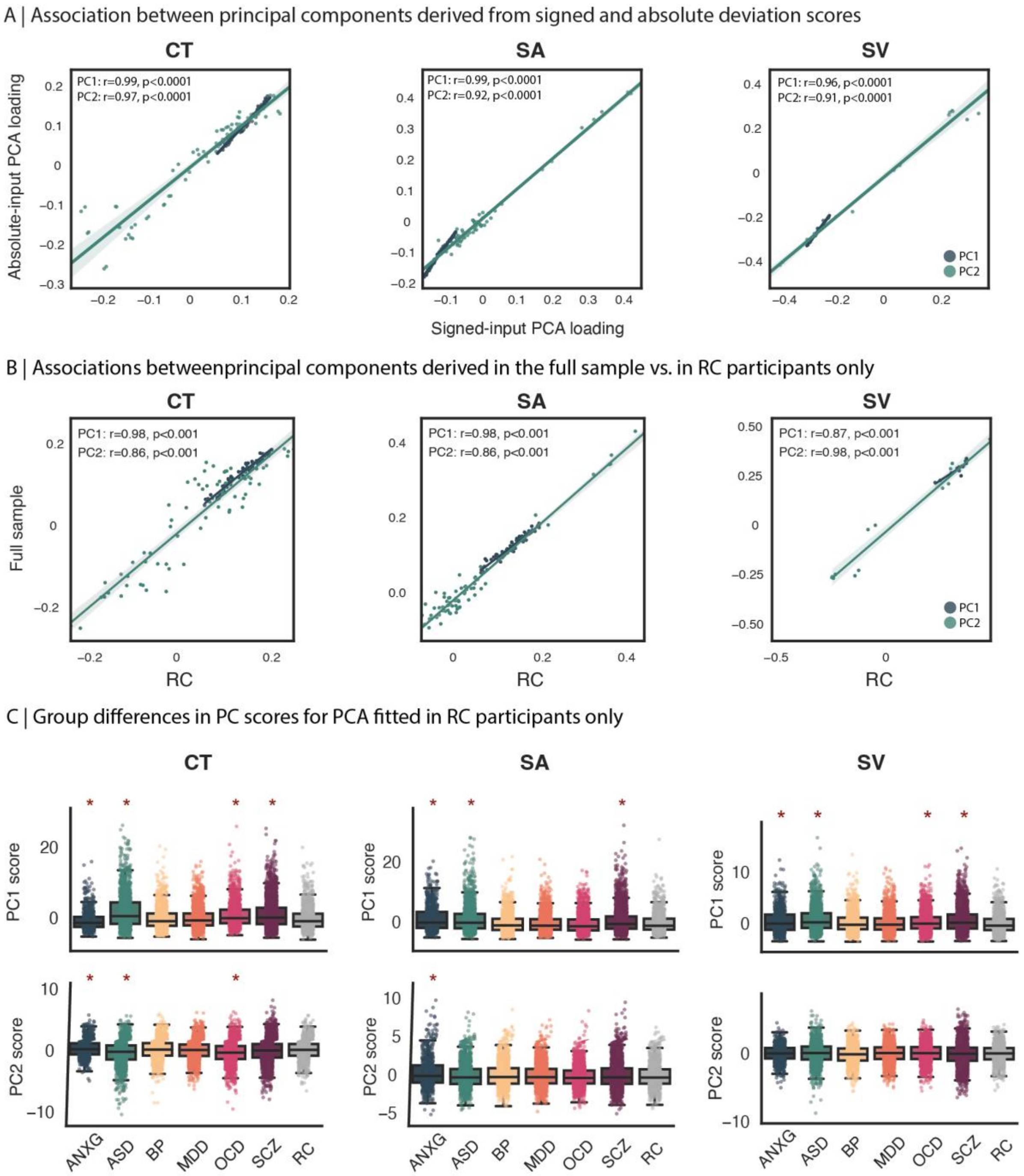

**Figure S11** *Capturing dominant axes of structural deviations based on signed deviation scores and in the reference cohort (RC).*

A) Regression plots depict the similarities between principal components computed based on signed or absolute deviation scores, for the full sample. B) Regression plots depict the spatial association between principal components (PCs) derived from 50% of the RC cohort and PCs derived in the full sample. Significance was assessed variogram permutation tests[14] for cortical and variogram tests for subcortical data. C) Case-comparison differences in PC scores were computed based on scores derived from fitting the principal component analysis in 50% of the RC only, and applying the fitted PCA to the individuals with neurodevelopmental and psychiatric conditions (NDPC)s as well as the remaining 50% of the RC. NDPC-comparison differences were computed between patients and the held out 50% of RCs, where individuals were matched based on propensity score matching, taking age and sex into account. * = $p_{FDR} < 0.05$. CT = Cortical thickness; SA = Surface area; SV =

Subcortical volume. ANXG = Generalized anxiety disorder, ASD = Autism, BP = Bipolar disorder, MDD = Major depressive disorder, OCD = obsessive-compulsive disorder, SCZ = Schizophrenia spectrum disorder.

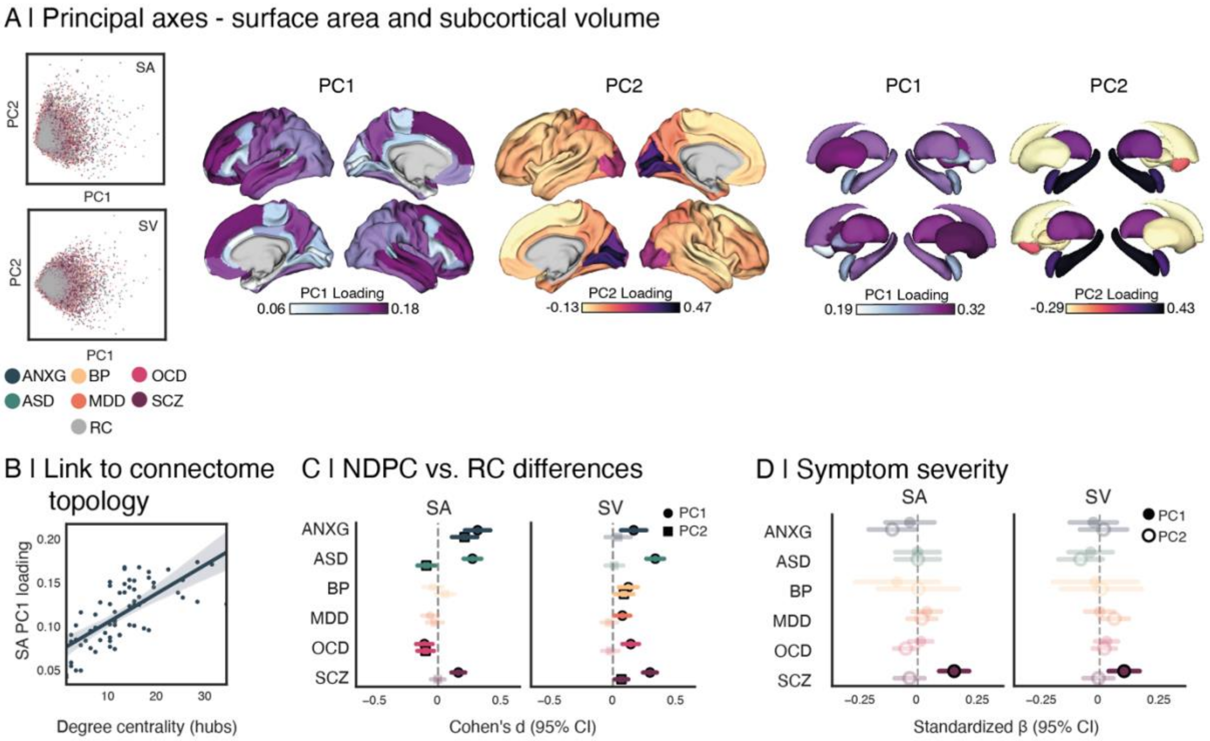

**Figure S12** *Principles of structural deviations in surface area and subcortical volume.*
A) Two main axes of structural deviations derived in the full sample for surface area (SA; left) and subcortical volumes (SV; right). Scatters depict the principal component scores for each individual, showing large overlap between the diagnoses. B) Spatial association between the principal component (PC1) for surface area and a normative connectome hub map derived in a subset of unrelated participants of the Human Connectome Young Adults (HCP-YA) sample (see Main). C) Cohen's d group differences in PC scores between individuals with a neurodevelopmental or psychiatric condition (NDPC) and reference comparators (RC). D) Association between symptom severity and PC scores for surface area and subcortical volume. In C) and D), horizontal bars indicate 95% confidence intervals. ANXG = Generalized anxiety disorder, ASD = Autism, BP = Bipolar disorder, MDD = Major depressive disorder, OCD = obsessive-compulsive disorder, SCZ = Schizophrenia spectrum disorder.

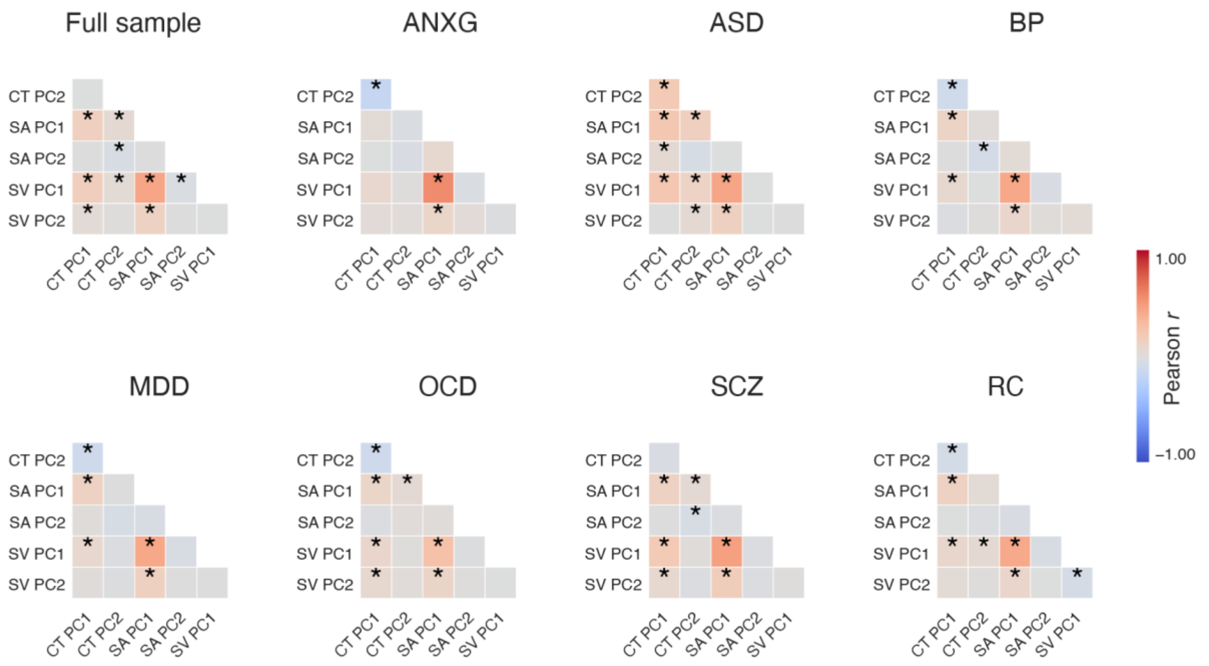

**Figure S13** *Associations between PC scores across modalities.*

In order to test whether individuals who show stronger expressions of deviations along an organizational axis identified in one modality also show strong stronger expression in another modality, we computed pairwise Pearson's correlations between the scores. * = $p_{FDR} < 0.05$. ANXG = Generalized anxiety disorder, ASD = Autism, BP = Bipolar disorder, MDD = Major depressive disorder, OCD = obsessive-compulsive disorder, SCZ = Schizophrenia spectrum disorder. CT = cortical thickness, SA = surface area, SV = subcortical volumes.

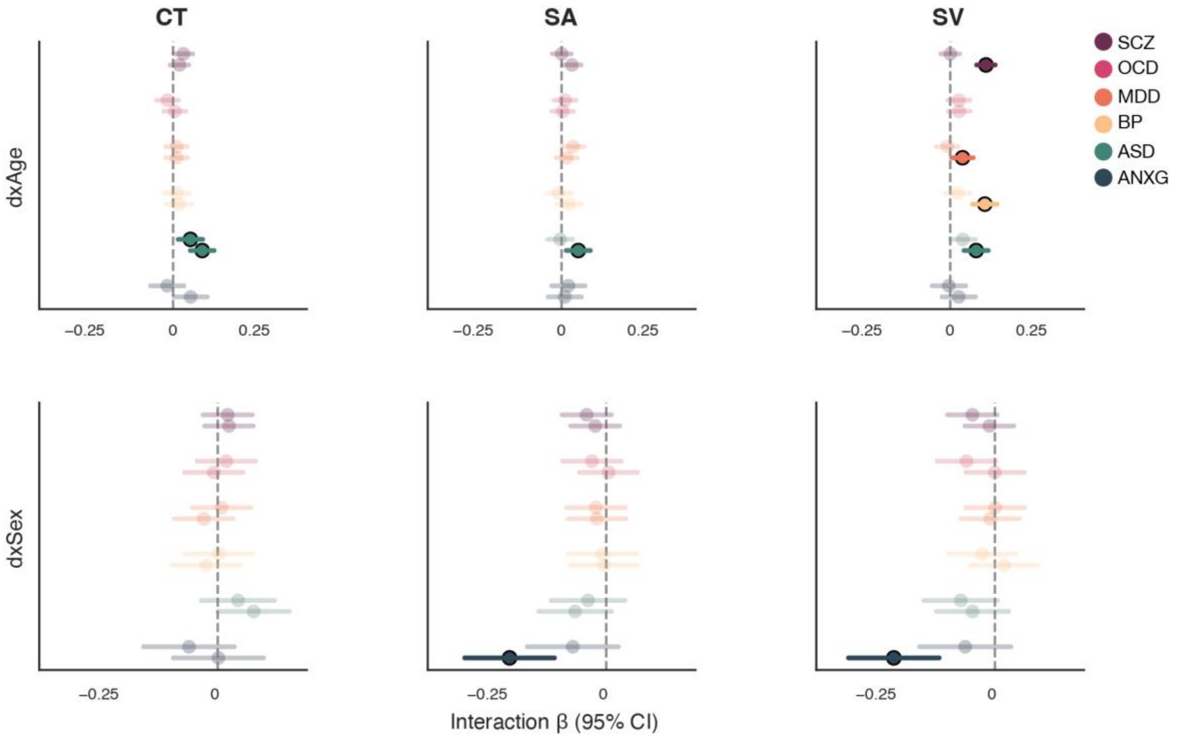

**Figure S14** *Interaction effects of diagnosis with age and sex on principal component scores across structural features.*

Forest plots show standardized interaction effects (beta) and 95% confidence intervals for diagnosis-by-age (top row) and diagnosis-by-sex (bottom row) interactions on principal component (PC) scores. Points represent effect estimates for each disorder, and horizontal bars indicate 95% confidence intervals (CI). Solid and dashed positions within each disorder correspond to PC1 and PC2, respectively. Significant effects after false discovery rate (FDR; <0.05) correction are shown with full opacity, whereas non-significant effects are faded. ANXG = Generalized anxiety disorder, ASD = Autism, BP = Bipolar disorder, MDD = Major depressive disorder, OCD = obsessive-compulsive disorder, SCZ = Schizophrenia spectrum disorder. CT = cortical thickness, SA = surface area, SV = subcortical volumes.

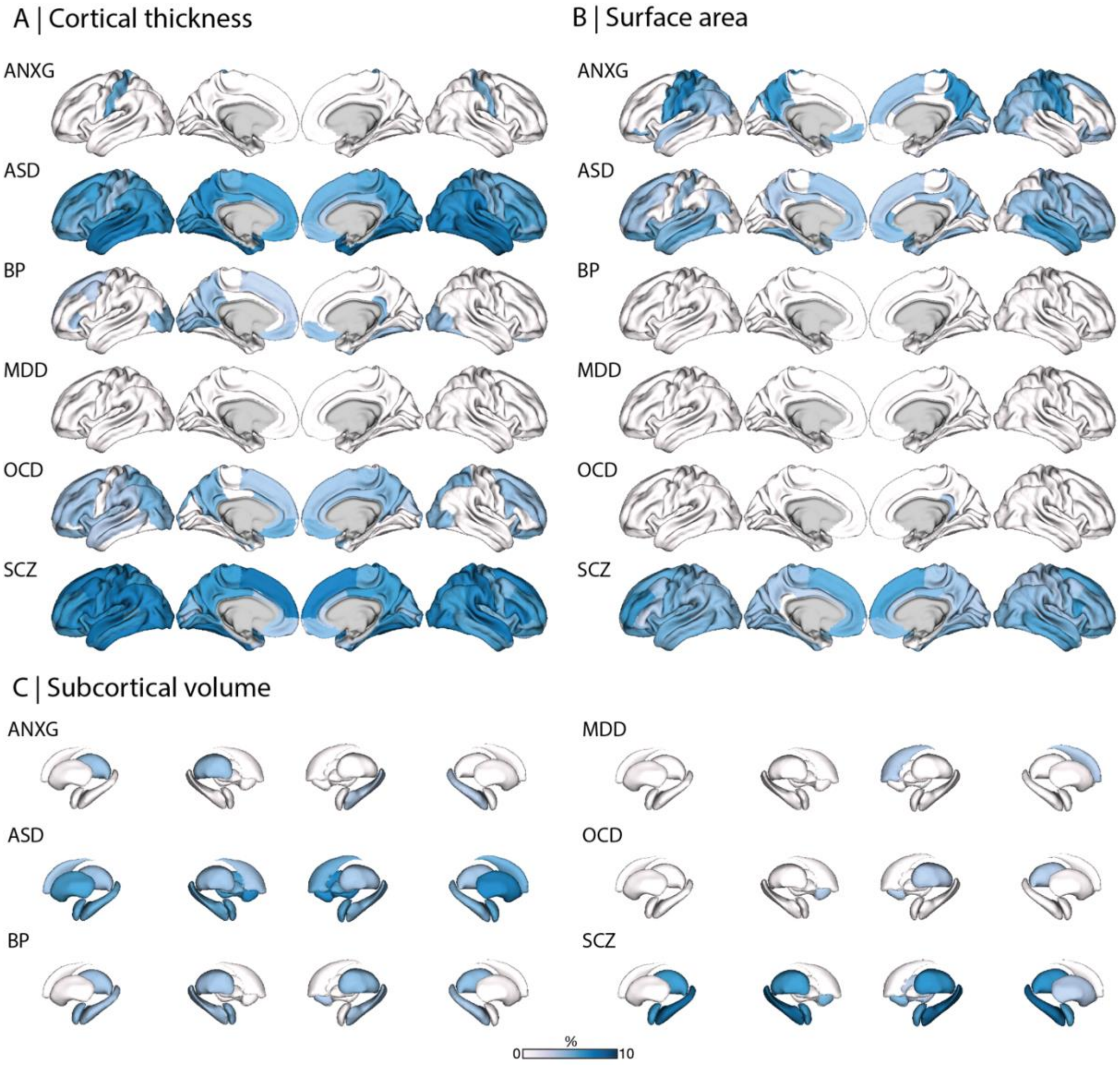

**Figure S15** *Overlap of extreme negative deviations*.
We defined values in the lower 2.5% percentiles (z < 1.96) as extreme negative deviations and computed how often (%) individuals with the same diagnosis show an extreme negative deviation in a given region, contrasting individuals with a neurodevelopmental or psychiatric condition vs reference comparators (RC; 10,000 permutations; p<0.05 FDR). RCs were matched via propensity score matching, taking age and sex into account. ANXG = Generalized anxiety disorder, ASD = Autism spectrum diagnosis, BP = Bipolar disorder, MDD = Major depressive disorder, OCD = obsessive-compulsive disorder, SCZ = Schizophrenia spectrum disorder.

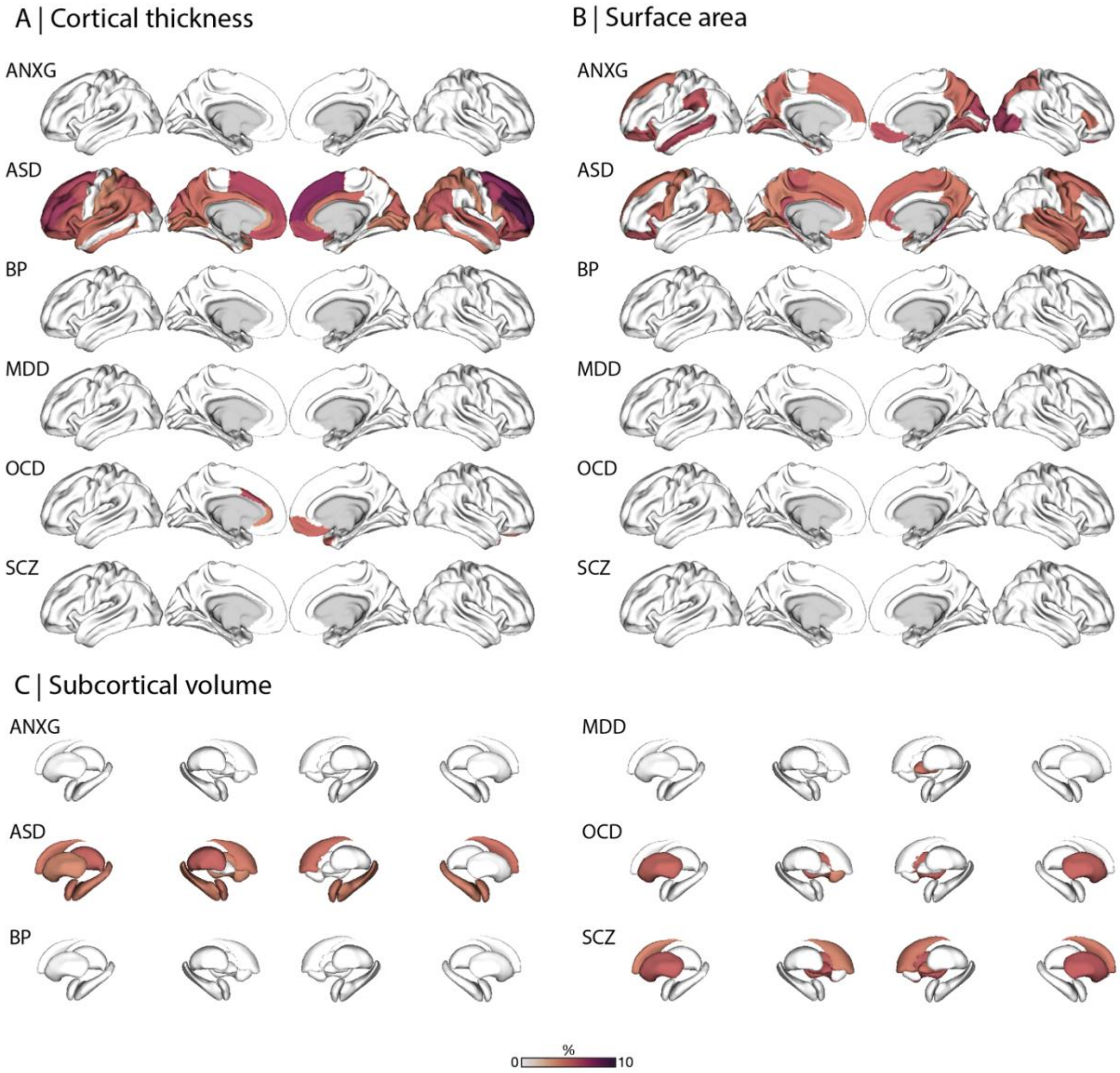

**Figure S16** *Overlap of extreme positive deviations.*
We defined values in the upper 2.5% percentiles (z > 1.96) as extreme negative deviations and computed how often (%) individuals with the same diagnosis show an extreme negative deviation in a given region, contrasting individuals with a neurodevelopmental or psychiatric condition vs reference comparators (RC; 10,000 permutations; p<0.05 FDR). RCs were matched via propensity score matching, taking age and sex into account. ANXG = Generalized anxiety disorder, ASD = Autism, BP = Bipolar disorder, MDD = Major depressive disorder, OCD = obsessive-compulsive disorder, SCZ = Schizophrenia spectrum disorder.

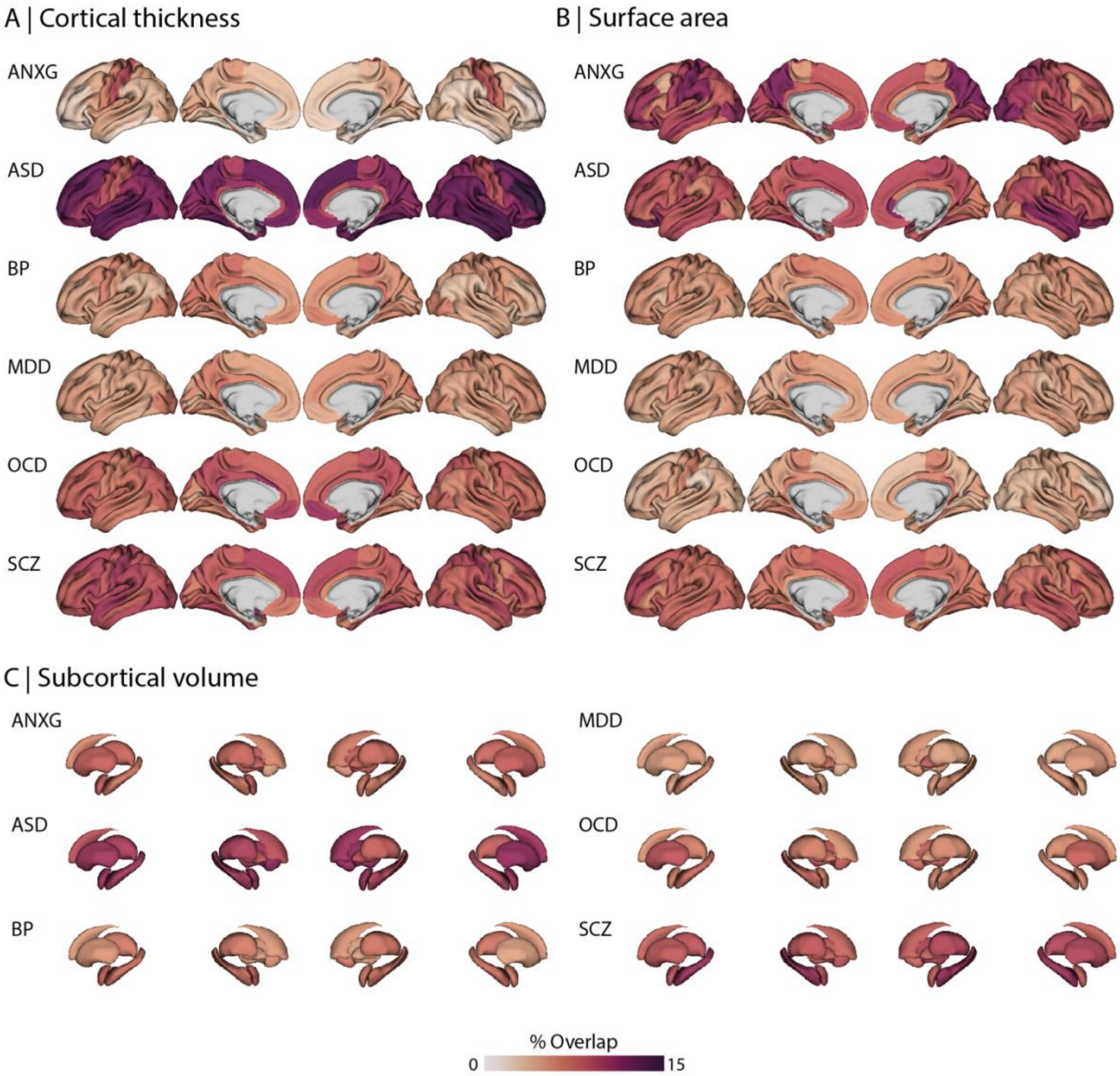

**Figure S17** *Cortical and subcortical maps depicting the percentage overlap of any extreme deviation (|z|>1.96) per region, computed within each diagnostic group.*

Maps are unthresholded. ANXG = Generalized anxiety disorder, ASD = Autism, BP = Bipolar disorder, MDD = Major depressive disorder, OCD = obsessive-compulsive disorder, SCZ = Schizophrenia spectrum disorder. CT = cortical thickness, SA = surface area, SV = subcortical volumes.

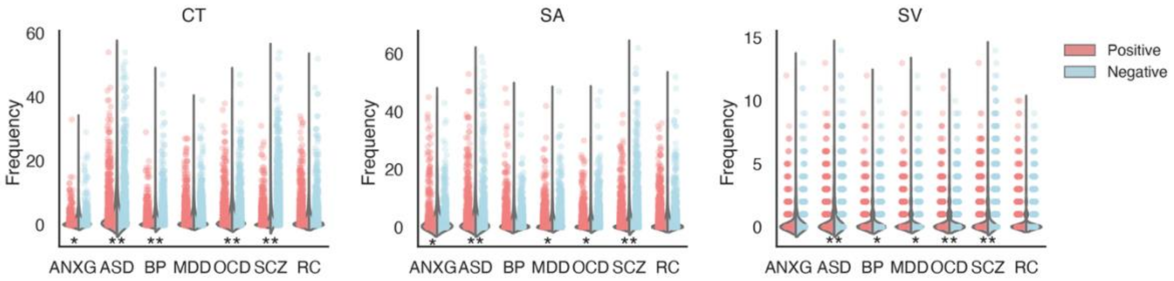

**Figure S18** *Overall number of extreme deviations per individual, across the cortex / subcortex.*
A deviation was labelled as extreme when it was below the 2.5th or higher than the 97.5th percentiles (|z|>1.96).
* Indicates significantly more extreme deviations in individuals with neurodevelopmental or psychiatric conditions than RCs (FDR<0.05 within modality and direction of deviation). ANXG= generalized anxiety disorder, ASD = autism, BP = Bipolar disorder, MDD = Major depressive disorder, OCD = obsessive-compulsive disorder, SCZ = Schizophrenia spectrum disorder, RC = Reference cohort, CT = cortical thickness, SA = surface area, SV = subcortical volumes.

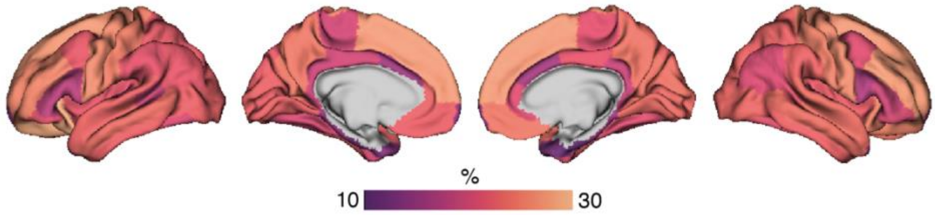

**Figure S19** *Hubs of extreme deviations computed for surface area.*
Hub computation was performed in the same way as described for cortical thickness in the Main, using an unrelated subset of the Human Connectome Young Adult sample as the reference connectome.

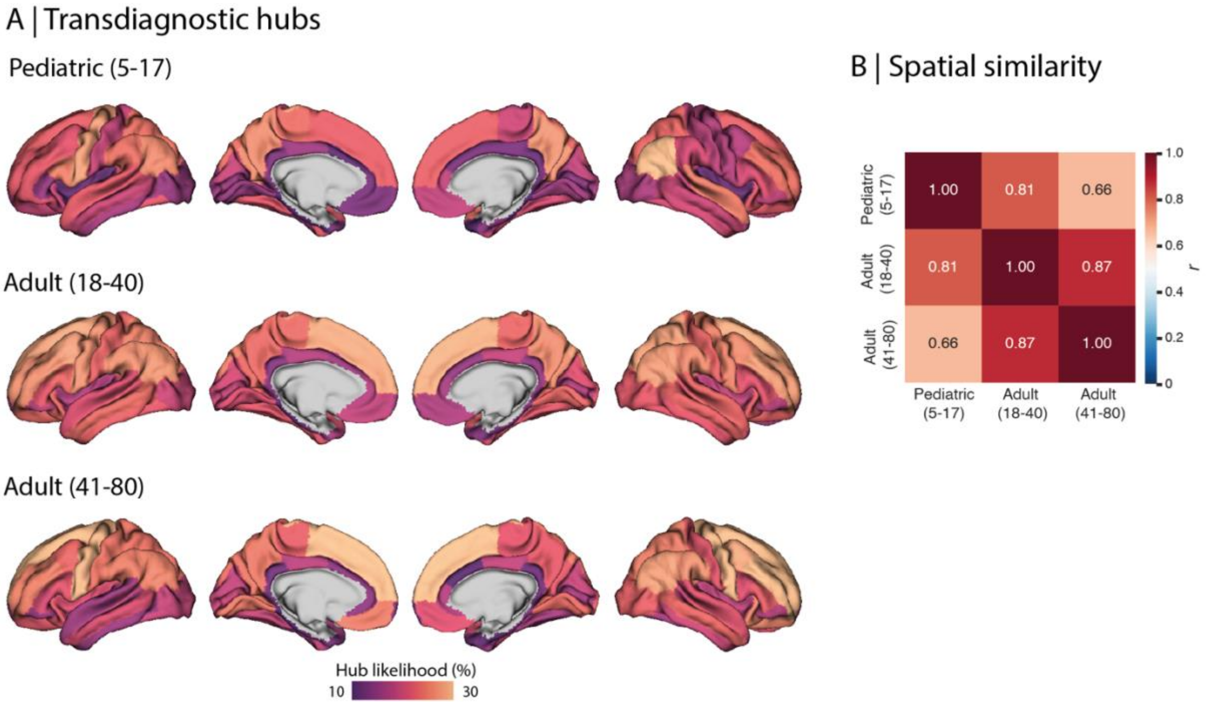

**Figure S20** *Transdiagnostic hubs of extreme deviations across age strata.*

**A)** The pediatric map (5-17 years old) includes data from individuals with an autism spectrum diagnosis (ASD), major depressive disorder (MDD), generalized anxiety (ANXG), and obsessive-compulsive disorder (OCD). The adult (18-40 years old) map includes individuals with ASD, ANX, bipolar disorder (BP), MDD, OCD, and schizophrenia (SCZ). The map of relatively older adults (41-80 years) includes individuals with ANXG, BP, OCD, MDD, SCZ. **B)** Depicts Spearman's rho correlation between hub maps across the three age strata.

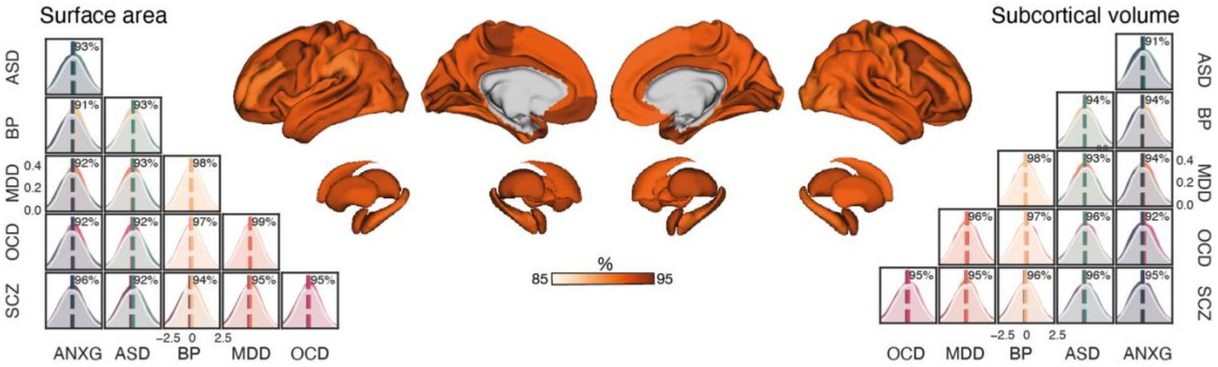

**Figure S21** *Transdiagnostic distribution overlap in surface area and subcortical volumes.*
Distribution overlap was computed as the shared area under two distributions. Distribution plots: Pairwise distribution overlap in cortical thickness across individuals and (sub-)cortical regions. Cortical and subcortical plots: Spatial variation in transdiagnostic surface area (top) and subcortical volumes (bottom) overlap, computed as the average of all pairwise comparisons.

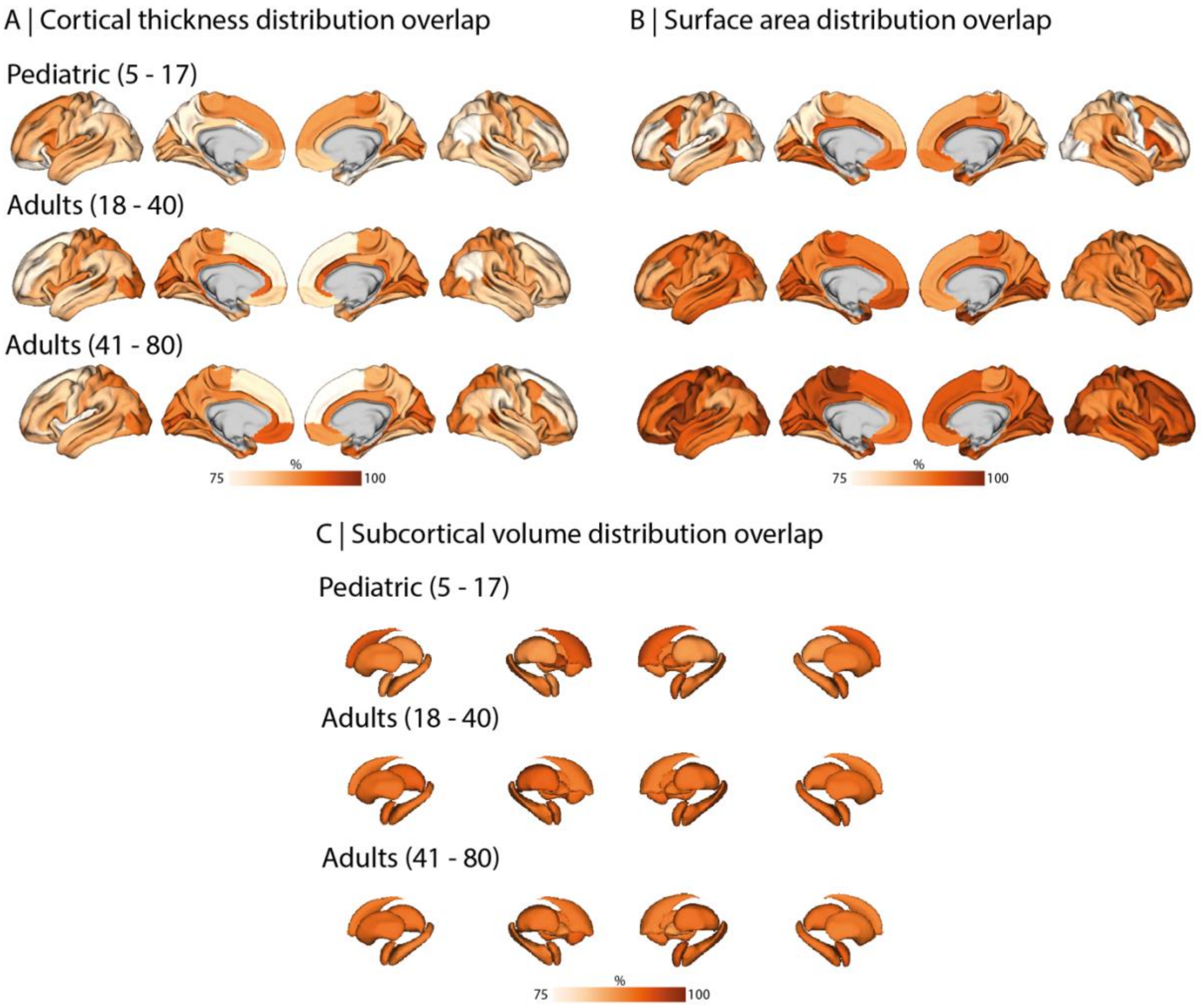

**Figure S22** *Distribution overlap across age strata.*
The pediatric map (5-17 years old) includes data from individuals with an autism spectrum diagnosis (ASD), major depressive disorder (MDD), generalized anxiety (ANXG), and obsessive-compulsive disorder (OCD). The adult (18-40y) map includes individuals with ASD, ANX, bipolar disorder (BP), MDD, OCD, and schizophrenia (SCZ). The map of relatively older adults (41-80 years old) includes individuals with ANXG, BP, OCD, MDD, SCZ. Distribution overlap was computed as the shared area under two distributions for cortical thickness **(A)**, surface area **(B),** and subcortical volumes **(C)**.

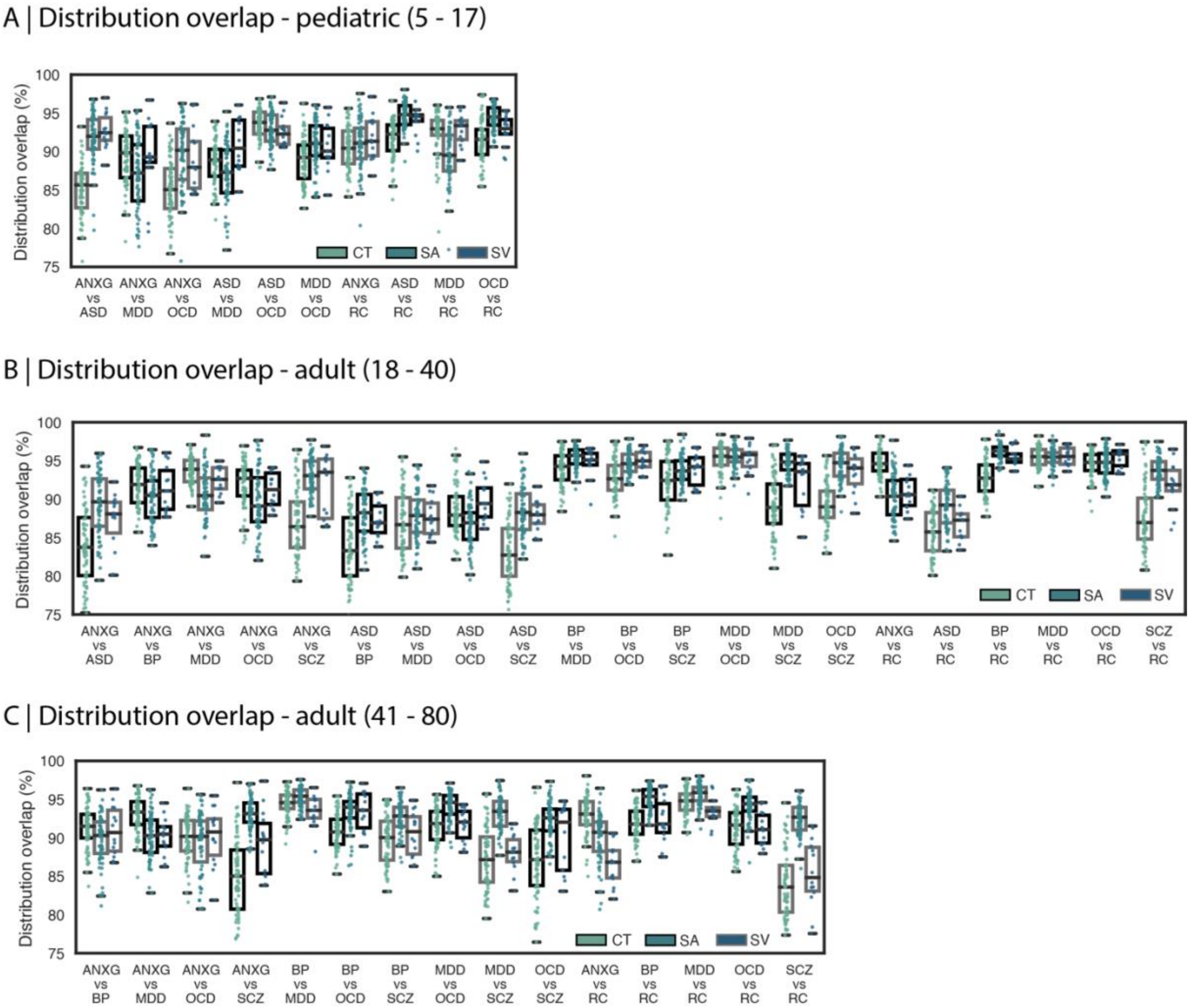

**Figure S23** *Regional distribution overlap across age strata.*

Pairwise comparisons were limited to diagnostic groups for which at least 100 individuals were available per age strata. ANXG = generalized anxiety disorder, ASD = autism spectrum diagnosis, BP = bipolar disorder, MDD = major depressive disorder, OCD = obsessive-compulsive disorder, SCZ = schizophrenia spectrum disorder, RC = reference cohort.

#### *Supplementary references*

1. Satterthwaite, T. D. *et al.* The Philadelphia Neurodevelopmental Cohort: A publicly available resource for the study of normal and abnormal brain development in youth. *NeuroImage* **124**, 1115–1119 (2016).
2. Alexander, L. M. *et al.* An open resource for transdiagnostic research in pediatric mental health and learning disorders. *Sci. Data* **4**, 170181 (2017).
3. Glasser, M. F. *et al.* The minimal preprocessing pipelines for the Human Connectome Project. *Neuroimage* **80**, 105–124 (2013).
4. Larivière, S. *et al.* The ENIGMA Toolbox: multiscale neural contextualization of multisite neuroimaging datasets. *Nat. Methods* **18**, 698–700 (2021).
5. Desikan, R. S. *et al.* An automated labeling system for subdividing the human cerebral cortex on MRI scans into gyral based regions of interest. *Neuroimage* **31**, 968–980 (2006).
6. Griffa, A., Aleman-Gomez, Y. & Hagmann, P. Structural and functional connectome from 70 young healthy adults. https://zenodo.org/records/2872624 (2019) doi:https://zenodo.org/record/2872624.
7. Hansen, J. Y. *et al.* Local molecular and global connectomic contributions to cross-disorder cortical abnormalities. *Nat. Commun.* **13**, 4682 (2022).
8. Daducci, A. *et al.* The connectome mapper: an open-source processing pipeline to map connectomes with MRI. *PloS One* **7**, e48121 (2012).
9. Betzel, R. F., Griffa, A., Hagmann, P. & Mišić, B. Distance-dependent consensus thresholds for generating group-representative structural brain networks. *Netw. Neurosci.* **3**, 475–496 (2019).
10. Power, J. D., Barnes, K. A., Snyder, A. Z., Schlaggar, B. L. & Petersen, S. E. Spurious but systematic correlations in functional connectivity MRI networks arise from subject motion. *NeuroImage* **59**, 2142–2154 (2012).
11. de Boer, A. A. A. *et al.* Non-Gaussian normative modelling with hierarchical Bayesian regression. *Imaging Neurosci.* **2**, imag–2–00132 (2024).
12. Bayer, J. M. M. *et al.* Accommodating site variation in neuroimaging data using normative and hierarchical Bayesian models. *NeuroImage* **264**, 119699 (2022).
13. Fortin, J.-P. *et al.* Harmonization of cortical thickness measurements across scanners and sites. *NeuroImage* **167**, 104–120 (2018).
14. Burt, J. B., Helmer, M., Shinn, M., Anticevic, A. & Murray, J. D. Generative modeling of brain maps with spatial autocorrelation. *NeuroImage* **220**, 117038 (2020).
15. Taiwan Aging and Mental Illness Cohort - Taiwan Medical AI and Data Portal. https://data.dmc.nycu.edu.tw/dataset/taiwan-aging-and-mental-illness-cohort-tami.
16. Zhu, J.-D., Wu, Y.-F., Tsai, S.-J., Lin, C.-P. & Yang, A. C. Investigating brain aging trajectory deviations in different brain regions of individuals with schizophrenia using multimodal magnetic resonance imaging and brain-age prediction: a multicenter study. *Transl. Psychiatry* **13**, 82 (2023).
17. Yang, A. C. *et al.* Complexity of spontaneous BOLD activity in default mode network is correlated with cognitive function in normal male elderly: a multiscale entropy analysis. *Neurobiol. Aging* **34**, 428–438 (2013).

18. van Rooij, D. *et al.* Cortical and Subcortical Brain Morphometry Differences Between Patients With Autism Spectrum Disorder and Healthy Individuals Across the Lifespan: Results From the ENIGMA ASD Working Group. *Am J Psychiatry* **175**, 359–369 (2018).
19. Hibar, D. P. *et al.* Cortical abnormalities in bipolar disorder: an MRI analysis of 6503 individuals from the ENIGMA Bipolar Disorder Working Group. *Mol Psychiatry* **23**, 932–942 (2018).
20. Schmaal, L. *et al.* Cortical abnormalities in adults and adolescents with major depression based on brain scans from 20 cohorts worldwide in the ENIGMA Major Depressive Disorder Working Group. *Mol Psychiatry* **22**, 900–909 (2017).
21. Boedhoe, P. S. W. *et al.* Cortical Abnormalities Associated With Pediatric and Adult Obsessive-Compulsive Disorder: Findings From the ENIGMA Obsessive-Compulsive Disorder Working Group. *Am J Psychiatry* **175**, 453–462 (2018).
22. van Erp, T. G. M. *et al.* Cortical Brain Abnormalities in 4474 Individuals With Schizophrenia and 5098 Control Subjects via the Enhancing Neuro Imaging Genetics Through Meta Analysis (ENIGMA) Consortium. *Biol Psychiatry* **84**, 644–654 (2018).
